# Proteomic Profile of Human Colon Organoids: Effects of a multi-mineral intervention alone and in the presence of pro-Inflammatory and anti-inflammatory treatments

**DOI:** 10.1101/2025.03.13.25323857

**Authors:** Muhammad N Aslam, Shannon D McClintock, Gillian Moraga, Daniyal M Nadeem, Isabelle Harber, James Varani

## Abstract

**Introduction:** Aquamin, a multi-mineral product derived from fossilized red marine algae, has been shown to improve colon barrier structure and function. Mesalamine is commonly used as maintenance therapy for patients with mild-to-moderate ulcerative colitis (UC) or those in remission. Our long-term aim is to evaluate if Aquamin can be part of a UC maintenance regimen, examining potential complementary efficacy or synergy with Mesalamine, as well as any possible drug interactions.

**Methods:** Human colon organoids were maintained under controlled conditions or exposed to a pro-inflammatory stimulus to mimic the environment in mild-to-moderate UC. Organoids were treated with Aquamin alone, Mesalamine alone, or the two agents in combination for 14 days. At the end of the treatment period, proteomic analysis was conducted to evaluate protein changes induced by the two agents (individually and in combination).

**Results:** Colon organoids treated with Aquamin or Mesalamine exhibited distinct protein expression profiles. Aquamin enhanced the expression of colon barrier proteins (e.g., cadherin-17 and desmoglein-2). Mesalamine by itself had minimal impact on these moieties; when present with Aquamin, it did not alter Aquamin’s response. By itself, treatment with Mesalamine alone resulted in up-regulation of basement membrane proteins; the combination of Aquamin and Mesalamine was more effective than either alone. In contrast to these results, Mesalamine up-regulated numerous proteins directly related to inflammation including members of the complement and clotting/fibrinolytic cascades. These were down-regulated with Aquamin. When the two agents were utilized in combination, changes in the expression of inflammation-related proteins resembled the profile seen with Mesalamine alone more than the profile obtained with Aquamin. Of interest, the presence of a pro-inflammatory stimulus further highlighted the unique responses to the two interventions, with Mesalamine aligning more closely with the pro-inflammatory stimulus in its effect on the expression of inflammation-associated proteins.

**Conclusion:** The data presented here suggest that the induction of barrier proteins by Aquamin would not be counteracted by the concomitant presence of Mesalamine. The current studies also found no evidence to suggest that the presence of Aquamin would interfere with the capacity of Mesalamine to alter the expression of proteins that are part of the anti-inflammatory shield.

## INTRODUCTION

Ulcerative colitis (UC) is a chronic bowel disease characterized by inflammation of the colonic lining and the formation of superficial ulcers in the bowel wall (1-4). Current therapeutic strategies are aimed at mitigating inflammation. Traditional treatment options include broadly acting anti-inflammatory agents like corticosteroids (5) and non-steroidal anti-inflammatories such as Mesalamine (6). In recent years, advancements in our understanding of the underlying immunopathological mechanisms of UC have led to the development and increased use of biological agents. These biologics are designed to target specific components of the inflammatory cascade. Notable among these are tumor necrosis factor-α (TNF-α) inhibitors, interleukin-blocking molecules, agents that disrupt leukocyte trafficking, and those that interfere with upstream signaling pathways leading to cytokine production and immune activation (7-9).

Abnormal barrier structure and function in the gastrointestinal tract, marked by increased mucosal permeability, is a well-known pathological feature of UC (10-14). While barrier dysfunction is a consequence of colonic inflammation, recent studies suggest that barrier impairment may precede the onset of inflammation in the pathogenesis of UC (15-17). However, no current therapies specifically address this fundamental aspect of disease pathophysiology (18,19). Previous studies in our laboratory utilizing human colon organoid cultures have shown that Aquamin, a multi-mineral product derived from the skeletal remains of red marine algae, promotes the production of various proteins that contribute to barrier structure (20-23). Included were significant increase in several adherens junction and desmosomal proteins. Proteins of the mucous layer, including mucins and trefoils, as well as components of the basement membrane and other cell-cell or cell-matrix adhesion molecule were also up-regulated. These changes in barrier protein expression were associated with improved permeability control and increased organoid cohesion. Notably, the same Aquamin-induced changes in barrier protein expression observed in colon organoids from healthy donor tissue were also seen in colonoids derived from UC patient tissue (24).

These preclinical findings from organoid culture, i.e. – increased barrier protein expression in response to Aquamin – have been validated in a 90-day interventional trial with healthy adult subjects (25). More recently, we carried out a 180-day interventional trial with UC patients at the mild-to-moderate disease state or in remission (26). Improved barrier protein expression was observed in subjects receiving Aquamin compared to those receiving a placebo. No safety or tolerability issues were identified in either trial. Based on these *in vitro* and *in vivo* observations, along with previous findings demonstrating Aquamin’s ability to suppress gut inflammation in mice (27-29), we envision the use of Aquamin as an ancillary intervention to help improve the gastrointestinal barrier in individuals with mild-to-moderate UC or with disease in remission.

Mesalamine (or one of the other anti-inflammatory amino salicylate drugs) is commonly used as part of their maintenance regimen for most individuals with mild-to-moderate or in-remission UC. As such, it would be important to know if and to what extent the use of Aquamin along with one of these drugs would provide complementary efficacy or if, in fact, there might be significant synergy between them. Whether drug-interference might occur would also be important to know. As a first step in addressing these issues, human colon organoid cultures were treated with Aquamin alone, Mesalamine alone or the two agents in combination. At the end of a two-week treatment period, the proteomic signature of control and treated organoids was assessed. In parallel, organoids were challenged with a combination of pro-inflammatory effectors (i.e., bacterial lipopolysaccharide [LPS], tumor necrosis factor-α [TNF-α], interleukin-1β [IL-1β] and interferon-γ [IFN-γ]) and treated in the same manner (i.e., with either intervention alone or with the two in combination). The findings are described herein.

## MATERIALS AND METHODS

### Aquamin

Aquamin is a product rich in calcium, magnesium, and multiple trace elements, sourced from the skeletal remains of the red marine algae Lithothamnion sp (30) (Marigot Ltd, Cork, Ireland). The ratio of calcium to magnesium in Aquamin^®^ is about 12:1, and it includes measurable amounts of seventy-two trace minerals. An independent laboratory (Advanced Laboratories in Salt Lake City, Utah) determined the mineral composition of Aquamin using Inductively Coupled Plasma Optical Emission Spectrometry. Supplement Table 1 lists the detected elements in Aquamin^®^ and their relative quantities. Available as a dietary supplement (GRAS 000028), Aquamin is incorporated into various products for human consumption in Europe, Asia, Australia, and North America. In the ex vivo studies discussed here, Aquamin was used at a concentration of 0.925 mg/mL, providing a final calcium concentration of 3 mM. This specific concentration was selected based on prior studies showing increases in multiple cell-cell adhesion proteins (cadherins and desmogleins) and an augmentation of desmosomes as observed through electron microscopy (20,21,23,24).

### Mesalamine

Mesalamine (5-aminosalicylic acid [5-ASA]) was purchased from Sigma-Aldrich (product# A3537). A stock solution of 200 mg/mL was prepared in PBS and diluted in cell culture medium from there. Mesalamine was initially tested over a range of concentrations (50 µg to 250 µg/mL) for organoid toxicity at day-14 with cultures established from two subjects. As shown in the Supplement Figure 1, no toxicity was evident at these doses. Toxicity was defined based on morphological evidence of organoid growth suppression (smaller organoids at the end of the treatment period), organoid failure to demonstrate features of differentiation (formation of thick walls and a decrease in budding structures) and loss of tissue integrity. Additionally, the proteomic signature (by reviewing a list of top canonical pathways) of these mesalamine treated colon organoids suggest using a higher dose (Supplement Figure 2). A Mesalamine concentration of 200 µg/mL (1.306 mM) was eventually chosen for the studies described in this manuscript based on the previous studies exploring mechanisms of mesalamine in intestinal cell lines (31,32).

### LPS-cytokine mix

To replicate the conditions of a chronically inflamed colon, a combination of lipopolysaccharide (LPS) from Escherichia coli and three pro-inflammatory cytokines was employed (33). The stock solution included LPS (1 µg/mL, Sigma), tumor necrosis factor-α (TNF-α; 50 ng/mL, Sigma), interleukin-1β (IL-1β; 25 ng/mL, Shenandoah Biotech), and interferon-γ (IFN-γ; 50 ng/mL, Sigma). We tested this pro-inflammatory mixture across a broad concentration range for organoid toxicity, as detailed in our previous report (23). In the current study, experiments used a 1:250 dilution of the LPS-cytokine stock solution, delivering 4000 pg of LPS, 200 pg each of TNF-α and IFN-γ, and 100 pg of IL-1β per mL of organoid culture medium.

### Organoid culture

Healthy colon tissue was obtained endoscopically from the sigmoid colon from four healthy subjects. Demographic characteristics (age, gender and ethnicity) of the subjects providing tissue are present in Supplement Table 2. The collection and use of human colonic tissue was approved by the Institutional Review Board (IRBMED) at the University of Michigan and all subjects provided written informed consent prior to biopsy. This study was conducted according to the principles stated in the Declaration of Helsinki.

Briefly, cryopreserved organoid tissue samples were obtained from the Michigan Medicine Translational Tissue Modeling Laboratory (https://www.umichttml.org/) and expanded in Matrigel (Corning) over a 3-4 week period. During the expansion phase, culture medium consisted of a 1:1 mix of Advanced DMEM/F12 (Invitrogen) and the same medium that had been conditioned by the growth of L cells genetically modified to produce recombinant forms of Wnt3a, R-spondin-3 and Noggin (i.e., L-WRN) (34). The growth medium also contained 10% fetal bovine serum (Gibco) and the final calcium concentration was 1.0 mM. The medium was supplemented with 1X N2 (Invitrogen), 1X B-27 without vitamin A (Invitrogen), 1 mM N-Acetyl-L-cysteine (Sigma), 10 mM HEPES (Invitrogen), 2 mM Glutamax (Invitrogen), 100 μg/mL Primocin (InvivoGen) and small molecule inhibitors (10 μM Y27632 [Tocris]; as a ROCK inhibitor, 500 nM A83-01 [Tocris]; a TGF-β inhibitor, 10 μM SB202190 [Sigma]; a p38 inhibitor, along with 100 ng/mL EGF [R&D]). For the first 10 two days at each passage the medium was also supplemented with 2.5 µM CHIR99021 (Tocris). During this expansion phase, the colon organoids were passaged 3-6 times (in Matrigel) before undergoing experimental treatments.

During the experimental phase, established organoids were maintained in L-WRN plus 10 µM Y27632 (but without the additional small molecules) diluted 1:4 with KGM Gold. KGM-Gold is a serum-free, calcium-free culture medium optimized for epithelial cell growth (Lonza). The final serum concentration in the L-WRN – KGM Gold culture medium was 2.5% and the calcium concentration was 0.25 mM. This control treatment medium was compared to the same medium supplemented with the pro-inflammatory (LPS-cytokines) mix at a 1:250 dilution of the stock material. Control organoid cultures and those exposed to the pro-inflammatory mix were maintained without additional treatment or treated with Aquamin in an amount to provide a final calcium concentration of 3.0 mM or Mesalamine 200 µg/mL and combination of two treatments. Organoids were evaluated by phase-contrast microscopy (Hoffman Modulation Contrast - Olympus IX70 with a DP71 digital camera) for change in size and shape during the 14-day in-life portion of the study and at harvest. At harvest, organoids were prepared for proteomic assessment as described below.

### Proteomic assessment

Colon organoids were harvested from two wells per condition in a 6-well plate for proteomics, with each experiment conducted separately for each subject, as described in our previous reports (20,23,24). These organoids were isolated from Matrigel using 2 mM EDTA (E-5134; Sigma) in DPBS (14190-144; Gibco) for 15 minutes, followed by centrifugation at 100 × g and 4°C for 3 minutes, with three washes using DPBS. The organoids were then exposed to Radioimmunoprecipitation Assay (RIPA) lysis and extraction buffer (Pierce, #89901; ThermoFisher Scientific) for protein isolation at 4°C (or on ice) for 10 minutes, and centrifuged for 15 minutes at 14,000 × g and 4°C. Proteomic assessments were conducted by using mass spectrometry-based Tandem Mass Tagging system (TMT, ThermoFisher Scientific) in the Proteomics Resource Facility (PRF) housed in the Department of Pathology at the University of Michigan. For this, four subjects were assessed individually using TMT 18-plex kits (A52045; Thermo Scientific). The first experiment was conducted as part of a range-finding study with Mesalamine and compared to control medium (L-WRN – KGM Gold culture medium). Four subsequent proteomic experiments were conducted with complete sets of samples; control medium alone with supplementing Aquamin and Mesalamine individually and in combination. Parallel studies in the same subjects utilized addition of LPS-cytokines to control medium with and without Aquamin and Mesalamine, individually or in combination.

Fifty micrograms (at a concentration of 1µg/µl) of organoid protein from each condition was digested separately with trypsin and individual samples labeled with one of six isobaric mass tags according to the manufacturer’s protocol. After labeling, equal amounts of peptides from each condition were mixed together. In order to achieve in-depth characterization of the proteome, the labeled peptides were fractionated using 2D-LC (basic pH reverse-phase separation followed by acidic pH reverse phase) and analyzed on a high-resolution, tribrid mass spectrometer (Orbitrap Fusion Tribrid, ThermoFisher Scientific) using conditions optimized at the PRF. MultiNotch MS3 analysis was employed to obtain accurate quantitation of the identified proteins/peptides (35). Data analysis was performed using Proteome Discoverer (v3.0, ThermoFisher Scientific). MS2 spectra were searched against UniProt human protein database (20350 sequences; downloaded on 2023-03-01) using the following search parameters: MS1 and MS2 tolerance were set to 10 ppm and 0.6 Da, respectively; carbamidomethylation of cysteines (57.02146 Da) and TMT labeling of lysine and N-termini of peptides (304.207 Da) were considered static modifications; oxidation of methionine (15.9949 Da) and deamidation of asparagine and glutamine (0.98401 Da) were considered variable. Identified proteins and peptides were filtered to retain only those that passed ≤2% false discovery rate (FDR) threshold of detection. Quantitation was performed using high-quality MS3 spectra. Average signal-to-noise threshold was set to 8. Proteins names were retrieved using Uniprot.org. Only Proteins with a ≤2% FDR confidence of detection were included in the analyses. Differential protein expression profiling (for each subject separately) was established by considering the respective control (L-WRN – KGM Gold medium) as a comparator and evaluated results from the other conditions in relation to this control.

Next, resulting data from the four datasets were combined. The initial analysis was based on an unbiased proteome-wide screen of all proteins modified by Aquamin, Mesalamine and LPS-cytokines interventions in relation to the control using a cutoff of 1.5-fold. Follow-up analysis involved a targeted approach towards differentiation, barrier-related, cell-cell and cell adhesion proteins, mineral uptake and iron metabolism involving inflammation. QIAGEN Ingenuity Pathway Analysis (IPA) was used to find significantly altered canonical pathways by the participating proteins. IPA also provided predictions on the activated or inhibited status of these pathways. This commercial software is based on a knowledge database which can identify specific pathways and it can generate biological networks influenced by a given set of proteins and their observed expression. GraphPad Prism (v10.2) was utilized to generate heatmaps. Heatmaps were chosen for data visualization due to their ability to effectively condense complex datasets into a single figure. To identify common proteins and create Venn diagrams, we employed a web-based tool called InteractiVenn (interactivenn.net). The mass spectrometry proteomics data were deposited to the ProteomeXchange Consortium via the PRIDE partner repository (with the dataset identifier: pending) for open access.

### Statistical methods

Protein abundance ratios for individual proteins were obtained as calculated by the reporter ion quantifier in proteome discoverer by setting the control as denominator. Significance was calculated by the p-value based on t-test (background based). The adjusted p-value is by Benjamini-Hochberg method. For pathways enrichment analysis, IPA uses the Fisher’s Exact Test to calculate a statistical significance (p-value) of overlap of the dataset molecules with various sets of molecules that represent annotations such as Canonical Pathways. A p-value <0.05 was considered significant. IPA calculates the z-score by comparing observed expression to the expected expression in the knowledgebase, predicting up or down-regulation and weighted by the underlying findings.

## RESULTS

### Characteristics of organoids treated with Aquamin or Mesalamine: alone and in combination

After an initial series of dose-ranging experiments (see Materials and Methods for details; Supplement Figure 1), colon organoids from four subjects were incubated for a two-week period with subculture at the end of week-one. Incubation conditions included control culture medium alone or the LPS-cytokine mix in the same culture medium. Organoids from both groups were incubated without further treatment or concomitantly treated with either Aquamin (providing 3 mM calcium) or Mesalamine (200µg/mL) alone or with the two interventions in combination. At the beginning of the treatment phase and immediately after the initial subculture, organoid fragments were approximately 40 µm in size on average. At the time of harvest (after the first week and at the end of the 14-day culture period), individual organoids had increased to an average size of approximately 500 µm in diameter. Morphological features of organoids in all eight treatment groups are shown in Figure 1. All eight treatment groups were similar in appearance; cultures contained organoids with more differentiated features (thick-walled, round or oval-shaped, single lobed or multi-lobed structures) and a few with cystic appearance. The organoid growth characteristics and morphological features shown in Figure 1 are similar to what has been described previously (20,23,24).

**Figure 1.**
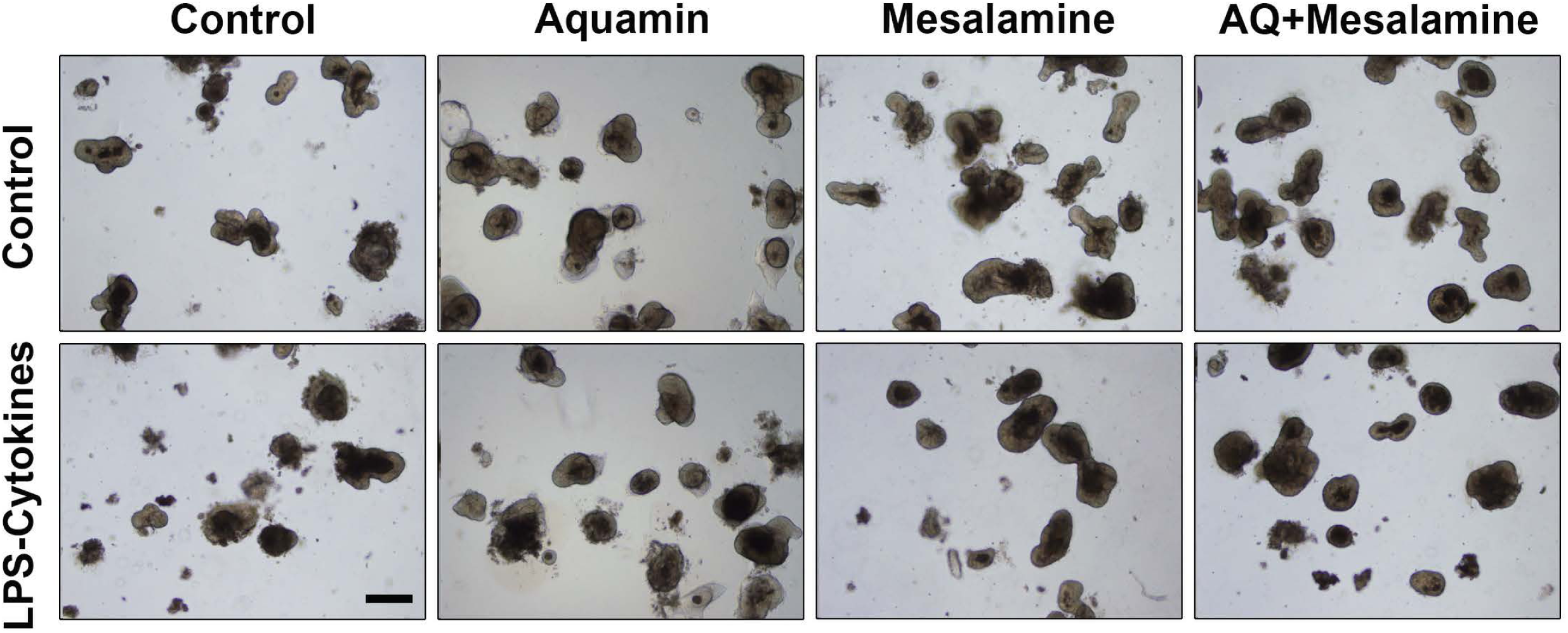
Human colon organoid appearance assessed by phase-contrast microscopy. At the end of the incubation period, intact colon organoids were examined by phase-contrast microscopy. Organoids were present as thick-walled structures with few surface buds. A wide range of sizes and shapes were seen under all conditions. Scale bar=500µm.

### Effects of intervention on protein expression profile: Unbiased assessment

Lysates were prepared from each of the eight treatment groups and the protein signature was assessed using the TMT-mass spectrometry – based proteomic approach as described in the Materials and Methods Section. As noted above, organoids established from each individual tissue donor were assessed separately with findings from the four subjects merged at the end. Figure 2 summarizes findings from the unbiased search portion of the study. Panel A of Figure 2 shows the number of proteins up-regulated and down-regulated by Aquamin alone, Mesalamine alone and the combination of the two interventions compared to control at three different fold-change levels (1.2-, 1.5- and 2-fold difference). The Venn plots show overlap among the three treatment groups at the 1.5-fold change level. As regards up-regulated proteins, a substantial number of proteins were induced by each intervention separately (138 with Aquamin and 324 with Mesalamine at 1.5-fold). However, only 51 of these proteins were present in both treatment groups. When the two interventions were present together throughout the 14-day incubation period, a total of 371 proteins were up-regulated but only 41 proteins were common to all three-treatment group. The conclusion from these experiments is that while Aquamin and Mesalamine each up-regulated multiple proteins, the groups of proteins up-regulated in response to the two interventions were largely independent of one another.

**Figure 2.**
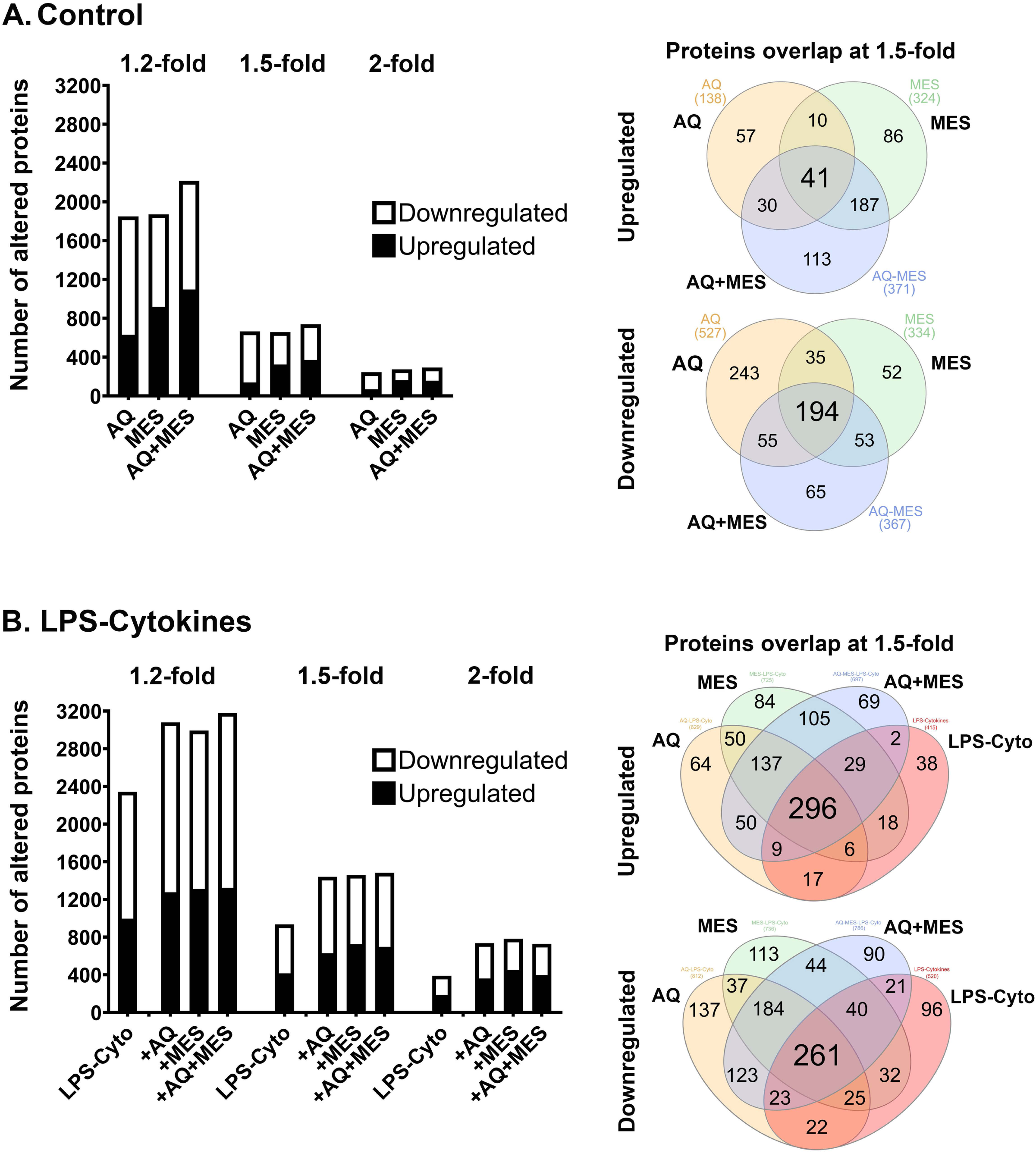
Proteomic profile of human colon organoids treated with Aquamin, Mesalamine and LPS-cytokines: Unbiased assessment. At the end of the incubation period, lysates were subjected to TMT mass spectroscopy-based proteomic analysis. **A**: <u>Control conditions</u>: Aquamin alone, Mesalamine alone and Aquamin plus Mesalamine compared to control. <u>Left</u>: Bar graph showing the number of proteins up-regulated and down-regulated at 1.2-fold, 1.5-fold and 2.0-fold change from control with <2% FDR. <u>Right</u>: Venn plots showing proteins altered (increased or decreased) by an average of 1.5-fold or greater compared to control. **B**: <u>LPS-cytokine challenged</u>: LPS-cytokines alone, LPS-cytokines plus Aquamin, LPS-cytokines plus Mesalamine, and LPS-cytokine plus Aquamin and Mesalamine compared to control. <u>Left</u>: Bar graph showing the number of proteins up-regulated and down-regulated at 1.2-fold, 1.5-fold and 2.0-fold change from control with <2% FDR. <u>Right</u>: Venn plots showing proteins altered (increased or decreased) by an average of 1.5-fold or greater compared to control. The data are based on all unique proteins identified across organoid cultures from four subjects. Organoid specimens from each subject were subjected to TMT-mass spectronomy – based proteomic analysis separately with data from the four merged at the end.

Down-regulated proteins demonstrated a similar trend in that a large number of proteins were responsive to each intervention (Figure 2A). While there was a larger overlap between the two interventions than was seen with up-regulated proteins, the majority of affected proteins were intervention-specific. Supplement Tables 3 and 4 are lists of the proteins making up each of the Venn plot domains. Protein distribution under control conditions shown by fold-change and p-value is included in the accompanying volcano plots (Supplement Figure 3A-C).

Figure 2B presents findings from organoid cultures in which the LPS-cytokine mix was included alone as an intervention as well as when it was present in combination with Aquamin and/or Mesalamine. Clearly evident is that the pro-inflammatory stimulus by itself had a profound effect on protein signature. Both increases and decreases were seen. How the profile of LPS-cytokines – induced protein changes was modified by Aquamin and Mesalamine can also be seen in Figure 2B. While some of the protein changes driven by the pro-inflammatory mix were blunted, most were not. In fact, there was a substantial overlap between proteins up-regulated by the pro-inflammatory mix and proteins responsive to Mesalamine. Supplement Tables 5 and 6 are lists of the proteins making up each of the Venn plot domains. Protein distribution under pro-inflammatory challenge shown by fold-change and p-value is included in the accompanying volcano plots (Supplement Figure 3D-G). Not surprisingly, many of the proteins responsive to treatment with LPS and cytokines are ones already well-known to be associated with the pro-inflammatory state (as we have seen in our previous study; 23). The following sections present results of a directed search for protein changes relevant to barrier formation in the colon and changes thought to impact the inflammatory process directly.

### Proteomic changes with intervention: Effects of Aquamin and Mesalamine (alone and in combination) on proteins contributing to barrier integrity

The same tissue lysates used in the unbiased assessment were interrogated specifically for proteins related to barrier formation in the colon. We used findings from our earlier organoid culture studies with Aquamin (20-24) as well as from our two interventional trials with the same agent (25,26) to guide the search.

Consistent with findings from our previous studies in colon organoids, Aquamin had little effect on expression of most proteins associated with tight junctions (Figure 3A). In contrast (but also consistent with past findings), cadherins/adherens junction proteins (cadherin-17, protocadherin-1, and cadherin-3) were strongly up-regulated with Aquamin, as were desmosomal proteins (desmoglein-2 and desmocollin-2) (Figure 3A). While the present study utilized a proteomic approach to obtain a broad survey of protein changes, our previous efforts made use of immunohistology, confocal immunofluorescence microscopy, western blotting and quantitative electron microscopy to validate proteomic findings related to these same barrier proteins (specifically, occludin, cadherin 17 and desmoglein 2) (20-24). Furthermore, functional assays (colon organoid cohesion and transepithelial electrical resistance) were used to establish biological relevance of changes observed in the proteomic screen (21-23). Of direct relevance to the present study, treatment with Mesalamine alone had no effect on these same cell-cell adhesion molecules. Equally important, Aquamin-responsive barrier protein changes were unaffected by co-incubation with Mesalamine (Figure 3A). Perhaps unexpectedly, increased expression of four proteins (CAD13, PCDH12, PCDHGC3 and DSG3) was seen with Mesalamine but not Aquamin. At this point, we do not know the significance of this, but it should be noted that none of these is recognized as being a significant contributor to the barrier in the gastrointestinal tract (36-39).

**Figure 3.**
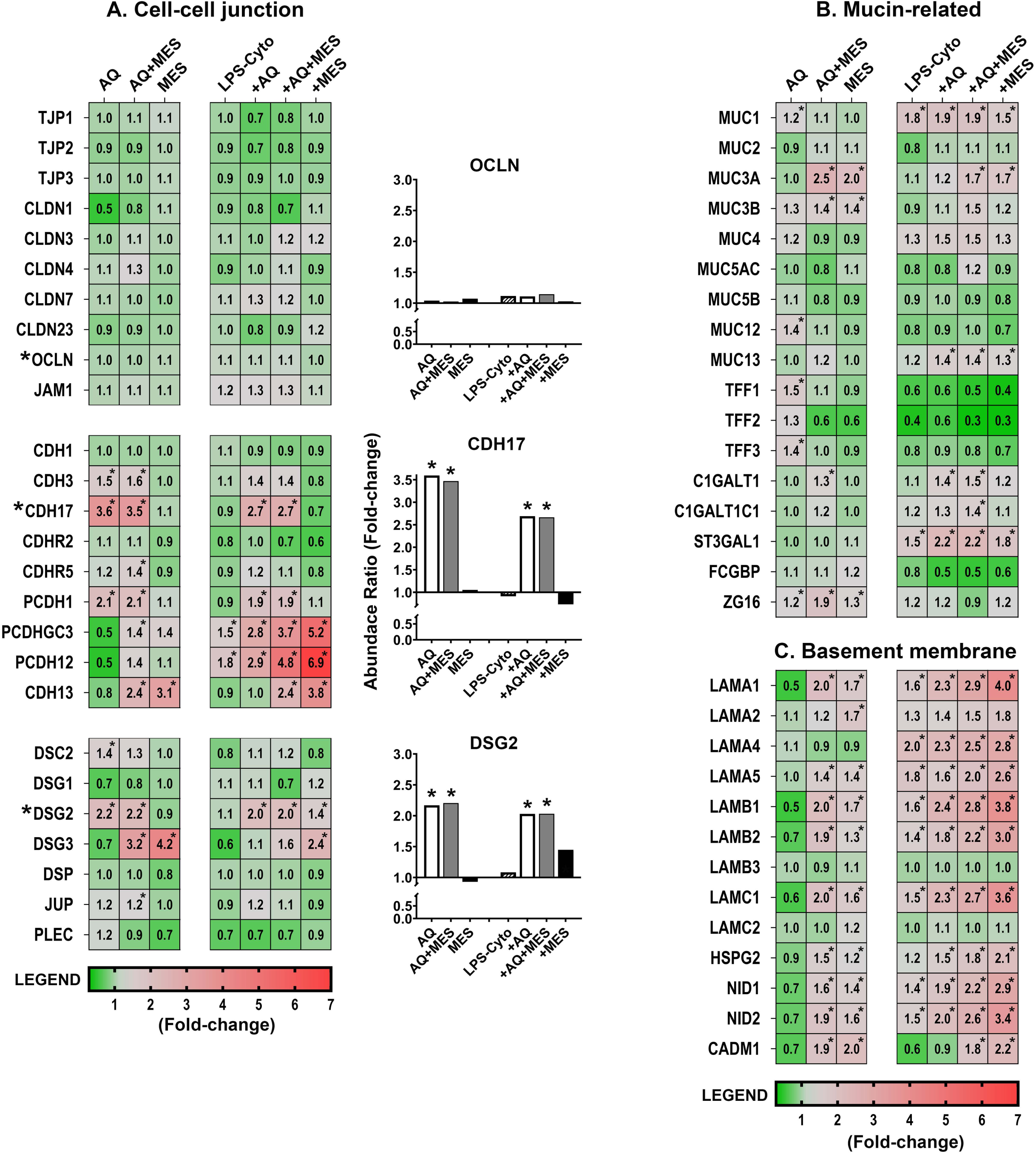
Heatmaps showing proteomic profile of human colon organoids treated with Aquamin, Mesalamine and LPS-cytokines: Mucosal Barrier proteins. Values represent average abundance ratio from treated organoids as compared to untreated (control) organoids. Values are based on merged data from n=4 subjects. Asterisks (*) represent significantly altered proteins as compared to the control (p<0.05). A: Cell-cell adhesion molecules; B: Mucin / trefoil related proteins; C: Basement membrane-related proteins. Protein designations follow their gene IDs as provided by Uniprot. Supplement Table 7A provides an index to complete protein names. Bar graph inserts (occludin – OCLN, cadherin 17 – CDH17 and desmoglein 2 – DSG2) identify critical proteins previously validated by western blotting, confocal immunofluorescence microscopy, immunohistology and semi-quantitative electron microscopy (20,21,23,24).

As part of this study, a pro-inflammatory stimulus (i.e., a mix of LPS and three potent cytokines) was examined for effects on barrier protein expression. By itself, the pro-inflammatory stimulus had only a modest effect on the majority of the detected proteins including those critical to formation of cell-cell adhesion structures. Specifically, there was little change in expression (either down- or up- regulation) among tight junctional proteins, cadherins and desmosomal proteins (Figure 3A). Importantly, when the pro-inflammatory stimulus was present with Aquamin (alone or combined with Mesalamine), the effects were similar to those observed with Aquamin in the absence of the pro-inflammatory stimulus. That is, tight junction protein expression exhibited minimal change, but the cadherins and desmosomal proteins essential for tissue strength and cohesion were still substantially up-regulated, as we have demonstrated earlier (23).

Effects of Aquamin on proteins that contribute to the mucinous layer (muc proteins and trefoils) are shown in Figure 3B. While most of the detected muc proteins were not responsive to Aquamin, levels of the three trefoil proteins were increased (consistent with past observations [20,21,23,24]). Mesalamine, by itself, had little effect on most of the detected muc proteins (the exceptions being muc 3A and 3B which were up-regulated) and little effect on trefoil-1 and -3. Trefoil-2, however, was down-regulated by Mesalamine (Figure 3B). A search for moieties involved in muc protein synthesis/trafficking revealed several candidates. Most showed little response to Aquamin alone and only ZG16 (zymogen granule-16 [40]) responded strongly (up-regulated) with the combination of Aquamin and Mesalamine. The majority of the same muc proteins were largely unaffected by the LPS-cytokine stimulus. The only substantial changes observed with the pro-inflammatory stimulus alone were with Muc1 (1.8-fold up-regulation), as has been shown previously (23) and down-regulated of all three trefoils (Figure 3B). The concomitant presence of Aquamin and/or Mesalamine did not alter responses to the pro-inflammatory stimulus.

A number of basement membrane components were also detected in the proteomic screen. While there was little response to Aquamin alone, there was a substantial up-regulation of several basement membrane proteins (i.e., laminin subunits, nidogen-1 and -2 and heparin sulfate proteoglycan-2 [HSPG-2]) with Mesalamine alone and an even stronger induction of several of these moieties in the presence of the two interventions together (Figure 3C). The same trend was observed with CADM1, a syndecam family member that mediates a variety of cell-cell and cell-matrix interactions and is shown to enhance intestinal barrier function (Figure 3C) (41). Finally (and perhaps unexpectedly), most of the basement membrane proteins were increased in response to the LPS-cytokine stimulus by itself. Perhaps, even more unexpected, basement membrane protein levels increased with all three interventions and were the highest when all three interventions (i.e., Aquamin, Mesalamine and the LPS-cytokine mix) were present together (Figure 3C). Supplement Table 7A provides the complete names of the directed proteins designated by the gene symbols shown in Figure 3.

### Proteomic changes with intervention: Effects of Aquamin and Mesalamine (alone and in combination) on proteins associated with inflammation

We next utilized the same lysates to examine for proteins associated with inflammation – including those that may directly contribute to inflammatory tissue injury or those that could counter inflammatory damage. Among the most noteworthy findings seen with the two interventions in the absence of the LPS-cytokine mix were with components of the complement cascade and the clotting/fibrinolytic cascade (Figure 4A-B). Multiple proteins in both cascades were responsive, but changes induced by the two interventions separately were dramatically different from each other. Specifically, virtually all of the complement cascade proteins were down-regulated with Aquamin but strongly up-regulated with Mesalamine. Strong up-regulation was also observed with the combination treatment though some of the individual complement components were expressed at slightly lower levels with the combined treatment than was seen with Mesalamine alone (Figure 4A). A similar trend was observed with proteins belonging to the clotting/fibrinolytic cascade (Figure 4B). All three fibrinogen subunits and kininogen-1 were down-regulated in response to Aquamin, along with several other components of the cascade. These changes were not observed with Mesalamine alone. Quite the opposite, all three fibrinogen chains and kininogen, itself, were up-regulated in response to Mesalamine as were virtually all of the other clotting/fibrinolytic cascade moieties. When the two agents were present in concert, the protein signature, like that of the complement protein signature, was reflective, primarily, of Mesalamine. The major exception was the fibrinogen-a chain (FGA), which was down-regulated in the presence of the two interventions together as much as with Aquamin alone (Figure 4B).

**Figure 4.**
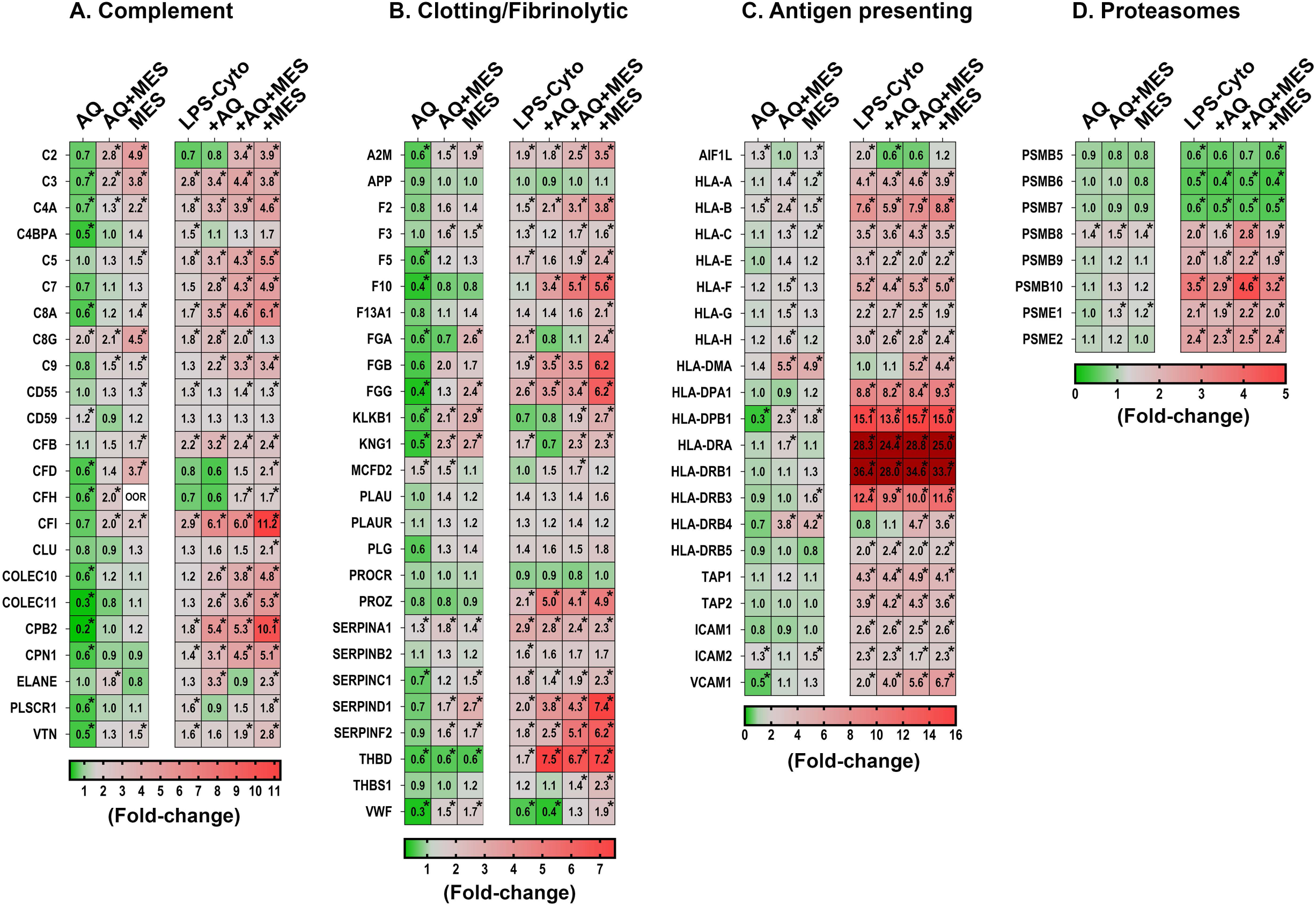
Heatmaps showing proteomic profile of human colon organoids treated with Aquamin, Mesalamine and LPS-cytokines: Inflammation-related proteins. Values represent average abundance ratio from treated organoids as compared to untreated (control) organoids. Values are based on merged data from n=4 subjects. Asterisks (*) represent significantly altered proteins as compared to the control (p<0.05). A: Complement cascade; B: Coagulation/fibrinolytic cascade; C: Antigen recognition and presentation D: Proteosome – immunoproteosome proteins. Protein designations follow their gene IDs as provided by Uniprot. Supplement Table 7B provides an index to complete protein names. Note: The expression levels of the two proteins, HLA-DRA and HLA-DRB1, under pro-inflammatory conditions were well above the maximum value on the legend scale.

Figure 4A and 4B demonstrate how the potent pro-inflammatory stimulus (alone) affected expression of the same complement and clotting/fibrinolytic cascade proteins and how this expression pattern was further altered by the concomitant presence of Aquamin and/or Mesalamine. It can be seen, first of all, that when the pro-inflammatory stimulus was present without the two interventions, most of the detected proteins in both cascades were up-regulated. It can be seen, furthermore, that when Aquamin was concomitantly present, the majority (though not all) of the changes induced by the LPS-cytokine stimulus were also up-regulated. The exceptions were the complement C2, CFD and CFH proteins (important components of the alternative pathway) and fibrinogen-a chain. The strong down-regulation of these moieties observed with Aquamin in the absence of the pro-inflammatory stimulus was maintained. When Mesalamine (alone or in combination with Aquamin) was present in conjunction with the pro-inflammatory stimulus, expression of the majority of both complement and clotting/fibrinolytic cascade proteins remained high; in some cases, higher levels were seen than was the case with either the pro-inflammatory stimulus alone or Mesalamine alone (Figure 4A-B).

The mix of LPS and the three pro-inflammatory cytokines provided a powerful stimulus that up- or down-regulated numerous additional proteins beyond those involved in the complement and clotting/fibrinolytic cascades. Among the most LPS-cytokines responsive moieties were those involved in antigen recognition and processing. This included several HLA isoforms as well as certain other adhesion molecules that mediate interactions between inflammatory cells and the vascular wall (e.g., ICAM-1, -2, VCAM and TAP-1, -2). With the majority of these proteins, Aquamin and Mesalamine had only modest effects by themselves, while concomitant exposure to Aquamin and/or Mesalamine along with the LPS-cytokine mix had little additional effect over what was observed with the pro-inflammatory stimulus alone (Figure 4C). One interesting exception was that allograft inhibitory factor-1 (AIF1L) was up-regulated with LPS-cytokines, but significantly down-regulated with Aquamin (both with and without Mesalamine) in the presence of a pro-inflammatory stimulus. Higher levels of AIF1L are associated with intestinal inflammation (42).

Along with proteins that affect the process of antigen recognition and processing directly, we also detected forty-seven proteins that contribute to proteosome-mediated protein degradation. Figure 4D presents the eight moieties from this group that were the most-highly responsive to the LPS-cytokine combination. As can be seen, subunits that are part of the classical proteosome (PSMB-5, -6 and -7) were all significantly down-regulated, while moieties contributing to the immunoproteosome (PSMB-8, -9 and -10 as well as PSME-1 and -2) showed strong up-regulation. Neither Aquamin nor Mesalamine (alone or in combination) affected the levels of the various proteosome components in the absence of the LPS-cytokine stimulus, and neither intervention substantially altered the response to the pro-inflammatory mix. Supplement Table 7B provides the complete names of the directed proteins designated by the gene symbols shown in Figure 4.

### Effects of Aquamin and Mesalamine (alone and in combination) on proteins involved in mineral uptake and iron-dependent reactions

Finally, the proteomic database was searched for proteins involved in metal ion transport and, in particular, for proteins contributing to iron-dependent reactions. Several potentially important proteins were identified. Proteins that regulate uptake and intracellular storage of zinc and copper (i.e., members of the SLC30 and SLC39 families [zinc] and SLC31 family [copper] were prominent. These same proteins along with others identified in the screen can also bind and help regulate levels of other trace elements including cobalt, magnesium, manganese, molybdenum, and iron. Several of these proteins proved to be responsive to the multimineral intervention, though the degree of responsiveness was modest (Figure 5A). Mesalamine alone also had a modest effect on these proteins as did the combination of Aquamin and Mesalamine. The one exception was SLC39A5 which was strongly down-regulated by Mesalamine (alone and in combination with Aquamin). Finally, two metallothionine-1 isoforms (MT1E and MT1H) were identified in the database. In contrast to what was observed with members of the solute carrier superfamily, both MT1E and MT1H were down-regulated with Aquamin alone. Mesalamine, in contrast, modestly increased MT1E expression but had no effect on MT1H (Figure 5A). The LPS-cytokine mix had no significant effect on levels of either protein when present alone and did not interfere with down-regulation of MT1H by Aquamin.

**Figure 5.**
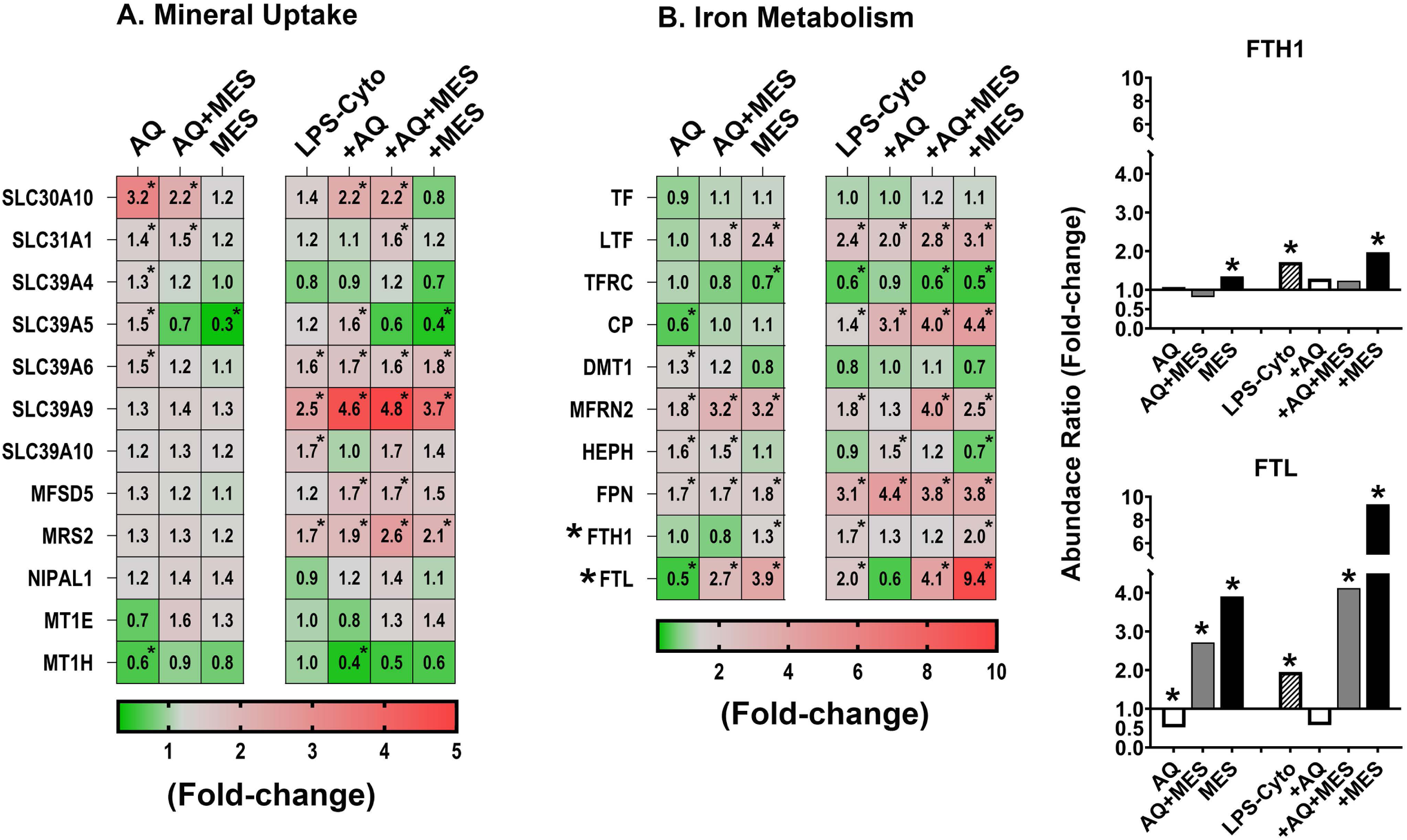
Heatmaps showing proteomic profile of human colon organoids treated with Aquamin, Mesalamine and LPS-cytokines: Metal ion transport and iron-related proteins. Values represent average abundance ratio from treated organoids as compared to untreated (control) organoids. Values are based on merged data from n=4 subjects. Asterisks (*) represent significantly altered proteins as compared to the control A: Metal ion transport; B: Iron transport and binding. Protein designations follow their gene IDs as provided by Uniprot. Supplement Table 7C provides an index to complete protein names.

Figure 5B identifies proteins that directly affect iron uptake, storage and participation in critical cellular reactions. Iron-dependent oxidative cell injury is well-established as a major contributor to tissue damage in inflammation (43). At the same time, dysregulation of iron-uptake contributes to colon cancer incidence and progression (44). Molecules that transport iron in the extracellular space (serotransferrin [TF] and lactotransferrin [LTF] as well as the major transferrin receptor [TFRC]) were all detected but none of the three was significantly affected by Aquamin alone (Figure 5B). Ceruloplasmin (CP), another plasma protein that transports iron (as well as copper) extracellularly, was detected and this moiety was significantly down-regulated with Aquamin. Although none of the proteins that carry iron in the circulation were up-regulated in response to Aquamin, four membrane-associated iron transporters – i.e., haephestin (HEPH), ferroportin (FPN), dimethyl transporter-1 (DMT-1) and mitoferrin-2 (MFRN2) – were up-regulated (Figure 5B). Finally, both isoforms of ferritin, the major cellular iron storage protein, were also detected. The heavy chain (FTH1) showed no response to Aquamin but ferritin light chain (FTL) was reduced in Aquamin-treated organoids.

How these same ten iron-regulating proteins responded to Mesalamine and to LPS-cytokine stimulation is also shown in Figure 5B. Neither the profile of Mesalamine-sensitive proteins nor that of the LPS-cytokine mix resembled the profile of proteins responsive to Aquamin. However, protein changes induced by Mesalamine and those induced by the pro-inflammatory stimulus mimicked one another closely. Of particular note, both the pro-inflammatory stimulus and Mesalamine strongly up-regulated both ferritin isoforms. Supplement Table 7C provides the complete names of the directed proteins designated by the gene symbols shown in Figure 5.

### Pathways altered with intervention: Effects of Aquamin and Mesalamine (alone and in combination)

As a final step in the analysis, we utilized Qiagen IPA software to search for the most highly-enriched pathways influenced by the proteins identified in Figures 4 and 5. Shown in Figure 6 are pathways that met the criteria of a -4 to +5 activation z-score. Not surprisingly, the majority of the identified pathways were related to the inflammatory state – either as contributors to inflammatory pathophysiology or as part of the host response. The z-score activation values provide a comparison among the different treatment groups for effects on critical pathways. Evident from the figure is the dichotomy between Aquamin and Mesalamine in their effects on the inflammation-related pathways under control conditions. Also evident is that when the two agents were provided together, effects on critical pathways reflected the action of Mesalamine more than that of Aquamin. Finally, it is clear from Figure 6 that when the pro-inflammatory stimulus was present, it was sufficient by itself to strongly modulate (either up or down) the majority of the pathways. As was observed with individual proteins, the effects of LPS-cytokine stimulation were similar to, though generally stronger than, effects seen with mesalamine alone. In contrast, with the majority of affected pathways, the actions of Aquamin and LPS-cytokines were distinct.

**Figure 6.**
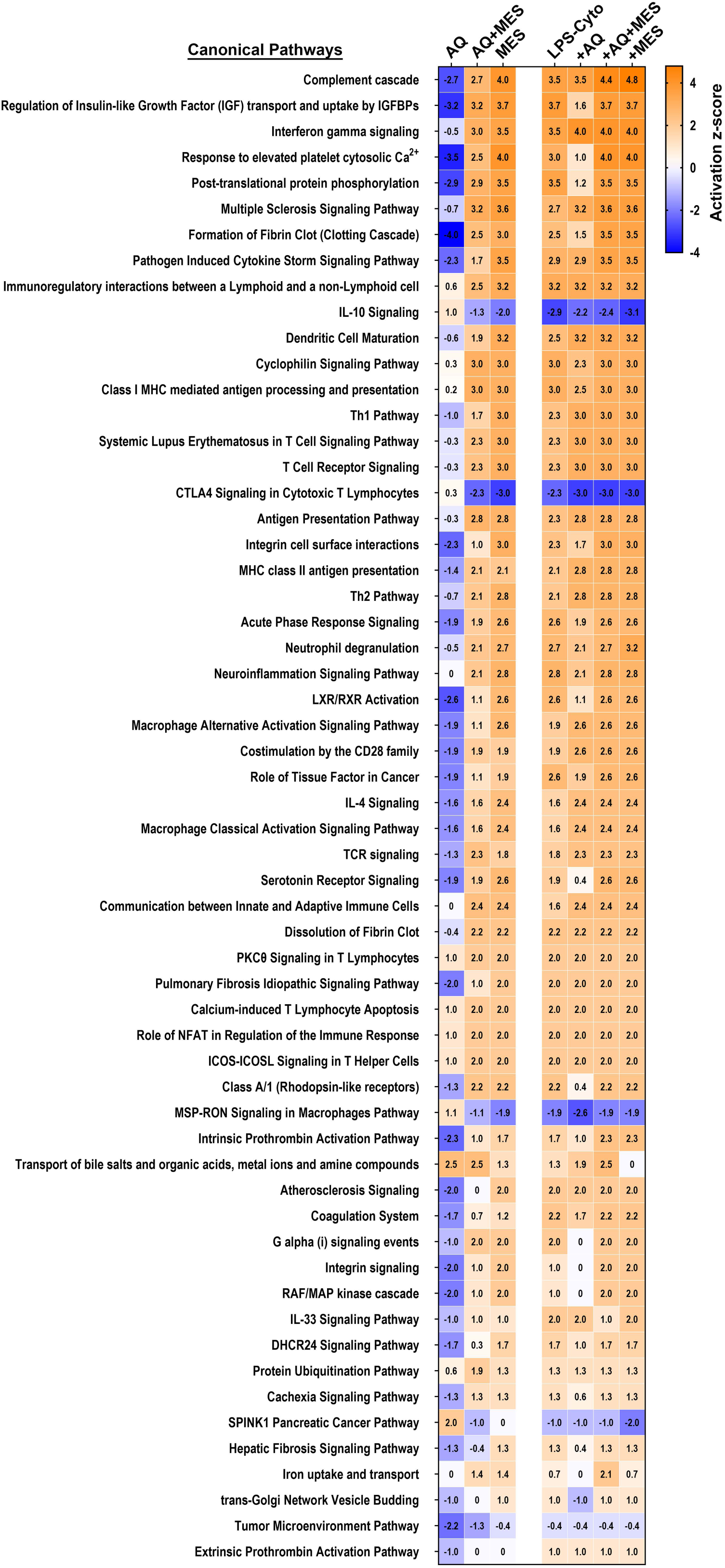
Heatmaps showing pathways affected in human colon organoids in response to treatment with Aquamin, Mesalamine and LPS-cytokines. The top canonical pathways affected by proteins modified as a result of the interventions were identified. These pathways were curated using QIAGEN Ingenuity^®^ Pathway Analysis (IPA) and ranked according to their z-score activation.

Finally, the IPA database was used to search for networks predicted to be altered by the proteins responsive to each intervention. Supplement Figure 4 shows one of the top networks, “Inflammatory Disease and Organismal Injury.” Supplement Table 8 identifies participating molecules and shows their expression levels and relationship types. The network data confirm the unique expression pattern of the interacting proteins influenced by Aquamin compared to each of the other interventions (Supplement Figure 4). Proteins involved in immune response (like HLA-DRA and HLA-DRB5) show significant changes, reflecting the impact of cytokine treatments, possibly related to immune cell activation. Aquamin and Mesalamine are largely independent of one another in terms of anti-inflammatory mechanisms, and support our view that Aquamin primarily works to improve the barrier.

## DISCUSSION

The present study is part of our effort to determine if Aquamin may be useful as a treatment for colonic barrier improvement in individuals with UC. Previous studies in mice have demonstrated a reduction in gastrointestinal (and systemic) inflammation when Aquamin was included in the diet (27-29,45,46). Subsequent studies with human colon tissue in organoid culture found that Aquamin was able to up-regulate proteins that contribute to barrier formation (20-24). Improved barrier protein expression was associated with increased organoid cohesion and with increased trans-epithelial electrical resistance. Based on those findings we conducted two interventional trials – one in healthy human subjects (ClinicalTrials.gov ID: NCT02647671) (25) and the other involving individuals with mild UC or UC in remission (ClinicalTrials.gov ID: NCT03869905) (26). In both trials we saw increases in the same barrier proteins as seen in organoid culture. In neither trial were any safety or tolerability issues reported.

The participants who enrolled in the UC trial were maintained on their physician-prescribed standard-of-care UC treatment as part of the protocol. Mesalamine is the most frequently included drug in the maintenance regimen for individuals with UC in remission or at the mild-to-moderate stage (6,47), and that was the case in our study. Given the common use of Mesalamine as a maintenance therapy in UC, it would be of value to know if and to what extent Aquamin and Mesalamine might complement one another or if, in fact, there could be significant synergy between the two. Whether interference might occur would also be important to know. As a first step in addressing these questions, we utilized human colon organoid cultures to assess the effects of Aquamin and Mesalamine alone and in combination (under control conditions and with pro-inflammatory stimulus) on protein expression patterns in the tissue.

The data presented here (both in the non-biased approach [Figure 2 and Supplement Tables 3-6] and in the directed searches [Figures 3-5]) demonstrated that both Aquamin and Mesalamine up-regulate numerous proteins in colon organoid culture. The signatures of the two interventions are, not surprisingly, very different. By itself, Aquamin up-regulated cell-cell adhesion molecules (adherens junction and desmosomal proteins) that are important for tissue integrity and strength in the colonic epithelium. This activity was largely unaffected by the concomitant presence of Mesalamine in the treatment medium (Figure 3). Of interest, while Mesalamine did not affect Aquamin’s ability to up-regulate the adherens junction and desmosomal proteins, there appeared to be an additive effect on components of the basement membrane. These proteins were consistently higher in the presence of the two interventions together than seen with either one separately (Figure 3). Our conclusion from these studies is that there is no evidence to suggest that the concomitant presence of Mesalamine would prevent Aquamin from acting as a treatment for barrier improvement in UC. Quite the contrary, the effects of the two interventions together on basement membrane component expression could suggest room for a synergistic (or at least additive) improvement. Of interest, a recent study has suggested that a direct impact on the colonic barrier might also occur with Mesalamine use (48). While the data presented here and those reported in the previous publication are quite different, both suggest a potentially novel mechanism to explain Mesalamine’s beneficial activity in UC. Additional work will be needed to determine the extent to which this novel hypothesis is valid.

As part of the study protocol, Aquamin and Mesalamine were examined for effects on protein expression in the presence of a strong pro-inflammatory stimulus. We have previously reported that the same potent pro-inflammatory challenge as used here was able to disrupt barrier function in human colon organoids and colon organoid-derived epithelial cells in monolayer culture. At the same time, however, the combination of LPS and pro-inflammatory cytokines did not prevent barrier protein up-regulation with Aquamin alone and did not suppress the improvement in barrier function associated with barrier protein up-regulation (23). In the current study, neither the pro-inflammatory stimulus alone nor the combination of the stimulus and Mesalamine altered the ability of Aquamin to up-regulate barrier protein expression.

While the primary goal of the present study was to determine if the concomitant presence of Mesalamine might disrupt Aquamin’s effect on barrier protein expression, the approach also allowed us to assess changes in a wide range of proteins that may contribute to beneficial activity with either Aquamin or Mesalamine. The response to Mesalamine was of particular interest since this agent has not been assessed previously in this manner. Mesalamine is widely used as maintenance therapy for patents with UC (6,49,50) but the exact mechanism(s) of action is/are still not fully understood. Most of the proposed mechanisms relate to interference with one aspect or another of the inflammatory response. Mesalamine is structurally related to aspirin, and reducing the generation of pro-inflammatory metabolites from arachidonic acid metabolism is a widely-accepted possibility (51,52). Mesalamine also has the capacity to interfere with oxidant injury by limiting free radical production (53) and/or blocking radical-induced lipid peroxidation (53-55). Other studies have shown that Mesalamine can interfere with cytokine generation, possibly by targeting NF-kB – mediated transcription of cytokine genes (56) or by blocking signaling pathways that influence NF-kB (57,58).

While these mechanisms, undoubtedly, contribute to Mesalamine’s effectiveness in UC, the proteomic screen identified changes in other proteins / protein families that may be relevant. For example, we saw up-regulation in the expression of two groups of proteins – i.e., members of the complement cascade and members of the clotting/fibrinolytic cascade – that have not been noted previously. Complement activation (59,60) and activation of the clotting/fibrinolytic cascade (61,62) occur during inflammation. Both cascades are part of the host innate response to infection and inflammation; when fully activated, they contribute to tissue damage.

The complement cascade, in particular, functions to bring inflammatory cells to sites of bacterial infection and to kill and remove (through membrane lysis or through opsonization) invasive pathogens from the host. While the liver is thought to be the main contributor to circulating complement proteins, gastrointestinal cells also elaborate complement components (63-65). Local production in the gastrointestinal tract is thought to be part of the mechanism keeping colonic bacteria from penetrating the colonic wall and initiating an inflammatory response (66,67). Thus, the complement cascade functions as a key part of the normal surveillance system. However, when excessive stimulation occurs – e.g., during an ongoing inflammatory response – tissue damage is the result. Most studies suggest that in UC, a higher level of complement activation is associated with more severe inflammation (63,68). In murine models of acute colonic inflammation, a higher level of complement activation and greater tissue damage is also correlated. In these mouse models, therapeutic interventions to reduce activation can lead to a reduction in damage (69). At the same time, complement activation has been shown to be protective in chronic UC models (69-71).

Like the complement cascade, the clotting/fibrinolytic cascade plays multiple roles in inflammation that may be protective or contribute to tissue damage (61,62). Fibrinogen and fibrinogen-derived peptides as well as other components of the cascade (e.g., thrombin and plasminogen) mediate interactions between inflammatory cells and potentially harmful microorganisms to help eliminate the infectious agents at an early stage of infection (72,73). The same events can bring about leukocyte binding and damage to the vascular wall. Most importantly, of course, the clotting/fibrinolytic cascade’s major role is in hemostasis and tissue repair (72,73). The findings presented here do not allow us to distinguish between beneficial effects that may accrue with Mesalamine and those that are potentially damaging with either cascade. They do, however, suggest yet another mechanism through which Mesalamine may exert an effect in the gastrointestinal tract. Equally important, these findings also suggest no significant modulation of Mesalamine activity by concomitant presence of Aquamin.

Inflammatory tissue injury is driven, in part, through iron-dependent cell killing mechanisms (43,74,75). As part of the current study, the proteomic data base was searched for proteins that influence metal ion transport and iron-dependent reactions. While both Aquamin and Mesalamine altered the expression of several transporters, only a modest effect on most of the detected moieties was seen. In contrast, the ferritin light chain (FTL) isoform was down-regulated by Aquamin, while both the ferritin heavy and light chains were up-regulated in response to Mesalamine. The pro-inflammatory stimulus also increased the levels of both isoforms, with the highest expression levels seen with the combination of pro- and anti-inflammatory interventions. An increase in ferritin (especially the heavy chain) is well-known to occur with inflammation (76,77); this is assumed to be a cellular attempt to sequester iron and reduce the size of the labile pool. Since iron utilization by bacteria for growth and iron’s crucial role in generation of tissue-destructive oxygen radicals could both lead to tissue damage (77,78), controlling the size of the labile iron pool may serve the purpose of preventing both occurrences. Again, the data presented here do not provide for detailed mechanistic understanding but suggest yet one more possible way in which Mesalamine could function to counter inflammation in UC.

Finally, the proteomic screen identified substantial changes in proteins belonging to the antigen recognition and processing pathway (i.e., various class I and II MHC molecules, immune cell adhesion molecules and components of the immunoproteosome). While modest changes were induced in these proteins by Aquamin and Mesalamine, a much stronger induction occurred in response to the LPS-cytokine mix. When Aquamin and/or Mesalamine were included along with the pro-inflammatory stimulus, the changes seen in response to the pro-inflammatory stimulus alone were not affected further. Up-regulation of proteins involved in antigen recognition and processing occurs in both UC and Crohn’s Disease as well as in other inflammatory bowel conditions (79-81). This pathway is highly-sensitive to LPS and the individual cytokines included here (82). For better or worse, neither Aquamin nor Mesalamine (alone or in concert) would appear to impact this critical component of the host response to inflammation.

In summary, the primary goal of these studies was to identify potential interactions between Aquamin and Mesalamine that may occur if and when the two agents are used together as part of a maintenance regime in UC. Most importantly, the data presented here suggest that barrier protein induction by Aquamin would not be counteracted by the concomitant presence of Mesalamine. To the extent that cell-matrix interactions contribute to barrier function, the increase in basement membrane protein expression seen with combined treatment may suggest, ultimately, that improved barrier function could result from the presence of the two interventions together. Finally, the current studies found no evidence to suggest that the presence of Aquamin would interfere with the capacity of Mesalamine to alter the expression of proteins that are part of the anti-inflammatory shield. When the data from this study are considered *in toto*, they allow us to suggest that Aquamin and Mesalamine could, ultimately, prove useful together as part of a maintenance regime for individuals with UC. While this could be beneficial for all individuals with bowel inflammation, people with UC in remission or with mild-to-moderate disease are likely to benefit the most from the approach. The inclusion of Aquamin as part of a maintenance therapy along with Mesalamine could provide the first meaningful change in the treatment of these individuals since the introduction Mesalamine in the 1980s.

Beyond this goal, the TMT-mass spectronomy – based proteomic approach used here allowed us to survey a broad range of protein changes occurring in human colon organoid cultures exposed to Aquamin alone, Mesalamine alone or the two agents in combination. While we have utilized this approach previously to assess the effects of Aquamin on colon organoid protein signature, this study (to our knowledge) provides the first look at the broad range of protein changes occurring in human colon organoids in response to Mesalamine. The observations made here provide novel and intriguing clues to possible new mechanisms underlying Mesalamine’s action in colon inflammation.

## Supplemental Figures

**Supplement Figure 1. Assessment of colon organoid appearance in response to varying doses of Mesalamine using phase-contrast microscopy.** At the end of the incubation period, intact colon organoids were examined with phase-contrast microscopy to identify differences across a Mesalamine dose range of 50-250 µg. Under all conditions, a diverse range of sizes and shapes was observed. Scale bar=500µm.

**Supplement Figure 2. Heatmaps showing canonical pathways affected in human colon organoids in response to varying doses of Mesalamine**. The top canonical pathways impacted by the modified proteins in response to a Mesalamine dose range of 50-250 µg were identified. These pathways were curated using QIAGEN Ingenuity^®^ Pathway Analysis (IPA) and ranked according to their z-score activation.

**Supplement Figure 3. Distribution of proteins**. Proteins that are either up-regulated (red dots) or down-regulated (green dots) by 1.5-fold in response to various treatments under control and pro-inflammatory conditions. Grey dots represent proteins with changes of less than 1.5-fold. Panels A-C depict protein changes under control conditions with A) Aquamin, B) Aquamin and Mesalamine together, and C) Mesalamine alone. Panels D-G show protein changes under the influence of a pro-inflammatory stimulus (LPS-Cytokines), with each panel representing D) LPS-Cytokines alone, E) LPS-Cytokines with Aquamin, F) LPS-Cytokines with Mesalamine, and G) LPS-Cytokines with Aquamin and Mesalamine. Protein values in the control condition were normalized to 1.0, and the seven treatment group values were compared against this baseline. The x-axis displays the log2 fold-change of individual proteins, while the y-axis indicates the -log10 p-value (n=4 subjects).

**Supplement Figure 4. Qiagen IPA generated network analysis in response to treatment with Aquamin, Mesalamine and LPS-cytokines.** One of the top networks, presented in a subcellular layout, is related to "Cellular Movement, Inflammatory Disease, Organismal Injury, and Abnormalities" and includes 24 molecules from our dataset. Each network is capped at 35 molecules (the IPA default setting) for clarity. The likelihood of these molecules being part of the network is determined by a p-value. Networks were created from proteins altered by the interventions used here. The sidebar details confidence levels for predicted activities and interactions based on measured expression levels, using color-coding with different intensities to indicate expression and interaction differences. Supplement Table 8 presents interactions and relationship details.

## Supplemental Tables

**Supplement Table 1: Mineral Composition of Aquamin**

**Supplement Table 2: Demographic characteristics of tissue donors**

**Supplement Table 3: Protein identification**. Proteins that were up-regulated and common among the three interventions under control conditions were identified, using a 1.5-fold change and an ≤2%FDR, and sorted in each of the domains shown in the Venn plot in Figure 2A (Up-regulated proteins).

**Supplement Table 4: Protein identification**. Proteins that were down-regulated and common among the three interventions under control conditions were identified, using a 1.5-fold change and an ≤2%FDR, and sorted in each of the domains shown in the Venn plot in Figure 2A (Down-regulated proteins).

**Supplement Table 5: Protein identification**. Proteins that were up-regulated and common among the four interventions under pro-inflammatory conditions were identified, using a 1.5-fold change and an ≤2%FDR, and sorted in each of the domains shown in the Venn plot in Figure 2B (Up-regulated proteins).

**Supplement Table 6: Protein identification**. Proteins that were down-regulated and common among the four interventions under pro-inflammatory conditions were identified, using a 1.5-fold change and an <2%FDR, and sorted in each of the domains shown in the Venn plot in Figure 2B (Down-regulated proteins).

**Supplement Table 7: Complete names of proteins designated by gene symbols in Figures 3-5.**

**Supplement Table 8: Predicted Protein Interactions in the Network - Cellular Movement, Inflammatory Disease, Organismal Injury and Abnormalities**. A) Molecules involved; B) Their interactions and relationship between molecules.

## Data availability statement

The datasets presented in this study can be found as supplemental files or in an online repository. The mass spectrometry proteomics dataset will be available in the repository (ProteomeXchange Consortium - PRIDE partner repository), with the dataset identifier pending.

## Ethics statement

For the present 3D culture study, de-identified colon tissue was gathered during routine colonoscopies from healthy subjects or collected from deceased donors through the Gift of Life program. Tissue collection was performed under a protocol reviewed and approved by the Institutional Review Board at the University of Michigan (IRBMED). Patients’ protected health information was not shared with the investigators. The patients or participants signed a written informed consent document prior to the procedure, allowing the use of the colonic tissue for research purposes.

## Author contributions

All authors have contributed to this project and meet the ICMJE criteria for authorship. JV and MA were responsible for the study’s conception and design. SM conducted the majority of the organoid culture work. All authors participated in data acquisition, analysis, or interpretation. MA performed the statistical analysis. JV and MA composed the first draft of the manuscript. All authors contributed to the article and approved the submitted version.

## Acknowledgments

We thank the Michigan Medicine Translational Tissue Modeling Laboratory (TTML) for providing human colon organoid cultures and the Proteomics Resource Facility at the University of Michigan for assistance with proteomic data acquisition. We also acknowledge Marigot Ltd. (Cork, Ireland) for providing Aquamin^®^ as a gift.

