## Supplementary figures and images for "Proteomic Profile of Human Colon Organoids: Effects of a multi-mineral intervention alone and in the presence of pro-Inflammatory and anti-inflammatory treatments"

### S Figure 1

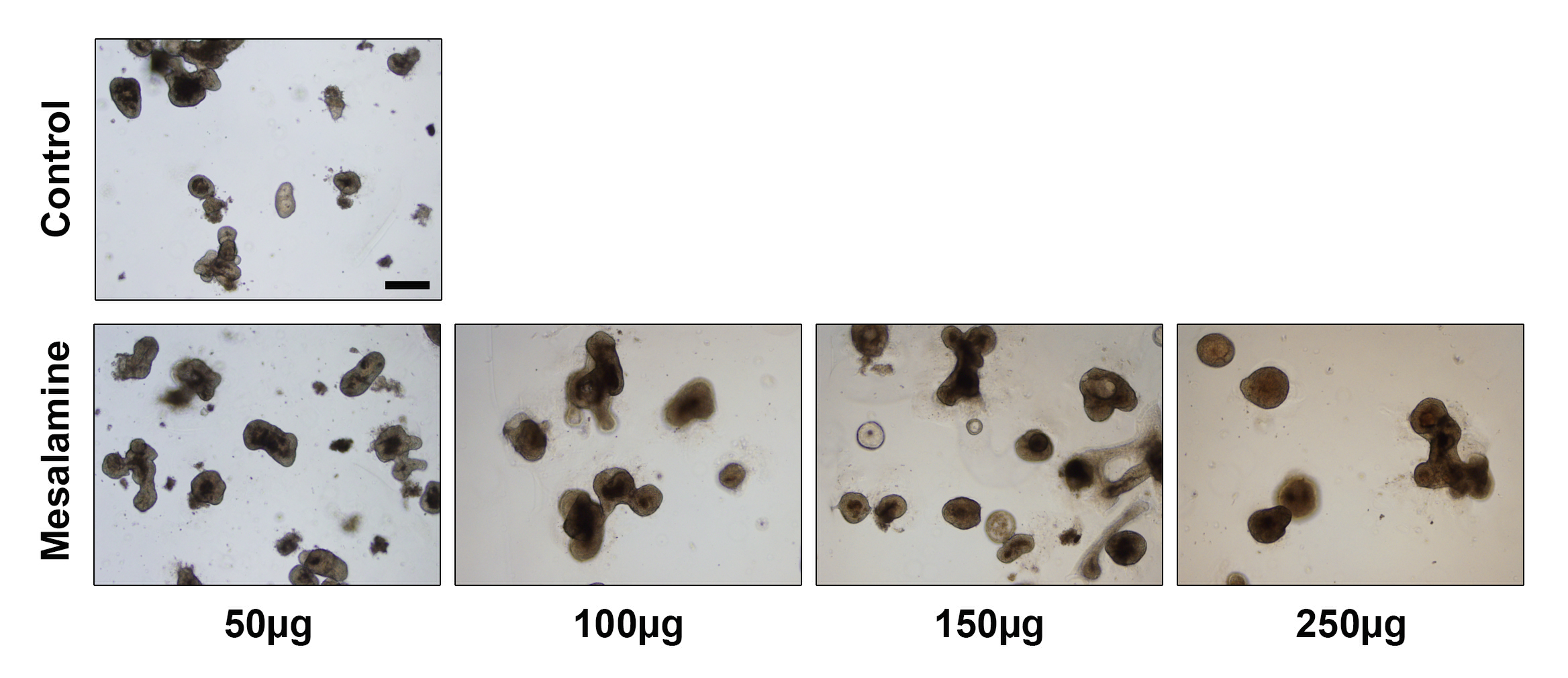

### S Figure 2

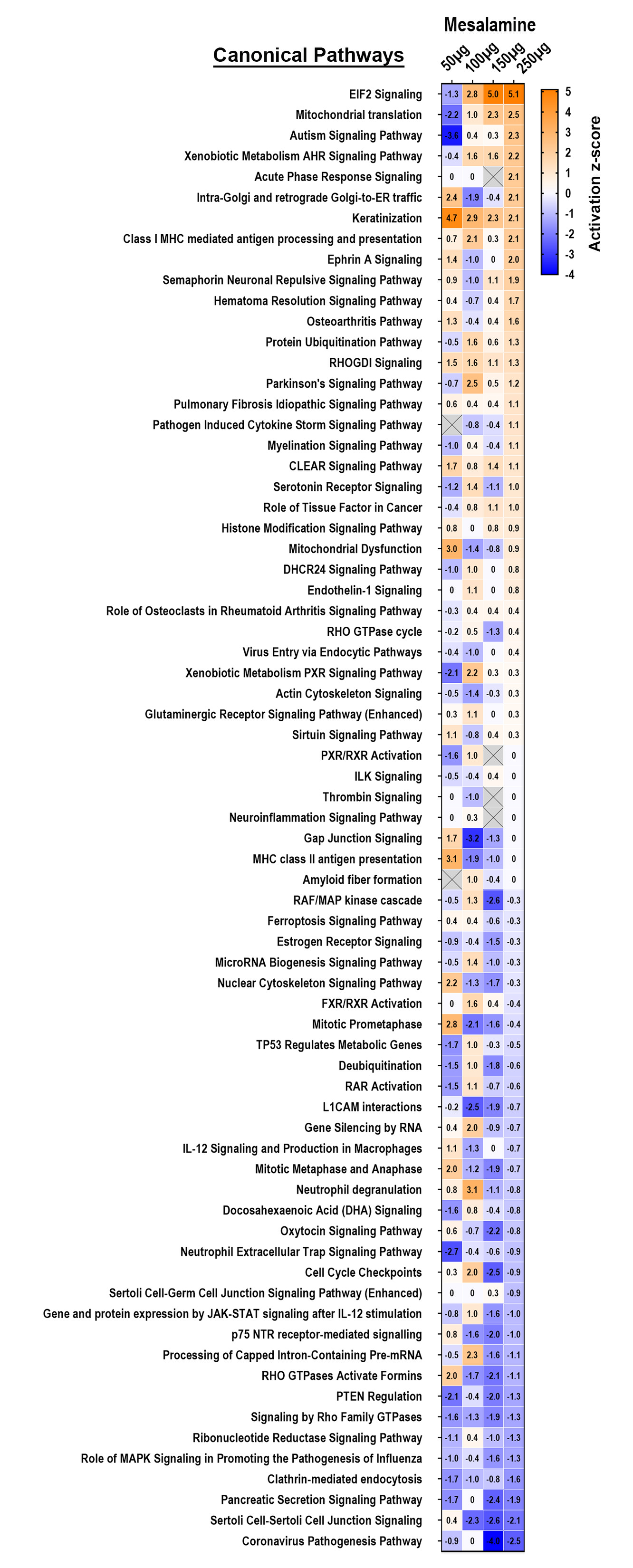

### S Figure 3

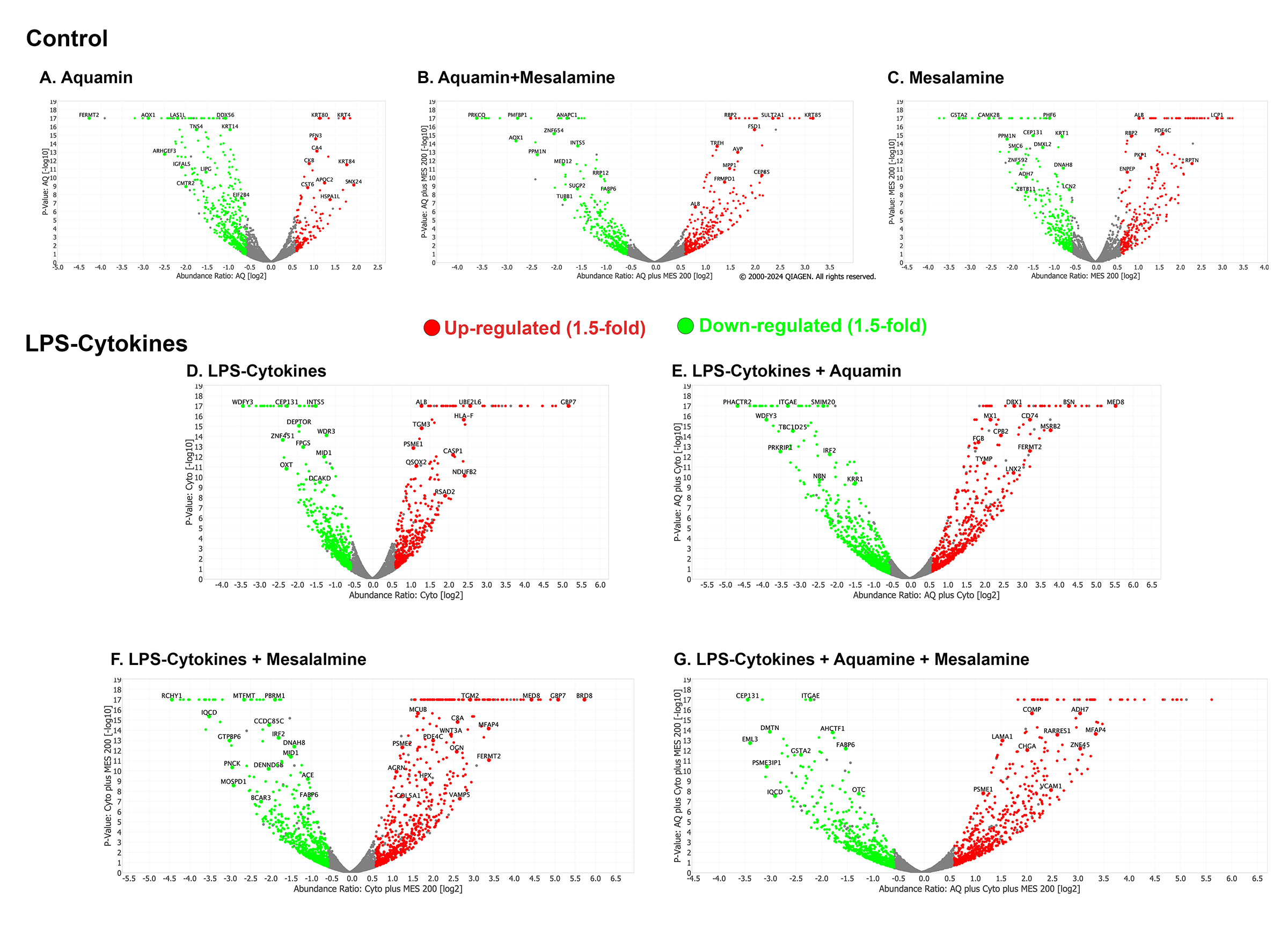

### S Figure 4

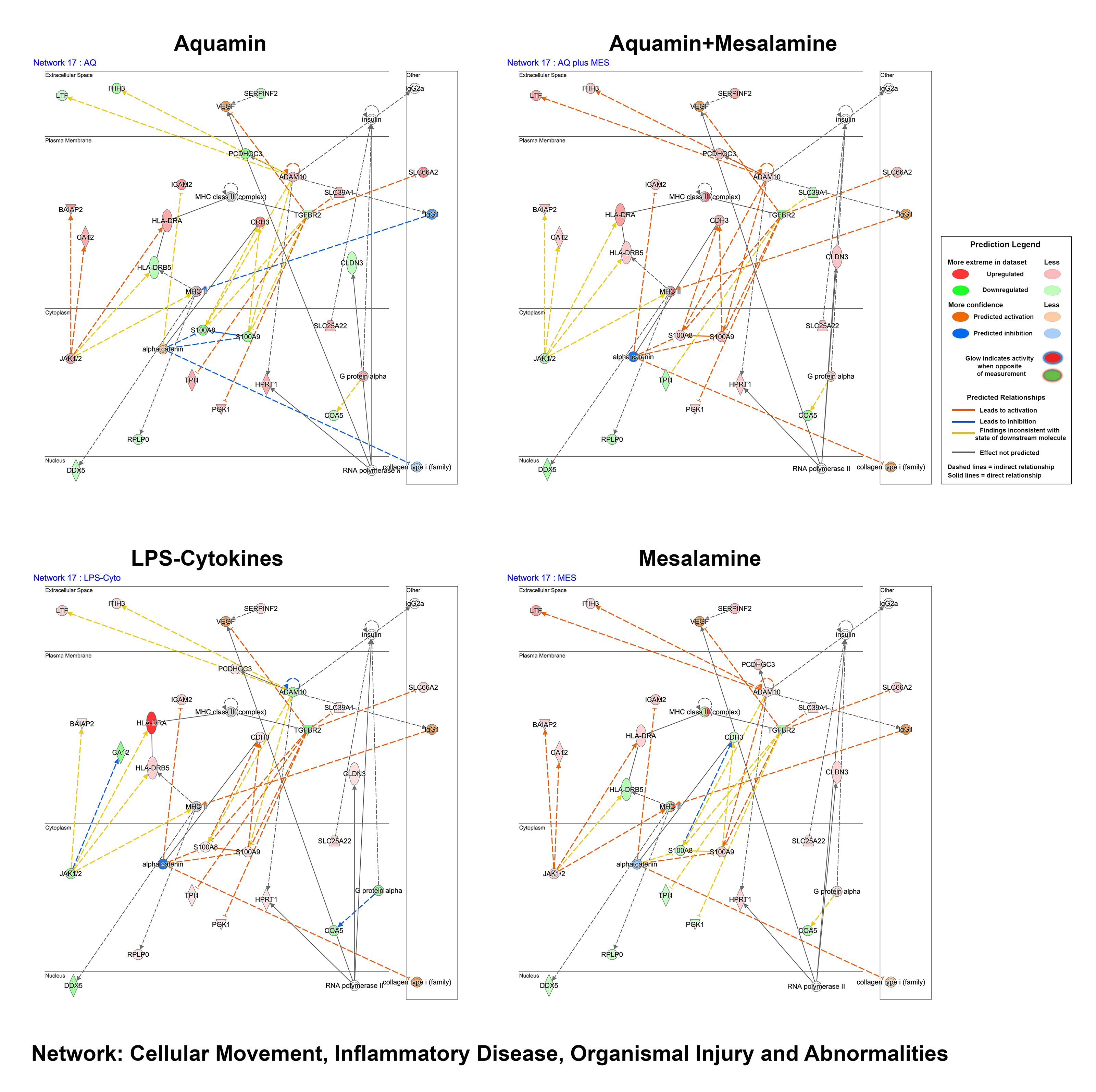
