## Supplementary material for "Proteomic Profile of Human Colon Organoids: Effects of a multi-mineral intervention alone and in the presence of pro-Inflammatory and anti-inflammatory treatments": S Table 2

**Supplement Table x. Demographic characteristics of tissue donors (subjects)**

| Sample ID | Age (Y) | Sex | Ethnicity | Biopsy Site |
|---|---|---|---|---|
| Colon-E1* | 46-50 | F | White (Not Hispanic) | Ascending colon |
| Colon-E7* | 21-25 | M | White (Not Hispanic) | Ascending colon |
| Colon-E8* | 31-35 | F | White (Not Hispanic) | Ascending colon |
| Colon-O5 | 61-65 | M | White (Not Hispanic) | Sigmoid colon |

*Source: Gift of Life, Michigan
