## Supplementary material for "Proteomic Profile of Human Colon Organoids: Effects of a multi-mineral intervention alone and in the presence of pro-Inflammatory and anti-inflammatory treatments": S Table 3

**Supplement Table 3. Up-regulated proteins influenced by Aquamin and Mesalamine under control conditions (with 1.5-fold).**

***A. Common among three groups – Aquamin (AQ), Mesalamine (MES) and Aquamin plus Mesalamine (AQ+MES) [41 proteins]***

| | | Interventions | | | | | | |
|---|---|---|---|---|---|---|---|---|
| | | Control | | | *With LPS & Cytokines* | | | |
| Proteins | Genes | **AQ** | **AQ+MES** | **MES** | *LPS-Cyto* | *AQ* | *AQ+MES* | *MES* |
| Sorting nexin-24 | SNX24 | 3.83* | 4.44* | 3.16* | 5.18* | 5.74* | 3.20* | 4.30* |
| Putative beta-actin-like protein 3 | POTEKP | 3.53* | 4.54* | 4.91* | 1.20 | 1.16 | 1.95* | 5.20* |
| FXYD domain-containing ion transport regulator 5 | FXYD5 | 3.37* | 3.09* | 2.13* | 2.24* | 5.59* | 4.94* | 4.29* |
| Keratin, type II cytoskeletal 4 | KRT4 | 3.27* | 1.56* | 9.03* | 0.70 | 10.27* | 1.13 | 6.70* |
| Transmembrane and immunoglobulin domain-containing protein 1 | TMIGD1 | 2.53* | 7.38* | 3.86* | 3.17* | 3.35* | 4.41* | 2.89* |
| Equilibrative nucleoside transporter 1 | SLC29A1 | 2.51* | 2.80* | 2.70* | 4.14* | 4.67* | 4.44* | 3.78* |
| Solute carrier family 35 member B1 | SLC35B1 | 2.33* | 2.96* | 3.17* | 1.47 | 2.58* | 2.91* | 2.01* |
| Keratin, type I cytoskeletal 23 | KRT23 | 2.31* | 1.81* | 6.31* | 0.44* | 2.65* | 0.70 | 2.04* |
| Triggering receptor expressed on myeloid cells 1 | TREM1 | 2.25* | 2.93* | 1.88* | 1.88* | 4.12* | 1.94* | 3.89* |
| Myeloid leukemia factor 2 | MLF2 | 2.19* | 1.97* | 3.37* | 2.90* | 2.56* | 1.98* | 2.36* |
| Small nuclear ribonucleoprotein F | SNRPF | 2.07* | 1.62* | 1.69* | 1.59* | 0.18* | 0.89 | 0.54 |
| NADH dehydrogenase [ubiquinone] 1 beta subcomplex subunit 2, mitochondrial | NDUFB2 | 2.06* | 1.92* | 3.85* | 5.33* | 9.24* | 8.41* | 5.38* |
| Spliceosome-associated protein CWC27 homolog | CWC27 | 2.02* | 1.81* | 1.94* | 1.33 | 0.51 | 0.99 | 2.11* |
| Nuclear receptor coactivator 6 | NCOA6 | 2.02* | 3.40* | 3.87* | 2.35* | 0.89 | 2.98* | 4.19* |
| Xaa-Pro aminopeptidase 2 | XPNPEP2 | 2.00* | 2.97* | 1.79* | 1.17 | 1.85* | 1.67* | 1.38 |
| Complement component C8 gamma chain | C8G | 1.96* | 2.13* | 4.51* | 1.81* | 2.79* | 1.96* | 1.26 |
| Transcription factor ETV6 | ETV6 | 1.90* | 1.88* | 2.40* | 1.54* | 1.45 | 2.02* | 1.81 |
| Prostatic acid phosphatase | ACP3 | 1.83* | 8.61* | 2.97* | 1.52* | 2.09* | 1.08 | 1.62 |
| Mitoferrin-2 | SLC25A28 | 1.83* | 3.25* | 3.19* | 1.78* | 1.29 | 3.96* | 2.50* |
| Cystatin-M | CST6 | 1.81* | 3.67* | 3.47* | 0.54* | 1.96* | 0.58 | 1.84* |
| Transmembrane protein 236 | TMEM236 | 1.79* | 4.00* | 2.65* | 1.34 | 1.43 | 2.59* | 1.66 |
| RING1 and YY1-binding protein | RYBP | 1.77* | 1.81* | 2.11* | 1.28 | 1.80 | 0.92 | 2.13* |
| Dynein axonemal heavy chain 17 | DNAH17 | 1.74* | 2.19* | 1.86* | 0.67 | 0.06* | 0.20* | 0.23* |
| Semenogelin-1 | SEMG1 | 1.74* | 4.10* | 2.91* | 0.71 | 2.56* | 0.55* | 2.67* |
| F-box only protein 50 | NCCRP1 | 1.73* | 2.05* | 3.97* | 0.77 | 1.78* | 0.88 | 1.38 |
| Cornulin | CRNN | 1.73* | 2.38* | 5.70* | 0.44* | 3.63* | 0.61 | 2.57* |
| Desmocollin-3 | DSC3 | 1.72* | 9.40* | 34.18* | 0.65 | 1.84* | 0.93 | 0.01* |
| Bleomycin hydrolase | BLMH | 1.70* | 1.51* | 1.70* | 1.98* | 1.46 | 1.45 | 1.59 |
| Nuclear speckle splicing regulatory protein 1 | NSRP1 | 1.69* | 2.06* | 2.33* | 1.09 | 0.02* | 0.40* | 1.14 |

| | | | | | | | | |
|---|---|---|---|---|---|---|---|---|
| Cilia- and flagella-associated protein 45 | CFAP45 | 1.69* | 1.98* | 2.43* | 1.19 | 3.11* | 2.25* | 2.51* |
| Acetylcholinesterase | ACHE | 1.68* | 3.16* | 2.26* | 1.09 | 1.69 | 2.30* | 1.62 |
| Solute carrier family 40 member 1 | SLC40A1 | 1.66* | 1.70* | 1.81* | 3.07* | 4.44* | 3.81* | 3.78* |
| Protein C19orf12 | C19orf12 | 1.61* | 2.28* | 1.77* | 1.55 | 1.49 | 1.94* | 1.86 |
| Histidine ammonia-lyase | HAL | 1.60* | 2.11* | 5.94* | 0.35* | 2.50* | 0.56 | 2.12* |
| Retinoic acid receptor responder protein 1 | RARRES1 | 1.60* | 2.72* | 3.25* | 3.23* | 3.43* | 6.05* | 6.66* |
| Trehalase | TREH | 1.60* | 2.35* | 1.52* | 0.90 | 1.42* | 1.88* | 1.13 |
| HLA class II histocompatibility antigen, DM beta chain | HLA-DMB | 1.59* | 13.66* | 8.99* | 1.13 | 1.05 | 10.01* | 8.97* |
| Small integral membrane protein 20 | SMIM20 | 1.53* | 1.65* | 1.62* | 1.27* | 0.19* | 0.38* | 0.30* |
| NK-tumor recognition protein | NKTR | 1.53* | 1.80* | 1.57 | 1.59 | 1.50 | 1.67 | 1.19 |
| Alsin | ALS2 | 1.51* | 1.57 | 1.67* | 1.30 | 1.37 | 1.56 | 1.50 |
| Ubiquinol-cytochrome-c reductase complex assembly factor 6 | UQCC6 | 1.50* | 1.63* | 1.62* | 1.17 | 0.37* | 0.43* | 0.39* |

***B. Up-regulated proteins unique to Aquamin (AQ) [57 proteins]***

| | | Interventions | | | | | | |
|---|---|---|---|---|---|---|---|---|
| | | Control | | | *With LPS & Cytokines* | | | |
| Proteins | Genes | **AQ** | AQ+MES | MES | *LPS-Cyto* | *AQ* | *AQ+MES* | *MES* |
| Keratin, type II cuticular Hb4 | KRT84 | 3.41* | 1.07 | 1.03 | 1.40 | 0.95 | 1.04 | 3.69* |
| Mediator of RNA polymerase II transcription subunit 8 | MED8 | 3.06* | 0.51* | 0.49* | 22.20* | 45.95* | 32.32* | 21.41* |
| Putative histone H2B type 2-C | H2BC20P | 2.73* | 0.76 | 0.85 | 10.59* | 34.44* | 25.80* | 21.48* |
| Heat shock 70 kDa protein 1-like | HSPA1L | 2.61* | 1.48 | 1.29 | 8.97* | 15.03* | 9.11* | 10.55* |
| Apolipoprotein C-II | APOC2 | 2.38* | 0.68 | 0.99 | 9.94* | 21.64* | 14.48* | 17.84* |
| Deleted in malignant brain tumors 1 protein | DMBT1 | 2.24* | 0.89 | 1.16* | 1.67* | 1.57* | 0.60* | 0.73* |
| Stanniocalcin-2 | STC2 | 2.21* | 1.18 | 1.08 | 1.05 | 0.95 | 1.24 | 0.87 |
| Keratin, type II cytoskeletal 80 | KRT80 | 2.20* | 1.11 | 1.12 | 0.99 | 1.72* | 0.99 | 1.07 |
| Cytochrome P450 2C19 | CYP2C19 | 2.12* | 1.15 | 0.61* | 1.64* | 1.98* | 0.88 | 0.62 |
| Carbonic anhydrase 4 | CA4 | 2.10* | 1.45* | 1.09 | 1.18 | 1.36 | 0.85 | 0.69 |
| Profilin-3 | PFN3 | 2.06* | 0.44* | 0.48* | 0.82 | 0.81 | 0.84 | 0.37* |
| Protein disulfide-isomerase A2 | PDIA2 | 2.04* | 1.32 | 1.24 | 1.68* | 0.72 | 0.87 | 1.13 |
| Nucleoplasmin-3 | NPM3 | 1.99* | 1.31 | 1.49 | 2.28* | 0.37* | 0.65 | 0.30* |
| Group IID secretory phospholipase A2 | PLA2G2D | 1.94* | 0.98 | 0.85 | 3.07* | 7.57* | 9.66* | 8.40* |
| Guanylate-binding protein 4 | GBP4 | 1.91* | 0.78 | 0.94 | 22.11* | 26.71* | 25.20* | 24.08* |
| 2-aminomuconic semialdehyde dehydrogenase | ALDH8A1 | 1.90* | 1.21 | 1.32 | 2.50* | 13.33* | 30.36* | 5.64* |
| Glycerophosphoinositol inositolphosphodiesterase GDPD2 | GDPD2 | 1.86* | 1.32 | 0.87 | 0.91 | 1.44 | 1.40 | 0.80 |
| Creatine kinase B-type | CKB | 1.86* | 1.48* | 1.47* | 0.69* | 0.99 | 1.05 | 0.58* |
| UDP-glucuronosyltransferase 2A3 | UGT2A3 | 1.71* | 1.27 | 0.93 | 0.94 | 1.03 | 0.73 | 0.51* |

| | | | | | | | | |
|---|---|---|---|---|---|---|---|---|
| Pterin-4-alpha-carbinolamine dehydratase 2 | PCBD2 | 1.71* | 1.26 | 1.07 | 1.40 | 1.81* | 1.88* | 1.22 |
| Bromodomain-containing protein 8 | BRD8 | 1.65* | 0.96 | 0.74 | 26.61* | 35.18* | 48.74* | 52.93* |
| Hephaestin | HEPH | 1.64* | 1.49* | 1.05 | 0.94 | 1.50* | 1.23 | 0.72* |
| Neuropilin-2 | NRP2 | 1.63* | 1.43 | 1.45* | 1.26 | 1.69* | 1.56 | 1.33 |
| Olfactomedin-4 | OLFM4 | 1.62* | 0.97 | 0.77* | 0.88 | 1.06 | 0.78 | 0.91 |
| Protein PET100 homolog, mitochondrial | PET100 | 1.62* | 1.23 | 0.99 | 1.10 | 1.09 | 1.11 | 0.97 |
| Major facilitator superfamily domain-containing protein 8 | MFSD8 | 1.61* | 1.27 | 1.09 | 2.56* | 4.01* | 3.96* | 3.35* |
| Transmembrane 4 L6 family member 20 | TM4SF20 | 1.61* | 0.93 | 0.65* | 0.63* | 1.51* | 0.97 | 0.50* |
| Aminopeptidase N | ANPEP | 1.60* | 1.32* | 0.97 | 0.91 | 0.80 | 0.95 | 0.67* |
| Amiloride-sensitive sodium channel subunit alpha | SCNN1A | 1.60* | 1.38 | 0.85 | 0.99 | 0.86 | 1.44 | 0.65 |
| V-type immunoglobulin domain-containing suppressor of T-cell activation | VSIR | 1.60* | 1.43 | 1.49* | 2.30* | 5.02* | 3.78* | 3.72* |
| C-C motif chemokine 15 | CCL15 | 1.60* | 1.01 | 0.78 | 0.96 | 0.70 | 0.82 | 0.67 |
| Very low-density lipoprotein receptor | VLDLR | 1.58* | 1.36 | 1.44* | 1.24 | 1.93* | 1.56 | 1.43 |
| Nuclease EXOG, mitochondrial | EXOG | 1.57* | 1.36 | 1.45 | 2.19* | 3.66* | 3.66* | 3.13* |
| IgG receptor FcRn large subunit p51 | FCGRT | 1.56* | 1.42 | 0.97 | 1.38 | 1.32 | 1.30 | 1.05 |
| Hexokinase HKDC1 | HKDC1 | 1.56* | 1.03 | 0.87 | 1.07 | 1.23 | 0.93 | 0.82 |
| Exosome complex component RRP42 | EXOSC7 | 1.55* | 1.10 | 1.10 | 1.22 | 1.16 | 1.15 | 1.05 |
| Matrix-remodeling-associated protein 7 | MXRA7 | 1.55* | 1.50 | 1.19 | 1.41 | 1.04 | 1.02 | 0.86 |
| Solute carrier family 66 member 2 | SLC66A2 | 1.55* | 1.44* | 1.44* | 1.99* | 4.16* | 3.88* | 3.56* |
| Epsin-2 | EPN2 | 1.54* | 1.34* | 1.44* | 1.52* | 1.37 | 1.38 | 1.54* |
| Sulfiredoxin-1 | SRXN1 | 1.54* | 1.21 | 1.01 | 1.11 | 1.42 | 1.20 | 1.33 |
| A-kinase anchor protein 9 | AKAP9 | 1.54* | 0.94 | 0.85 | 1.12 | 1.04 | 0.78 | 0.70 |
| Transmembrane protein 164 | TMEM164 | 1.54* | 1.23 | 0.99 | 0.79 | 1.15 | 1.18 | 0.69 |
| Telomeric repeat-binding factor 2 | TERF2 | 1.54* | 1.23 | 1.29 | 1.43 | 0.60 | 0.56 | 0.66 |
| Keratinocyte-associated protein 2 | KRTCAP2 | 1.54* | 1.29 | 1.18 | 2.55* | 4.86* | 4.63* | 2.87* |
| PWWP domain-containing DNA repair factor 4 | PWWP4 | 1.54* | 1.13 | 1.24 | 0.54* | 1.27 | 1.13 | 0.67 |
| Trefoil factor 1 | TFF1 | 1.54* | 1.08 | 0.91 | 0.61* | 0.63* | 0.48* | 0.37* |
| Transmembrane protein 125 | TMEM125 | 1.53* | 1.26 | 0.95 | 1.26 | 1.86* | 0.98 | 1.15 |
| Complement C1r subcomponent-like protein | C1RL | 1.53* | 1.26 | 1.35 | 1.05 | 1.08 | 0.47* | 0.76 |
| Inactive tyrosine-protein kinase transmembrane receptor ROR1 | ROR1 | 1.53* | 0.91 | 1.21 | 1.01 | 1.12 | 0.82 | 1.05 |
| Matrilysin | MMP7 | 1.52* | 1.02 | 1.05 | 1.66* | 1.74* | 1.24 | 1.42* |
| Twisted gastrulation protein homolog 1 | TWSG1 | 1.51* | 1.20 | 1.09 | 1.12 | 0.77 | 0.85 | 1.00 |
| PC4 and SFRS1-interacting protein | PSIP1 | 1.51* | 1.06 | 1.31* | 1.61* | 0.48* | 0.47* | 1.46* |
| Trinucleotide repeat-containing gene 6B protein | TNRC6B | 1.51* | 1.07 | 0.90 | 0.59* | 0.65 | 1.27 | 0.62 |
| DnaJ homolog subfamily B member 6 | DNAJB6 | 1.51* | 1.38 | 1.48 | 0.98 | 1.06 | 1.27 | 0.98 |
| PAT complex subunit Asterix | WDR83OS | 1.51* | 1.46 | 1.31 | 1.84* | 5.64* | 4.74* | 4.39* |

| | | | | | | | | |
|---|---|---|---|---|---|---|---|---|
| Condensin complex subunit 2 | NCAPH | 1.51* | 1.22 | 1.07 | 1.36 | 2.49* | 2.98* | 1.72* |
| Metalloproteinase inhibitor 2 | TIMP2 | 1.50* | 1.16 | 1.12 | 1.27 | 1.40 | 1.26 | 1.02 |

***C. Up-regulated proteins unique to Aquamin plus Mesalamine (AQ+MES) [113 proteins]***

| | | Interventions | | | | | | |
|---|---|---|---|---|---|---|---|---|
| | | Control | | | *With LPS & Cytokines* | | | |
| Proteins | Genes | AQ | **AQ+MES** | MES | *LPS-Cyto* | *AQ* | *AQ+MES* | *MES* |
| Lysozyme g-like protein 2 | LYG2 | 0.84 | 31.25* | 1.48 | 0.85 | 1.13 | 1.33 | 1.98* |
| Keratin, type II cuticular Hb2 | KRT82 | 0.91 | 23.09* | 1.49* | 0.74 | 1.00 | 0.40* | 1.04 |
| Keratin, type II cuticular Hb5 | KRT85 | 0.66* | 8.93* | 1.06 | 0.69* | 0.55* | 0.40* | 1.22 |
| Maltase-glucoamylase | MGAM | 1.32* | 3.05* | 1.31* | 1.32* | 1.62* | 1.41* | 0.94 |
| 55 kDa erythrocyte membrane protein | MPP1 | 1.16 | 2.80* | 1.47* | 1.10 | 1.36 | 1.92* | 1.21 |
| Dynein regulatory complex protein 10 | IQCD | 0.69 | 2.52* | 1.32 | 0.48* | 0.17* | 0.13* | 0.09* |
| Aspartate dehydrogenase domain-containing protein | ASPDH | 1.49* | 2.23* | 1.22 | 1.34 | 1.82* | 1.07 | 1.07 |
| Sorbin and SH3 domain-containing protein 1 | SORBS1 | 0.97 | 2.18* | 1.43 | 1.40 | 1.86* | 1.35 | 1.24 |
| Apolipoprotein D | APOD | 1.10 | 2.13* | 1.32 | 4.26* | 2.98* | 1.00 | 2.56* |
| 5'-3' exonuclease PLD3 | PLD3 | 1.31* | 2.06* | 1.16 | 1.16 | 1.31 | 1.17 | 1.12 |
| Seipin | BSCL2 | 1.44* | 2.03* | 1.20 | 0.85 | 1.45 | 1.27 | 1.26 |
| Guanylate-binding protein 7 | GBP7 | 1.02 | 2.02* | 0.88 | 35.72* | 32.72* | 30.59* | 33.79* |
| Olfactory receptor 1M1 | OR1M1 | 1.48* | 1.95* | 1.32 | 1.06 | 1.87* | 2.10* | 1.19 |
| Protein ABHD13 | ABHD13 | 0.71 | 1.91* | 1.42 | 0.70 | 0.92 | 1.38 | 1.31 |
| Laminin subunit beta-2 | LAMB2 | 0.67* | 1.91* | 1.30* | 1.38* | 1.83* | 2.20* | 3.05* |
| Zymogen granule membrane protein 16 | ZG16 | 1.23* | 1.91* | 1.33* | 1.18 | 1.16 | 0.87 | 1.20 |
| Adhesion G-protein coupled receptor F1 | ADGRF1 | 1.32* | 1.87* | 1.24 | 1.65* | 1.98* | 1.80* | 1.81* |
| Melanotransferrin | MELTF | 1.46* | 1.86* | 1.26 | 1.24 | 1.78 | 1.93* | 1.35 |
| Glutaminyl-peptide cyclotransferase-like protein | QPCTL | 1.26 | 1.86* | 1.14 | 1.24 | 1.57 | 1.58 | 1.49 |
| Sodium-dependent phosphate transporter 1 | SLC20A1 | 1.47* | 1.85* | 1.42* | 0.90 | 0.54 | 0.70 | 0.57 |
| Neutrophil elastase | ELANE | 0.97 | 1.85* | 0.83 | 1.30 | 3.28* | 0.92 | 2.27* |
| Serpin B7 | SERPINB7 | 1.00 | 1.84* | 1.21 | 1.19 | 2.08* | 2.49* | 1.71* |
| Group 10 secretory phospholipase A2 | PLA2G10 | 1.13 | 1.83* | 1.30 | 0.87 | 0.14* | 0.82 | 0.58 |
| Mitochondrial pyruvate carrier 1 | MPC1 | 0.99 | 1.83* | 1.33 | 1.58 | 2.75* | 2.41* | 2.70* |
| Glutathione S-transferase Mu 4 | GSTM4 | 1.18 | 1.83* | 0.87 | 0.96 | 0.87 | 1.26 | 0.72 |
| DnaJ homolog subfamily B member 9 | DNAJB9 | 1.26 | 1.81* | 1.41 | 1.32 | 1.26 | 1.76 | 1.72 |
| Collagen alpha-1(XV) chain | COL15A1 | 0.83 | 1.81* | 1.43 | 1.57 | 1.31 | 1.62 | 2.31* |
| UDP-GlcNAc:betaGal beta-1,3-N-acetylglucosaminyltransferase 7 | B3GNT7 | 1.09 | 1.78* | 1.50* | 0.94 | 1.56 | 1.64* | 1.41 |
| Enhancer of filamentation 1 | NEDD9 | 1.46* | 1.77* | 1.30 | 0.93 | 1.23 | 1.75* | 0.91 |
| Alpha-1-antitrypsin | SERPINA1 | 1.26* | 1.76* | 1.41* | 2.93* | 2.81* | 2.44* | 2.34* |

| | | | | | | | | |
|---|---|---|---|---|---|---|---|---|
| HLA class II histocompatibility antigen, DR alpha chain | HLA-DRA | 1.05 | 1.75* | 1.05 | 28.26* | 24.40* | 28.76* | 24.96* |
| 1-phosphatidylinositol 4,5-bisphosphate phosphodiesterase delta-1 | PLCD1 | 1.04 | 1.75* | 1.10 | 0.94 | 0.93 | 0.91 | 0.88 |
| Tigger transposable element-derived protein 3 | TIGD3 | 0.93 | 1.74* | 0.94 | 1.15 | 1.30 | 1.69 | 1.19 |
| Deoxyhypusine synthase | DHPS | 0.95 | 1.73* | 1.01 | 1.75* | 1.58* | 1.88* | 1.77* |
| Arylsulfatase D | ARSD | 0.83 | 1.73* | 1.02 | 0.94 | 0.75 | 1.04 | 0.99 |
| Small integral membrane protein 24 | SMIM24 | 1.45 | 1.71* | 0.90 | 1.51 | 1.28 | 1.22 | 0.90 |
| Sodium-dependent lysophosphatidylcholine symporter 1 | MFSD2A | 1.38 | 1.71* | 1.13 | 1.34 | 0.87 | 1.25 | 1.57 |
| Electron transfer flavoprotein regulatory factor 1 | ETFRF1 | 1.41* | 1.71* | 1.48* | 1.68* | 1.53 | 1.47 | 1.73* |
| Bromodomain-containing protein 9 | BRD9 | 0.70* | 1.71* | 1.15 | 1.38* | 2.07* | 3.25* | 5.06* |
| Cytochrome P450 4F11 | CYP4F11 | 0.97 | 1.71* | 1.13 | 1.31 | 1.55 | 1.55 | 1.32 |
| Threonylcarbamoyl-AMP synthase | YRDC | 1.20 | 1.69* | 1.25 | 1.89* | 0.94 | 1.91* | 1.10 |
| Superoxide dismutase [Mn], mitochondrial | SOD2 | 1.20* | 1.69* | 1.12 | 1.74* | 1.96* | 2.13* | 2.03* |
| 2-acylglycerol O-acyltransferase 2 | MOGAT2 | 1.18 | 1.68* | 1.15 | 1.14 | 1.23 | 1.38 | 0.98 |
| Cytokine receptor common subunit gamma | IL2RG | 1.27 | 1.67* | 1.27 | 1.28 | 1.27 | 1.55 | 0.97 |
| Protein MMP24OS | MMP24OS | 1.36 | 1.67* | 1.29 | 1.11 | 1.61 | 1.43 | 1.33 |
| Disco-interacting protein 2 homolog A | DIP2A | 1.26 | 1.67* | 1.47* | 1.17 | 1.38 | 1.38 | 1.00 |
| PWWP domain-containing protein 2A | PWWP2A | 1.12 | 1.65* | 1.22 | 1.07 | 0.59 | 0.61 | 0.74 |
| Galactosylgalactosylxylosylprotein 3-beta-glucuronosyltransferase 3 | B3GAT3 | 1.46* | 1.65* | 1.29 | 1.47* | 1.77 | 2.12* | 1.07 |
| E3 ubiquitin ligase RNF121 | RNF121 | 1.22 | 1.65* | 1.28 | 1.18 | 1.43 | 1.63 | 1.17 |
| Amino acid transporter heavy chain SLC3A1 | SLC3A1 | 1.30 | 1.64* | 1.12 | 1.08 | 1.43 | 1.21 | 0.94 |
| Protein GUCD1 | GUCD1 | 1.02 | 1.64* | 1.44 | 0.74 | 1.48 | 1.65 | 1.75 |
| Motile sperm domain-containing protein 1 | MOSPD1 | 1.07 | 1.64* | 1.30 | 0.65 | 0.27* | 0.58 | 0.13* |
| UMP-CMP kinase 2, mitochondrial | CMPK2 | 0.96 | 1.64* | 1.39 | 1.53 | 1.76* | 2.93* | 2.32* |
| Putative HLA class I histocompatibility antigen, alpha chain H | HLA-H | 1.19 | 1.64* | 1.17 | 2.96* | 2.63* | 2.82* | 2.42* |
| Transcription elongation factor SPT4 | SUPT4H1 | 1.14 | 1.63* | 0.98 | 1.44 | 1.15 | 1.33 | 1.64 |
| Ras-related protein Rab-30 | RAB30 | 1.13 | 1.62* | 1.22 | 1.04 | 1.12 | 1.31 | 0.98 |
| Voltage-gated potassium channel subunit beta-2 | KCNAB2 | 1.44* | 1.62* | 1.25 | 1.16 | 1.22 | 0.94 | 0.98 |
| Arylacetamide deacetylase | AADAC | 1.04 | 1.62 | 1.20 | 1.20 | 1.10 | 1.11 | 1.66 |
| ATP-binding cassette sub-family D member 1 | ABCD1 | 1.20 | 1.61* | 1.10 | 0.92 | 1.09 | 1.12 | 0.80 |
| Signal peptide peptidase-like 2B | SPPL2B | 1.26 | 1.61* | 1.48* | 1.77* | 2.29* | 2.26* | 2.37* |
| Immunoglobulin superfamily member 8 | IGSF8 | 1.11 | 1.61* | 1.44* | 1.53* | 2.10* | 1.93* | 1.61* |
| TLC domain-containing protein 4 | TLCD4 | 1.45* | 1.60* | 1.10 | 1.12 | 1.45 | 1.40 | 1.02 |
| Tissue factor | F3 | 0.97 | 1.60* | 1.48* | 1.34* | 1.23 | 1.69* | 1.58* |
| Intestinal-type alkaline phosphatase | ALPI | 1.25 | 1.60* | 1.15 | 1.01 | 1.45 | 1.54 | 1.09 |
| Phosphatidylinositol 4-kinase type 2-beta | PI4K2B | 1.29 | 1.60* | 1.40 | 1.23 | 1.27 | 1.36 | 1.01 |
| Tetraspanin-14 | TSPAN14 | 1.01 | 1.60* | 0.95 | 1.09 | 1.38* | 1.29 | 1.22 |

| | | | | | | | | |
|---|---|---|---|---|---|---|---|---|
| Mitochondrial import receptor subunit TOM40B | TOMM40L | 1.43* | 1.59* | 1.46* | 1.94* | 2.74* | 2.90* | 2.29* |
| Prothrombin | F2 | 0.84 | 1.59 | 1.40 | 1.51* | 2.10* | 3.12* | 3.81* |
| Dixin | DIXDC1 | 1.10 | 1.59* | 1.27 | 1.43 | 0.87 | 1.95* | 1.47 |
| Protein FAM83G | FAM83G | 1.12 | 1.58* | 1.13 | 0.85 | 0.46* | 0.56 | 0.54 |
| Bis(5'-adenosyl)-triphosphatase | FHIT | 1.09 | 1.58* | 1.32 | 1.78* | 1.32 | 1.52 | 1.77* |
| HIG1 domain family member 1A, mitochondrial | HIGD1A | 1.46 | 1.58* | 1.10 | 1.43 | 1.83* | 2.31* | 1.94* |
| Pannexin-1 | PANX1 | 0.84 | 1.58 | 0.82 | 1.18 | 1.37 | 1.21 | 1.26 |
| Ubiquinol-cytochrome-c reductase complex assembly factor 3 | UQCC3 | 1.21 | 1.58 | 1.28 | 0.89 | 0.90 | 0.92 | 0.88 |
| Lysosomal thioesterase PPT2 | PPT2 | 0.78 | 1.58* | 0.68 | 1.00 | 0.92 | 0.56 | 0.82 |
| Proton-coupled folate transporter | SLC46A1 | 1.06 | 1.56 | 1.28 | 1.04 | 0.94 | 0.91 | 0.83 |
| Phosphatidylethanolamine N-methyltransferase | PEMT | 1.16 | 1.56 | 1.27 | 1.00 | 1.29 | 1.88* | 1.29 |
| STARD3 N-terminal-like protein | STARD3NL | 1.12 | 1.56* | 1.24 | 1.68* | 1.74 | 1.55 | 1.57 |
| Metallothionein-1E | MT1E | 0.72 | 1.56* | 1.26 | 1.03 | 0.83 | 1.26 | 1.43 |
| Dual oxidase 2 | DUOX2 | 1.39* | 1.56* | 1.15* | 0.94 | 1.23 | 1.30 | 0.88 |
| Collagen alpha-1(VI) chain | COL6A1 | 0.88 | 1.56* | 1.49* | 1.06 | 1.51* | 2.05* | 2.64* |
| Ubiquitin/ISG15-conjugating enzyme E2 L6 | UBE2L6 | 1.34 | 1.56* | 1.29 | 5.90* | 5.70* | 6.47* | 6.58* |
| Sacsin | SACS | 1.37* | 1.56* | 1.45* | 1.67* | 2.82* | 2.91* | 2.32* |
| Nidogen-1 | NID1 | 0.75* | 1.56* | 1.44* | 1.41* | 1.86* | 2.21* | 2.89* |
| PDZK1-interacting protein 1 | PDZK1IP1 | 1.21 | 1.54* | 1.40* | 1.11 | 1.42 | 1.37 | 1.40 |
| P2X purinoceptor 4 | P2RX4 | 1.27* | 1.54* | 1.15 | 0.99 | 1.36 | 1.25 | 0.92 |
| Neurotensin/neuromedin N | NTS | 0.74 | 1.54* | 1.30 | 1.01 | 0.50* | 1.03 | 0.72 |
| Solute carrier family 52, riboflavin transporter, member 3 | SLC52A3 | 1.21 | 1.54 | 1.45 | 1.29 | 1.72 | 1.52 | 1.49 |
| b(0,+)-type amino acid transporter 1 | SLC7A9 | 1.23 | 1.53* | 1.26 | 1.20 | 1.33 | 1.37 | 0.91 |
| Programmed cell death 1 ligand 1 | CD274 | 0.43* | 1.53 | 1.44 | 16.94* | 16.35* | 19.58* | 15.89* |
| Protein O-linked-mannose beta-1,4-N-acetylglucosaminyltransferase 2 | POMGNT2 | 1.04 | 1.53 | 1.41 | 1.30 | 1.55 | 1.88 | 1.04 |
| Collagen alpha-1(I) chain | COL1A1 | 0.89 | 1.53* | 1.40* | 1.45* | 1.99* | 2.46* | 3.30* |
| Mitochondrial adenyl nucleotide antiporter SLC25A25 | SLC25A25 | 1.30 | 1.53 | 1.24 | 1.58* | 2.23* | 2.58* | 1.80* |
| Acyl-coenzyme A synthetase ACSM3, mitochondrial | ACSM3 | 1.15 | 1.53 | 1.29 | 1.43 | 1.41 | 1.74* | 1.81* |
| Sorcin | SRI | 1.45* | 1.52* | 1.05 | 0.89 | 1.13 | 1.04 | 0.79 |
| Elongation factor 1-alpha 2 | EEF1A2 | 0.84 | 1.52* | 1.19 | 1.02 | 1.66* | 1.71* | 1.43 |
| Glutaminyl-peptide cyclotransferase | QPCT | 1.49* | 1.52* | 1.48* | 0.83 | 1.22 | 0.98 | 1.26 |
| Organic solute transporter subunit beta | SLC51B | 1.09 | 1.52* | 1.30* | 1.00 | 1.09 | 1.25 | 1.17 |
| Thymidine phosphorylase | TYMP | 1.24 | 1.52* | 1.28 | 4.52* | 3.92* | 4.88* | 4.23* |
| Proteasome subunit beta type-8 | PSMB8 | 1.37* | 1.51* | 1.39* | 2.00* | 1.62* | 2.82* | 1.92* |
| Transmembrane protein 171 | TMEM171 | 1.40 | 1.51 | 0.90 | 1.34 | 1.23 | 0.97 | 0.84 |
| GPI inositol-deacylase | PGAP1 | 1.24 | 1.51* | 1.27 | 1.13 | 1.52 | 1.91* | 1.21 |

| | | | | | | | | |
|---|---|---|---|---|---|---|---|---|
| Protein S100-A9 | S100A9 | 0.60* | 1.51* | 1.41* | 1.53* | 1.35 | 1.39 | 2.46* |
| Exostosin-2 | EXT2 | 1.09 | 1.51* | 1.42* | 1.25 | 1.38 | 1.43 | 1.85* |
| BAH and coiled-coil domain-containing protein 1 | BAHCC1 | 0.54* | 1.51* | 0.94 | 0.43* | 0.49* | 0.40* | 0.22* |
| Broad substrate specificity ATP-binding cassette transporter ABCG2 | ABCG2 | 1.44* | 1.51* | 1.23 | 1.08 | 1.22 | 1.41 | 1.17 |
| Mitochondrial fission process protein 1 | MTFP1 | 1.06 | 1.51* | 1.46* | 1.44 | 1.61* | 1.62* | 1.80* |
| Rootletin | CROCC | 0.49* | 1.51* | 0.96 | 0.74 | 0.50* | 0.53* | 0.43* |
| ER membrane protein complex subunit 7 | EMC7 | 0.97 | 1.51* | 1.43* | 1.54* | 1.50 | 1.59 | 1.99* |
| High affinity copper uptake protein 1 | SLC31A1 | 1.36* | 1.51* | 1.17 | 1.17 | 1.11 | 1.59* | 1.16 |
| Ectonucleotide pyrophosphatase/phosphodiesterase family member 3 | ENPP3 | 1.33 | 1.50 | 0.93 | 1.01 | 1.39 | 1.12 | 1.00 |
| Zymogen granule protein 16 homolog B | EECP | 1.16 | 1.50* | 1.36* | 1.18 | 2.47* | 1.02 | 2.66* |
| Acyl-CoA:lysophosphatidylglycerol acyltransferase 1 | LPGAT1 | 1.10 | 1.50* | 1.18 | 1.37* | 1.49* | 1.82* | 1.33 |

***D. Up-regulated proteins unique to Mesalamine (MES) [86 proteins]***

| | | Interventions | | | | | | |
|---|---|---|---|---|---|---|---|---|
| | | Control | | | *With LPS & Cytokines* | | | |
| Proteins | Genes | AQ | AQ+MES | **MES** | *LPS-Cyto* | *AQ* | *AQ+MES* | *MES* |
| Complement C1r subcomponent | C1R | 0.94 | 1.31 | 14.51* | 0.63 | 0.54 | 1.27 | 0.15* |
| Metalloreductase STEAP4 | STEAP4 | 0.92 | 1.01 | 12.59* | 0.82 | 0.89 | 0.98 | 1.29 |
| Immunoglobulin lambda variable 1-51 | IGLV1-51 | 0.85 | 1.31 | 8.76* | 0.95 | 0.92 | 0.94 | 0.55* |
| Immunoglobulin lambda constant 2 | IGLC2 | 0.66* | 1.33 | 4.95* | 1.42 | 3.25* | 2.14* | 1.55 |
| Protein S100-A7A | S100A7A | 0.27* | 0.40* | 4.36* | 0.20* | 0.48* | 0.17* | 0.46* |
| Histidine-rich glycoprotein | HRG | 0.76 | 1.27 | 4.14* | 0.61 | 0.90 | 1.03 | 1.39 |
| Haptoglobin | HP | 0.70* | 1.32* | 3.72* | 1.15 | 3.32* | 1.47* | 1.19 |
| Complement factor D | CFD | 0.59* | 1.43 | 3.72* | 0.81 | 0.58 | 1.54 | 2.05* |
| Mediator of RNA polymerase II transcription subunit 16 | MED16 | 1.30 | 1.31 | 3.61* | 0.75 | 1.32 | 0.83 | 0.80 |
| Immunoglobulin heavy constant alpha 1 | IGHA1 | 0.68* | 1.29* | 3.28* | 1.11 | 2.69* | 1.30 | 2.32* |
| Mitotic-spindle organizing protein 1 | MZT1 | 1.14 | 1.09 | 2.73* | 0.92 | 0.63 | 0.71 | 2.13* |
| Immunoglobulin lambda-1 light chain | | 0.85 | 1.19 | 2.66* | 1.84* | 3.41* | 4.29* | 5.54* |
| Fibrinogen alpha chain | FGA | 0.63* | 0.68 | 2.55* | 2.12* | 0.76 | 1.05 | 2.43* |
| Probable phosphoglycerate mutase 4 | PGAM4 | 0.53* | 1.15 | 2.54* | 0.51* | 0.63 | 1.94* | 2.77* |
| Fibrinogen gamma chain | FGG | 0.35* | 1.28 | 2.38* | 2.58* | 3.47* | 3.42* | 6.21* |
| Tetranectin | CLEC3B | 0.72 | 1.19 | 2.37* | 2.09* | 1.76* | 2.13* | 2.78* |
| Serpin B12 | SERPINB12 | 0.96 | 1.21 | 2.32* | 1.07 | 3.22* | 0.93 | 2.21* |
| Ubiquitin thioesterase OTU1 | YOD1 | 1.22 | 1.23 | 2.20* | 0.87 | 0.79 | 0.55 | 0.91 |
| Tudor domain-containing protein 3 | TDRD3 | 1.39 | 1.39 | 2.19* | 0.80 | 0.14* | 0.30* | 0.42* |
| Complement C4-A | C4A | 0.73* | 1.29* | 2.17* | 1.82* | 3.29* | 3.86* | 4.65* |

| | | | | | | | | |
|---|---|---|---|---|---|---|---|---|
| T-complex protein 10A homolog 1 | TCP10L | 0.85 | 1.26 | 2.12* | 0.87 | 0.83 | 2.79* | 2.40* |
| Zinc finger protein 687 | ZNF687 | 1.25 | 1.02 | 1.97* | 0.80 | 1.33 | 1.36 | 0.92 |
| Hemoglobin subunit epsilon | HBE1 | 0.77 | 1.36 | 1.95* | 1.80* | 2.27* | 2.55* | 3.81* |
| Pigment epithelium-derived factor | SERPINF1 | 0.57* | 1.24 | 1.90* | 1.24 | 0.96 | 2.19* | 3.38* |
| Filamin-C | FLNC | 1.23 | 0.84 | 1.90* | 2.91* | 3.79* | 4.00* | 2.41* |
| Alpha-2-macroglobulin | A2M | 0.61* | 1.48* | 1.87* | 1.88* | 1.77* | 2.51* | 3.48* |
| SPARC | SPARC | 0.54* | 1.40* | 1.85* | 1.86* | 1.88* | 3.08* | 3.36* |
| Serpin B3 | SERPINB3 | 0.72* | 1.14 | 1.82* | 1.19 | 2.93* | 0.78 | 1.96* |
| Immortalization up-regulated protein | IMUP | 0.66* | 1.26 | 1.81* | 1.60* | 1.16 | 1.10 | 1.75* |
| N-acetylglucosamine-1-phosphodiester alpha-N-acetylglucosaminidase | NAGPA | 0.82 | 1.28 | 1.80* | 0.75 | 0.93 | 1.45 | 1.58 |
| Protein BCAP | ODF2L | 0.43* | 1.24 | 1.79* | 2.23* | 2.03* | 2.99* | 3.70* |
| N-acetyltransferase ESCO1 | ESCO1 | 1.20 | 0.85 | 1.78* | 1.70* | 1.20 | 0.90 | 1.49 |
| Cilia- and flagella-associated protein 100 | CFAP100 | 0.89 | 1.31 | 1.78* | 1.65* | 1.48 | 2.37* | 2.52* |
| Alpha-fetoprotein | AFP | 0.54* | 1.16 | 1.75* | 1.41* | 1.41* | 2.14* | 3.27* |
| Tumor necrosis factor receptor superfamily member 12A | TNFRSF12A | 0.88 | 1.46 | 1.75* | 1.47 | 1.35 | 1.30 | 1.52 |
| Stromal interaction molecule 2 | STIM2 | 0.77 | 1.30 | 1.75* | 1.84* | 1.54 | 2.36* | 3.21* |
| Alpha-2-HS-glycoprotein | AHSG | 0.70* | 1.45* | 1.73* | 1.82* | 1.47* | 2.35* | 2.40* |
| Laminin subunit alpha-2 | LAMA2 | 1.06 | 1.20 | 1.71* | 1.33 | 1.36 | 1.54 | 1.76 |
| Thymosin beta-4 | TMSB4X | 0.55* | 0.94 | 1.71* | 1.20 | 1.01 | 1.17 | 1.93* |
| Protein-glutamine gamma-glutamyltransferase K | TGM1 | 1.13 | 1.39 | 1.71* | 0.66 | 3.12* | 0.49* | 2.44* |
| Collagen alpha-1(IV) chain | COL4A1 | 0.50* | 1.17 | 1.70* | 1.82* | 2.23* | 2.22* | 3.55* |
| Transmembrane protein 51 | TMEM51 | 1.19 | 1.38 | 1.70* | 1.29 | 0.27* | 0.66 | 0.80 |
| Serum paraoxonase/arylesterase 1 | PON1 | 0.69 | 1.33 | 1.69* | 1.47 | 3.15* | 4.26* | 4.58* |
| Lysyl oxidase homolog 3 | LOXL3 | 0.95 | 1.24 | 1.68* | 0.97 | 1.28 | 1.51 | 1.85* |
| Plexin domain-containing protein 2 | PLXDC2 | 1.16 | 1.24 | 1.67* | 2.61* | 5.06* | 7.61* | 8.74* |
| Glutathione S-transferase Mu 2 | GSTM2 | 0.74 | 1.19 | 1.67* | 1.22 | 3.12* | 3.94* | 3.58* |
| Complement factor B | CFB | 1.12 | 1.46 | 1.67* | 2.18* | 3.23* | 2.38* | 2.44* |
| tRNA methyltransferase 10 homolog A | TRMT10A | 0.87 | 1.03 | 1.66* | 0.99 | 1.01 | 1.30 | 2.12* |
| Gasdermin-A | GSDMA | 1.05 | 1.25 | 1.66* | 1.00 | 2.19* | 0.72 | 0.83 |
| Crooked neck-like protein 1 | CRNKL1 | 0.97 | 1.39 | 1.66* | 0.83 | 0.82 | 1.05 | 1.05 |
| Glycoprotein endo-alpha-1,2-mannosidase | MANEA | 1.08 | 1.29 | 1.66* | 0.72 | 1.03 | 1.41 | 1.28 |
| Pro-glucagon | GCG | 0.65* | 1.31* | 1.64* | 0.78 | 0.99 | 0.72 | 0.86 |
| HLA class II histocompatibility antigen, DR beta 3 chain | HLA-DRB3 | 0.90 | 1.00 | 1.64* | 12.42* | 9.92* | 10.03* | 11.55* |
| Non-histone chromosomal protein HMG-17 | HMGN2 | 0.56* | 1.04 | 1.61* | 0.87 | 0.80 | 1.21 | 1.52 |
| Kynureninase | KYNU | 0.82 | 1.00 | 1.61* | 0.52* | 0.92 | 1.21 | 1.74* |
| Vitamin D-binding protein | GC | 0.71* | 1.25 | 1.61* | 1.69* | 1.51* | 2.10* | 2.04* |
| Sugar phosphate exchanger 3 | SLC37A3 | 1.09 | 1.42 | 1.60* | 0.81 | 1.47 | 1.15 | 1.29 |

| | | | | | | | | |
|---|---|---|---|---|---|---|---|---|
| Hepatocyte growth factor-like protein | MST1 | 1.03 | 1.45* | 1.60* | 1.62* | 2.56* | 3.35* | 4.24* |
| Interferon-induced protein with tetratricopeptide repeats 5 | IFIT5 | 1.04 | 1.34 | 1.60* | 2.00* | 1.09 | 1.96* | 2.05* |
| Cyclin-dependent kinase 2 | CDK2 | 0.76 | 1.14 | 1.60* | 0.69 | 1.16 | 0.98 | 1.35 |
| Inter-alpha-trypsin inhibitor heavy chain H4 | ITIH4 | 0.56* | 1.42 | 1.59* | 2.09* | 1.62* | 2.06* | 3.18* |
| Probable serine carboxypeptidase CPVL | CPVL | 0.97 | 1.32 | 1.59* | 1.34 | 1.15 | 1.65* | 2.03* |
| Mediator of RNA polymerase II transcription subunit 30 | MED30 | 0.87 | 1.43 | 1.58* | 0.93 | 0.94 | 1.28 | 1.01 |
| Hemopexin | HPX | 0.65* | 1.28 | 1.58* | 1.68* | 1.54 | 1.99* | 3.50* |
| Collagen alpha-3(VI) chain | COL6A3 | 0.29* | 0.50* | 1.58* | 1.31 | 0.81 | 1.25 | 1.92* |
| Insulin-like growth factor-binding protein 4 | IGFBP4 | 0.88 | 1.45* | 1.57* | 0.83 | 0.89 | 1.38 | 1.56 |
| Vesicle-associated membrane protein 5 | VAMP5 | 1.13 | 1.43 | 1.57 | 7.65* | 6.90* | 6.61* | 6.29* |
| Midasin | MDN1 | 0.66* | 1.29 | 1.56* | 1.48* | 1.53* | 2.03* | 2.59* |
| Nuclear envelope pore membrane protein POM 121 | POM121 | 0.75 | 1.26 | 1.56* | 0.87 | 0.22* | 1.01 | 1.38 |
| Uncharacterized protein MISP3 | MISP3 | 1.18 | 1.36 | 1.56* | 1.30 | 1.66 | 1.70 | 2.06* |
| Antithrombin-III | SERPINC1 | 0.69* | 1.19 | 1.55* | 1.84* | 1.44* | 1.87* | 2.27* |
| Serine protease HTRA1 | HTRA1 | 0.84 | 1.16 | 1.55* | 0.94 | 1.32 | 1.42 | 1.99* |
| Inter-alpha-trypsin inhibitor heavy chain H1 | ITIH1 | 0.48* | 0.96 | 1.54* | 0.48* | 0.48* | 1.13 | 1.53 |
| Transcription initiation factor IIA subunit 2 | GTF2A2 | 1.36 | 1.35 | 1.54* | 1.37 | 0.59 | 0.36* | 0.48* |
| Cystatin-S | CST4 | 0.83 | 1.14 | 1.53* | 1.06 | 2.72* | 1.49 | 2.96* |
| Frizzled-5 | FZD5 | 1.21 | 1.25 | 1.53* | 1.21 | 1.09 | 1.03 | 1.13 |
| Secretoglobin family 1D member 2 | SCGB1D2 | 0.91 | 1.39 | 1.53* | 1.04 | 3.37* | 0.82 | 2.35* |
| Tumor necrosis factor receptor superfamily member 5 | CD40 | 0.84 | 1.23 | 1.53 | 3.99* | 4.56* | 4.44* | 4.22* |
| Heat shock 70 kDa protein 13 | HSPA13 | 0.96 | 1.49* | 1.51* | 1.27 | 1.61* | 1.99* | 2.46* |
| Vitronectin | VTN | 0.53* | 1.33 | 1.51* | 1.58* | 1.55 | 1.93* | 2.76* |
| SAP domain-containing ribonucleoprotein | SARNP | 1.10 | 1.28 | 1.51* | 0.98 | 0.53* | 0.92 | 0.93 |
| Ras-related protein Rab-33B | RAB33B | 0.79 | 0.93 | 1.51* | 0.76 | 0.86 | 0.78 | 0.96 |
| Interferon-induced GTP-binding protein Mx1 | MX1 | 1.07 | 1.34 | 1.50* | 3.61* | 4.42* | 5.17* | 4.06* |
| Apolipoprotein E | APOE | 0.90 | 1.42* | 1.50* | 1.64* | 1.69* | 1.93* | 2.28* |
| MANSC domain-containing protein 1 | MANSC1 | 0.94 | 1.34 | 1.50* | 1.10 | 2.34* | 2.35* | 1.72* |
| 5-formyltetrahydrofolate cyclo-ligase | MTHFS | 1.17 | 1.44 | 1.50* | 1.19 | 0.75 | 0.95 | 1.15 |

***E. Common up-regulated proteins between Aquamin and Aquamin plus Mesalamine [30 proteins]***

| | | Interventions | | | | | | |
|---|---|---|---|---|---|---|---|---|
| | | Control | | | *With LPS & Cytokines* | | | |
| Proteins | Genes | **AQ** | **AQ+MES** | MES | *LPS-Cyto* | *AQ* | *AQ+MES* | *MES* |
| Fibronectin type III and SPRY domain-containing protein 1 | FSD1 | 3.62* | 3.96* | 1.02 | 0.70 | 2.52* | 1.94* | 0.49* |
| Cadherin-17 | CDH17 | 3.59* | 3.47* | 1.05 | 0.91 | 2.69* | 2.67* | 0.73* |

| | | | | | | | | |
|---|---|---|---|---|---|---|---|---|
| Calcium/manganese antiporter SLC30A10 | SLC30A10 | 3.21* | 2.20* | 1.22 | 1.36 | 2.24* | 2.25* | 0.82 |
| Putative nucleoside diphosphate kinase | NME2P1 | 2.99* | 1.90* | 1.48* | 2.39* | 0.55* | 0.75 | 0.60* |
| Sulfate transporter | SLC26A2 | 2.55* | 1.80* | 0.90 | 0.65* | 0.48* | 0.77 | 0.44* |
| Cytochrome b | MT-CYB | 2.22* | 1.67* | 1.38 | 3.21* | 5.40* | 5.01* | 3.12* |
| Desmoglein-2 | DSG2 | 2.17* | 2.21* | 0.93 | 1.08 | 2.03* | 2.03* | 1.45* |
| Malignant fibrous histiocytoma-amplified sequence 1 | MFHAS1 | 2.14* | 1.68* | 1.35 | 1.22 | 1.41 | 1.16 | 1.12 |
| Protocadherin-1 | PCDH1 | 2.10* | 2.09* | 1.14 | 0.88 | 1.93* | 1.95* | 1.13 |
| Ubiquinone biosynthesis protein COQ4 homolog, mitochondrial | COQ4 | 1.99* | 1.66* | 0.97 | 1.02 | 0.88 | 0.76 | 0.40* |
| TPA-induced transmembrane protein | TTMP | 1.81* | 1.96* | 1.42 | 2.23* | 4.60* | 4.29* | 3.70* |
| Tetraspanin-33 | TSPAN33 | 1.76* | 1.53 | 1.49 | 2.39* | 3.91* | 2.87* | 2.46* |
| Membrane-spanning 4-domains subfamily A member 10 | MS4A10 | 1.73* | 2.21* | 1.28 | 1.40 | 2.17* | 1.20 | 1.03 |
| [Pyruvate dehydrogenase [acetyl-transferring]]-phosphatase 2, mitochondrial | PDP2 | 1.70* | 1.56* | 1.29 | 1.09 | 0.59 | 0.86 | 0.90 |
| Sodium/myo-inositol cotransporter | SLC5A3 | 1.68* | 1.70* | 1.41 | 2.45* | 3.59* | 4.91* | 2.00* |
| Testis-expressed protein 9 | TEX9 | 1.68* | 1.53* | 0.95 | 1.42* | 1.70* | 2.57* | 2.77* |
| Arginase-2, mitochondrial | ARG2 | 1.67* | 1.74* | 1.25 | 1.44 | 1.55 | 1.51 | 0.93 |
| Glycerophosphodiester phosphodiesterase 1 | GDE1 | 1.65* | 1.63* | 1.36* | 2.17* | 3.02* | 3.04* | 1.72* |
| Sodium-dependent neutral amino acid transporter B(0)AT1 | SLC6A19 | 1.63* | 2.12* | 0.93 | 1.71* | 1.74* | 1.12 | 0.47* |
| Protein O-glucosyltransferase 3 | POGLUT3 | 1.62* | 1.67* | 1.49* | 0.96 | 0.48* | 0.78 | 1.14 |
| Ubiquilin-2 | UBQLN2 | 1.61* | 1.52* | 1.41* | 1.44* | 0.79 | 0.82 | 0.89 |
| Endoplasmic reticulum membrane adapter protein XK | XK | 1.59* | 1.81* | 1.36 | 2.33* | 3.37* | 4.41* | 2.84* |
| Fatty-acid amide hydrolase 2 | FAAH2 | 1.58* | 1.52 | 1.12 | 1.00 | 1.24 | 1.44 | 0.97 |
| Solute carrier family 53 member 1 | XPR1 | 1.58* | 1.51 | 1.38 | 1.17 | 1.61 | 1.70* | 1.32 |
| Homeobox protein DBX1 | DBX1 | 1.57* | 1.54* | 1.49* | 3.07* | 6.91* | 6.60* | 5.35* |
| Battenin | CLN3 | 1.57* | 2.13* | 1.44* | 1.94* | 1.87* | 2.41* | 2.39* |
| Protein YIF1A | YIF1A | 1.55* | 1.60 | 1.42 | 2.21* | 3.72* | 3.42* | 2.33* |
| Cadherin-3 | CDH3 | 1.54* | 1.58* | 1.00 | 1.05 | 1.39 | 1.36 | 0.82 |
| LHFPL tetraspan subfamily member 2 protein | LHFPL2 | 1.51* | 1.61* | 1.23 | 1.92* | 2.46* | 1.60 | 1.57 |
| ATP-binding cassette sub-family C member 2 | ABCC2 | 1.50* | 2.42* | 1.43* | 1.53* | 1.48* | 1.89* | 1.60* |

***F. Common up-regulated proteins between Aquamin and Mesalamine [10 proteins]***

| | | Interventions | | | | | | |
|---|---|---|---|---|---|---|---|---|
| | | Control | | | *With LPS & Cytokines* | | | |
| Proteins | Genes | **AQ** | AQ+MES | **MES** | *LPS-Cyto* | *AQ* | *AQ+MES* | *MES* |
| Arachidonate 12-lipoxygenase, 12R-type | ALOX12B | 2.22* | 1.37 | 7.85* | 0.47* | 2.58* | 0.92 | 1.08 |
| Repetin | RPTN | 1.60* | 1.00 | 4.84* | 0.42* | 2.18* | 0.67 | 3.40* |

| | | | | | | | | |
|---|---|---|---|---|---|---|---|---|
| Serine/threonine-protein kinase 31 | STK31 | 2.24* | 1.32 | 4.57* | 0.59* | 2.06* | 0.73 | 1.99* |
| Interferon-related developmental regulator 1 | IFRD1 | 2.60* | 0.80 | 1.96* | 3.12* | 1.60 | 1.01 | 0.77 |
| Apolipoprotein L5 | APOL5 | 1.57* | 0.98 | 1.90* | 1.78* | 1.11 | 1.62 | 1.76 |
| Zinc finger protein 284 | ZNF284 | 1.90* | 1.45 | 1.70* | 1.88* | 0.68 | 0.99 | 2.29* |
| Meiosis regulator and mRNA stability factor 1 | MARF1 | 1.56* | 1.10 | 1.65* | 0.81 | 1.40 | 0.92 | 1.12 |
| Ribonucleoside-diphosphate reductase subunit M2 | RRM2 | 1.74* | 1.38 | 1.58* | 0.79 | 0.67 | 0.60 | 0.55* |
| High affinity cationic amino acid transporter 1 | SLC7A1 | 1.76* | 1.42* | 1.56* | 2.39* | 3.41* | 3.56* | 2.53* |
| Ferredoxin-2, mitochondrial | FDX2 | 1.78* | 1.45 | 1.55* | 1.59* | 2.48* | 1.91* | 1.27 |

***G. Common up-regulated proteins between Mesalamine and Aquamin plus Mesalamine [187 proteins]***

| | | Interventions | | | | | | |
|---|---|---|---|---|---|---|---|---|
| | | Control | | | *With LPS & Cytokines* | | | |
| Proteins | Genes | AQ | **AQ+MES** | **MES** | *LPS-Cyto* | *AQ* | *AQ+MES* | *MES* |
| C-X-C motif chemokine 10 | CXCL10 | 0.72 | 34.05* | 43.44* | 0.97 | 0.83 | 30.10* | 22.62* |
| Immunoglobulin kappa variable 4-1 | IGKV4-1 | 0.63* | 2.29* | 37.17* | 0.69 | 0.85 | 1.24 | 0.95 |
| Immunoglobulin heavy constant gamma 4 | IGHG4 | 0.64* | 1.95* | 32.94* | 0.66* | 0.72 | 1.67* | 0.96 |
| Complement factor H | CFH | 0.62* | 2.03* | 27.05* | 0.74 | 0.62 | 1.68* | 1.72* |
| Immunoglobulin heavy variable 3-49 | IGHV3-49 | 0.94 | 2.96* | 25.72* | 0.91 | 1.41 | 2.01* | 0.78 |
| Immunoglobulin heavy variable 3-7 | IGHV3-7 | 0.93 | 2.12* | 23.53* | 0.83 | 1.48 | 1.53 | 0.58 |
| CD5 antigen-like | CD5L | 0.66 | 1.53 | 23.17* | 0.77 | 0.69 | 0.82 | 0.29* |
| Immunoglobulin heavy constant alpha 2 | IGHA2 | 1.13 | 3.35* | 14.54* | 0.74 | 1.90* | 1.16 | 4.45* |
| Calmodulin-like protein 3 | CALML3 | 1.02 | 52.36* | 12.16* | 0.75 | 1.15 | 0.24* | 0.82 |
| Proline-rich protein 9 | PRR9 | 1.25 | 15.97* | 11.14* | 0.74 | 1.51 | 0.43* | 0.33* |
| Apolipoprotein A-II | APOA2 | 0.90 | 2.24* | 10.88* | 0.84 | 1.17 | 1.89* | 2.53* |
| Retroviral-like aspartic protease 1 | ASPRV1 | 0.92 | 3.16* | 10.29* | 0.39* | 1.54* | 0.49* | 1.97* |
| cAMP-dependent protein kinase inhibitor beta | PKIB | 0.88 | 6.48* | 9.41* | 2.44* | 2.06* | 11.12* | 18.67* |
| Apolipoprotein A-IV | APOA4 | 0.95 | 3.94* | 8.02* | 1.53* | 1.25 | 2.60* | 3.78* |
| Plastin-2 | LCP1 | 0.68* | 2.28* | 7.32* | 0.81 | 0.92 | 1.85* | 2.39* |
| Alpha-2-macroglobulin-like protein 1 | A2ML1 | 1.27* | 2.46* | 5.56* | 0.41* | 1.85* | 0.34* | 1.36* |
| Solute carrier family 2, facilitated glucose transporter member 5 | SLC2A5 | 1.25* | 6.90* | 5.49* | 1.35* | 1.55* | 6.09* | 4.52* |
| Collagen alpha-1(II) chain | COL2A1 | 1.18 | 4.35* | 5.26* | 1.58* | 1.09 | 3.94* | 5.38* |
| Chromogranin-A | CHGA | 0.64* | 5.56* | 5.04* | 0.86 | 1.34 | 4.03* | 6.57* |
| HLA class II histocompatibility antigen, DM alpha chain | HLA-DMA | 1.43 | 5.47* | 4.95* | 1.02 | 1.13 | 5.22* | 4.36* |
| Complement C2 | C2 | 0.70 | 2.82* | 4.91* | 0.68 | 0.79 | 3.36* | 3.92* |
| Immunoglobulin kappa constant | IGKC | 0.71* | 1.56* | 4.90* | 1.34* | 2.92* | 1.88* | 2.00* |
| Galectin-7 | LGALS7 | 0.90 | 4.40* | 4.72* | 1.18 | 2.20* | 0.97 | 1.65 |
| Follistatin-related protein 1 | FSTL1 | 0.95 | 4.04* | 4.57* | 1.47* | 1.61 | 3.52* | 5.73* |

| | | | | | | | | |
|---|---|---|---|---|---|---|---|---|
| Secretogranin-2 | SCG2 | 0.74 | 3.08* | 4.35* | 0.99 | 1.09 | 3.27* | 5.45* |
| Desmoglein-3 | DSG3 | 0.68 | 3.24* | 4.23* | 0.59 | 0.94 | 1.57 | 2.39* |
| Synaptic vesicle membrane protein VAT-1 homolog-like | VAT1L | 0.71 | 3.05* | 4.20* | 0.93 | 1.19 | 2.75* | 4.58* |
| HLA class II histocompatibility antigen, DR beta 4 chain | HLA-DRB4 | 0.71 | 3.84* | 4.17* | 0.84 | 1.08 | 4.68* | 3.60* |
| Centrosomal protein of 85 kDa | CEP85 | 0.93 | 4.39* | 4.17* | 1.04 | 1.23 | 2.87* | 4.16* |
| Receptor-type tyrosine-protein phosphatase zeta | PTPRZ1 | 0.81 | 2.75* | 4.03* | 0.97 | 1.19 | 2.32* | 3.43* |
| Phospholipase A2, membrane associated | PLA2G2A | 1.31 | 4.25* | 3.95* | 3.18* | 8.37* | 15.64* | 17.81* |
| Ferritin light chain | FTL | 0.52* | 2.72* | 3.90* | 1.95* | 0.58 | 4.12* | 9.36* |
| Plasmalemma vesicle-associated protein | PLVAP | 0.75 | 3.13* | 3.90* | 1.02 | 1.13 | 3.00* | 3.92* |
| Ubiquitin D | UBD | 1.03 | 4.25* | 3.84* | 2.84* | 3.54* | 12.39* | 12.06* |
| Complement C3 | C3 | 0.68* | 2.23* | 3.76* | 2.82* | 3.40* | 4.41* | 3.80* |
| EH domain-containing protein 3 | EHD3 | 0.76 | 3.83* | 3.66* | 1.19 | 1.06 | 2.38* | 3.59* |
| Tenascin-X | TNXB | 0.89 | 2.86* | 3.62* | 0.93 | 1.02 | 2.74* | 3.87* |
| 2'-5'-oligoadenylate synthase 2 | OAS2 | 0.80 | 3.38* | 3.47* | 1.71* | 2.58* | 9.01* | 8.23* |
| Beta-parvin | PARVB | 0.82 | 2.48* | 3.45* | 0.96 | 1.17 | 2.19* | 3.29* |
| Guanine nucleotide-binding protein subunit alpha-15 | GNA15 | 0.90 | 2.12* | 3.41* | 0.64 | 1.28 | 1.26 | 1.03 |
| Ferroxidase HEPHL1 | HEPHL1 | 0.77 | 61.07* | 3.28* | 0.93 | 1.02 | 0.31* | 1.91 |
| Moesin | MSN | 0.69* | 1.72* | 3.19* | 1.45* | 1.46* | 2.21* | 3.03* |
| Phospholipid transfer protein | PLTP | 0.70 | 2.51* | 3.16* | 0.97 | 0.87 | 2.48* | 3.00* |
| Cadherin-13 | CDH13 | 0.84 | 2.41* | 3.15* | 0.88 | 1.01 | 2.44* | 3.83* |
| Vasopressin-neurophysin 2-copeptin | AVP | 0.94 | 3.13* | 3.08* | 1.27 | 1.08 | 2.41* | 3.99* |
| Fibroleukin | FGL2 | 0.77 | 3.23* | 3.07* | 0.79 | 0.93 | 3.82* | 3.15* |
| Membrane primary amine oxidase | AOC3 | 0.85 | 2.15* | 3.07* | 0.93 | 1.18 | 1.67 | 2.86* |
| Interleukin-1 receptor accessory protein | IL1RAP | 0.61* | 2.35* | 2.99* | 0.79 | 0.92 | 2.31* | 3.58* |
| cAMP-specific 3',5'-cyclic phosphodiesterase 4C | PDE4C | 0.89 | 2.07* | 2.99* | 0.79 | 1.82* | 2.80* | 3.99* |
| Myeloblastin | PRTN3 | 0.84 | 3.59* | 2.95* | 0.55* | 1.93* | 0.45* | 1.61 |
| Alpha-1-antichymotrypsin | SERPINA3 | 1.08 | 1.90* | 2.88* | 1.28 | 3.87* | 1.63 | 2.12* |
| Plasma kallikrein | KLKB1 | 0.59* | 2.06* | 2.86* | 0.72 | 0.82 | 1.89* | 2.72* |
| Glycolipid transfer protein domain-containing protein 2 | GLTPD2 | 1.47* | 2.85* | 2.85* | 1.93* | 2.81* | 2.12* | 3.41* |
| Selenoprotein P | SELENOP | 0.80 | 1.87* | 2.83* | 0.81 | 1.24 | 1.76* | 1.95* |
| Protein S100-A3 | S100A3 | 1.12 | 26.05* | 2.82* | 0.58* | 1.60 | 1.77 | 3.12* |
| A disintegrin and metalloproteinase with thrombospondin motifs 13 | ADAMTS13 | 0.68 | 2.03* | 2.82* | 0.65 | 0.85 | 2.28* | 2.94* |
| Oncoprotein-induced transcript 3 protein | OIT3 | 0.70* | 2.44* | 2.81* | 0.81 | 0.94 | 2.23* | 3.12* |
| Contactin-1 | CNTN1 | 0.65* | 2.25* | 2.79* | 0.80 | 0.88 | 2.29* | 3.42* |
| Retinol-binding protein 4 | RBP4 | 0.67* | 2.60* | 2.77* | 0.92 | 0.90 | 2.14* | 2.97* |
| Carboxypeptidase A2 | CPA2 | 1.24 | 3.77* | 2.77* | 0.76 | 1.74 | 4.32* | 2.79* |
| Mammaglobin-B | SCGB2A1 | 1.32 | 5.33* | 2.76* | 0.64 | 1.80* | 0.64 | 1.75 |

| | | | | | | | | |
|---|---|---|---|---|---|---|---|---|
| Fibromodulin | FMOD | 0.64* | 2.45* | 2.75* | 1.06 | 0.95 | 2.23* | 3.19* |
| Heparin cofactor 2 | SERPIND1 | 0.75 | 1.73* | 2.75* | 2.04* | 3.83* | 4.33* | 7.38* |
| Collagen alpha-1(XI) chain | COL11A1 | 0.53* | 2.32* | 2.75* | 0.87 | 0.94 | 2.20* | 3.10* |
| Kininogen-1 | KNG1 | 0.47* | 2.27* | 2.73* | 1.71* | 0.68 | 2.29* | 2.32* |
| Alpha-1B-glycoprotein | A1BG | 0.55* | 1.99* | 2.73* | 0.77 | 0.76 | 2.05* | 3.14* |
| Sulfotransferase 2A1 | SULT2A1 | 1.49* | 5.11* | 2.72* | 1.25 | 1.90* | 2.73* | 1.62 |
| All-trans-retinol dehydrogenase [NAD(+)] ADH4 | ADH4 | 1.07 | 2.55* | 2.71* | 2.95* | 2.62* | 2.01* | 1.81* |
| Phosphatidylcholine-sterol acyltransferase | LCAT | 0.65* | 2.10* | 2.67* | 0.76 | 0.94 | 2.06* | 2.94* |
| C-type mannose receptor 2 | MRC2 | 0.88 | 2.19* | 2.65* | 0.96 | 1.06 | 2.07* | 2.88* |
| Interferon-induced protein with tetratricopeptide repeats 3 | IFIT3 | 1.24 | 2.65* | 2.64* | 1.98* | 2.49* | 4.45* | 4.39* |
| Mitochondrial glutamate carrier 2 | SLC25A18 | 0.86 | 1.95* | 2.62* | 0.94 | 1.58 | 2.10* | 3.70* |
| FERM and PDZ domain-containing protein 1 | FRMPD1 | 0.54* | 2.61* | 2.58* | 0.47* | 0.94 | 1.85* | 2.22* |
| Beta-2-glycoprotein 1 | APOH | 1.26 | 2.33* | 2.52* | 2.43* | 2.14* | 2.35* | 3.00* |
| Cerebellin-4 | CBLN4 | 0.41* | 1.58 | 2.52* | 0.57* | 0.78 | 1.73 | 1.95* |
| EGF-containing fibulin-like extracellular matrix protein 1 | EFEMP1 | 0.62* | 2.26* | 2.51* | 0.81 | 0.53* | 1.88* | 2.37* |
| Retinoic acid receptor responder protein 2 | RARRES2 | 0.54* | 1.61* | 2.51* | 0.66 | 0.68 | 1.46 | 2.08* |
| Sex hormone-binding globulin | SHBG | 0.50* | 2.02* | 2.50* | 0.70 | 0.75 | 1.95* | 2.89* |
| Neural cell adhesion molecule 1 | NCAM1 | 0.68* | 2.18* | 2.49* | 0.81 | 0.77 | 2.12* | 3.03* |
| Apolipoprotein C-I | APOC1 | 0.88 | 1.73* | 2.48* | 1.47* | 3.07* | 4.84* | 3.83* |
| Hepatocyte growth factor activator | HGFAC | 0.62* | 2.13* | 2.48* | 0.77* | 0.77 | 2.27* | 3.01* |
| Collagen alpha-2(I) chain | COL1A2 | 0.53* | 2.15* | 2.47* | 0.72 | 0.65 | 1.88* | 2.71* |
| Serum amyloid A-4 protein | SAA4 | 0.63* | 2.09* | 2.47* | 3.32* | 9.01* | 8.67* | 16.73* |
| Carboxypeptidase E | CPE | 0.79 | 2.20* | 2.47* | 0.75 | 0.99 | 1.70 | 2.53* |
| NEDD4 family-interacting protein 2 | NDFIP2 | 1.07 | 2.34* | 2.44* | 0.95 | 1.12 | 2.19* | 2.96* |
| Tyrosine-protein kinase receptor Tie-1 | TIE1 | 0.68 | 1.62* | 2.43* | 0.79 | 0.81 | 2.09* | 2.76* |
| Afamin | AFM | 0.52* | 2.16* | 2.43* | 0.71* | 0.67* | 2.00* | 2.67* |
| Metalloproteinase inhibitor 3 | TIMP3 | 0.80 | 3.49* | 2.39* | 0.78 | 0.88 | 2.09* | 1.51 |
| Latent-transforming growth factor beta-binding protein 4 | LTBP4 | 0.82 | 2.90* | 2.39* | 1.01 | 1.40 | 1.61 | 3.16* |
| Lactotransferrin | LTF | 0.98 | 1.77* | 2.37* | 2.39* | 2.02* | 2.78* | 3.12* |
| Collagen alpha-1(XXI) chain | COL21A1 | 0.75 | 1.78* | 2.36* | 0.97 | 0.96 | 1.81* | 2.34* |
| TRPM8 channel-associated factor 2 | TCAF2 | 1.14 | 1.76* | 2.34* | 1.03 | 1.43 | 1.71 | 1.71 |
| ERC protein 2 | ERC2 | 0.70 | 2.27* | 2.27* | 0.79 | 0.70 | 1.76 | 2.58* |
| Scrapie-responsive protein 1 | SCRG1 | 0.64* | 2.21* | 2.25* | 0.80 | 0.99 | 1.82* | 2.12* |
| Lymphocyte antigen 6D | LY6D | 0.89 | 1.56 | 2.25* | 2.10* | 2.22* | 1.53 | 1.51 |
| Keratin, type I cuticular Ha4 | KRT34 | 0.84 | 63.49* | 2.24* | 0.74 | 1.05 | 0.91 | 3.16* |
| Cytochrome c oxidase assembly protein COX11, mitochondrial | COX11 | 1.28 | 1.96* | 2.23* | 1.39 | 2.04* | 2.52* | 3.10* |

| | | | | | | | | |
|---|---|---|---|---|---|---|---|---|
| Desmoglein-4 | DSG4 | 1.01 | 65.35* | 2.23* | 0.77 | 1.18 | 1.51 | 2.16* |
| Glutathione S-transferase A5 | GSTA5 | 0.75 | 1.99* | 2.22* | 0.78 | 0.90 | 1.92* | 2.62* |
| Collagen alpha-1(III) chain | COL3A1 | 0.82 | 1.61* | 2.22* | 0.72 | 0.87 | 1.66* | 2.04* |
| Pro-opiomelanocortin | POMC | 0.96 | 2.08* | 2.21* | 2.71* | 4.33* | 6.13* | 10.26* |
| Procollagen C-endopeptidase enhancer 1 | PCOLCE | 0.69 | 1.78* | 2.20* | 0.96 | 2.34* | 2.41* | 2.65* |
| Dynein axonemal heavy chain 1 | DNAH1 | 0.51* | 1.54* | 2.15* | 1.10 | 0.62* | 2.04* | 2.51* |
| Complement factor I | CFI | 0.68 | 2.00* | 2.08* | 2.94* | 6.10* | 6.03* | 11.20* |
| Plakophilin-1 | PKP1 | 0.84 | 3.19* | 2.07* | 0.84 | 2.73* | 0.56* | 2.07* |
| Protein piccolo | PCLO | 0.56* | 1.75* | 2.05* | 0.55* | 0.72 | 2.03* | 2.36* |
| Cell adhesion molecule 1 | CADM1 | 0.65* | 1.89* | 2.04* | 0.63 | 0.89 | 1.76* | 2.25* |
| Insulin-like growth factor II | IGF2 | 0.62* | 2.07* | 2.04* | 0.74 | 0.48* | 1.65* | 2.22* |
| Keratin, type I cuticular Ha1 | KRT31 | 0.68* | 49.23* | 2.04* | 0.65* | 0.92 | 0.80 | 2.96* |
| Albumin | ALB | 0.63* | 1.73* | 2.02* | 2.42* | 1.96* | 2.70* | 2.09* |
| Mucin-3A | MUC3A | 1.00 | 2.47* | 2.01* | 1.05 | 1.21 | 1.75* | 1.67* |
| Interferon-induced transmembrane protein 3 | IFITM3 | 0.94 | 2.48* | 1.97* | 1.60* | 2.12* | 4.68* | 3.56* |
| Protein bassoon | BSN | 0.50* | 2.53* | 1.96* | 5.37* | 19.19* | 17.24* | 29.63* |
| Ubiquitin carboxyl-terminal hydrolase 40 | USP40 | 1.12 | 1.64* | 1.96* | 0.80 | 1.08 | 0.97 | 1.03 |
| Inositol-trisphosphate 3-kinase C | ITPKC | 1.17 | 2.04* | 1.95* | 1.21 | 1.37 | 1.86* | 1.46 |
| Secretogranin-3 | SCG3 | 1.21 | 1.79* | 1.95* | 2.47* | 2.79* | 3.40* | 4.48* |
| BPI fold-containing family A member 1 | BPIFA1 | 1.11 | 2.30* | 1.94* | 1.49* | 2.48* | 1.40 | 3.23* |
| Retinal dehydrogenase 2 | ALDH1A2 | 0.81 | 1.80* | 1.94* | 0.84 | 1.03 | 1.97* | 2.31* |
| Tropomodulin-2 | TMOD2 | 0.97 | 2.08* | 1.93* | 2.25* | 1.95* | 2.91* | 4.70* |
| Keratin, type I cuticular Ha5 | KRT35 | 1.08 | 26.02* | 1.92* | 0.71 | 0.74 | 0.68 | 0.97 |
| Transcription cofactor vestigial-like protein 4 | VGLL4 | 0.92 | 1.59* | 1.92* | 0.98 | 0.94 | 0.98 | 0.60 |
| mRNA-decapping enzyme 1B | DCP1B | 1.26 | 2.01* | 1.92* | 0.58* | 1.35 | 0.76 | 1.52 |
| Decreased expression in renal and prostate cancer protein | DERPC | 1.02 | 1.57 | 1.92* | 1.41 | 1.36 | 1.21 | 0.86 |
| EF-hand domain-containing protein D1 | EFHD1 | 0.66 | 43.85* | 1.91* | 0.69 | 0.69 | 0.84 | 3.11* |
| Hemoglobin subunit beta | HBB | 1.03 | 2.23* | 1.89* | 2.18* | 2.36* | 3.19* | 4.34* |
| Guanidinoacetate N-methyltransferase | GAMT | 0.55* | 1.98* | 1.89* | 0.67 | 0.57 | 1.54 | 1.94* |
| Sterile alpha motif domain-containing protein 9-like | SAMD9L | 1.07 | 1.96* | 1.88* | 1.41 | 2.01* | 3.01* | 3.00* |
| Tropomyosin beta chain | TPM2 | 0.68* | 1.91* | 1.88* | 0.86 | 0.83 | 1.59 | 2.29* |
| Polyglutamine-binding protein 1 | PQBP1 | 1.06 | 1.73* | 1.85* | 1.25 | 0.26* | 0.69 | 1.16 |
| Apolipoprotein L1 | APOL1 | 1.46 | 1.73* | 1.82* | 2.68* | 3.15* | 3.75* | 2.64* |
| Lumican | LUM | 0.67* | 1.73* | 1.81* | 1.41* | 1.54* | 2.50* | 3.78* |
| HLA class II histocompatibility antigen, DP beta 1 chain | HLA-DPB1 | 0.31* | 2.33* | 1.80* | 15.14* | 13.55* | 15.68* | 15.01* |
| Nucleolar and spindle-associated protein 1 | NUSAP1 | 1.44* | 1.64* | 1.78* | 1.33 | 0.54 | 0.27* | 0.86 |
| Retinol-binding protein 2 | RBP2 | 1.29* | 2.84* | 1.78* | 1.20 | 2.02* | 1.36* | 1.25* |

| | | | | | | | | |
|---|---|---|---|---|---|---|---|---|
| Secreted and transmembrane protein 1 | SECTM1 | 1.01 | 1.76* | 1.78* | 14.70* | 8.17* | 7.82* | 13.21* |
| Collagen alpha-1(V) chain | COL5A1 | 1.02 | 1.90* | 1.77* | 1.10 | 1.65* | 2.12* | 2.62* |
| Desumoylating isopeptidase 1 | DESI1 | 0.95 | 1.72* | 1.76* | 1.06 | 1.21 | 1.64 | 2.27* |
| Inter-alpha-trypsin inhibitor heavy chain H2 | ITIH2 | 0.86 | 1.71* | 1.75* | 1.61* | 2.05* | 2.42* | 3.06* |
| CD166 antigen | ALCAM | 1.12 | 1.95* | 1.74* | 2.10* | 1.22 | 2.14* | 1.78* |
| Sulfhydryl oxidase 2 | QSOX2 | 1.47* | 1.88* | 1.74* | 2.20* | 3.05* | 3.18* | 3.04* |
| Microtubule-associated proteins 1A/1B light chain 3 beta 2 | MAP1LC3B2 | 1.26 | 1.67* | 1.73* | 1.34 | 1.32 | 1.61 | 1.83* |
| Transforming growth factor-beta-induced protein ig-h3 | TGFBI | 0.93 | 1.95* | 1.72* | 1.09 | 1.36 | 1.69* | 2.50* |
| Opioid growth factor receptor | OGFR | 1.32* | 2.03* | 1.72* | 1.66* | 1.39 | 1.91* | 2.03* |
| DNA-binding protein SMUBP-2 | IGHMBP2 | 0.58* | 1.53* | 1.72* | 2.47* | 3.42* | 3.38* | 5.07* |
| Mammaglobin-A | SCGB2A2 | 1.06 | 1.93* | 1.71* | 1.32 | 4.09* | 1.54 | 3.31* |
| Keratin, type I cuticular Ha3-I | KRT33A | 0.87 | 24.84* | 1.71* | 0.94 | 0.91 | 1.29 | 2.44* |
| Neugrin | NGRN | 1.04 | 1.57* | 1.71* | 1.11 | 0.65 | 0.56* | 0.90 |
| Sushi domain-containing protein 2 | SUSD2 | 1.30 | 2.95* | 1.70* | 1.09 | 1.57 | 1.60 | 0.92 |
| SPARC-related modular calcium-binding protein 2 | SMOC2 | 0.84 | 1.62* | 1.69* | 1.38 | 2.05* | 1.83* | 1.95* |
| Transgelin | TAGLN | 0.82 | 2.04* | 1.68* | 1.62* | 1.71 | 2.44* | 2.91* |
| Cornifin-B | SPRR1B | 0.79 | 1.62* | 1.68* | 1.14 | 4.30* | 0.46* | 3.71* |
| ER degradation-enhancing alpha-mannosidase-like protein 2 | EDEM2 | 1.23 | 1.67* | 1.68* | 1.01 | 1.30 | 1.56 | 1.89* |
| Alpha-2-antiplasmin | SERPINF2 | 0.86 | 1.62* | 1.67* | 1.84* | 2.47* | 5.09* | 6.18* |
| Fibrinogen beta chain | FGB | 0.56* | 2.04* | 1.67* | 1.92* | 3.53* | 3.54* | 6.17* |
| Exocyst complex component 3-like protein 4 | EXOC3L4 | 0.82 | 1.70* | 1.67* | 0.79 | 0.71 | 1.71* | 1.62 |
| F-box only protein 2 | FBXO2 | 1.08 | 2.13* | 1.67* | 0.90 | 1.73* | 2.19* | 1.63 |
| Laminin subunit alpha-1 | LAMA1 | 0.55* | 2.01* | 1.66* | 1.60* | 2.30* | 2.88* | 3.96* |
| Lymphocyte function-associated antigen 3 | CD58 | 1.37* | 1.92* | 1.66* | 1.76* | 2.20* | 2.42* | 2.22* |
| Glutamyl aminopeptidase | ENPEP | 0.97 | 2.32* | 1.66* | 1.38* | 1.43* | 1.34 | 0.94 |
| Apolipoprotein C-III | APOC3 | 0.91 | 2.22* | 1.66* | 2.09* | 2.28* | 2.65* | 3.96* |
| Laminin subunit beta-1 | LAMB1 | 0.54* | 2.04* | 1.65* | 1.58* | 2.39* | 2.83* | 3.78* |
| Protransforming growth factor alpha | TGFA | 1.48* | 1.58* | 1.65* | 1.77* | 3.24* | 3.42* | 3.06* |
| Ileal sodium/bile acid cotransporter | SLC10A2 | 1.33* | 2.98* | 1.64* | 1.53* | 1.67* | 2.16* | 1.42 |
| MARVEL domain-containing protein 3 | MARVELD3 | 1.31 | 1.82* | 1.63* | 1.72* | 0.97 | 1.89* | 1.56 |
| Laminin subunit gamma-1 | LAMC1 | 0.61* | 1.97* | 1.62* | 1.54* | 2.28* | 2.68* | 3.65* |
| Ethanolaminephosphotransferase 1 | SELENOI | 1.27 | 2.10* | 1.62* | 0.85 | 1.42 | 1.42 | 1.22 |
| Nidogen-2 | NID2 | 0.73 | 1.89* | 1.61* | 1.45* | 2.02* | 2.55* | 3.43* |
| DnaJ homolog subfamily C member 15 | DNAJC15 | 1.21 | 1.83* | 1.61* | 2.28* | 3.11* | 2.09* | 2.92* |
| Trophoblast glycoprotein | TPBG | 1.42* | 1.68* | 1.61* | 1.90* | 2.67* | 2.69* | 1.69 |
| Dual oxidase maturation factor 2 | DUOXA2 | 1.45* | 1.75* | 1.60* | 1.11 | 1.43 | 1.63* | 1.04 |

| | | | | | | | | |
|---|---|---|---|---|---|---|---|---|
| Protein preY, mitochondrial | PYURF | 1.41* | 1.93* | 1.58* | 1.27 | 1.06 | 1.27 | 1.34 |
| N-acetyllactosaminide beta-1,3-N-acetylglucosaminyltransferase 2 | B3GNT2 | 1.29 | 1.91* | 1.58* | 1.36 | 2.26* | 2.13* | 2.17* |
| Leukocyte receptor cluster member 8 | LENG8 | 1.48* | 1.79* | 1.57* | 1.20 | 0.21* | 0.51* | 0.13* |
| Collagen alpha-1(XII) chain | COL12A1 | 0.71 | 1.58 | 1.57* | 2.12* | 3.69* | 6.05* | 6.67* |
| Renin | REN | 1.34 | 1.56* | 1.57* | 0.66 | 0.96 | 0.87 | 0.83 |
| Growth-regulated alpha protein | CXCL1 | 0.91 | 1.50 | 1.56* | 1.02 | 1.15 | 1.36 | 1.44 |
| 2'-5'-oligoadenylate synthase-like protein | OASL | 1.25 | 1.70* | 1.54* | 1.09 | 1.26 | 1.71* | 1.54 |
| Coiled-coil domain-containing protein 127 | CCDC127 | 1.41 | 1.65* | 1.53 | 1.85* | 2.71* | 2.49* | 2.29* |
| Inositol-3-phosphate synthase 1 | ISYNA1 | 1.33* | 1.61* | 1.53* | 2.28* | 3.96* | 2.92* | 3.27* |
| Succinate dehydrogenase assembly factor 1, mitochondrial | SDHAF1 | 1.26 | 1.62* | 1.52 | 1.27 | 0.77 | 0.74 | 1.25 |
| Hemoglobin subunit alpha | HBA1 | 0.78* | 1.53* | 1.52* | 1.62* | 1.74* | 2.44* | 3.35* |
| HLA class I histocompatibility antigen, B alpha chain | HLA-B | 1.47* | 2.37* | 1.52* | 7.62* | 5.91* | 7.94* | 8.75* |
| Cysteine protease ATG4C | ATG4C | 1.22 | 1.51 | 1.51* | 1.04 | 0.50* | 0.27* | 0.39* |
| Periostin | POSTN | 0.66* | 1.63* | 1.51* | 1.02 | 2.43* | 2.54* | 3.38* |
| Keratinocyte-associated transmembrane protein 2 | KCT2 | 1.12 | 1.69* | 1.51* | 2.82* | 2.19* | 2.57* | 2.90* |
| Ankyrin repeat domain-containing protein 27 | ANKRD27 | 1.10 | 1.61* | 1.51* | 1.81* | 1.46* | 1.06 | 1.92* |
| Solute carrier family 66 member 3 | SLC66A3 | 1.36 | 2.04* | 1.50* | 0.98 | 1.35 | 1.63 | 1.02 |

Values represent the abundance ratio from organoids (n=4 subjects) compared to the control. These proteins were up-regulated at a 1.5-fold change (<2% FDR). Corresponding abundance ratios from the other treatment groups are provided for comparison. Proteins common among groups and unique to individual groups under control conditions are presented. *Indicates significance compared to the control (at p<0.05).
