## Supplementary material for "Proteomic Profile of Human Colon Organoids: Effects of a multi-mineral intervention alone and in the presence of pro-Inflammatory and anti-inflammatory treatments": S Table 4

**Supplement Table 4. Down-regulated proteins influenced by Aquamin and Mesalamine under control conditions (with 1.5-fold).**

***A. Common among three groups – Aquamin (AQ), Mesalamine (MES) and Aquamin plus Mesalamine (AQ+MES) [194 proteins]***

| | | Interventions | | | | | | |
|---|---|---|---|---|---|---|---|---|
| | | Control | | | *With LPS & Cytokines* | | | |
| Proteins | Genes | **AQ** | **AQ+MES** | **MES** | *LPS-Cyto* | *AQ* | *AQ+MES* | *MES* |
| Fermitin family homolog 2 | FERMT2 | 0.05* | 0.35* | 0.25* | 2.05* | 9.27* | 9.50* | 10.36* |
| Tubulin alpha-3C chain | TUBA3C | 0.07* | 0.27* | 0.11* | 1.78* | 10.04* | 7.83* | 8.39* |
| Telomere-associated protein RIF1 | RIF1 | 0.11* | 0.18* | 0.18* | 0.26* | 0.35* | 0.29* | 0.28* |
| Calcium/calmodulin-dependent protein kinase type II subunit beta | CAMK2B | 0.12* | 0.10* | 0.17* | 0.26* | 0.64* | 0.53* | 0.47* |
| Aldehyde oxidase | AOX1 | 0.14* | 0.14* | 0.27* | 2.11* | 11.69* | 8.47* | 9.08* |
| Survival motor neuron protein | SMN1 | 0.18* | 0.25* | 0.29* | 0.40* | 0.38* | 0.38* | 0.43* |
| Rho guanine nucleotide exchange factor 3 | ARHGEF3 | 0.18* | 0.44* | 0.36* | 0.12* | 0.48* | 0.17* | 0.24* |
| Heparan sulfate glucosamine 3-O-sulfotransferase 1 | HS3ST1 | 0.19* | 0.35* | 0.29* | 1.17 | 4.51* | 3.55* | 5.50* |
| Kelch-like ECH-associated protein 1 | KEAP1 | 0.19* | 0.33* | 0.37* | 0.18* | 0.39* | 0.32* | 0.29* |
| Rho GTPase-activating protein 29 | ARHGAP29 | 0.19* | 0.35* | 0.41* | 0.33* | 0.58 | 0.50* | 0.35* |
| Zinc finger protein 654 | ZNF654 | 0.19* | 0.24* | 0.21* | 0.15* | 0.36* | 0.33* | 0.29* |
| Gamma-tubulin complex component 3 | TUBGCP3 | 0.19* | 0.27* | 0.29* | 0.31* | 0.86 | 0.80 | 0.82 |
| Zinc finger protein 136 | ZNF136 | 0.20* | 0.18* | 0.20* | 0.20* | 0.26* | 0.23* | 0.17* |
| TBC1 domain family member 25 | TBC1D25 | 0.21* | 0.09* | 0.08* | 0.10* | 0.11* | 0.15* | 0.07* |
| Keratin, type II cytoskeletal 2 epidermal | KRT2 | 0.21* | 0.26* | 0.47* | 0.33* | 0.50* | 0.23* | 0.35* |
| Tubulin beta-1 chain | TUBB1 | 0.21* | 0.28* | 0.24* | 1.96* | 9.06* | 7.77* | 9.62* |
| Leydig cell tumor 10 kDa protein homolog | C19orf53 | 0.21* | 0.19* | 0.20* | 0.43* | 0.37* | 0.36* | 0.45* |
| Bone morphogenetic protein 1 | BMP1 | 0.22* | 0.37* | 0.31* | 0.91 | 3.52* | 2.50* | 3.63* |
| Ribosomal biogenesis protein LAS1L | LAS1L | 0.22* | 0.26* | 0.25* | 0.28* | 0.62* | 0.51* | 0.40* |
| WD repeat and FYVE domain-containing protein 3 | WDFY3 | 0.23* | 0.09* | 0.12* | 0.09* | 0.07* | 0.18* | 0.06* |
| Insulin-like growth factor-binding protein complex acid labile subunit | IGFALS | 0.23* | 0.37* | 0.33* | 2.38* | 12.16* | 9.41* | 11.08* |
| Guanine nucleotide-binding protein-like 3-like protein | GNL3L | 0.23* | 0.38* | 0.37* | 0.14* | 0.22* | 0.31* | 0.20* |
| AT-rich interactive domain-containing protein 2 | ARID2 | 0.24* | 0.38* | 0.30* | 0.30* | 0.48* | 0.35* | 0.48* |
| Filaggrin-2 | FLG2 | 0.24* | 0.30* | 0.44* | 0.94 | 0.69* | 0.34* | 0.69* |
| Mediator of RNA polymerase II transcription subunit 12 | MED12 | 0.25* | 0.27* | 0.31* | 0.42* | 0.97 | 0.81 | 0.84 |
| Zinc finger protein 592 | ZNF592 | 0.25* | 0.28* | 0.27* | 0.33* | 0.59 | 0.55 | 0.49* |
| Histone-lysine N-methyltransferase EHMT2 | EHMT2 | 0.25* | 0.28* | 0.29* | 0.40* | 1.50 | 1.10 | 1.16 |
| Zinc finger and BTB domain-containing protein 11 | ZBTB11 | 0.25* | 0.37* | 0.31* | 0.47* | 0.42* | 0.29* | 0.49* |

| | | | | | | | | |
|---|---|---|---|---|---|---|---|---|
| Cap-specific mRNA (nucleoside-2'-O-)-methyltransferase 2 | CMTR2 | 0.25* | 0.45* | 0.57* | 0.26* | 0.82 | 0.62 | 0.87 |
| Protein kinase C theta type | PRKCQ | 0.26* | 0.08* | 0.11* | 0.16* | 0.14* | 0.22* | 0.11* |
| Zinc finger protein with KRAB and SCAN domains 4 | ZKSCAN4 | 0.27* | 0.19* | 0.22* | 0.15* | 0.37* | 0.19* | 0.22* |
| Activating transcription factor 7-interacting protein 1 | ATF7IP | 0.27* | 0.43* | 0.38* | 0.47* | 0.46* | 0.50* | 0.46* |
| Integrin alpha-E | ITGAE | 0.27* | 0.09* | 0.14* | 0.13* | 0.10* | 0.22* | 0.09* |
| Mediator of RNA polymerase II transcription subunit 23 | MED23 | 0.28* | 0.39* | 0.32* | 0.32* | 0.85 | 0.69 | 0.70 |
| Protein ECT2 | ECT2 | 0.28* | 0.38* | 0.40* | 0.25* | 0.64 | 0.62 | 0.55* |
| Oxytocin-neurophysin 1 | OXT | 0.28* | 0.52* | 0.47* | 0.20* | 0.20* | 0.50* | 0.37* |
| Structural maintenance of chromosomes protein 6 | SMC6 | 0.28* | 0.28* | 0.27* | 0.41* | 1.10 | 0.92 | 1.02 |
| All-trans-retinol dehydrogenase [NAD(+)] ADH7 | ADH7 | 0.29* | 0.43* | 0.31* | 2.22* | 11.41* | 8.22* | 10.30* |
| Trafficking kinesin-binding protein 1 | TRAK1 | 0.29* | 0.38* | 0.48* | 0.36* | 0.46* | 0.61 | 0.58 |
| Primary cilium assembly protein FAM149B1 | FAM149B1 | 0.29* | 0.24* | 0.24* | 0.30* | 0.85 | 0.39* | 0.60 |
| SURP and G-patch domain-containing protein 2 | SUGP2 | 0.29* | 0.33* | 0.39* | 0.28* | 0.24* | 0.23* | 0.19* |
| Structural maintenance of chromosomes protein 5 | SMC5 | 0.30* | 0.36* | 0.38* | 0.34* | 0.60 | 0.46* | 0.61* |
| Tensin-4 | TNS4 | 0.30* | 0.47* | 0.46* | 0.43* | 0.67 | 0.52* | 0.45* |
| Transformation/transcription domain-associated protein | TRRAP | 0.30* | 0.35* | 0.40* | 0.54* | 1.21 | 1.16 | 1.04 |
| Biogenesis of lysosome-related organelles complex 1 subunit 3 | BLOC1S3 | 0.30* | 0.42* | 0.36* | 0.49* | 0.50* | 0.39* | 0.62 |
| Betaine--homocysteine S-methyltransferase 1 | BHMT | 0.31* | 0.41* | 0.31* | 2.44* | 13.21* | 10.27* | 6.92* |
| General transcription factor 3C polypeptide 3 | GTF3C3 | 0.31* | 0.41* | 0.39* | 0.71 | 0.94 | 0.93 | 0.91 |
| E3 SUMO-protein ligase ZNF451 | ZNF451 | 0.32* | 0.37* | 0.35* | 0.19* | 0.43* | 0.44* | 0.36* |
| E3 ubiquitin-protein ligase Midline-1 | MID1 | 0.32* | 0.37* | 0.40* | 0.41* | 0.38* | 0.40* | 0.35* |
| Echinoderm microtubule-associated protein-like 3 | EML3 | 0.32* | 0.50* | 0.48* | 0.47* | 0.21* | 0.10* | 0.30* |
| PHD finger protein 6 | PHF6 | 0.32* | 0.43* | 0.46* | 0.26* | 0.65* | 0.64* | 0.69* |
| Anaphase-promoting complex subunit 1 | ANAPC1 | 0.33* | 0.29* | 0.32* | 0.60* | 2.35* | 1.97* | 2.10* |
| Probable ATP-dependent RNA helicase DDX20 | DDX20 | 0.33* | 0.54* | 0.49* | 0.51* | 0.75 | 0.70 | 0.73 |
| Matrix metalloproteinase-28 | MMP28 | 0.33* | 0.49* | 0.56* | 0.80 | 3.03* | 2.32* | 2.79* |
| Gametogenetin-binding protein 2 | GGNBP2 | 0.34* | 0.37* | 0.50* | 0.34* | 0.51* | 0.46* | 0.40* |
| Transmembrane protein 209 | TMEM209 | 0.34* | 0.43* | 0.44* | 0.42* | 1.00 | 0.80 | 0.87 |
| DNA topoisomerase 2-binding protein 1 | TOPBP1 | 0.34* | 0.50* | 0.60* | 0.31* | 0.44* | 0.44* | 0.51* |
| Helicase-like transcription factor | HLTF | 0.34* | 0.41* | 0.37* | 0.28* | 0.52 | 0.63 | 0.51 |
| Protein furry homolog | FRY | 0.34* | 0.64* | 0.59* | 0.65 | 0.30* | 0.36* | 0.55 |
| Centrosomal protein of 131 kDa | CEP131 | 0.35* | 0.36* | 0.35* | 0.21* | 0.33* | 0.09* | 0.09* |
| Parathyroid hormone/parathyroid hormone-related peptide receptor | PTH1R | 0.35* | 0.33* | 0.47* | 1.60* | 8.24* | 7.16* | 7.08* |
| Endothelial lipase | LIPG | 0.35* | 0.46* | 0.43* | 1.12 | 4.07* | 3.10* | 3.50* |
| NEDD4-binding protein 3 | N4BP3 | 0.35* | 0.47* | 0.43* | 0.20* | 0.36* | 0.29* | 0.26* |

| | | | | | | | | |
|---|---|---|---|---|---|---|---|---|
| HAUS augmin-like complex subunit 3 | HAUS3 | 0.35* | 0.39* | 0.40* | 0.45* | 0.66 | 0.62 | 0.59 |
| Molybdenum cofactor sulfurase | MOCOS | 0.36* | 0.52* | 0.58* | 0.29* | 0.24* | 0.26* | 0.25* |
| Nuclear receptor corepressor 1 | NCOR1 | 0.36* | 0.52* | 0.51* | 0.38* | 0.32* | 0.33* | 0.51* |
| C-X-C motif chemokine 14 | CXCL14 | 0.36* | 0.37* | 0.42* | 0.28* | 0.28* | 0.42* | 0.39* |
| Keratin, type II cytoskeletal 71 | KRT71 | 0.36* | 0.52* | 0.41* | 0.94 | 1.15 | 0.39* | 0.51* |
| E3 ubiquitin-protein transferase MAEA | MAEA | 0.36* | 0.45* | 0.47* | 0.45* | 0.72 | 0.67 | 0.61* |
| Beclin-1 | BECN1 | 0.36* | 0.54* | 0.52* | 0.41* | 0.67 | 0.66 | 0.61* |
| WD repeat-containing protein 3 | WDR3 | 0.37* | 0.34* | 0.36* | 0.43* | 0.82 | 0.73 | 0.70* |
| AP-4 complex subunit beta-1 | AP4B1 | 0.37* | 0.55* | 0.63* | 0.50* | 0.67 | 0.68 | 0.55 |
| TBC1 domain family member 2B | TBC1D2B | 0.37* | 0.52* | 0.55* | 0.35* | 0.89 | 0.73 | 0.60 |
| Nucleolar MIF4G domain-containing protein 1 | NOM1 | 0.38* | 0.40* | 0.40* | 0.60* | 0.37* | 0.27* | 0.32* |
| Small subunit processome component 20 homolog | UTP20 | 0.39* | 0.36* | 0.37* | 0.42* | 0.86 | 0.84 | 1.24 |
| Serine/threonine-protein kinase tousled-like 2 | TLK2 | 0.39* | 0.59* | 0.61* | 0.53* | 0.32* | 0.30* | 0.34* |
| Liprin-alpha-4 | PPFIA4 | 0.39* | 0.45* | 0.66* | 1.37 | 0.17* | 0.20* | 0.32* |
| Polyamine-modulated factor 1-binding protein 1 | PMFBP1 | 0.39* | 0.15* | 0.13* | 0.14* | 0.15* | 0.25* | 0.08* |
| Myotubularin-related protein 13 | SBF2 | 0.39* | 0.59* | 0.60* | 0.32* | 0.67 | 0.42* | 0.40* |
| Anaphase-promoting complex subunit 5 | ANAPC5 | 0.39* | 0.38* | 0.39* | 0.53* | 1.56* | 1.01 | 1.02 |
| Integrator complex subunit 4 | INTS4 | 0.39* | 0.43* | 0.46* | 0.61* | 1.10 | 1.22 | 1.14 |
| Transcription factor Sp1 | SP1 | 0.39* | 0.49* | 0.53* | 0.26* | 0.40* | 0.44* | 0.46* |
| Integrator complex subunit 5 | INTS5 | 0.39* | 0.34* | 0.33* | 0.35* | 0.86 | 0.81 | 0.84 |
| Deoxycytidine kinase | DCK | 0.40* | 0.45* | 0.56* | 0.34* | 0.63 | 0.57* | 0.54* |
| PCNA-interacting partner | PARPBP | 0.40* | 0.11* | 0.08* | 0.13* | 0.12* | 0.16* | 0.06* |
| Probable protein phosphatase 1N | PPM1N | 0.40* | 0.19* | 0.23* | 0.27* | 0.19* | 0.42* | 0.17* |
| Baculoviral IAP repeat-containing protein 2 | BIRC2 | 0.41* | 0.58* | 0.49* | 0.57* | 1.37 | 1.08 | 1.31 |
| Keratin, type II cytoskeletal 1 | KRT1 | 0.42* | 0.49* | 0.57* | 1.10 | 1.59* | 0.44* | 1.02 |
| Uncharacterized protein C2orf42 | C2orf42 | 0.42* | 0.67* | 0.53* | 0.45* | 0.41* | 0.45* | 0.28* |
| Mitotic deacetylase-associated SANT domain protein | MIDEAS | 0.42* | 0.58* | 0.54* | 0.52* | 0.31* | 0.47* | 0.43* |
| General transcription factor 3C polypeptide 1 | GTF3C1 | 0.42* | 0.44* | 0.37* | 0.48* | 0.68 | 0.68 | 0.43* |
| E3 ubiquitin-protein ligase NRDP1 | RNF41 | 0.43* | 0.55* | 0.45* | 0.33* | 0.81 | 0.61 | 0.52 |
| Circadian locomoter output cycles protein kaput | CLOCK | 0.43* | 0.48* | 0.56* | 0.34* | 0.60 | 0.53* | 0.46* |
| Transducin beta-like protein 3 | TBL3 | 0.43* | 0.52* | 0.55* | 0.64* | 0.60 | 0.68 | 0.74 |
| Bromodomain adjacent to zinc finger domain protein 1A | BAZ1A | 0.43* | 0.62* | 0.56* | 0.43* | 0.58 | 0.43* | 0.43* |
| Zinc finger protein 701 | ZNF701 | 0.43* | 0.34* | 0.36* | 0.45* | 0.46* | 0.42* | 0.48* |
| Protein Wiz | WIZ | 0.43* | 0.61* | 0.61* | 0.58* | 0.15* | 0.17* | 0.32* |
| Mitogen-activated protein kinase kinase kinase 20 | MAP3K20 | 0.44* | 0.46* | 0.61* | 0.40* | 0.79 | 0.65 | 0.61* |
| Translation machinery-associated protein 7 | TMA7 | 0.44* | 0.46* | 0.65* | 0.80 | 0.44* | 0.40* | 0.71 |
| WD repeat-containing protein 37 | WDR37 | 0.44* | 0.58* | 0.50* | 0.48* | 0.72 | 0.69 | 0.64 |

| | | | | | | | | |
|---|---|---|---|---|---|---|---|---|
| Putative ATP-dependent RNA helicase DHX57 | DHX57 | 0.45* | 0.47* | 0.46* | 0.30* | 0.71 | 0.63 | 0.46* |
| Sin3 histone deacetylase corepressor complex component SDS3 | SUDS3 | 0.45* | 0.57* | 0.52* | 0.61* | 0.92 | 0.93 | 0.83 |
| Methylosome subunit pICln | CLNS1A | 0.45* | 0.50* | 0.51* | 0.84 | 0.84 | 0.80 | 1.06 |
| Transcription initiation factor TFIID subunit 5 | TAF5 | 0.45* | 0.59* | 0.49* | 0.70 | 1.60 | 1.45 | 1.68* |
| Caspase-14 | CASP14 | 0.45* | 0.59* | 0.53* | 1.31* | 0.79 | 0.78 | 1.12 |
| Serine/threonine-protein phosphatase 6 regulatory ankyrin repeat subunit A | ANKRD28 | 0.46* | 0.44* | 0.46* | 0.44* | 0.67 | 0.55 | 0.51* |
| Inactive ubiquitin carboxyl-terminal hydrolase 53 | USP53 | 0.46* | 0.63* | 0.51* | 0.31* | 0.32* | 0.36* | 0.22* |
| Keratin, type I cytoskeletal 9 | KRT9 | 0.46* | 0.44* | 0.46* | 1.11 | 1.99* | 0.51* | 0.89 |
| Structural maintenance of chromosomes protein 4 | SMC4 | 0.46* | 0.45* | 0.55* | 0.46* | 1.22 | 1.09 | 0.96 |
| Integrator complex subunit 7 | INTS7 | 0.46* | 0.58* | 0.57* | 0.49* | 1.12 | 0.88 | 0.83 |
| Ubiquitin-associated and SH3 domain-containing protein B | UBASH3B | 0.46* | 0.57* | 0.63* | 0.43* | 0.52* | 0.51* | 0.44* |
| NFX1-type zinc finger-containing protein 1 | ZNFX1 | 0.46* | 0.56* | 0.59* | 0.49* | 0.55* | 0.63 | 0.56* |
| Zinc finger protein 22 | ZNF22 | 0.46* | 0.37* | 0.55* | 0.77 | 0.49* | 0.35* | 0.36* |
| E3 ubiquitin-protein ligase MSL2 | MSL2 | 0.47* | 0.40* | 0.51* | 2.74* | 11.31* | 10.04* | 11.22* |
| ATPase family gene 2 protein homolog B | AFG2B | 0.47* | 0.50* | 0.62* | 0.54* | 0.54 | 0.54 | 0.60 |
| RRP12-like protein | RRP12 | 0.47* | 0.46* | 0.54* | 0.47* | 0.45* | 0.46* | 0.49* |
| Lysine-specific demethylase 9 | RSBN1 | 0.47* | 0.57* | 0.46* | 0.83 | 0.08* | 0.42* | 0.30* |
| Neurabin-1 | PPP1R9A | 0.47* | 0.58* | 0.64* | 0.46* | 0.49* | 0.39* | 0.47* |
| Alpha-endosulfine | ENSA | 0.47* | 0.44* | 0.55* | 0.53* | 0.36* | 0.35* | 0.55* |
| Mismatch repair endonuclease PMS2 | PMS2 | 0.47* | 0.47* | 0.58* | 0.54* | 1.06 | 0.65 | 0.77 |
| Mitogen-activated protein kinase 7 | MAPK7 | 0.47* | 0.62* | 0.67* | 0.45* | 0.68 | 0.63 | 0.63 |
| Probable JmjC domain-containing histone demethylation protein 2C | JMJD1C | 0.47* | 0.42* | 0.33* | 0.50* | 0.72 | 0.42* | 0.29* |
| Calcium-binding and coiled-coil domain-containing protein 2 | CALCOCO2 | 0.47* | 0.60* | 0.63* | 0.55* | 0.31* | 0.29* | 0.33* |
| DNA (cytosine-5)-methyltransferase 1 | DNMT1 | 0.48* | 0.32* | 0.41* | 0.70 | 3.00* | 2.01* | 2.38* |
| Loricrin | LORICRIN | 0.48* | 0.44* | 0.64* | 1.79* | 1.63* | 0.30* | 1.10 |
| RING finger and CHY zinc finger domain-containing protein 1 | RCHY1 | 0.48* | 0.63* | 0.65* | 0.35* | 0.05* | 0.12* | 0.05* |
| Probable ATP-dependent RNA helicase DDX56 | DDX56 | 0.48* | 0.51* | 0.57* | 0.42* | 0.34* | 0.34* | 0.30* |
| Cytosolic iron-sulfur assembly component 3 | CIAO3 | 0.48* | 0.47* | 0.55* | 0.58* | 0.69 | 0.48* | 0.58* |
| Keratin, type I cytoskeletal 27 | KRT27 | 0.48* | 0.64 | 0.52* | 1.06 | 0.68 | 0.48* | 1.03 |
| Zinc finger CCCH domain-containing protein 8 | ZC3H8 | 0.48* | 0.52* | 0.58* | 0.44* | 0.51* | 0.43* | 0.45* |
| DNA-directed RNA polymerase III subunit RPC1 | POLR3A | 0.49* | 0.56* | 0.66 | 0.97 | 1.00 | 0.97 | 1.20 |
| DmX-like protein 2 | DMXL2 | 0.50* | 0.46* | 0.41* | 0.44* | 0.53* | 0.52* | 0.44* |
| Vam6/Vps39-like protein | VPS39 | 0.50* | 0.46* | 0.46* | 0.37* | 0.63 | 0.54* | 0.58* |

| | | | | | | | | |
|---|---|---|---|---|---|---|---|---|
| TATA-binding protein-associated factor 172 | BTAF1 | 0.50* | 0.62* | 0.58* | 0.51* | 0.58 | 0.54 | 0.52* |
| NF-X1-type zinc finger protein NFXL1 | NFXL1 | 0.50* | 0.58* | 0.58* | 0.40* | 0.39* | 0.35* | 0.40* |
| Macrophage mannose receptor 1 | MRC1 | 0.51* | 0.40* | 0.41* | 1.72* | 6.02* | 7.32* | 7.08* |
| Replication factor C subunit 5 | RFC5 | 0.51* | 0.49* | 0.62* | 0.29* | 0.55* | 0.70 | 0.43* |
| Ribonuclease P protein subunit p29 | POP4 | 0.51* | 0.61* | 0.62* | 0.63* | 1.31 | 1.21 | 1.20 |
| E3 ubiquitin-protein ligase RNF113A | RNF113A | 0.51* | 0.59* | 0.63* | 0.48* | 0.18* | 0.21* | 0.31* |
| Ephrin type-B receptor 3 | EPHB3 | 0.51* | 0.62* | 0.60* | 1.22 | 4.74* | 3.26* | 4.09* |
| Keratin, type I cytoskeletal 14 | KRT14 | 0.51* | 0.49* | 0.57* | 1.84* | 0.71* | 0.52* | 0.61* |
| TNF receptor-associated factor 2 | TRAF2 | 0.52* | 0.60* | 0.58* | 0.57* | 0.95 | 0.93 | 0.89 |
| AN1-type zinc finger protein 1 | ZFAND1 | 0.53* | 0.50* | 0.58* | 0.40* | 0.41* | 0.34* | 0.71 |
| Kinetochore-associated protein NSL1 homolog | NSL1 | 0.53* | 0.63* | 0.65* | 0.56* | 0.45* | 0.26* | 0.50* |
| SUN domain-containing protein 1 | SUN1 | 0.53* | 0.64* | 0.58* | 0.57* | 0.65 | 0.69 | 0.49* |
| BTB/POZ domain-containing protein KCTD3 | KCTD3 | 0.53* | 0.58* | 0.51* | 0.54* | 0.97 | 1.07 | 0.69 |
| DNA replication licensing factor MCM5 | MCM5 | 0.54* | 0.55* | 0.65* | 0.72 | 0.60 | 0.67 | 1.03 |
| Remodeling and spacing factor 1 | RSF1 | 0.54* | 0.63* | 0.57* | 0.64* | 0.40* | 0.41* | 0.53* |
| E3 ubiquitin-protein ligase TRIM32 | TRIM32 | 0.54* | 0.56* | 0.56* | 0.48* | 0.95 | 0.87 | 0.72 |
| Desmocollin-1 | DSC1 | 0.54* | 0.35* | 0.53* | 0.79 | 1.12 | 0.55* | 0.56* |
| Guanine nucleotide-binding protein subunit beta-like protein 1 | GNB1L | 0.54* | 0.61* | 0.64* | 0.48* | 0.50* | 0.58 | 0.57 |
| Structural maintenance of chromosomes protein 2 | SMC2 | 0.55* | 0.44* | 0.54* | 0.42* | 0.65 | 0.74 | 0.48* |
| tRNA (uracil-5-)-methyltransferase homolog A | TRMT2A | 0.55* | 0.50* | 0.52* | 0.55* | 0.58 | 0.55* | 0.62 |
| Thrombomodulin | THBD | 0.55* | 0.62* | 0.60* | 1.70* | 7.47* | 6.68* | 7.22* |
| Serine/threonine-protein kinase Chk2 | CHEK2 | 0.56* | 0.63* | 0.61* | 0.38* | 0.31* | 0.25* | 0.25* |
| ATP-dependent RNA helicase DDX24 | DDX24 | 0.56* | 0.55* | 0.62* | 0.81 | 0.51* | 0.51* | 0.67* |
| U3 small nucleolar RNA-associated protein 6 homolog | UTP6 | 0.56* | 0.49* | 0.59* | 0.67 | 0.68 | 0.68 | 0.75 |
| Non-structural maintenance of chromosomes element 3 homolog | NSMCE3 | 0.56* | 0.62* | 0.61* | 0.60* | 0.87 | 0.88 | 0.88 |
| Vacuolar protein sorting-associated protein 18 homolog | VPS18 | 0.56* | 0.56* | 0.55* | 0.49* | 0.67* | 0.70 | 0.59* |
| Zinc finger CCHC domain-containing protein 8 | ZCCHC8 | 0.56* | 0.55* | 0.56* | 0.60* | 0.26* | 0.25* | 0.39* |
| Peroxisomal ATPase PEX1 | PEX1 | 0.56* | 0.64* | 0.64* | 0.39* | 0.43* | 0.48* | 0.31* |
| Ribosomal RNA-processing protein 7 homolog A | RRP7A | 0.57* | 0.49* | 0.52* | 0.47* | 0.39* | 0.41* | 0.46* |
| Zinc finger and BTB domain-containing protein 7A | ZBTB7A | 0.57* | 0.54* | 0.56* | 0.56* | 0.52* | 0.45* | 0.56* |
| Serine/threonine-protein kinase MRCK alpha | CDC42BPA | 0.58* | 0.63* | 0.57* | 0.52* | 0.64 | 0.58* | 0.40* |
| Probable ribosome biogenesis protein RLP24 | RSL24D1 | 0.58* | 0.47* | 0.54* | 0.50* | 0.22* | 0.21* | 0.27* |
| Condensin complex subunit 3 | NCAPG | 0.58* | 0.53* | 0.54* | 0.45* | 0.97 | 0.80 | 0.67 |
| Epithelial splicing regulatory protein 2 | ESRP2 | 0.58* | 0.58* | 0.66* | 0.42* | 0.50* | 0.38* | 0.45* |
| E3 ubiquitin-protein ligase makorin-2 | MKRN2 | 0.59* | 0.67 | 0.65* | 0.42* | 0.60 | 0.52* | 0.45* |

| | | | | | | | | |
|---|---|---|---|---|---|---|---|---|
| Dynamin-binding protein | DNMBP | 0.59* | 0.48* | 0.50* | 0.52* | 0.66 | 0.52 | 0.46* |
| Replication factor C subunit 2 | RFC2 | 0.59* | 0.62* | 0.63* | 0.34* | 0.52* | 0.54* | 0.44* |
| Transmembrane channel-like protein 6 | TMC6 | 0.59* | 0.47* | 0.51* | 0.62* | 0.63 | 0.64 | 0.61 |
| Tetratricopeptide repeat protein 7A | TTC7A | 0.60* | 0.57* | 0.61* | 0.46* | 0.66 | 0.77 | 0.73 |
| DNA mismatch repair protein Msh2 | MSH2 | 0.60* | 0.57* | 0.67* | 0.70 | 0.66 | 0.64 | 0.75 |
| S1 RNA-binding domain-containing protein 1 | SRBD1 | 0.60* | 0.53* | 0.64* | 0.43* | 0.58 | 0.40* | 0.23* |
| WD repeat-containing protein 36 | WDR36 | 0.60* | 0.52* | 0.56* | 0.76 | 0.69 | 0.62 | 0.71 |
| U3 small nucleolar RNA-associated protein 4 homolog | UTP4 | 0.61* | 0.58* | 0.61* | 0.80 | 1.19 | 1.21 | 1.14 |
| Phosphoinositide 3-kinase regulatory subunit 4 | PIK3R4 | 0.61* | 0.65* | 0.59* | 0.64* | 0.72 | 0.62 | 0.58 |
| 3'-5' RNA helicase YTHDC2 | YTHDC2 | 0.61* | 0.59* | 0.64* | 0.82 | 0.72 | 0.60* | 0.78 |
| Protein AATF | AATF | 0.61* | 0.52* | 0.65* | 0.82 | 0.54* | 0.54* | 0.65 |
| TRAF family member-associated NF-kappa-B activator | TANK | 0.61* | 0.64 | 0.66* | 0.91 | 0.50* | 0.50* | 0.55 |
| Transcriptional adapter 1 | TADA1 | 0.62* | 0.65* | 0.58* | 0.95 | 1.19 | 1.06 | 1.29 |
| E3 ubiquitin-protein ligase MIB1 | MIB1 | 0.62* | 0.64* | 0.55* | 0.69 | 0.71 | 0.56 | 0.72 |
| Gap junction beta-1 protein | GJB1 | 0.62* | 0.56* | 0.56* | 0.75 | 0.66 | 0.53* | 0.58* |
| Translation initiation factor eIF-2B subunit delta | EIF2B4 | 0.62* | 0.59* | 0.55* | 0.64* | 0.57* | 0.57* | 0.61* |
| DNA polymerase delta catalytic subunit | POLD1 | 0.62* | 0.55* | 0.53* | 0.73 | 0.61 | 0.62 | 0.85 |
| E3 ubiquitin-protein ligase BRE1B | RNF40 | 0.62* | 0.66* | 0.61* | 0.62* | 0.57* | 0.49* | 0.64 |
| ATPase WRNIP1 | WRNIP1 | 0.62* | 0.60* | 0.63* | 0.35* | 0.39* | 0.35* | 0.38* |
| A-kinase anchor protein 8 | AKAP8 | 0.63* | 0.53* | 0.59* | 0.50* | 0.42* | 0.47* | 0.67 |
| Tetratricopeptide repeat protein 12 | TTC12 | 0.64* | 0.64* | 0.64* | 0.57* | 0.54* | 0.51* | 0.44* |
| Syndecan-1 | SDC1 | 0.64* | 0.52* | 0.65* | 0.81 | 0.67 | 0.59 | 0.73 |
| Activity-dependent neuroprotector homeobox protein | ADNP | 0.65* | 0.65* | 0.67* | 0.68* | 0.77 | 0.72* | 0.86 |
| Probable dimethyladenosine transferase | DIMT1 | 0.65* | 0.59* | 0.64* | 0.70 | 0.59 | 0.75 | 0.90 |
| Tubulin alpha chain-like 3 | TUBAL3 | 0.66* | 0.67* | 0.56* | 0.39* | 0.33* | 0.45* | 0.17* |
| Nucleolar protein 6 | NOL6 | 0.67* | 0.57* | 0.61* | 0.81 | 0.75 | 0.64 | 0.74 |

***B. Down-regulated proteins unique to Aquamin (AQ) [243 proteins]***

| | | Interventions | | | | | | |
|---|---|---|---|---|---|---|---|---|
| | | Control | | | *With LPS & Cytokines* | | | |
| Proteins | Genes | **AQ** | AQ+MES | MES | *LPS-Cyto* | *AQ* | *AQ+MES* | *MES* |
| Reticulocalbin-3 | RCN3 | 0.17* | 0.72 | 0.78 | 1.01 | 3.31* | 4.57* | 5.68* |
| Carboxypeptidase B2 | CPB2 | 0.23* | 1.02 | 1.24 | 1.76* | 5.38* | 5.30* | 10.11* |
| Collectin-11 | COLEC11 | 0.28* | 0.79 | 1.14 | 1.30 | 2.59* | 3.59* | 5.27* |
| Mimecan | OGN | 0.29* | 0.85 | 0.94 | 1.34 | 3.21* | 4.58* | 5.97* |
| HLA class II histocompatibility antigen, DP beta 1 chain | HLA-DPB1 | 0.31* | 2.33* | 1.80* | 15.14* | 13.55* | 15.68* | 15.01* |
| Dihydropyrimidinase | DPYS | 0.33* | 0.71 | 0.75 | 1.13 | 2.82* | 3.76* | 4.94* |

| | | | | | | | | |
|---|---|---|---|---|---|---|---|---|
| Protein S100-A7 | S100A7 | 0.33* | 0.70 | 0.94 | 1.71* | 0.60 | 1.01 | 1.30 |
| Rho guanine nucleotide exchange factor 40 | ARHGEF40 | 0.35* | 0.74 | 1.05 | 0.31* | 0.49* | 0.88 | 0.97 |
| Fibrinogen gamma chain | FGG | 0.35* | 1.28 | 2.38* | 2.58* | 3.47* | 3.42* | 6.21* |
| Olfactomedin-like protein 3 | OLFML3 | 0.35* | 1.30 | 0.87 | 1.59* | 3.28* | 5.46* | 7.10* |
| Ankyrin repeat domain-containing protein SOWAHB | SOWAHB | 0.35* | 0.73 | 0.67 | 0.39* | 0.53 | 0.48* | 0.46* |
| Mucin-like protein 1 | MUCL1 | 0.37* | 0.77 | 0.72 | 1.23 | 0.90 | 0.72 | 1.47 |
| Vasorin | VASN | 0.39* | 0.96 | 0.86 | 1.44 | 2.45* | 3.55* | 5.06* |
| Zinc finger protein 536 | ZNF536 | 0.40* | 1.44 | 1.10 | 1.81* | 2.08* | 2.70* | 5.62* |
| ABC-type oligopeptide transporter ABCB9 | ABCB9 | 0.40* | 0.67 | 1.05 | 1.77* | 0.59 | 0.93 | 2.42* |
| Suprabasin | SBSN | 0.41* | 0.70* | 0.75* | 1.09 | 0.96 | 0.80 | 1.12 |
| Cerebellin-4 | CBLN4 | 0.41* | 1.58 | 2.52* | 0.57* | 0.78 | 1.73 | 1.95* |
| Coagulation factor X | F10 | 0.41* | 0.77 | 0.81 | 1.13 | 3.39* | 5.07* | 5.60* |
| RAB6A-GEF complex partner protein 2 | RGP1 | 0.41* | 0.67 | 0.93 | 0.65 | 0.92 | 1.04 | 0.97 |
| Coiled-coil domain-containing protein 39 | CCDC39 | 0.41* | 0.77 | 1.31 | 2.33* | 2.48* | 2.44* | 2.90* |
| Microtubule-associated protein 1B | MAP1B | 0.42* | 0.77 | 0.85 | 1.20 | 2.79* | 2.86* | 3.79* |
| Protein BCAP | ODF2L | 0.43* | 1.24 | 1.79* | 2.23* | 2.03* | 2.99* | 3.70* |
| Programmed cell death 1 ligand 1 | CD274 | 0.43* | 1.53 | 1.44 | 16.94* | 16.35* | 19.58* | 15.89* |
| Cdc42 effector protein 5 | CDC42EP5 | 0.44* | 0.80 | 0.94 | 1.05 | 0.88 | 0.85 | 0.47* |
| Ribosomal protein eL22-like | RPL22L1 | 0.45* | 0.89 | 1.44 | 0.86 | 0.54 | 1.05 | 1.90* |
| Coiled-coil domain-containing protein 91 | CCDC91 | 0.46* | 0.97 | 0.95 | 0.49* | 0.54* | 0.49* | 0.49* |
| Neural proliferation differentiation and control protein 1 | NPDC1 | 0.46* | 0.76 | 0.88 | 0.47* | 0.26* | 0.24* | 0.25* |
| Carbamoyl-phosphate synthase [ammonia], mitochondrial | CPS1 | 0.47* | 1.17 | 0.96 | 1.52* | 4.01* | 4.48* | 5.28* |
| Protocadherin-12 | PCDH12 | 0.47* | 1.35 | 1.06 | 1.79* | 2.87* | 4.80* | 6.92* |
| Kininogen-1 | KNG1 | 0.47* | 2.27* | 2.73* | 1.71* | 0.68 | 2.29* | 2.32* |
| Protein FAM117B | FAM117B | 0.47* | 0.87 | 0.70 | 0.60* | 0.12* | 0.28* | 0.71 |
| Ral GTPase-activating protein subunit alpha-2 | RALGAPA2 | 0.47* | 0.70 | 0.69* | 0.52* | 0.48* | 0.38* | 0.37* |
| Protein S100-A8 | S100A8 | 0.48* | 1.04 | 0.90 | 1.09 | 1.06 | 1.27 | 1.27 |
| Nucleus accumbens-associated protein 1 | NACC1 | 0.48* | 0.68* | 0.69* | 0.50* | 0.31* | 0.32* | 0.34* |
| Inter-alpha-trypsin inhibitor heavy chain H1 | ITIH1 | 0.48* | 0.96 | 1.54* | 0.48* | 0.48* | 1.13 | 1.53 |
| Integrin alpha-7 | ITGA7 | 0.48* | 0.78 | 0.74 | 5.21* | 0.84 | 0.49* | 1.76 |
| Fermitin family homolog 3 | FERMT3 | 0.48* | 1.02 | 0.99 | 1.11 | 2.94* | 4.31* | 4.74* |
| C4b-binding protein alpha chain | C4BPA | 0.48* | 1.00 | 1.41 | 1.50* | 1.06 | 1.25 | 1.68 |
| Rootletin | CROCC | 0.49* | 1.51* | 0.96 | 0.74 | 0.50* | 0.53* | 0.43* |
| Adenylyl cyclase-associated protein 2 | CAP2 | 0.49* | 1.23 | 1.16 | 1.51* | 3.15* | 3.71* | 4.96* |
| Lysine-specific demethylase 2A | KDM2A | 0.49* | 0.82 | 0.77 | 0.73 | 0.44* | 0.45* | 0.70 |
| Keratin, type II cytoskeletal 6A | KRT6A | 0.49* | 1.00 | 1.23 | 1.61* | 0.93 | 1.36 | 1.29 |
| Smad nuclear-interacting protein 1 | SNIP1 | 0.49* | 0.69 | 0.71 | 0.53* | 0.69 | 0.66 | 0.48* |

| | | | | | | | | |
|---|---|---|---|---|---|---|---|---|
| Protein Wnt-3a | WNT3A | 0.49* | 0.92 | 0.90 | 1.17 | 3.24* | 4.10* | 5.41* |
| Protein bassoon | BSN | 0.50* | 2.53* | 1.96* | 5.37* | 19.19* | 17.24* | 29.63* |
| Sex hormone-binding globulin | SHBG | 0.50* | 2.02* | 2.50* | 0.70 | 0.75 | 1.95* | 2.89* |
| Collagen alpha-1(IV) chain | COL4A1 | 0.50* | 1.17 | 1.70* | 1.82* | 2.23* | 2.22* | 3.55* |
| Beta-enolase | ENO3 | 0.50* | 0.75 | 0.76* | 1.95* | 5.84* | 4.93* | 4.93* |
| Apolipoprotein M | APOM | 0.51* | 0.86 | 1.24* | 1.56* | 2.35* | 3.87* | 5.61* |
| C-type lectin domain family 11 member A | CLEC11A | 0.51* | 1.23 | 1.22 | 1.20 | 2.68* | 3.66* | 4.12* |
| DNA-binding protein SATB2 | SATB2 | 0.51* | 0.92 | 0.75* | 0.48* | 0.45* | 0.42* | 0.63* |
| Protein unc-93 homolog A | UNC93A | 0.51* | 1.04 | 1.49 | 2.62* | 1.95* | 2.34* | 3.58* |
| Dynein axonemal heavy chain 1 | DNAH1 | 0.51* | 1.54* | 2.15* | 1.10 | 0.62* | 2.04* | 2.51* |
| Afamin | AFM | 0.52* | 2.16* | 2.43* | 0.71* | 0.67* | 2.00* | 2.67* |
| Myomegalin | PDE4DIP | 0.52* | 0.90 | 0.94 | 0.87 | 0.30* | 0.31* | 0.60* |
| Ferritin light chain | FTL | 0.52* | 2.72* | 3.90* | 1.95* | 0.58 | 4.12* | 9.36* |
| Claudin-1 | CLDN1 | 0.52* | 0.82 | 1.07 | 0.92 | 0.85 | 0.65 | 1.06 |
| Collagen alpha-1(XI) chain | COL11A1 | 0.53* | 2.32* | 2.75* | 0.87 | 0.94 | 2.20* | 3.10* |
| CDK-activating kinase assembly factor MAT1 | MNAT1 | 0.53* | 0.93 | 1.05 | 0.79 | 0.52 | 0.89 | 1.51 |
| Tensin-3 | TNS3 | 0.53* | 0.80 | 0.74* | 0.43* | 0.72 | 0.61 | 0.47* |
| Collagen alpha-2(I) chain | COL1A2 | 0.53* | 2.15* | 2.47* | 0.72 | 0.65 | 1.88* | 2.71* |
| Coiled-coil domain-containing protein 9B | CCDC9B | 0.53* | 0.96 | 1.03 | 0.55* | 0.41* | 0.73 | 0.79 |
| AN1-type zinc finger protein 6 | ZFAND6 | 0.53* | 0.75* | 0.85 | 0.35* | 0.06* | 0.11* | 0.22* |
| Probable phosphoglycerate mutase 4 | PGAM4 | 0.53* | 1.15 | 2.54* | 0.51* | 0.63 | 1.94* | 2.77* |
| Vitronectin | VTN | 0.53* | 1.33 | 1.51* | 1.58* | 1.55 | 1.93* | 2.76* |
| DEP domain-containing mTOR-interacting protein | DEPTOR | 0.53* | 0.74 | 0.75* | 0.26* | 0.46* | 0.39* | 0.27* |
| Ubiquitin-associated protein 2 | UBAP2 | 0.54* | 0.90 | 1.00 | 0.59* | 0.07* | 0.16* | 0.10* |
| Vascular cell adhesion protein 1 | VCAM1 | 0.54* | 1.13 | 1.26 | 2.04* | 3.97* | 5.56* | 6.73* |
| Protocadherin gamma-C3 | PCDHGC3 | 0.54* | 1.39 | 1.37* | 1.48* | 2.77* | 3.66* | 5.21* |
| FERM and PDZ domain-containing protein 1 | FRMPD1 | 0.54* | 2.61* | 2.58* | 0.47* | 0.94 | 1.85* | 2.22* |
| Laminin subunit beta-1 | LAMB1 | 0.54* | 2.04* | 1.65* | 1.58* | 2.39* | 2.83* | 3.78* |
| BAH and coiled-coil domain-containing protein 1 | BAHCC1 | 0.54* | 1.51* | 0.94 | 0.43* | 0.49* | 0.40* | 0.22* |
| Retinoic acid receptor responder protein 2 | RARRES2 | 0.54* | 1.61* | 2.51* | 0.66 | 0.68 | 1.46 | 2.08* |
| SPARC | SPARC | 0.54* | 1.40* | 1.85* | 1.86* | 1.88* | 3.08* | 3.36* |
| Alpha-fetoprotein | AFP | 0.54* | 1.16 | 1.75* | 1.41* | 1.41* | 2.14* | 3.27* |
| Adenosine 3'-phospho 5'-phosphosulfate transporter 2 | SLC35B3 | 0.55* | 0.96 | 0.84 | 0.96 | 1.05 | 0.94 | 1.05 |
| Rho GTPase-activating protein 42 | ARHGAP42 | 0.55* | 0.74 | 0.71 | 0.78 | 0.67 | 0.97 | 0.45* |
| Nuclear factor 1 C-type | NFIC | 0.55* | 0.74 | 0.78 | 1.06 | 1.84* | 1.42 | 1.30 |
| Nucleolar complex protein 3 homolog | NOC3L | 0.55* | 0.80 | 0.98 | 1.23 | 0.41* | 0.42* | 0.82 |
| Laminin subunit alpha-1 | LAMA1 | 0.55* | 2.01* | 1.66* | 1.60* | 2.30* | 2.88* | 3.96* |

| | | | | | | | | |
|---|---|---|---|---|---|---|---|---|
| Thymosin beta-4 | TMSB4X | 0.55* | 0.94 | 1.71* | 1.20 | 1.01 | 1.17 | 1.93* |
| Guanidinoacetate N-methyltransferase | GAMT | 0.55* | 1.98* | 1.89* | 0.67 | 0.57 | 1.54 | 1.94* |
| Thymosin beta-10 | TMSB10 | 0.55* | 0.69 | 1.12 | 1.08 | 0.57 | 0.67 | 0.93 |
| Alpha-1B-glycoprotein | A1BG | 0.55* | 1.99* | 2.73* | 0.77 | 0.76 | 2.05* | 3.14* |
| Gamma-taxilin | TXLNG | 0.56* | 0.82 | 0.80 | 0.73 | 0.81 | 0.70 | 0.86 |
| Myosin light chain 6B | MYL6B | 0.56* | 0.71* | 0.75 | 0.80 | 1.66* | 1.68* | 1.15 |
| Fibrinogen beta chain | FGB | 0.56* | 2.04* | 1.67* | 1.92* | 3.53* | 3.54* | 6.17* |
| ATPase family gene 2 protein homolog A | AFG2A | 0.56* | 0.72 | 0.72 | 0.67 | 0.78 | 0.76 | 0.77 |
| Carboxypeptidase N catalytic chain | CPN1 | 0.56* | 0.92 | 0.92 | 1.44* | 3.11* | 4.54* | 5.08* |
| Endoribonuclease ZC3H12A | ZC3H12A | 0.56* | 0.69* | 0.73* | 0.46* | 0.80 | 0.84 | 0.70 |
| Inter-alpha-trypsin inhibitor heavy chain H4 | ITIH4 | 0.56* | 1.42 | 1.59* | 2.09* | 1.62* | 2.06* | 3.18* |
| Non-histone chromosomal protein HMG-17 | HMGN2 | 0.56* | 1.04 | 1.61* | 0.87 | 0.80 | 1.21 | 1.52 |
| Phospholipid scramblase 1 | PLSCR1 | 0.56* | 1.04 | 1.09 | 1.60* | 0.94 | 1.55 | 1.76* |
| Protein piccolo | PCLO | 0.56* | 1.75* | 2.05* | 0.55* | 0.72 | 2.03* | 2.36* |
| Sphingomyelin phosphodiesterase 2 | SMPD2 | 0.56* | 1.01 | 1.40 | 0.73 | 0.70 | 1.09 | 0.90 |
| Tripartite motif-containing protein 26 | TRIM26 | 0.56* | 0.73* | 0.71* | 0.63* | 0.53* | 0.56* | 0.62* |
| Dynamin-1 | DNM1 | 0.57* | 1.02 | 0.79 | 1.22 | 2.61* | 1.81* | 2.14* |
| Pigment epithelium-derived factor | SERPINF1 | 0.57* | 1.24 | 1.90* | 1.24 | 0.96 | 2.19* | 3.38* |
| Pregnancy zone protein | PZP | 0.57* | 1.21 | 1.00 | 1.42 | 0.69 | 1.38 | 1.86 |
| Keratin, type I cytoskeletal 16 | KRT16 | 0.57* | 0.74 | 0.81 | 1.57* | 0.80 | 0.85 | 1.03 |
| Protein polybromo-1 | PBRM1 | 0.57* | 0.81 | 0.74* | 0.47* | 0.32* | 0.33* | 0.27* |
| Deoxynucleotidyltransferase terminal-interacting protein 2 | DNTTIP2 | 0.57* | 0.77 | 0.76 | 0.51* | 0.67 | 0.67 | 0.56* |
| Phosphoinositide 3-kinase adapter protein 1 | PIK3AP1 | 0.57* | 0.81 | 0.75 | 0.97 | 1.01 | 0.90 | 0.93 |
| Cartilage oligomeric matrix protein | COMP | 0.57* | 0.84 | 1.22 | 1.73* | 2.91* | 4.29* | 6.18* |
| Abl interactor 2 | ABI2 | 0.57* | 0.79 | 0.83 | 0.91 | 1.11 | 0.97 | 1.21 |
| Retinoblastoma-associated protein | RB1 | 0.57* | 0.73 | 0.82 | 0.71 | 1.02 | 1.15 | 0.84 |
| AMP deaminase 3 | AMPD3 | 0.58* | 0.83 | 0.81 | 0.65 | 0.64 | 0.73 | 0.77 |
| Cerebellar degeneration-related protein 2 | CDR2 | 0.58* | 0.93 | 0.86 | 0.81 | 1.26 | 0.91 | 1.09 |
| Cytosolic carboxypeptidase 1 | AGTPBP1 | 0.58* | 0.73 | 0.86 | 0.64* | 0.62 | 0.35* | 0.67 |
| Splicing regulator ARVCF | ARVCF | 0.58* | 0.91 | 0.98 | 0.40* | 0.46* | 0.37* | 0.29* |
| BRCA1-associated protein | BRAP | 0.58* | 0.73 | 0.74 | 0.59* | 0.33* | 0.42* | 0.44* |
| DNA-binding protein SMUBP-2 | IGHMBP2 | 0.58* | 1.53* | 1.72* | 2.47* | 3.42* | 3.38* | 5.07* |
| E3 ubiquitin-protein ligase TRIM15 | TRIM15 | 0.58* | 0.81 | 0.82 | 0.72 | 0.74 | 0.86 | 0.75 |
| Endoribonuclease Dicer | DICER1 | 0.58* | 0.75 | 0.84 | 0.53* | 0.94 | 0.86 | 0.85 |
| Inactive tyrosine-protein kinase 7 | PTK7 | 0.59* | 0.75 | 0.86 | 0.69 | 1.12 | 0.98 | 0.88 |
| Ribosomal protein eL42-like | RPL36AL | 0.59* | 0.84 | 0.84 | 0.70 | 0.61 | 0.72 | 0.65 |
| Plasma kallikrein | KLKB1 | 0.59* | 2.06* | 2.86* | 0.72 | 0.82 | 1.89* | 2.72* |

| | | | | | | | | |
|---|---|---|---|---|---|---|---|---|
| Gamma-tubulin complex component 2 | TUBGCP2 | 0.59* | 0.71* | 0.77* | 0.71* | 0.92 | 0.94 | 0.96 |
| Complement factor D | CFD | 0.59* | 1.43 | 3.72* | 0.81 | 0.58 | 1.54 | 2.05* |
| Extracellular sulfatase Sulf-2 | SULF2 | 0.59* | 0.93 | 1.25* | 0.86 | 0.35* | 0.37* | 0.73* |
| Replication factor C subunit 4 | RFC4 | 0.59* | 0.80 | 0.92 | 0.63* | 0.81 | 0.77 | 0.95 |
| Regucalcin | RGN | 0.59* | 1.19 | 1.30 | 1.08 | 1.30 | 1.94* | 2.66* |
| Calmodulin-regulated spectrin-associated protein 1 | CAMSAP1 | 0.59* | 0.92 | 1.06 | 0.77 | 0.86 | 0.61 | 0.70 |
| Poly(A)-specific ribonuclease PARN | PARN | 0.59* | 0.68* | 0.67* | 0.62* | 0.66* | 0.71 | 0.71 |
| Alpha-amylase 1B | AMY1B | 0.60* | 1.01 | 1.13 | 1.52* | 3.33* | 3.94* | 5.43* |
| Protein S100-A9 | S100A9 | 0.60* | 1.51* | 1.41* | 1.53* | 1.35 | 1.39 | 2.46* |
| Low-density lipoprotein receptor-related protein 2 | LRP2 | 0.60* | 1.46 | 1.02 | 1.45 | 4.10* | 5.03* | 6.63* |
| Elongator complex protein 4 | ELP4 | 0.60* | 0.73 | 0.75 | 0.66 | 0.60 | 0.53 | 0.49* |
| OTU domain-containing protein 3 | OTUD3 | 0.60* | 0.71 | 0.81 | 0.39* | 0.22* | 0.16* | 0.43* |
| Interferon-induced protein 44-like | IFI44L | 0.60* | 1.47 | 1.36 | 1.22 | 1.34 | 1.82 | 3.21* |
| NADPH oxidase organizer 1 | NOXO1 | 0.60* | 0.72 | 0.81 | 0.36* | 0.61 | 0.52 | 0.31* |
| Protein pelota homolog | PELO | 0.60* | 0.72* | 0.69* | 0.59* | 0.70 | 0.70 | 0.65* |
| Filaggrin | FLG | 0.60* | 0.72* | 0.83 | 1.47* | 0.97 | 0.88 | 1.06 |
| Complement component C8 alpha chain | C8A | 0.60* | 1.16 | 1.38* | 1.68* | 3.50* | 4.58* | 6.06* |
| Glucocorticoid modulatory element-binding protein 2 | GMEB2 | 0.60* | 0.73 | 0.76 | 0.54* | 0.68 | 0.74 | 0.57 |
| Sex-determining region Y protein | SRY | 0.61* | 1.19 | 1.25 | 1.77* | 1.72 | 1.71 | 2.48* |
| Coiled-coil domain-containing protein 85C | CCDC85C | 0.61* | 0.72* | 0.70* | 0.39* | 0.15* | 0.22* | 0.24* |
| Immediate early response 3-interacting protein 1 | IER3IP1 | 0.61* | 1.26 | 1.09 | 1.05 | 1.64* | 1.16 | 0.89 |
| von Willebrand factor A domain-containing protein 1 | VWA1 | 0.61* | 1.07 | 1.14 | 1.04 | 2.29* | 2.51* | 3.15* |
| E3 ubiquitin-protein ligase TRIM68 | TRIM68 | 0.61* | 0.90 | 0.91 | 0.76 | 1.24 | 1.19 | 0.78 |
| Laminin subunit gamma-1 | LAMC1 | 0.61* | 1.97* | 1.62* | 1.54* | 2.28* | 2.68* | 3.65* |
| Synaptojanin-2 | SYNJ2 | 0.61* | 0.68* | 0.76 | 0.49* | 0.41* | 0.32* | 0.33* |
| ADP-ribosylation factor-binding protein GGA2 | GGA2 | 0.61* | 0.80 | 0.81 | 0.40* | 0.69 | 0.44* | 0.48* |
| Interleukin-1 receptor accessory protein | IL1RAP | 0.61* | 2.35* | 2.99* | 0.79 | 0.92 | 2.31* | 3.58* |
| Coagulation factor V | F5 | 0.61* | 1.22 | 1.26 | 1.74* | 1.61 | 1.89* | 2.43* |
| Metallothionein-1H | MT1H | 0.61* | 0.90 | 0.77 | 0.97 | 0.42* | 0.55 | 0.62 |
| Probable helicase with zinc finger domain | HELZ | 0.61* | 0.70 | 0.67* | 0.49* | 1.11 | 1.01 | 1.01 |
| Regulatory-associated protein of mTOR | RPTOR | 0.61* | 0.70 | 0.73* | 0.69 | 0.90 | 0.81 | 0.91 |
| Histone-lysine N-methyltransferase SETD1A | SETD1A | 0.61* | 0.70 | 0.79 | 0.76 | 0.60 | 0.55 | 0.68 |
| Helicase with zinc finger domain 2 | HELZ2 | 0.61* | 0.88 | 0.77 | 0.53* | 0.36* | 0.97 | 0.57 |
| Alpha-2-macroglobulin | A2M | 0.61* | 1.48* | 1.87* | 1.88* | 1.77* | 2.51* | 3.48* |
| Protein TANC1 | TANC1 | 0.62* | 0.71 | 0.76 | 0.69 | 0.87 | 0.59 | 0.63 |
| UPF0538 protein C2orf76 | C2orf76 | 0.62* | 0.76 | 0.83 | 0.77 | 0.71 | 0.58 | 0.73 |
| RalBP1-associated Eps domain-containing protein 2 | REPS2 | 0.62* | 0.72 | 0.76 | 0.43* | 0.43* | 0.24* | 0.25* |

| | | | | | | | | |
|---|---|---|---|---|---|---|---|---|
| Kinesin-like protein KIF13A | KIF13A | 0.62* | 0.77 | 0.74* | 0.69 | 0.66 | 0.63 | 0.66 |
| Vacuolar protein sorting-associated protein 72 homolog | VPS72 | 0.62* | 0.73 | 0.82 | 0.28* | 0.92 | 0.17* | 0.70 |
| EGF-containing fibulin-like extracellular matrix protein 1 | EFEMP1 | 0.62* | 2.26* | 2.51* | 0.81 | 0.53* | 1.88* | 2.37* |
| Insulin-like growth factor II | IGF2 | 0.62* | 2.07* | 2.04* | 0.74 | 0.48* | 1.65* | 2.22* |
| CCHC-type zinc finger nucleic acid binding protein | CNBP | 0.62* | 0.99 | 0.95 | 1.03 | 0.55* | 0.93 | 1.04 |
| Cyclin-dependent kinase 1 | CDK1 | 0.62* | 0.67* | 0.68* | 0.40* | 1.51 | 1.34 | 1.54* |
| Collectin-10 | COLEC10 | 0.62* | 1.16 | 1.14 | 1.23 | 2.63* | 3.78* | 4.85* |
| Complement factor H | CFH | 0.62* | 2.03* | 27.05* | 0.74 | 0.62 | 1.68* | 1.72* |
| Low-density lipoprotein receptor-related protein 5 | LRP5 | 0.62* | 1.06 | 0.81 | 0.61* | 0.55 | 0.72 | 0.76 |
| WD repeat-containing protein 26 | WDR26 | 0.62* | 0.73 | 0.89 | 1.06 | 1.12 | 1.18 | 1.38 |
| Plasminogen | PLG | 0.62 | 1.31 | 1.39 | 1.44 | 1.62 | 1.48 | 1.84 |
| Hepatocyte growth factor activator | HGFAC | 0.62* | 2.13* | 2.48* | 0.77* | 0.77 | 2.27* | 3.01* |
| CCR4-NOT transcription complex subunit 6 | CNOT6 | 0.63* | 0.77 | 0.73 | 1.15 | 2.38* | 1.85* | 2.10* |
| UDP-N-acetylglucosamine transferase subunit ALG14 homolog | ALG14 | 0.63 | 0.81 | 1.10 | 1.15 | 1.16 | 0.88 | 0.64 |
| Spectrin beta chain, non-erythrocytic 2 | SPTBN2 | 0.63* | 0.94 | 1.28 | 0.86 | 0.84 | 1.25 | 1.43 |
| Ceruloplasmin | CP | 0.63* | 1.04 | 1.10 | 1.39* | 3.14* | 3.98* | 4.43* |
| Immunoglobulin kappa variable 4-1 | IGKV4-1 | 0.63* | 2.29* | 37.17* | 0.69 | 0.85 | 1.24 | 0.95 |
| Serum amyloid A-4 protein | SAA4 | 0.63* | 2.09* | 2.47* | 3.32* | 9.01* | 8.67* | 16.73* |
| Vinexin | SORBS3 | 0.63* | 1.02 | 0.97 | 0.84 | 0.76 | 0.96 | 1.01 |
| Amphiregulin | AREG | 0.63 | 1.46 | 1.11 | 1.32 | 1.51 | 1.25 | 3.02* |
| Mitochondrial ribosome-associated GTPase 2 | MTG2 | 0.63* | 0.83 | 0.94 | 0.26* | 0.29* | 0.28* | 0.25* |
| Albumin | ALB | 0.63* | 1.73* | 2.02* | 2.42* | 1.96* | 2.70* | 2.09* |
| Ubiquitin carboxyl-terminal hydrolase 3 | USP3 | 0.63* | 0.71 | 0.70 | 0.52* | 0.67 | 0.41* | 0.28* |
| Proto-oncogene c-Rel | REL | 0.63* | 0.88 | 0.79 | 1.17 | 1.50 | 0.99 | 1.70* |
| Phosphatidylinositol 3,4,5-trisphosphate 5-phosphatase 2 | INPPL1 | 0.63* | 0.89 | 0.77 | 0.77 | 0.97 | 0.98 | 1.27 |
| RNA polymerase II subunit A C-terminal domain phosphatase | CTDP1 | 0.63* | 0.67* | 0.70* | 0.62* | 0.63 | 0.55* | 0.62 |
| Fibrinogen alpha chain | FGA | 0.63* | 0.68 | 2.55* | 2.12* | 0.76 | 1.05 | 2.43* |
| Immunoglobulin heavy constant gamma 4 | IGHG4 | 0.64* | 1.95* | 32.94* | 0.66* | 0.72 | 1.67* | 0.96 |
| E3 ubiquitin-protein ligase RING2 | RNF2 | 0.64* | 0.81 | 0.86 | 0.85 | 0.15* | 0.24* | 0.38* |
| Metastasis-associated protein MTA3 | MTA3 | 0.64* | 0.69 | 0.82 | 0.71 | 0.94 | 0.80 | 1.02 |
| Tetraspanin-9 | TSPAN9 | 0.64 | 0.93 | 0.79 | 2.49* | 7.29* | 7.91* | 7.67* |
| Collagen alpha-2(VI) chain | COL6A2 | 0.64* | 1.24 | 1.33 | 1.16 | 1.60 | 2.85* | 3.58* |
| Scrapie-responsive protein 1 | SCRG1 | 0.64* | 2.21* | 2.25* | 0.80 | 0.99 | 1.82* | 2.12* |
| tRNA (guanine(26)-N(2))-dimethyltransferase | TRMT1 | 0.64* | 0.82 | 0.82 | 0.50* | 0.50* | 0.39* | 0.72 |
| Transcriptional repressor p66-alpha | GATAD2A | 0.64* | 0.72 | 0.99 | 0.78 | 0.50* | 0.69 | 0.85 |

| | | | | | | | | |
|---|---|---|---|---|---|---|---|---|
| DNA replication licensing factor MCM3 | MCM3 | 0.64* | 0.71 | 0.84 | 1.19 | 0.90 | 1.07 | 1.30 |
| Fibromodulin | FMOD | 0.64* | 2.45* | 2.75* | 1.06 | 0.95 | 2.23* | 3.19* |
| Ral GTPase-activating protein subunit alpha-1 | RALGAPA1 | 0.64* | 0.71 | 0.75 | 0.76 | 0.62 | 0.55 | 0.61 |
| Chromogranin-A | CHGA | 0.64* | 5.56* | 5.04* | 0.86 | 1.34 | 4.03* | 6.57* |
| Histone H3-7 | H3-7 | 0.64* | 0.78 | 0.93 | 0.99 | 1.01 | 1.06 | 1.31 |
| Dermcidin | DCD | 0.64* | 0.68* | 0.85 | 0.99 | 1.23 | 0.95 | 1.65* |
| Spectrin beta chain, erythrocytic | SPTB | 0.65 | 0.70 | 1.24 | 1.53 | 3.31* | 3.32* | 4.60* |
| Single-stranded DNA-binding protein 4 | SSBP4 | 0.65* | 0.96 | 0.80 | 0.58* | 1.08 | 1.15 | 1.07 |
| Overexpressed in colon carcinoma 1 protein | OCC1 | 0.65* | 0.70 | 0.87 | 0.63* | 0.55* | 0.62 | 0.87 |
| Contactin-1 | CNTN1 | 0.65* | 2.25* | 2.79* | 0.80 | 0.88 | 2.29* | 3.42* |
| TBC1 domain family member 8B | TBC1D8B | 0.65* | 0.82 | 0.74* | 0.61* | 0.63 | 0.67 | 0.63 |
| Protein SDA1 homolog | SDAD1 | 0.65* | 0.70 | 0.69* | 0.69 | 0.35* | 0.42* | 0.47* |
| Beta-2-syntrophin | SNTB2 | 0.65* | 0.94 | 0.98 | 0.71 | 1.15 | 1.29 | 1.33 |
| Deoxynucleoside triphosphate triphosphohydrolase SAMHD1 | SAMHD1 | 0.65 | 1.18 | 1.07 | 1.16 | 2.07* | 1.70 | 1.86 |
| Pleckstrin homology-like domain family B member 2 | PHLDB2 | 0.65* | 0.72 | 0.85 | 0.71 | 0.85 | 0.74 | 0.70 |
| Hemopexin | HPX | 0.65* | 1.28 | 1.58* | 1.68* | 1.54 | 1.99* | 3.50* |
| Synergin gamma | SYNRG | 0.65* | 0.69 | 0.84 | 0.56* | 0.42* | 0.37* | 0.55 |
| BMP-binding endothelial regulator protein | BMPER | 0.65 | 1.20 | 1.13 | 1.57* | 3.53* | 3.94* | 4.84* |
| Cell adhesion molecule 1 | CADM1 | 0.65* | 1.89* | 2.04* | 0.63 | 0.89 | 1.76* | 2.25* |
| Coiled-coil domain-containing protein 50 | CCDC50 | 0.65* | 0.81 | 0.99 | 1.02 | 0.62 | 0.67 | 1.29 |
| Zinc finger CCCH domain-containing protein 15 | ZC3H15 | 0.65* | 0.74* | 0.80* | 0.74* | 0.67* | 0.67 | 0.80 |
| Homologous recombination OB-fold protein | HROB | 0.65* | 1.35* | 1.48* | 1.73* | 1.51* | 2.16* | 1.96* |
| Pro-glucagon | GCG | 0.65* | 1.31* | 1.64* | 0.78 | 0.99 | 0.72 | 0.86 |
| Phosphatidylcholine-sterol acyltransferase | LCAT | 0.65* | 2.10* | 2.67* | 0.76 | 0.94 | 2.06* | 2.94* |
| TBC domain-containing protein kinase-like protein | TBCK | 0.65* | 0.69 | 0.71 | 0.71 | 0.87 | 0.74 | 0.79 |
| Gap junction beta-2 protein | GJB2 | 0.66 | 0.71 | 0.82 | 0.91 | 1.23 | 1.20 | 1.26 |
| DNA mismatch repair protein Msh3 | MSH3 | 0.66* | 0.67* | 0.70* | 0.60* | 0.66 | 0.65 | 0.69 |
| CD5 antigen-like | CD5L | 0.66 | 1.53 | 23.17* | 0.77 | 0.69 | 0.82 | 0.29* |
| Serine/arginine repetitive matrix protein 2 | SRRM2 | 0.66* | 0.89 | 1.08 | 0.93 | 0.71 | 0.87 | 1.25 |
| Midasin | MDN1 | 0.66* | 1.29 | 1.56* | 1.48* | 1.53* | 2.03* | 2.59* |
| BRCA2 and CDKN1A-interacting protein | BCCIP | 0.66* | 0.96 | 0.88 | 1.09 | 0.82 | 1.09 | 1.09 |
| Transcription factor 20 | TCF20 | 0.66* | 0.99 | 1.00 | 2.22* | 0.80 | 0.51 | 1.59 |
| Guanine nucleotide-binding protein-like 3 | GNL3 | 0.66* | 0.68 | 0.79 | 0.55* | 0.56 | 0.46* | 0.47* |
| MAP kinase-activated protein kinase 5 | MAPKAPK5 | 0.66* | 0.99 | 0.88 | 0.49* | 0.63 | 0.44* | 0.45* |
| LIM and cysteine-rich domains protein 1 | LMCD1 | 0.66 | 0.81 | 0.92 | 1.00 | 0.74 | 0.76 | 0.91 |
| Immunoglobulin lambda constant 2 | IGLC2 | 0.66* | 1.33 | 4.95* | 1.42 | 3.25* | 2.14* | 1.55 |

| | | | | | | | | |
|---|---|---|---|---|---|---|---|---|
| E3 ubiquitin-protein ligase SH3RF1 | SH3RF1 | 0.66 | 0.85 | 0.79 | 0.70 | 0.69 | 0.55 | 0.45* |
| Actin filament-associated protein 1 | AFAP1 | 0.66 | 1.00 | 0.86 | 0.61* | 0.41* | 0.40* | 0.55 |
| Immortalization up-regulated protein | IMUP | 0.66* | 1.26 | 1.81* | 1.60* | 1.16 | 1.10 | 1.75* |
| Mixed lineage kinase domain-like protein | MLKL | 0.66 | 1.00 | 0.98 | 1.10 | 0.76 | 1.01 | 1.45 |
| Calmodulin-like protein 5 | CALML5 | 0.66 | 0.73 | 1.08 | 1.49 | 0.89 | 0.91 | 1.44 |
| Periostin | POSTN | 0.66* | 1.63* | 1.51* | 1.02 | 2.43* | 2.54* | 3.38* |
| Dual specificity tyrosine-phosphorylation-regulated kinase 1A | DYRK1A | 0.66* | 0.71* | 0.76* | 0.82 | 1.55* | 1.35 | 1.36 |
| Protein MAK16 homolog | MAK16 | 0.66* | 0.74 | 0.78 | 0.81 | 1.18 | 1.12 | 1.31 |
| EF-hand domain-containing protein D1 | EFHD1 | 0.66 | 43.85* | 1.91* | 0.69 | 0.69 | 0.84 | 3.11* |
| Keratin, type II cuticular Hb5 | KRT85 | 0.66* | 8.93* | 1.06 | 0.69* | 0.55* | 0.40* | 1.22 |
| Histone H2B type 1-B | H2BC3 | 0.66* | 0.83 | 0.98 | 1.05 | 0.58 | 0.63 | 0.97 |
| Inter-alpha-trypsin inhibitor heavy chain H3 | ITIH3 | 0.66* | 1.15 | 1.11 | 1.68* | 3.30* | 4.88* | 6.20* |
| Diacylglycerol kinase theta | DGKQ | 0.66 | 1.19 | 0.97 | 0.63* | 0.46* | 0.64 | 0.53* |
| Thrombospondin-4 | THBS4 | 0.67* | 1.01 | 1.08 | 1.82* | 3.14* | 4.76* | 6.66* |
| Desmoglein-1 | DSG1 | 0.67 | 0.82 | 0.96 | 1.11 | 1.13 | 0.69 | 1.18 |
| Muskelin | MKLN1 | 0.67* | 0.77 | 0.73 | 0.79 | 1.11 | 1.17 | 1.24 |
| Steroid hormone receptor ERR1 | ESRRA | 0.67* | 0.77 | 0.76 | 0.66 | 0.67 | 0.61 | 0.61 |
| Retinol-binding protein 4 | RBP4 | 0.67* | 2.60* | 2.77* | 0.92 | 0.90 | 2.14* | 2.97* |

***C. Down-regulated proteins unique to Aquamin plus Mesalamine (AQ+MES) [65 proteins]***

| | | Interventions | | | | | | |
|---|---|---|---|---|---|---|---|---|
| | | Control | | | *With LPS & Cytokines* | | | |
| Proteins | Genes | AQ | **AQ+MES** | MES | *LPS-Cyto* | *AQ* | *AQ+MES* | *MES* |
| HEAT repeat-containing protein 3 | HEATR3 | 0.74 | 0.44* | 0.85 | 0.72 | 0.52 | 0.37* | 0.80 |
| Leucine-rich alpha-2-glycoprotein | LRG1 | 1.03 | 0.46* | 0.83 | 0.52* | 0.97 | 0.66 | 0.54* |
| Mitogen-activated protein kinase kinase kinase kinase 1 | MAP4K1 | 0.76* | 0.46* | 1.00 | 0.70 | 0.76 | 0.67 | 0.69 |
| POTE ankyrin domain family member E | POTEE | 1.02 | 0.49* | 0.95 | 0.59* | 0.50* | 0.51* | 0.90 |
| Phosphatidate phosphatase LPIN3 | LPIN3 | 0.77 | 0.49* | 0.79 | 0.70 | 0.65 | 0.66 | 1.02 |
| Protein PALS2 | PALS2 | 0.98 | 0.52* | 0.71 | 0.52* | 1.81* | 1.54 | 1.29 |
| Dysferlin | DYSF | 0.78* | 0.53* | 0.82 | 0.51* | 0.36* | 0.42* | 0.35* |
| Keratin, type I cytoskeletal 13 | KRT13 | 0.75* | 0.53* | 0.92 | 0.69* | 0.87 | 0.64 | 1.46* |
| High mobility group protein HMGI-C | HMGA2 | 0.68 | 0.54* | 0.81 | 0.74 | 0.54 | 0.64 | 0.71 |
| Junctophilin-1 | JPH1 | 1.14 | 0.54* | 1.41 | 1.04 | 0.48* | 0.49* | 1.10 |
| Monocyte differentiation antigen CD14 | CD14 | 1.13 | 0.54* | 1.02 | 0.95 | 1.09 | 0.63 | 0.72 |
| Ribosomal protein uL30-like | RPL7L1 | 0.71* | 0.56* | 0.76 | 1.00 | 1.14 | 0.94 | 0.98 |

| | | | | | | | | |
|---|---|---|---|---|---|---|---|---|
| Nuclear ubiquitous casein and cyclin-dependent kinase substrate 1 | NUCKS1 | 0.78 | 0.56* | 0.78 | 0.82 | 0.70 | 0.71 | 0.91 |
| Polyamine deacetylase HDAC10 | HDAC10 | 1.11 | 0.56* | 0.92 | 0.67 | 1.07 | 0.85 | 0.88 |
| Histone PARylation factor 1 | HPF1 | 1.26 | 0.56* | 0.96 | 0.94 | 1.43 | 1.17 | 1.03 |
| Regenerating islet-derived protein 4 | REG4 | 0.73* | 0.56* | 0.98 | 0.66* | 0.45* | 0.49* | 0.52* |
| Methyl-CpG-binding domain protein 2 | MBD2 | 0.74 | 0.56* | 0.76 | 0.69 | 0.63 | 0.60 | 0.68 |
| General transcription factor IIF subunit 1 | GTF2F1 | 0.78 | 0.57* | 0.97 | 1.27 | 0.92 | 0.88 | 1.24 |
| Exportin-6 | XPO6 | 0.67 | 0.58* | 0.69 | 0.91 | 0.66 | 0.82 | 0.82 |
| WD repeat-containing protein 74 | WDR74 | 0.74 | 0.58* | 0.71 | 0.94 | 1.54 | 1.63 | 1.63 |
| Thioredoxin-related transmembrane protein 4 | TMX4 | 0.79 | 0.58* | 0.73 | 0.70 | 0.65 | 0.46* | 0.43* |
| DNA replication licensing factor MCM2 | MCM2 | 0.73 | 0.58* | 0.88 | 0.81 | 0.87 | 1.01 | 1.49 |
| Nik-related protein kinase | NRK | 0.87 | 0.59* | 0.80 | 1.03 | 0.57 | 0.92 | 1.26 |
| Breakpoint cluster region protein | BCR | 0.83 | 0.60* | 0.73 | 0.72 | 0.82 | 0.81 | 0.66 |
| WD repeat-containing protein 55 | WDR55 | 0.88 | 0.60* | 0.71 | 0.76 | 0.50* | 0.65 | 0.79 |
| Non-histone chromosomal protein HMG-14 | HMGN1 | 1.16 | 0.60* | 0.78 | 3.08* | 1.07 | 1.10 | 2.25* |
| Cystathionine beta-synthase | CBS | 1.00 | 0.60* | 0.69* | 0.83 | 0.89 | 0.84 | 0.84 |
| Integrator complex subunit 1 | INTS1 | 0.96 | 0.61 | 0.78 | 1.00 | 1.30 | 1.20 | 1.14 |
| Nuclear prelamin A recognition factor | NARF | 0.85 | 0.61* | 0.79 | 0.81 | 0.66 | 0.61 | 0.60 |
| WW domain-containing transcription regulator protein 1 | WWTR1 | 0.98 | 0.61 | 1.09 | 0.85 | 0.74 | 1.17 | 0.81 |
| Arf-GAP domain and FG repeat-containing protein 2 | AGFG2 | 0.81 | 0.62* | 0.76 | 0.61* | 0.61 | 0.45* | 0.54 |
| Myelin basic protein | MBP | 1.02 | 0.63 | 1.44 | 1.00 | 0.75 | 0.72 | 0.88 |
| Glucose-induced degradation protein 8 homolog | GID8 | 1.00 | 0.63* | 1.00 | 1.03 | 1.02 | 1.07 | 1.26 |
| E3 ubiquitin-protein ligase UHRF2 | UHRF2 | 0.79 | 0.63 | 1.05 | 1.05 | 0.61 | 0.74 | 0.85 |
| Probable ATP-dependent RNA helicase DHX37 | DHX37 | 0.71 | 0.63 | 0.72 | 0.77 | 0.91 | 1.05 | 0.96 |
| Sarcoplasmic/endoplasmic reticulum calcium ATPase 3 | ATP2A3 | 1.17 | 0.63* | 0.89 | 0.69 | 0.72 | 0.68 | 0.62 |
| Histone deacetylase complex subunit SAP30 | SAP30 | 0.72 | 0.63* | 0.71 | 0.57* | 0.95 | 0.62 | 0.72 |
| Kinesin-like protein KIF6 | KIF6 | 1.14 | 0.64 | 0.99 | 1.63* | 1.91* | 2.10* | 1.79 |
| Rab-like protein 2A | RABL2A | 0.67* | 0.64* | 0.77 | 0.32* | 0.45* | 0.43* | 0.28* |
| GTP-binding protein 4 | GTPBP4 | 0.72* | 0.64* | 0.86 | 0.85 | 0.76 | 0.74 | 1.03 |
| Kinesin-like protein KIF1B | KIF1B | 0.75 | 0.64* | 0.78 | 0.67 | 0.60 | 0.64 | 0.56 |
| Multivesicular body subunit 12A | MVB12A | 0.94 | 0.64* | 0.98 | 0.89 | 0.49* | 0.65 | 0.61 |
| Acid sphingomyelinase-like phosphodiesterase 3b | SMPDL3B | 0.89 | 0.64* | 0.68* | 0.86 | 1.05 | 0.69* | 0.68* |
| Kinesin-like protein KIF23 | KIF23 | 0.95 | 0.64* | 0.70* | 0.85 | 0.70* | 0.59* | 0.66* |
| Ras-related protein Rab-13 | RAB13 | 1.05 | 0.65* | 0.98 | 0.48* | 0.46* | 0.39* | 0.43* |
| Ribonucleases P/MRP protein subunit POP1 | POP1 | 0.83 | 0.65* | 0.73 | 0.99 | 0.71 | 0.80 | 1.04 |
| Microtubule-associated protein RP/EB family member 2 | MAPRE2 | 0.71 | 0.65* | 0.80 | 0.83 | 0.63 | 0.66 | 0.75 |
| Ribosome biogenesis protein BMS1 homolog | BMS1 | 0.71 | 0.65 | 0.98 | 0.73 | 0.51* | 0.57 | 0.79 |

| | | | | | | | | |
|---|---|---|---|---|---|---|---|---|
| DCC-interacting protein 13-alpha | APPL1 | 0.87 | 0.65* | 0.89 | 0.72* | 0.67 | 0.48* | 0.61* |
| Caspase-5 | CASP5 | 1.06 | 0.65* | 0.95 | 1.37 | 0.19* | 0.37* | 0.45* |
| PEST proteolytic signal-containing nuclear protein | PCNP | 0.88 | 0.65* | 0.93 | 0.78 | 0.53* | 0.64 | 0.76 |
| Prothymosin alpha | PTMA | 0.77 | 0.65* | 1.07 | 0.59* | 0.45* | 0.66 | 1.38* |
| Serine/threonine-protein kinase A-Raf | ARAF | 0.87 | 0.65* | 0.81 | 0.60* | 0.66 | 0.73 | 0.87 |
| E3 ubiquitin-protein ligase TRIM22 | TRIM22 | 0.69 | 0.65 | 0.73 | 0.93 | 0.97 | 0.99 | 0.93 |
| Ceramide kinase | CERK | 1.07 | 0.65 | 0.83 | 0.99 | 0.92 | 0.79 | 0.69 |
| Rac GTPase-activating protein 1 | RACGAP1 | 0.98 | 0.66* | 0.69* | 0.80 | 0.64* | 0.52* | 0.57* |
| IQCJ-SCHIP1 readthrough transcript protein | IQCJ-SCHIP1 | 0.72 | 0.66 | 1.10 | 0.93 | 0.89 | 0.80 | 1.02 |
| E3 ubiquitin-protein ligase NEDD4 | NEDD4 | 0.78 | 0.66* | 0.85 | 0.76 | 1.18 | 1.35 | 0.97 |
| Uncharacterized protein C11orf98 | C11orf98 | 1.07 | 0.66* | 0.84 | 0.91 | 0.21* | 0.21* | 0.25* |
| ATP-dependent RNA helicase DDX3Y | DDX3Y | 0.68 | 0.66 | 0.87 | 0.61* | 0.51* | 0.52* | 0.55* |
| Protein mono-ADP-ribosyltransferase PARP12 | PARP12 | 0.90 | 0.66 | 1.04 | 1.49 | 1.32 | 1.35 | 1.43 |
| Tyrosine-protein kinase BAZ1B | BAZ1B | 0.67* | 0.66* | 0.69* | 0.85 | 0.68 | 0.63 | 0.72 |
| Glutamate-rich WD repeat-containing protein 1 | GRWD1 | 0.78 | 0.67 | 0.82 | 0.95 | 0.75 | 0.73 | 0.90 |
| Keratin, type II cytoskeletal 78 | KRT78 | 0.71* | 0.67* | 1.26* | 1.50* | 1.92* | 0.51* | 1.49* |
| Translation factor GUF1, mitochondrial | GUF1 | 0.81 | 0.67* | 0.68* | 0.67* | 0.68 | 0.81 | 0.86 |

***D. Down-regulated proteins unique to Mesalamine (MES) [52 proteins]***

| | | Interventions | | | | | | |
|---|---|---|---|---|---|---|---|---|
| | | Control | | | *With LPS & Cytokines* | | | |
| Proteins | Genes | AQ | AQ+MES | **MES** | *LPS-Cyto* | *AQ* | *AQ+MES* | *MES* |
| Zinc transporter ZIP5 | SLC39A5 | 1.47* | 0.68 | 0.34* | 1.24 | 1.57 | 0.61 | 0.44* |
| Transmembrane 4 L6 family member 5 | TM4SF5 | 1.24 | 1.38 | 0.43* | 0.94 | 0.47* | 0.88 | 0.52 |
| Intelectin-2 | ITLN2 | 1.04 | 1.01 | 0.45* | 0.56* | 1.13 | 0.38* | 0.42* |
| Ectopic P granules protein 5 homolog | EPG5 | 1.07 | 0.91 | 0.47* | 0.87 | 0.85 | 0.69 | 0.75 |
| Growth arrest-specific protein 6 | GAS6 | 1.31* | 0.74 | 0.52* | 1.10 | 1.23 | 0.47* | 0.54* |
| Tuberin | TSC2 | 1.04 | 1.00 | 0.53* | 1.07 | 1.27 | 1.06 | 1.00 |
| AT-rich interactive domain-containing protein 4A | ARID4A | 0.88 | 0.73 | 0.53* | 1.09 | 0.88 | 0.72 | 0.87 |
| UDP-glucuronosyltransferase 2B15 | UGT2B15 | 0.87 | 0.69* | 0.53* | 0.73 | 0.92 | 0.62 | 0.61* |
| Protein Tob2 | TOB2 | 0.73 | 0.84 | 0.54* | 0.66 | 0.35* | 0.37* | 0.95 |
| Protein angel homolog 2 | ANGEL2 | 0.98 | 0.89 | 0.54* | 0.99 | 0.91 | 0.95 | 0.63 |
| Dimethylaniline monooxygenase [N-oxide-forming] 4 | FMO4 | 1.07 | 0.86 | 0.56* | 1.06 | 0.72 | 0.70 | 0.48* |
| Myelin regulatory factor-like protein | MYRFL | 0.96 | 0.80 | 0.57* | 1.19 | 0.89 | 0.77 | 0.61 |
| Serine incorporator 2 | SERINC2 | 0.90 | 0.86 | 0.58* | 0.50* | 0.89 | 0.86 | 0.55* |
| WD repeat domain phosphoinositide-interacting protein 4 | WDR45 | 0.99 | 0.70 | 0.58* | 0.84 | 0.62 | 0.94 | 0.79 |
| Zinc finger FYVE domain-containing protein 1 | ZFYVE1 | 1.06 | 0.74 | 0.59* | 0.76 | 0.82 | 0.87 | 0.67 |

| | | | | | | | | |
|---|---|---|---|---|---|---|---|---|
| Ly6/PLAUR domain-containing protein 8 | LYPD8 | 1.39 | 0.75 | 0.60* | 0.39* | 0.50* | 0.41* | 0.34* |
| Kallikrein-11 | KLK11 | 1.02 | 0.71 | 0.60* | 1.03 | 1.14 | 0.69 | 0.68 |
| ADP-ribosylation factor 3 | ARF3 | 0.75 | 1.05 | 0.60* | 1.25 | 1.12 | 1.03 | 1.12 |
| Deubiquitinating protein VCPIP1 | VCPIP1 | 0.74 | 0.83 | 0.60* | 0.92 | 0.99 | 0.96 | 1.02 |
| Cytochrome P450 2C19 | CYP2C19 | 2.12* | 1.15 | 0.61* | 1.64* | 1.98* | 0.88 | 0.62 |
| Transmembrane 7 superfamily member 3 | TM7SF3 | 0.93 | 0.97 | 0.61* | 1.41* | 1.17 | 1.51 | 1.23 |
| FAS-associated death domain protein | FADD | 0.90 | 0.79 | 0.61* | 0.96 | 0.80 | 0.76 | 1.03 |
| GTP-binding protein 10 | GTPBP10 | 1.01 | 0.90 | 0.62* | 0.52* | 0.78 | 0.66* | 0.52* |
| Phosphorylase b kinase gamma catalytic chain, liver/testis isoform | PHKG2 | 0.73 | 0.76 | 0.62* | 0.58* | 0.34* | 0.52 | 0.30* |
| Transmembrane 4 L6 family member 4 | TM4SF4 | 0.69 | 0.68 | 0.62* | 0.67 | 0.36* | 0.58 | 0.59 |
| Keratin, type II cytoskeletal 1b | KRT77 | 0.89 | 0.80 | 0.63* | 1.23 | 2.31* | 0.70 | 1.26 |
| Kynurenine formamidase | AFMID | 0.96 | 0.74 | 0.63* | 0.86 | 0.60* | 0.58* | 0.56* |
| Cancer-related nucleoside-triphosphatase | NTPCR | 0.70* | 0.79 | 0.64* | 0.86 | 0.55* | 0.65 | 0.85 |
| Neutrophil gelatinase-associated lipocalin | LCN2 | 1.07 | 0.81 | 0.64* | 1.17 | 1.23 | 1.09 | 0.94 |
| Calcium-activated chloride channel regulator 1 | CLCA1 | 0.89 | 1.05 | 0.64* | 0.88 | 1.13 | 0.71 | 1.17 |
| TBC1 domain family member 17 | TBC1D17 | 0.73 | 0.69 | 0.65* | 0.65* | 0.98 | 0.78 | 0.76 |
| Interferon-induced protein with tetratricopeptide repeats 1 | IFIT1 | 0.75 | 1.08 | 0.65* | 2.44* | 3.27* | 3.50* | 2.42* |
| Inositol 1,4,5-trisphosphate receptor type 2 | ITPR2 | 0.94 | 0.85 | 0.65* | 0.64 | 0.58 | 0.66 | 0.55 |
| Transmembrane 4 L6 family member 20 | TM4SF20 | 1.61* | 0.93 | 0.65* | 0.63* | 1.51* | 0.97 | 0.50* |
| Equilibrative nucleoside transporter 3 | SLC29A3 | 1.00 | 0.90 | 0.65* | 0.97 | 1.15 | 0.74 | 0.89 |
| Pericentriolar material 1 protein | PCM1 | 0.68* | 0.68* | 0.65* | 0.61* | 0.75 | 0.76 | 0.69 |
| Mast/stem cell growth factor receptor Kit | KIT | 0.70 | 1.13 | 0.66* | 0.79 | 3.01* | 2.43* | 2.74* |
| 5'-AMP-activated protein kinase subunit beta-1 | PRKAB1 | 0.87 | 0.87 | 0.66* | 0.70 | 0.87 | 0.80 | 0.83 |
| High affinity cGMP-specific 3',5'-cyclic phosphodiesterase 9A | PDE9A | 1.41* | 1.01 | 0.66* | 1.16 | 1.37 | 1.05 | 0.74 |
| tRNA endonuclease ANKZF1 | ANKZF1 | 0.80 | 0.74 | 0.66* | 0.81 | 0.76 | 0.67 | 0.70 |
| Protein RUFY3 | RUFY3 | 0.98 | 0.84 | 0.66 | 0.88 | 0.87 | 0.76 | 1.21 |
| Sorting nexin-30 | SNX30 | 1.10 | 0.73 | 0.66* | 0.80 | 0.73 | 0.69 | 0.56* |
| Selenoprotein W | SELENOW | 1.07 | 0.68 | 0.66* | 0.86 | 0.51* | 0.45* | 0.40* |
| AP-5 complex subunit beta-1 | AP5B1 | 0.82 | 0.77 | 0.66* | 0.61* | 0.56 | 0.72 | 0.69 |
| Serine/threonine-protein kinase Sgk2 | SGK2 | 0.83 | 0.88 | 0.66* | 0.55* | 0.63 | 0.60* | 0.50* |
| Uncharacterized protein C2orf72 | C2orf72 | 1.13 | 0.75 | 0.66* | 0.64* | 0.05* | 0.39* | 0.08* |
| Metabotropic glycine receptor | GPR158 | 0.90 | 0.76 | 0.66* | 0.84 | 0.89 | 0.81 | 0.82 |
| Ribonuclease 7 | RNASE7 | 0.79 | 0.80 | 0.67* | 0.64* | 1.10 | 0.43* | 1.35 |
| Carbonic anhydrase 1 | CA1 | 0.97 | 0.86 | 0.67* | 0.66* | 0.70 | 0.66 | 0.66* |
| Cleavage and polyadenylation specificity factor subunit 3 | CPSF3 | 0.70* | 0.67* | 0.67* | 0.70* | 0.73 | 0.74 | 0.72 |

| | | | | | | | | |
|---|---|---|---|---|---|---|---|---|
| Protein FAM107B | FAM107B | 0.81 | 0.92 | 0.67* | 0.64 | 0.61 | 0.69 | 0.69 |
| KICSTOR complex protein ITFG2 | ITFG2 | 0.90 | 0.77 | 0.67* | 0.92 | 0.59 | 0.61 | 0.81 |

***E. Common down-regulated proteins between Aquamin and Aquamin plus Mesalamine [55 proteins]***

| | | Interventions | | | | | | |
|---|---|---|---|---|---|---|---|---|
| | | Control | | | *With LPS & Cytokines* | | | |
| Proteins | Genes | **AQ** | **AQ+MES** | MES | *LPS-Cyto* | *AQ* | *AQ+MES* | *MES* |
| Protein S100-A7A | S100A7A | 0.27* | 0.40* | 4.36* | 0.20* | 0.48* | 0.17* | 0.46* |
| Keratin, type I cytoskeletal 10 | KRT10 | 0.29* | 0.34* | 0.79* | 0.85 | 0.85 | 0.37* | 0.57* |
| Collagen alpha-3(VI) chain | COL6A3 | 0.29* | 0.50* | 1.58* | 1.31 | 0.81 | 1.25 | 1.92* |
| Actin filament-associated protein 1-like 2 | AFAP1L2 | 0.38* | 0.66 | 0.77 | 0.37* | 0.13* | 0.36* | 0.30* |
| Ubiquitin carboxyl-terminal hydrolase 27 | USP27X | 0.39* | 0.49* | 0.71* | 0.43* | 0.82 | 0.81 | 0.63 |
| Transmembrane protein 225B | TMEM225B | 0.44* | 0.39* | 0.80 | 0.51* | 0.70 | 1.23 | 0.59 |
| Cytosolic iron-sulfur assembly component 2B | CIAO2B | 0.45* | 0.60* | 1.29 | 0.47* | 0.38* | 1.57 | 1.87 |
| Arginase-1 | ARG1 | 0.45* | 0.50* | 0.84 | 0.93 | 0.87 | 0.60 | 0.98 |
| Neurogenic locus notch homolog protein 1 | NOTCH1 | 0.46* | 0.54* | 0.85 | 0.49* | 0.63 | 0.65 | 0.50* |
| Helicase SRCAP | SRCAP | 0.47* | 0.38* | 0.76 | 0.62 | 0.78 | 0.59 | 0.85 |
| CDK5 and ABL1 enzyme substrate 1 | CABLES1 | 0.48* | 0.56* | 0.68* | 0.48* | 1.04 | 0.89 | 0.90 |
| KICSTOR subunit 2 | KICS2 | 0.48* | 0.38* | 1.09 | 1.29 | 0.39* | 0.38* | 0.78 |
| Coiled-coil domain-containing protein 97 | CCDC97 | 0.49* | 0.65 | 1.06 | 0.56* | 0.43* | 0.38* | 0.55 |
| Chromobox protein homolog 8 | CBX8 | 0.49* | 0.66* | 0.71* | 0.63* | 0.26* | 0.20* | 0.51* |
| Uncharacterized protein C9orf85 | C9orf85 | 0.49* | 0.66 | 0.86 | 0.54* | 0.21* | 0.17* | 0.36* |
| Small proline-rich protein 2D | SPRR2D | 0.49* | 0.44* | 1.18 | 2.59* | 0.70 | 0.50* | 0.58* |
| High mobility group protein HMG-I/HMG-Y | HMGA1 | 0.50* | 0.61* | 1.13 | 1.38* | 1.08 | 1.34 | 1.87* |
| Nuclear factor 1 A-type | NFIA | 0.51* | 0.65* | 0.67* | 0.57* | 0.73 | 0.68 | 0.60* |
| Paladin | PALD1 | 0.51* | 0.59* | 0.69* | 0.60* | 0.46* | 0.56* | 0.54* |
| eIF-2-alpha kinase GCN2 | EIF2AK4 | 0.51* | 0.63 | 0.70 | 0.65 | 0.82 | 0.77 | 0.79 |
| Neuroguidin | NGDN | 0.52* | 0.50* | 0.71* | 0.85 | 0.74 | 0.82 | 0.74 |
| Inositol polyphosphate-4-phosphatase type I A | INPP4A | 0.52* | 0.62 | 0.73 | 0.89 | 1.91* | 1.37 | 1.56 |
| Receptor-type tyrosine-protein phosphatase F | PTPRF | 0.52* | 0.48* | 0.95 | 0.70* | 0.47* | 0.47* | 0.64* |
| Nuclear receptor subfamily 2 group F member 6 | NR2F6 | 0.53* | 0.55* | 0.85 | 1.03 | 0.36* | 0.53 | 0.69 |
| Proliferation marker protein Ki-67 | MKI67 | 0.53* | 0.44* | 0.69 | 0.89 | 0.49* | 0.62 | 1.02 |
| Protein arginine N-methyltransferase 3 | PRMT3 | 0.53* | 0.56* | 0.71* | 0.61* | 0.69 | 0.66 | 0.73 |
| Serine/threonine-protein phosphatase 6 regulatory ankyrin repeat subunit C | ANKRD52 | 0.54* | 0.63 | 0.71 | 0.49* | 0.75 | 0.59 | 0.49* |
| Condensin complex subunit 1 | NCAPD2 | 0.55* | 0.51* | 0.67* | 0.68* | 0.71 | 0.72 | 0.95 |

| | | | | | | | | |
|---|---|---|---|---|---|---|---|---|
| Mucosa-associated lymphoid tissue lymphoma translocation protein 1 | MALT1 | 0.55* | 0.63 | 0.79 | 0.62 | 0.67 | 0.61 | 0.58 |
| Integrator complex subunit 11 | INTS11 | 0.56* | 0.66* | 0.68* | 0.59* | 0.68 | 0.79 | 0.96 |
| Protein FAM110B | FAM110B | 0.57* | 0.56* | 0.69* | 0.47* | 0.56* | 0.52* | 0.45* |
| Coiled-coil domain-containing protein 124 | CCDC124 | 0.57* | 0.56* | 0.69 | 0.84 | 0.69 | 0.60 | 0.56 |
| Nephronectin | NPNT | 0.57* | 0.57* | 0.73 | 0.71 | 1.35 | 1.21 | 1.73 |
| SWI/SNF-related matrix-associated actin-dependent regulator of chromatin subfamily A-like protein 1 | SMARCAL1 | 0.57* | 0.55* | 0.67* | 0.31* | 0.77 | 0.53* | 0.44* |
| Ribosome biogenesis regulatory protein homolog | RRS1 | 0.57* | 0.64* | 0.98 | 1.07 | 0.46* | 0.53* | 0.78 |
| Fos-related antigen 1 | FOSL1 | 0.58* | 0.55* | 0.85 | 0.79 | 0.21* | 0.42* | 0.78 |
| Target of EGR1 protein 1 | TOE1 | 0.59* | 0.60* | 0.69* | 0.57* | 0.94 | 0.84 | 0.79 |
| RNA-binding protein NOB1 | NOB1 | 0.59* | 0.55* | 0.71* | 0.61* | 0.48* | 0.50* | 0.54* |
| Periodic tryptophan protein 2 homolog | PWP2 | 0.60* | 0.58* | 0.67* | 0.55* | 0.61* | 0.61 | 0.60* |
| Ephrin type-A receptor 2 | EPHA2 | 0.60* | 0.62* | 1.01 | 1.03 | 0.74* | 0.77 | 0.99 |
| DNA replication licensing factor MCM6 | MCM6 | 0.61* | 0.66* | 0.70* | 0.91 | 0.69 | 0.76 | 1.02 |
| General transcription and DNA repair factor IIH helicase subunit XPD | ERCC2 | 0.62* | 0.64* | 0.68* | 0.80 | 0.94 | 0.90 | 1.08 |
| Jupiter microtubule associated homolog 1 | JPT1 | 0.63* | 0.58* | 0.86 | 0.78 | 0.58 | 0.49* | 0.65 |
| Zinc finger C2HC domain-containing protein 1A | ZC2HC1A | 0.63* | 0.65* | 0.81 | 0.62* | 0.45* | 0.51* | 0.54* |
| Serine/threonine-protein kinase 11-interacting protein | STK11IP | 0.63* | 0.66 | 0.79 | 0.32* | 0.11* | 0.24* | 0.36* |
| ATP-dependent RNA helicase DDX51 | DDX51 | 0.64* | 0.66* | 0.72* | 0.66* | 0.55* | 0.60 | 0.70 |
| ATP-dependent RNA helicase DDX54 | DDX54 | 0.64* | 0.60* | 0.72 | 0.75 | 0.24* | 0.34* | 0.56 |
| Cyclin-dependent kinase 7 | CDK7 | 0.65 | 0.58* | 0.98 | 0.68 | 0.73 | 0.84 | 0.93 |
| Telomerase-binding protein EST1A | SMG6 | 0.65* | 0.61* | 0.87 | 0.65 | 0.60 | 0.67 | 0.66 |
| Keratin, type II cytoskeletal 5 | KRT5 | 0.65* | 0.63* | 0.87 | 1.64* | 1.09 | 0.72* | 0.86 |
| Ubiquitin carboxyl-terminal hydrolase 48 | USP48 | 0.66* | 0.45* | 0.72 | 0.53* | 0.62 | 0.70 | 0.74 |
| Ribosome biogenesis protein BOP1 | BOP1 | 0.66* | 0.58* | 0.70* | 0.77 | 0.53* | 0.54* | 0.56 |
| Unconventional myosin-Vc | MYO5C | 0.66 | 0.63* | 0.70 | 0.62* | 0.67 | 0.78 | 0.66 |
| Acyl-CoA (8-3)-desaturase | FADS1 | 0.67* | 0.65* | 0.70* | 0.36* | 0.52* | 0.44* | 0.41* |
| Spondin-1 | SPON1 | 0.67* | 0.66* | 1.05 | 0.73* | 0.56* | 0.49* | 0.69* |

***F. Common down-regulated proteins between Aquamin and Mesalamine [35 proteins]***

| | | Interventions | | | | | | |
|---|---|---|---|---|---|---|---|---|
| | | Control | | | *With LPS & Cytokines* | | | |
| Proteins | Genes | **AQ** | AQ+MES | **MES** | *LPS-Cyto* | *AQ* | *AQ+MES* | *MES* |
| Extracellular serine/threonine protein kinase FAM20C | FAM20C | 0.34* | 0.67 | 0.26* | 2.60* | 9.33* | 8.03* | 9.49* |
| Protein Shroom1 | SHROOM1 | 0.34* | 0.73 | 0.64* | 0.26* | 0.49* | 0.40* | 0.24* |

| | | | | | | | | |
|---|---|---|---|---|---|---|---|---|
| Serine/threonine-protein kinase WNK2 | WNK2 | 0.36* | 0.74 | 0.58* | 0.39* | 0.72 | 0.77 | 0.79 |
| Coilin | COIL | 0.37* | 0.67 | 0.64* | 0.77 | 1.39 | 1.58 | 1.54 |
| Zinc finger FYVE domain-containing protein 26 | ZFYVE26 | 0.41* | 0.71 | 0.58* | 0.36* | 0.55 | 0.65 | 0.44* |
| Keratin, type I cytoskeletal 25 | KRT25 | 0.44* | 1.22 | 0.25* | 0.41* | 1.70* | 0.23* | 1.17 |
| Protein DENND6B | DENND6B | 0.44* | 1.30 | 0.61* | 0.48* | 0.07* | 0.15* | 0.24* |
| Dermokine | DMKN | 0.47* | 0.70 | 0.58* | 0.64* | 1.58 | 0.79 | 1.33 |
| Progesterone-induced-blocking factor 1 | PIBF1 | 0.51* | 0.67* | 0.65* | 0.46* | 0.97 | 1.00 | 0.67 |
| Rapamycin-insensitive companion of mTOR | RICTOR | 0.52* | 0.69 | 0.57* | 0.47* | 0.57 | 0.72 | 0.69 |
| Zinc finger FYVE domain-containing protein 16 | ZFYVE16 | 0.52* | 0.76 | 0.62* | 0.59* | 0.40* | 0.71 | 0.60 |
| Rho GTPase-activating protein 32 | ARHGAP32 | 0.53* | 0.71 | 0.65* | 0.42* | 0.52* | 0.63 | 0.46* |
| Chromosome alignment-maintaining phosphoprotein 1 | CHAMP1 | 0.53* | 0.76 | 0.62* | 0.74 | 0.17* | 0.25* | 0.60* |
| Mediator of RNA polymerase II transcription subunit 22 | MED22 | 0.54* | 0.67 | 0.61* | 0.62* | 1.13 | 1.42 | 1.22 |
| Polyhomeotic-like protein 2 | PHC2 | 0.54* | 0.75 | 0.61* | 0.74 | 0.79 | 0.56 | 0.55 |
| Microtubule-associated tumor suppressor 1 | MTUS1 | 0.55* | 0.71 | 0.59* | 0.56* | 0.91 | 0.83 | 0.68 |
| Transmembrane protein 201 | TMEM201 | 0.55* | 0.68* | 0.64* | 0.55* | 0.22* | 0.38* | 0.44* |
| Myosin-2 | MYH2 | 0.56* | 0.80 | 0.45* | 1.12 | 5.57* | 4.93* | 4.82* |
| Pleckstrin | PLEK | 0.56* | 1.21 | 0.63* | 1.27 | 4.09* | 3.74* | 4.36* |
| Single-stranded DNA-binding protein 3 | SSBP3 | 0.56* | 0.77 | 0.66* | 0.58* | 1.24 | 1.30 | 1.34 |
| E3 ubiquitin-protein ligase TRIM36 | TRIM36 | 0.58* | 0.75 | 0.66* | 0.49* | 0.45* | 0.50* | 0.27* |
| Cleavage and polyadenylation specificity factor subunit 4 | CPSF4 | 0.61* | 0.69* | 0.66* | 0.55* | 0.69 | 0.65 | 0.61* |
| Serine/threonine-protein phosphatase 6 regulatory subunit 2 | PPP6R2 | 0.61* | 0.71* | 0.58* | 0.47* | 0.49* | 0.54* | 0.32* |
| Secreted Ly-6/uPAR domain-containing protein 2 | SLURP2 | 0.62* | 0.79 | 0.42* | 2.19* | 4.45* | 1.19 | 3.77* |
| Rho GTPase-activating protein 21 | ARHGAP21 | 0.62* | 0.85 | 0.64* | 0.69 | 1.05 | 0.85 | 0.73 |
| Aftiphilin | AFTPH | 0.62* | 0.68 | 0.63* | 0.72 | 0.96 | 0.84 | 0.86 |
| 3-hydroxy-3-methylglutaryl-coenzyme A reductase | HMGCR | 0.62* | 0.67 | 0.59* | 0.55* | 0.62 | 0.57 | 0.53* |
| Intermembrane lipid transfer protein VPS13D | VPS13D | 0.63* | 0.69 | 0.51* | 0.69 | 0.82 | 0.95 | 0.49* |
| A-kinase anchor protein 8-like | AKAP8L | 0.64* | 0.71 | 0.65* | 0.46* | 0.25* | 0.36* | 0.45* |
| Dynein axonemal heavy chain 8 | DNAH8 | 0.65* | 0.96 | 0.58* | 0.19* | 0.36* | 2.06* | 0.37* |
| Tripartite motif-containing protein 5 | TRIM5 | 0.65* | 0.68 | 0.64* | 0.55* | 0.75 | 0.87 | 0.57 |
| Uridine-cytidine kinase 2 | UCK2 | 0.65* | 0.69 | 0.67* | 0.44* | 0.39* | 0.36* | 0.34* |
| Kinase D-interacting substrate of 220 kDa | KIDINS220 | 0.66 | 0.81 | 0.67 | 0.81 | 0.76 | 0.85 | 0.57 |
| Pogo transposable element with ZNF domain | POGZ | 0.66* | 0.71 | 0.59* | 0.72 | 1.02 | 0.89 | 0.89 |
| Lipid droplet assembly factor 1 | LDAF1 | 0.67* | 0.75 | 0.66 | 0.50* | 0.54 | 0.57 | 0.41* |

***G. Common down-regulated proteins between Mesalamine and Aquamin plus Mesalamine [53 proteins]***

| | | Interventions |
|---|---|---|

| Proteins | Genes | Control AQ | Control **AQ+MES** | Control **MES** | *With LPS & Cytokines* *LPS-Cyto* | *AQ* | *AQ+MES* | *MES* |
|---|---|---|---|---|---|---|---|---|
| Glutathione S-transferase A2 | GSTA2 | 1.36* | 0.40* | 0.10* | 2.33* | 1.56* | 0.19* | 0.50* |
| Calcium/calmodulin-dependent protein kinase type 1B | PNCK | 1.12 | 0.39* | 0.14* | 1.00 | 1.60 | 0.82 | 0.13* |
| 3 beta-hydroxysteroid dehydrogenase/Delta 5-->4-isomerase type 2 | HSD3B2 | 1.43* | 0.44* | 0.24* | 0.88 | 1.24 | 0.30* | 0.21* |
| Meprin A subunit beta | MEP1B | 1.18 | 0.37* | 0.25* | 1.11 | 1.11 | 0.36* | 0.29* |
| UPF0235 protein C15orf40 | C15orf40 | 0.86 | 0.40* | 0.33* | 1.30 | 0.91 | 0.45* | 0.67 |
| Carboxypeptidase O | CPO | 1.36* | 0.54* | 0.34* | 1.41* | 1.27 | 0.41* | 0.39* |
| Beta-1,3-N-acetylglucosaminyltransferase lunatic fringe | LFNG | 0.67* | 0.50* | 0.41* | 0.90 | 3.55* | 2.99* | 3.10* |
| Ornithine transcarbamylase, mitochondrial | OTC | 1.13 | 0.62* | 0.42* | 0.86 | 1.03 | 0.41* | 0.48* |
| Gastrotropin | FABP6 | 1.05 | 0.52* | 0.43* | 0.66* | 0.55* | 0.35* | 0.48* |
| Tripartite motif-containing protein 3 | TRIM3 | 0.85 | 0.53* | 0.45* | 0.64* | 0.79 | 0.92 | 0.52 |
| Arylamine N-acetyltransferase 1 | NAT1 | 0.88 | 0.42* | 0.46* | 0.46* | 0.77 | 0.60 | 0.62 |
| Iodotyrosine deiodinase 1 | IYD | 0.94 | 0.57* | 0.46* | 0.68* | 1.05 | 0.57* | 0.51* |
| Glutathione S-transferase A1 | GSTA1 | 0.72* | 0.38* | 0.47* | 0.45* | 0.49* | 0.64 | 0.41* |
| Mitochondrial inner membrane protease ATP23 homolog | ATP23 | 1.03 | 0.58* | 0.47* | 0.82 | 0.73 | 0.58 | 0.46* |
| Profilin-3 | PFN3 | 2.06* | 0.44* | 0.48* | 0.82 | 0.81 | 0.84 | 0.37* |
| Mediator of RNA polymerase II transcription subunit 8 | MED8 | 3.06* | 0.51* | 0.49* | 22.20* | 45.95* | 32.32* | 21.41* |
| Solute carrier family 13 member 2 | SLC13A2 | 0.79 | 0.59* | 0.49* | 0.99 | 0.97 | 0.46* | 0.81 |
| Integrin alpha-1 | ITGA1 | 0.90 | 0.54* | 0.51* | 0.80 | 0.98 | 0.51* | 0.52* |
| ADP-ribosylation factor-like protein 14 | ARL14 | 0.93 | 0.60* | 0.53* | 0.92 | 0.82 | 0.60 | 0.63 |
| Pleckstrin homology-like domain family B member 1 | PHLDB1 | 0.73* | 0.61* | 0.53* | 0.70* | 0.41* | 0.32* | 0.43* |
| Death-associated protein 1 | DAP | 0.68 | 0.53* | 0.55* | 0.77 | 0.45* | 0.43* | 0.48* |
| Fatty acyl-CoA reductase 2 | FAR2 | 0.82 | 0.49* | 0.55* | 0.52* | 0.11* | 0.25* | 0.27* |
| Mitochondrial tRNA methylthiotransferase CDK5RAP1 | CDK5RAP1 | 0.68 | 0.62* | 0.56* | 0.69 | 0.60 | 0.56 | 0.58 |
| DNA-directed RNA polymerase I subunit RPA1 | POLR1A | 0.74* | 0.60* | 0.57* | 0.61* | 0.73 | 0.64 | 0.71 |
| DNA replication licensing factor MCM7 | MCM7 | 0.82 | 0.61* | 0.58* | 0.83 | 0.93 | 0.94 | 1.16 |
| Bridge-like lipid transfer protein family member 1 | BLTP1 | 0.98 | 0.65 | 0.59* | 0.88 | 1.01 | 0.80 | 0.58 |
| Trefoil factor 2 | TFF2 | 1.26 | 0.61* | 0.60* | 0.42* | 0.59 | 0.34* | 0.33* |
| Alpha-2A adrenergic receptor | ADRA2A | 0.76 | 0.55* | 0.60* | 0.82 | 0.88 | 0.76 | 0.72 |
| DNA mismatch repair protein Msh6 | MSH6 | 0.85 | 0.62* | 0.61* | 0.60* | 0.81 | 0.76 | 0.63 |
| SH2 domain-containing protein 3A | SH2D3A | 0.73* | 0.66* | 0.61* | 0.66* | 0.45* | 0.51* | 0.50* |
| Keratin, type I cytoskeletal 17 | KRT17 | 0.71* | 0.64* | 0.61* | 1.72* | 0.68 | 0.55* | 0.77 |
| Inhibitor of growth protein 1 | ING1 | 1.02 | 0.47* | 0.62* | 0.86 | 1.16 | 0.73 | 0.59 |
| DNA methyltransferase 1-associated protein 1 | DMAP1 | 0.69 | 0.59* | 0.62* | 0.73 | 0.93 | 0.89 | 0.84 |
| Beta-chimaerin | CHN2 | 0.86 | 0.57* | 0.62* | 0.94 | 0.85 | 0.56* | 0.58* |

| | | | | | | | | |
|---|---|---|---|---|---|---|---|---|
| Mediator of DNA damage checkpoint protein 1 | MDC1 | 0.91 | 0.65 | 0.62* | 0.70 | 0.35* | 0.50* | 1.13 |
| Target of rapamycin complex subunit LST8 | MLST8 | 0.68* | 0.67* | 0.63* | 0.78 | 1.17 | 1.14 | 0.85 |
| Trafficking protein particle complex subunit 10 | TRAPPC10 | 0.69* | 0.66* | 0.63* | 0.55* | 0.71 | 0.72 | 0.54* |
| Ribosomal protein S6 kinase alpha-4 | RPS6KA4 | 0.69* | 0.47* | 0.63* | 0.61* | 0.49* | 0.63 | 0.55* |
| ATPase MORC2 | MORC2 | 0.67* | 0.63* | 0.64* | 0.76 | 1.04 | 0.99 | 1.02 |
| L-fucose kinase | FCSK | 0.87 | 0.67* | 0.64* | 0.79 | 0.87 | 0.77 | 0.88 |
| Pre-mRNA-splicing factor ATP-dependent RNA helicase PRP16 | DHX38 | 0.84 | 0.66* | 0.64* | 0.68* | 0.65 | 0.66 | 0.68* |
| F-box/LRR-repeat protein 18 | FBXL18 | 0.74* | 0.59* | 0.64* | 0.70 | 0.60 | 0.61 | 0.67 |
| Protein TASOR | TASOR | 0.74 | 0.65 | 0.64* | 0.68 | 0.54 | 0.68 | 0.76 |
| Ephrin-A2 | EFNA2 | 0.88 | 0.60* | 0.64* | 0.88 | 0.70 | 0.70 | 0.55* |
| TGF-beta receptor type-2 | TGFBR2 | 0.83 | 0.58* | 0.65* | 0.69 | 0.94 | 0.64 | 0.60 |
| Alanyl-tRNA editing protein Aarsd1 | AARSD1 | 0.76 | 0.66* | 0.65* | 0.82 | 0.74 | 0.73 | 0.80 |
| Transforming acidic coiled-coil-containing protein 1 | TACC1 | 0.94 | 0.62* | 0.65* | 0.84 | 0.56* | 0.47* | 0.61 |
| Eukaryotic translation initiation factor 2D | EIF2D | 0.75 | 0.61* | 0.65* | 0.73 | 0.40* | 0.77 | 0.65 |
| IQ motif and SEC7 domain-containing protein 2 | IQSEC2 | 0.74 | 0.64 | 0.66* | 0.69 | 0.89 | 0.72 | 0.85 |
| Alcohol dehydrogenase 6 | ADH6 | 1.07 | 0.66* | 0.66* | 0.79 | 0.97 | 0.65* | 0.74 |
| Large subunit GTPase 1 homolog | LSG1 | 0.76* | 0.64* | 0.66* | 0.65* | 0.54* | 0.52* | 0.52* |
| Atypical kinase COQ8B, mitochondrial | COQ8B | 0.71 | 0.60* | 0.66* | 0.60* | 0.50* | 0.61 | 0.72 |
| DNA-directed RNA polymerase II subunit RPB3 | POLR2C | 0.78 | 0.60* | 0.66* | 1.00 | 0.69 | 0.70 | 0.92 |

Values represent the abundance ratio from organoids (n=4 subjects) compared to the control. These proteins were down-regulated at a 1.5-fold change (<2% FDR). Corresponding abundance ratios from the other treatment groups are provided for comparison. Proteins common among groups and unique to individual groups under control conditions are presented. *Indicates significance compared to the control (at $p<0.05$).
