## Supplementary material for "Proteomic Profile of Human Colon Organoids: Effects of a multi-mineral intervention alone and in the presence of pro-Inflammatory and anti-inflammatory treatments": S Table 5

**Supplement Table 5. Up-regulated proteins influenced by LPS-Cytokines alone and with Aquamin and Mesalamine (with 1.5-fold)**

***A. Common among four groups: LPS-Cytokines, +Aquamin, +Mesalamine and +Aquamin plus Mesalamine [296 proteins]***

| | | Interventions | | | | | | |
|---|---|---|---|---|---|---|---|---|
| | | Control | | | *With LPS & Cytokines* | | | |
| Proteins | Genes | AQ | AQ+MES | MES | ***LPS-Cyto*** | ***AQ*** | ***AQ+MES*** | ***MES*** |
| HLA class II histocompatibility antigen, DRB1 beta chain | HLA-DRB1 | 0.99 | 1.11 | 1.30 | 36.37* | 27.95* | 34.64* | 33.75* |
| Guanylate-binding protein 7 | GBP7 | 1.02 | 2.02* | 0.88 | 35.72* | 32.72* | 30.59* | 33.79* |
| HLA class II histocompatibility antigen, DR alpha chain | HLA-DRA | 1.05 | 1.75* | 1.05 | 28.26* | 24.40* | 28.76* | 24.96* |
| Bromodomain-containing protein 8 | BRD8 | 1.65* | 0.96 | 0.74 | 26.61* | 35.18* | 48.74* | 52.93* |
| Mediator of RNA polymerase II transcription subunit 8 | MED8 | 3.06* | 0.51* | 0.49* | 22.20* | 45.95* | 32.32* | 21.41* |
| Guanylate-binding protein 4 | GBP4 | 1.91* | 0.78 | 0.94 | 22.11* | 26.71* | 25.20* | 24.08* |
| Programmed cell death 1 ligand 1 | CD274 | 0.43* | 1.53 | 1.44 | 16.94* | 16.35* | 19.58* | 15.89* |
| HLA class II histocompatibility antigen, DP beta 1 chain | HLA-DPB1 | 0.31* | 2.33* | 1.80* | 15.14* | 13.55* | 15.68* | 15.01* |
| Indoleamine 2,3-dioxygenase 1 | IDO1 | 0.88 | 0.96 | 0.89 | 15.01* | 11.04* | 10.46* | 12.89* |
| Secreted and transmembrane protein 1 | SECTM1 | 1.01 | 1.76* | 1.78* | 14.70* | 8.17* | 7.82* | 13.21* |
| HLA class II histocompatibility antigen, DR beta 3 chain | HLA-DRB3 | 0.90 | 1.00 | 1.64* | 12.42* | 9.92* | 10.03* | 11.55* |
| Tryptophan--tRNA ligase, cytoplasmic | WARS1 | 0.89 | 1.08 | 1.05 | 11.26* | 9.10* | 9.83* | 10.44* |
| Putative histone H2B type 2-C | H2BC20P | 2.73* | 0.76 | 0.85 | 10.59* | 34.44* | 25.80* | 21.48* |
| Apolipoprotein C-II | APOC2 | 2.38* | 0.68 | 0.99 | 9.94* | 21.64* | 14.48* | 17.84* |
| HLA class II histocompatibility antigen gamma chain | CD74 | 1.27 | 1.28 | 1.16 | 9.37* | 9.26* | 9.53* | 7.79* |
| Heat shock 70 kDa protein 1-like | HSPA1L | 2.61* | 1.48 | 1.29 | 8.97* | 15.03* | 9.11* | 10.55* |
| HLA class II histocompatibility antigen, DP alpha 1 chain | HLA-DPA1 | 0.99 | 0.88 | 1.17 | 8.78* | 8.23* | 8.36* | 9.34* |
| Vesicle-associated membrane protein 5 | VAMP5 | 1.13 | 1.43 | 1.57 | 7.65* | 6.90* | 6.61* | 6.29* |
| HLA class I histocompatibility antigen, B alpha chain | HLA-B | 1.47* | 2.37* | 1.52* | 7.62* | 5.91* | 7.94* | 8.75* |
| Protein-glutamine gamma-glutamyltransferase 2 | TGM2 | 1.11 | 1.32* | 1.16 | 7.41* | 7.72* | 8.15* | 7.52* |
| Guanylate-binding protein 1 | GBP1 | 1.00 | 1.01 | 1.08 | 5.93* | 5.84* | 6.03* | 6.15* |
| Ubiquitin/ISG15-conjugating enzyme E2 L6 | UBE2L6 | 1.34 | 1.56* | 1.29 | 5.90* | 5.70* | 6.47* | 6.58* |
| Protein bassoon | BSN | 0.50* | 2.53* | 1.96* | 5.37* | 19.19* | 17.24* | 29.63* |
| NADH dehydrogenase [ubiquinone] 1 beta subcomplex subunit 2, mitochondrial | NDUFB2 | 2.06* | 1.92* | 3.85* | 5.33* | 9.24* | 8.41* | 5.38* |
| HLA class I histocompatibility antigen, alpha chain F | HLA-F | 1.23 | 1.49* | 1.32 | 5.25* | 4.42* | 5.26* | 5.03* |
| Sorting nexin-24 | SNX24 | 3.83* | 4.44* | 3.16* | 5.18* | 5.74* | 3.20* | 4.30* |
| Thymidine phosphorylase | TYMP | 1.24 | 1.52* | 1.28 | 4.52* | 3.92* | 4.88* | 4.23* |
| Methionine-R-sulfoxide reductase B2, mitochondrial | MSRB2 | 1.00 | 1.10 | 0.78 | 4.44* | 13.63* | 14.13* | 14.06* |

| | | | | | | | | |
|---|---|---|---|---|---|---|---|---|
| Tapasin | TAPBP | 1.23* | 1.27* | 1.26* | 4.39* | 4.05* | 4.83* | 3.93* |
| Nitric oxide synthase, inducible | NOS2 | 1.07 | 1.14 | 1.22 | 4.35* | 3.78* | 5.27* | 3.94* |
| Signal transducer and activator of transcription 1-alpha/beta | STAT1 | 1.04 | 1.05 | 1.01 | 4.34* | 4.13* | 4.36* | 4.41* |
| Cytosol aminopeptidase | LAP3 | 0.96 | 1.09 | 0.97 | 4.32* | 4.45* | 4.43* | 3.93* |
| Caspase-1 | CASP1 | 1.07 | 1.17 | 1.45 | 4.31* | 3.38* | 3.74* | 4.07* |
| Antigen peptide transporter 1 | TAP1 | 1.06 | 1.23 | 1.08 | 4.28* | 4.37* | 4.87* | 4.06* |
| Equilibrative nucleoside transporter 1 | SLC29A1 | 2.51* | 2.80* | 2.70* | 4.14* | 4.67* | 4.44* | 3.78* |
| HLA class I histocompatibility antigen, A alpha chain | HLA-A | 1.14 | 1.36* | 1.18* | 4.09* | 4.31* | 4.64* | 3.93* |
| Tumor necrosis factor receptor superfamily member 5 | CD40 | 0.84 | 1.23 | 1.53 | 3.99* | 4.56* | 4.44* | 4.22* |
| Antigen peptide transporter 2 | TAP2 | 0.97 | 1.03 | 1.03 | 3.89* | 4.20* | 4.32* | 3.62* |
| Beta-2-microglobulin | B2M | 1.10 | 1.32* | 1.10 | 3.77* | 4.13* | 4.31* | 4.05* |
| S-adenosylmethionine-dependent nucleotide dehydratase RSAD2 | RSAD2 | 0.94 | 1.33 | 1.23 | 3.73* | 6.25* | 5.47* | 2.08* |
| Interferon-induced GTP-binding protein Mx1 | MX1 | 1.07 | 1.34 | 1.50* | 3.61* | 4.42* | 5.17* | 4.06* |
| Apolipoprotein L2 | APOL2 | 0.92 | 1.01 | 0.97 | 3.60* | 3.29* | 3.98* | 3.24* |
| Hepatocyte growth factor | HGF | 1.29* | 0.68* | 0.82 | 3.59* | 11.24* | 9.91* | 8.32* |
| Proteasome subunit beta type-10 | PSMB10 | 1.06 | 1.34 | 1.22 | 3.51* | 2.90* | 4.61* | 3.18* |
| HLA class I histocompatibility antigen, C alpha chain | HLA-C | 1.10 | 1.34* | 1.24* | 3.50* | 3.60* | 4.35* | 3.49* |
| Serum amyloid A-4 protein | SAA4 | 0.63* | 2.09* | 2.47* | 3.32* | 9.01* | 8.67* | 16.73* |
| Microfibril-associated glycoprotein 4 | MFAP4 | 0.79 | 0.80 | 1.01 | 3.31* | 11.58* | 10.17* | 10.31* |
| Interferon-induced protein with tetratricopeptide repeats 2 | IFIT2 | 1.14 | 0.95 | 1.00 | 3.31* | 4.23* | 5.12* | 2.98* |
| Lipocalin-1 | LCN1 | 1.05 | 0.73 | 0.87 | 3.28* | 5.55* | 3.52* | 5.52* |
| Gamma-interferon-inducible lysosomal thiol reductase | IFI30 | 1.04 | 1.13 | 1.17 | 3.26* | 3.09* | 3.31* | 2.96* |
| Protein FAM234B | FAM234B | 1.38 | 1.35 | 1.46* | 3.26* | 5.10* | 4.90* | 2.41* |
| Retinoic acid receptor responder protein 1 | RARRES1 | 1.60* | 2.72* | 3.25* | 3.23* | 3.43* | 6.05* | 6.66* |
| Cytochrome b | MT-CYB | 2.22* | 1.67* | 1.38 | 3.21* | 5.40* | 5.01* | 3.12* |
| Phospholipase A2, membrane associated | PLA2G2A | 1.31 | 4.25* | 3.95* | 3.18* | 8.37* | 15.64* | 17.81* |
| Transmembrane and immunoglobulin domain-containing protein 1 | TMIGD1 | 2.53* | 7.38* | 3.86* | 3.17* | 3.35* | 4.41* | 2.89* |
| HLA class I histocompatibility antigen, alpha chain E | HLA-E | 0.98 | 1.36 | 1.22 | 3.08* | 2.16* | 2.05* | 2.19* |
| Group IID secretory phospholipase A2 | PLA2G2D | 1.94* | 0.98 | 0.85 | 3.07* | 7.57* | 9.66* | 8.40* |
| Solute carrier family 40 member 1 | SLC40A1 | 1.66* | 1.70* | 1.81* | 3.07* | 4.44* | 3.81* | 3.78* |
| Homeobox protein DBX1 | DBX1 | 1.57* | 1.54* | 1.49* | 3.07* | 6.91* | 6.60* | 5.35* |
| Endoplasmic reticulum aminopeptidase 2 | ERAP2 | 1.30* | 1.34* | 1.20* | 3.05* | 2.93* | 2.65* | 2.76* |
| Sodium-coupled neutral amino acid transporter 5 | SLC38A5 | 0.85 | 1.24 | 1.29 | 3.03* | 4.78* | 3.88* | 4.40* |
| Uncharacterized protein KIAA0040 | KIAA0040 | 1.32 | 1.26 | 1.38 | 2.96* | 3.16* | 2.75* | 3.21* |

| | | | | | | | | |
|---|---|---|---|---|---|---|---|---|
| Putative HLA class I histocompatibility antigen, alpha chain H | HLA-H | 1.19 | 1.64* | 1.17 | 2.96* | 2.63* | 2.82* | 2.42* |
| All-trans-retinol dehydrogenase [NAD(+)] ADH4 | ADH4 | 1.07 | 2.55* | 2.71* | 2.95* | 2.62* | 2.01* | 1.81* |
| Butyrophilin subfamily 3 member A3 | BTN3A3 | 0.89 | 1.45 | 1.31 | 2.95* | 2.00* | 2.40* | 2.94* |
| Complement factor I | CFI | 0.68 | 2.00* | 2.08* | 2.94* | 6.10* | 6.03* | 11.20* |
| Alpha-1-antitrypsin | SERPINA1 | 1.26* | 1.76* | 1.41* | 2.93* | 2.81* | 2.44* | 2.34* |
| Cathepsin O | CTSO | 0.99 | 1.06 | 0.77 | 2.92* | 3.74* | 3.50* | 2.61* |
| Filamin-C | FLNC | 1.23 | 0.84 | 1.90* | 2.91* | 3.79* | 4.00* | 2.41* |
| Myeloid leukemia factor 2 | MLF2 | 2.19* | 1.97* | 3.37* | 2.90* | 2.56* | 1.98* | 2.36* |
| Zinc finger protein 45 | ZNF45 | 0.85 | 0.94 | 0.79 | 2.89* | 7.63* | 8.24* | 8.00* |
| Neutral amino acid transporter A | SLC1A4 | 1.27 | 1.32 | 1.48* | 2.87* | 5.21* | 5.15* | 4.25* |
| Calcium uniporter regulatory subunit MCUb, mitochondrial | MCUB | 1.24* | 1.28* | 1.27* | 2.86* | 3.39* | 3.50* | 3.08* |
| Sodium-coupled neutral amino acid symporter 2 | SLC38A2 | 1.13 | 1.31 | 1.44* | 2.86* | 2.53* | 3.21* | 3.48* |
| Xylosyl- and glucuronyltransferase LARGE1 | LARGE1 | 1.38 | 1.47 | 1.40 | 2.86* | 5.42* | 4.76* | 3.33* |
| Ubiquitin D | UBD | 1.03 | 4.25* | 3.84* | 2.84* | 3.54* | 12.39* | 12.06* |
| Complement C3 | C3 | 0.68* | 2.23* | 3.76* | 2.82* | 3.40* | 4.41* | 3.80* |
| Keratinocyte-associated transmembrane protein 2 | KCT2 | 1.12 | 1.69* | 1.51* | 2.82* | 2.19* | 2.57* | 2.90* |
| 2'-5'-oligoadenylate synthase 3 | OAS3 | 1.09 | 1.29 | 1.03 | 2.80* | 3.02* | 3.38* | 3.00* |
| Cystine/glutamate transporter | SLC7A11 | 1.28 | 0.94 | 1.01 | 2.78* | 5.27* | 4.98* | 2.84* |
| Neuronal calcium sensor 1 | NCS1 | 0.67 | 1.11 | 1.15 | 2.78* | 3.34* | 3.17* | 2.49* |
| Guanylate-binding protein 2 | GBP2 | 1.05 | 1.05 | 1.09 | 2.75* | 2.72* | 2.89* | 2.95* |
| E3 ubiquitin-protein ligase MSL2 | MSL2 | 0.47* | 0.40* | 0.51* | 2.74* | 11.31* | 10.04* | 11.22* |
| Pro-opiomelanocortin | POMC | 0.96 | 2.08* | 2.21* | 2.71* | 4.33* | 6.13* | 10.26* |
| PDZ and LIM domain protein 4 | PDLIM4 | 1.23 | 1.00 | 1.37 | 2.70* | 2.24* | 2.23* | 2.43* |
| Apolipoprotein L1 | APOL1 | 1.46 | 1.73* | 1.82* | 2.68* | 3.15* | 3.75* | 2.64* |
| Protein unc-93 homolog A | UNC93A | 0.51* | 1.04 | 1.49 | 2.62* | 1.95* | 2.34* | 3.58* |
| Plexin domain-containing protein 2 | PLXDC2 | 1.16 | 1.24 | 1.67* | 2.61* | 5.06* | 7.61* | 8.74* |
| Extracellular serine/threonine protein kinase FAM20C | FAM20C | 0.34* | 0.67 | 0.26* | 2.60* | 9.33* | 8.03* | 9.49* |
| Fibrinogen gamma chain | FGG | 0.35* | 1.28 | 2.38* | 2.58* | 3.47* | 3.42* | 6.21* |
| Intercellular adhesion molecule 1 | ICAM1 | 0.83 | 0.89 | 1.02 | 2.57* | 2.62* | 2.53* | 2.58* |
| Major facilitator superfamily domain-containing protein 8 | MFSD8 | 1.61* | 1.27 | 1.09 | 2.56* | 4.01* | 3.96* | 3.35* |
| Transmembrane protein 143 | TMEM143 | 1.27* | 1.40 | 1.43* | 2.56* | 4.25* | 3.90* | 3.12* |
| Keratinocyte-associated protein 2 | KRTCAP2 | 1.54* | 1.29 | 1.18 | 2.55* | 4.86* | 4.63* | 2.87* |
| WD repeat domain phosphoinositide-interacting protein 1 | WIPI1 | 1.14 | 1.11 | 1.19 | 2.55* | 6.55* | 2.34* | 3.16* |
| 2-aminomuconic semialdehyde dehydrogenase | ALDH8A1 | 1.90* | 1.21 | 1.32 | 2.50* | 13.33* | 30.36* | 5.64* |
| Tetraspanin-9 | TSPAN9 | 0.64 | 0.93 | 0.79 | 2.49* | 7.29* | 7.91* | 7.67* |

| | | | | | | | | |
|---|---|---|---|---|---|---|---|---|
| Zinc transporter ZIP9 | SLC39A9 | 1.28 | 1.40 | 1.31 | 2.48* | 4.55* | 4.80* | 3.72* |
| DNA-binding protein SMUBP-2 | IGHMBP2 | 0.58* | 1.53* | 1.72* | 2.47* | 3.42* | 3.38* | 5.07* |
| Secretogranin-3 | SCG3 | 1.21 | 1.79* | 1.95* | 2.47* | 2.79* | 3.40* | 4.48* |
| Guanylate-binding protein 5 | GBP5 | 0.75 | 0.94 | 0.97 | 2.46* | 2.69* | 2.45* | 2.83* |
| Sodium/myo-inositol cotransporter | SLC5A3 | 1.68* | 1.70* | 1.41 | 2.45* | 3.59* | 4.91* | 2.00* |
| Betaine--homocysteine S-methyltransferase 1 | BHMT | 0.31* | 0.41* | 0.31* | 2.44* | 13.21* | 10.27* | 6.92* |
| Interferon-induced protein with tetratricopeptide repeats 1 | IFIT1 | 0.75 | 1.08 | 0.65* | 2.44* | 3.27* | 3.50* | 2.42* |
| cAMP-dependent protein kinase inhibitor beta | PKIB | 0.88 | 6.48* | 9.41* | 2.44* | 2.06* | 11.12* | 18.67* |
| Proteasome activator complex subunit 2 | PSME2 | 1.08 | 1.16 | 1.02 | 2.43* | 2.30* | 2.48* | 2.36* |
| Beta-2-glycoprotein 1 | APOH | 1.26 | 2.33* | 2.52* | 2.43* | 2.14* | 2.35* | 3.00* |
| Albumin | ALB | 0.63* | 1.73* | 2.02* | 2.42* | 1.96* | 2.70* | 2.09* |
| Lactotransferrin | LTF | 0.98 | 1.77* | 2.37* | 2.39* | 2.02* | 2.78* | 3.12* |
| Tetraspanin-33 | TSPAN33 | 1.76* | 1.53 | 1.49 | 2.39* | 3.91* | 2.87* | 2.46* |
| High affinity cationic amino acid transporter 1 | SLC7A1 | 1.76* | 1.42* | 1.56* | 2.39* | 3.41* | 3.56* | 2.53* |
| Insulin-like growth factor-binding protein complex acid labile subunit | IGFALS | 0.23* | 0.37* | 0.33* | 2.38* | 12.16* | 9.41* | 11.08* |
| Heme transporter FLVCR1 | FLVCR1 | 1.18 | 1.20 | 1.29 | 2.36* | 4.38* | 4.25* | 3.94* |
| Ubiquitin-like protein ISG15 | ISG15 | 1.08 | 1.13 | 1.07 | 2.34* | 1.92* | 2.65* | 2.14* |
| Intercellular adhesion molecule 2 | ICAM2 | 1.34* | 1.14 | 1.49* | 2.34* | 2.32* | 1.74* | 2.27* |
| Coiled-coil domain-containing protein 39 | CCDC39 | 0.41* | 0.77 | 1.31 | 2.33* | 2.48* | 2.44* | 2.90* |
| Endoplasmic reticulum membrane adapter protein XK | XK | 1.59* | 1.81* | 1.36 | 2.33* | 3.37* | 4.41* | 2.84* |
| Peroxisome proliferator-activated receptor delta | PPARD | 1.01 | 1.05 | 1.11 | 2.31* | 3.67* | 4.20* | 6.07* |
| V-type immunoglobulin domain-containing suppressor of T-cell activation | VSIR | 1.60* | 1.43 | 1.49* | 2.30* | 5.02* | 3.78* | 3.72* |
| Inositol-3-phosphate synthase 1 | ISYNA1 | 1.33* | 1.61* | 1.53* | 2.28* | 3.96* | 2.92* | 3.27* |
| DnaJ homolog subfamily C member 15 | DNAJC15 | 1.21 | 1.83* | 1.61* | 2.28* | 3.11* | 2.09* | 2.92* |
| Tropomodulin-2 | TMOD2 | 0.97 | 2.08* | 1.93* | 2.25* | 1.95* | 2.91* | 4.70* |
| Signal transducer and activator of transcription 2 | STAT2 | 1.09 | 1.06 | 1.13 | 2.24* | 2.52* | 2.72* | 2.89* |
| FXYD domain-containing ion transport regulator 5 | FXYD5 | 3.37* | 3.09* | 2.13* | 2.24* | 5.59* | 4.94* | 4.29* |
| TPA-induced transmembrane protein | TTMP | 1.81* | 1.96* | 1.42 | 2.23* | 4.60* | 4.29* | 3.70* |
| Vang-like protein 1 | VANGL1 | 1.17 | 1.03 | 1.13 | 2.23* | 4.05* | 4.10* | 3.12* |
| Protein BCAP | ODF2L | 0.43* | 1.24 | 1.79* | 2.23* | 2.03* | 2.99* | 3.70* |
| All-trans-retinol dehydrogenase [NAD(+)] ADH7 | ADH7 | 0.29* | 0.43* | 0.31* | 2.22* | 11.41* | 8.22* | 10.30* |
| Cytochrome c oxidase assembly factor 1 homolog | COA1 | 1.14 | 1.44 | 1.12 | 2.21* | 4.42* | 3.51* | 3.77* |
| Protein YIF1A | YIF1A | 1.55* | 1.60 | 1.42 | 2.21* | 3.72* | 3.42* | 2.33* |
| Sulfhydryl oxidase 2 | QSOX2 | 1.47* | 1.88* | 1.74* | 2.20* | 3.05* | 3.18* | 3.04* |
| Liprin-alpha-3 | PPFIA3 | 1.23 | 1.43 | 1.34* | 2.20* | 3.75* | 3.49* | 2.77* |

| | | | | | | | | |
|---|---|---|---|---|---|---|---|---|
| HLA class I histocompatibility antigen, alpha chain G | HLA-G | 1.14 | 1.45* | 1.31 | 2.20* | 2.70* | 2.49* | 1.93* |
| Nuclease EXOG, mitochondrial | EXOG | 1.57* | 1.36 | 1.45 | 2.19* | 3.66* | 3.66* | 3.13* |
| Transmembrane protein 186 | TMEM186 | 1.20 | 1.41 | 1.16 | 2.18* | 2.89* | 2.98* | 2.50* |
| Peptidyl-prolyl cis-trans isomerase FKBP11 | FKBP11 | 0.99 | 0.87 | 1.33* | 2.18* | 5.12* | 4.89* | 4.49* |
| Complement factor B | CFB | 1.12 | 1.46 | 1.67* | 2.18* | 3.23* | 2.38* | 2.44* |
| Hemoglobin subunit beta | HBB | 1.03 | 2.23* | 1.89* | 2.18* | 2.36* | 3.19* | 4.34* |
| Glycerophosphodiester phosphodiesterase 1 | GDE1 | 1.65* | 1.63* | 1.36* | 2.17* | 3.02* | 3.04* | 1.72* |
| EF-hand calcium-binding domain-containing protein 14 | EFCAB14 | 1.26 | 1.38 | 1.36 | 2.16* | 2.55* | 2.99* | 2.12* |
| Mitochondrial glutamate carrier 1 | SLC25A22 | 1.07 | 1.35* | 1.12 | 2.16* | 2.36* | 2.38* | 2.04* |
| E3 ubiquitin-protein ligase TRIM21 | TRIM21 | 1.24 | 1.08 | 1.31* | 2.13* | 1.75* | 2.06* | 1.95* |
| Collagen alpha-1(XII) chain | COL12A1 | 0.71 | 1.58 | 1.57* | 2.12* | 3.69* | 6.05* | 6.67* |
| Aldehyde oxidase | AOX1 | 0.14* | 0.14* | 0.27* | 2.11* | 11.69* | 8.47* | 9.08* |
| Lymphocyte antigen 6D | LY6D | 0.89 | 1.56 | 2.25* | 2.10* | 2.22* | 1.53 | 1.51 |
| Interleukin-32 | IL32 | 1.22 | 1.36 | 1.26 | 2.10* | 2.35* | 2.49* | 2.12* |
| Inter-alpha-trypsin inhibitor heavy chain H4 | ITIH4 | 0.56* | 1.42 | 1.59* | 2.09* | 1.62* | 2.06* | 3.18* |
| Tetranectin | CLEC3B | 0.72 | 1.19 | 2.37* | 2.09* | 1.76* | 2.13* | 2.78* |
| Proteasome activator complex subunit 1 | PSME1 | 0.99 | 1.28* | 1.17* | 2.09* | 1.89* | 2.22* | 1.95* |
| Apolipoprotein C-III | APOC3 | 0.91 | 2.22* | 1.66* | 2.09* | 2.28* | 2.65* | 3.96* |
| Vitamin K-dependent protein Z | PROZ | 0.80 | 0.77 | 0.91 | 2.08* | 5.00* | 4.05* | 4.90* |
| Thrombospondin type-1 domain-containing protein 4 | THSD4 | 1.04 | 0.98 | 1.23 | 2.06* | 12.07* | 6.18* | 3.67* |
| Fermitin family homolog 2 | FERMT2 | 0.05* | 0.35* | 0.25* | 2.05* | 9.27* | 9.50* | 10.36* |
| Tapasin-related protein | TAPBPL | 1.19 | 1.24 | 1.12 | 2.05* | 1.86* | 2.18* | 1.93* |
| HLA class II histocompatibility antigen, DR beta 5 chain | HLA-DRB5 | 0.87 | 1.01 | 0.77 | 2.05* | 2.43* | 1.98* | 2.20* |
| Proteasome subunit beta type-9 | PSMB9 | 1.05 | 1.16 | 1.14 | 2.04* | 1.77* | 2.18* | 1.90* |
| Vascular cell adhesion protein 1 | VCAM1 | 0.54* | 1.13 | 1.26 | 2.04* | 3.97* | 5.56* | 6.73* |
| Laminin subunit alpha-4 | LAMA4 | 1.09 | 0.93 | 0.89 | 2.04* | 2.27* | 2.54* | 2.81* |
| Heparin cofactor 2 | SERPIND1 | 0.75 | 1.73* | 2.75* | 2.04* | 3.83* | 4.33* | 7.38* |
| Zinc finger CCCH-type with G patch domain-containing protein | ZGPAT | 1.03 | 0.97 | 0.95 | 2.00* | 1.95* | 2.17* | 2.19* |
| Proteasome subunit beta type-8 | PSMB8 | 1.37* | 1.51* | 1.39* | 2.00* | 1.62* | 2.82* | 1.92* |
| Beta-1,4-galactosyltransferase 5 | B4GALT5 | 1.06 | 1.31 | 1.42 | 2.00* | 3.85* | 3.02* | 3.50* |
| Probable ATP-dependent RNA helicase DDX60 | DDX60 | 1.09 | 1.18 | 1.04 | 2.00* | 2.38* | 2.40* | 2.03* |
| Shiftless antiviral inhibitor of ribosomal frameshifting protein | SHFL | 1.10 | 1.04 | 1.06 | 1.99* | 2.01* | 1.82* | 1.79 |
| Solute carrier family 66 member 2 | SLC66A2 | 1.55* | 1.44* | 1.44* | 1.99* | 4.16* | 3.88* | 3.56* |
| Interferon-induced protein with tetratricopeptide repeats 3 | IFIT3 | 1.24 | 2.65* | 2.64* | 1.98* | 2.49* | 4.45* | 4.39* |
| Tubulin beta-1 chain | TUBB1 | 0.21* | 0.28* | 0.24* | 1.96* | 9.06* | 7.77* | 9.62* |

| | | | | | | | | |
|---|---|---|---|---|---|---|---|---|
| Short transmembrane mitochondrial protein 1 | STMP1 | 1.49* | 1.43 | 1.46 | 1.95* | 5.09* | 4.29* | 4.19* |
| Beta-enolase | ENO3 | 0.50* | 0.75 | 0.76* | 1.95* | 5.84* | 4.93* | 4.93* |
| Battenin | CLN3 | 1.57* | 2.13* | 1.44* | 1.94* | 1.87* | 2.41* | 2.39* |
| Mitochondrial import receptor subunit TOM40B | TOMM40L | 1.43* | 1.59* | 1.46* | 1.94* | 2.74* | 2.90* | 2.29* |
| Pyridine nucleotide-disulfide oxidoreductase domain-containing protein 2 | PYROXD2 | 1.28 | 1.08 | 1.19 | 1.94* | 2.66* | 2.32* | 2.26* |
| Glycolipid transfer protein domain-containing protein 2 | GLTPD2 | 1.47* | 2.85* | 2.85* | 1.93* | 2.81* | 2.12* | 3.41* |
| Electrogenic aspartate/glutamate antiporter SLC25A12, mitochondrial | SLC25A12 | 1.34* | 1.20 | 1.21 | 1.93* | 2.84* | 2.60* | 2.64* |
| Protein mono-ADP-ribosyltransferase PARP9 | PARP9 | 0.96 | 0.91 | 0.90 | 1.92* | 1.60* | 1.98* | 2.10* |
| Fibrinogen beta chain | FGB | 0.56* | 2.04* | 1.67* | 1.92* | 3.53* | 3.54* | 6.17* |
| Cathepsin S | CTSS | 1.09 | 1.16 | 1.03 | 1.92* | 1.57* | 1.81* | 1.57* |
| LHFPL tetraspan subfamily member 2 protein | LHFPL2 | 1.51* | 1.61* | 1.23 | 1.92* | 2.46* | 1.60 | 1.57 |
| Trophoblast glycoprotein | TPBG | 1.42* | 1.68* | 1.61* | 1.90* | 2.67* | 2.69* | 1.69 |
| Visinin-like protein 1 | VSNL1 | 1.20* | 1.18 | 1.08 | 1.89* | 1.79* | 1.73* | 1.95* |
| Alpha-2-macroglobulin | A2M | 0.61* | 1.48* | 1.87* | 1.88* | 1.77* | 2.51* | 3.48* |
| Triggering receptor expressed on myeloid cells 1 | TREM1 | 2.25* | 2.93* | 1.88* | 1.88* | 4.12* | 1.94* | 3.89* |
| Divergent protein kinase domain 2A | DIPK2A | 1.22 | 1.08 | 1.01 | 1.88* | 2.86* | 3.03* | 2.68* |
| Neprilysin | MME | 1.22 | 1.33 | 1.17 | 1.86* | 3.35* | 3.31* | 2.68* |
| SPARC | SPARC | 0.54* | 1.40* | 1.85* | 1.86* | 1.88* | 3.08* | 3.36* |
| Legumain | LGMN | 1.14 | 1.28 | 1.09 | 1.85* | 2.08* | 2.39* | 1.53* |
| Coiled-coil domain-containing protein 127 | CCDC127 | 1.41 | 1.65* | 1.53 | 1.85* | 2.71* | 2.49* | 2.29* |
| Stromal interaction molecule 2 | STIM2 | 0.77 | 1.30 | 1.75* | 1.84* | 1.54 | 2.36* | 3.21* |
| PAT complex subunit Asterix | WDR83OS | 1.51* | 1.46 | 1.31 | 1.84* | 5.64* | 4.74* | 4.39* |
| Alpha-2-antiplasmin | SERPINF2 | 0.86 | 1.62* | 1.67* | 1.84* | 2.47* | 5.09* | 6.18* |
| Probable U3 small nucleolar RNA-associated protein 11 | UTP11 | 0.88 | 0.77 | 0.91 | 1.84* | 2.62* | 2.45* | 1.93* |
| Leukocyte surface antigen CD47 | CD47 | 1.08 | 1.22 | 1.19* | 1.84* | 2.10* | 2.04* | 1.74* |
| Immunoglobulin lambda-1 light chain | | 0.85 | 1.19 | 2.66* | 1.84* | 3.41* | 4.29* | 5.54* |
| Complement C5 | C5 | 1.02 | 1.33 | 1.48* | 1.83* | 3.14* | 4.28* | 5.49* |
| Laminin subunit alpha-5 | LAMA5 | 1.04 | 1.44 | 1.45* | 1.83* | 1.58 | 1.98* | 2.57* |
| Complement C4-A | C4A | 0.73* | 1.29* | 2.17* | 1.82* | 3.29* | 3.86* | 4.65* |
| Thrombospondin-4 | THBS4 | 0.67* | 1.01 | 1.08 | 1.82* | 3.14* | 4.76* | 6.66* |
| Collagen alpha-1(IV) chain | COL4A1 | 0.50* | 1.17 | 1.70* | 1.82* | 2.23* | 2.22* | 3.55* |
| Zinc finger protein 536 | ZNF536 | 0.40* | 1.44 | 1.10 | 1.81* | 2.08* | 2.70* | 5.62* |
| Mucin-1 | MUC1 | 1.22* | 1.12 | 1.05 | 1.81* | 1.92* | 1.94* | 1.52* |
| F-box only protein 6 | FBXO6 | 1.11 | 1.10 | 1.29 | 1.81* | 1.73* | 1.77* | 2.13* |
| Hemoglobin subunit epsilon | HBE1 | 0.77 | 1.36 | 1.95* | 1.80* | 2.27* | 2.55* | 3.81* |
| Protocadherin-12 | PCDH12 | 0.47* | 1.35 | 1.06 | 1.79* | 2.87* | 4.80* | 6.92* |

| | | | | | | | | |
|---|---|---|---|---|---|---|---|---|
| Tubulin alpha-3C chain | TUBA3C | 0.07* | 0.27* | 0.11* | 1.78* | 10.04* | 7.83* | 8.39* |
| Sex-determining region Y protein | SRY | 0.61* | 1.19 | 1.25 | 1.77* | 1.72 | 1.71 | 2.48* |
| MIT domain-containing protein 1 | MITD1 | 1.24 | 0.74 | 0.95 | 1.77* | 2.40* | 2.21* | 2.51* |
| Signal peptide peptidase-like 2B | SPPL2B | 1.26 | 1.61* | 1.48* | 1.77* | 2.29* | 2.26* | 2.37* |
| Protransforming growth factor alpha | TGFA | 1.48* | 1.58* | 1.65* | 1.77* | 3.24* | 3.42* | 3.06* |
| Metallophosphoesterase 1 | MPPE1 | 1.08 | 1.37 | 1.03 | 1.77* | 3.07* | 3.52* | 3.24* |
| Carboxypeptidase B2 | CPB2 | 0.23* | 1.02 | 1.24 | 1.76* | 5.38* | 5.30* | 10.11* |
| Lymphocyte function-associated antigen 3 | CD58 | 1.37* | 1.92* | 1.66* | 1.76* | 2.20* | 2.42* | 2.22* |
| Deoxyhypusine synthase | DHPS | 0.95 | 1.73* | 1.01 | 1.75* | 1.58* | 1.88* | 1.77* |
| Bcl-2 homologous antagonist/killer | BAK1 | 1.21 | 1.16 | 1.24 | 1.75* | 1.83* | 1.80* | 1.75* |
| Heat shock 70 kDa protein 6 | HSPA6 | 1.26* | 1.15 | 1.29* | 1.75* | 2.82* | 3.06* | 2.61* |
| Transmembrane reductase CYB561D2 | CYB561D2 | 1.24 | 1.23 | 1.32 | 1.74* | 2.33* | 1.99* | 1.98* |
| Magnesium transporter MRS2 homolog, mitochondrial | MRS2 | 1.30 | 1.34 | 1.19 | 1.74* | 1.90* | 2.64* | 2.09* |
| Thyroxine-binding globulin | SERPINA7 | 0.70* | 1.20 | 1.31* | 1.74* | 2.91* | 3.91* | 4.50* |
| Coagulation factor V | F5 | 0.61* | 1.22 | 1.26 | 1.74* | 1.61 | 1.89* | 2.43* |
| Argininosuccinate synthase | ASS1 | 1.01 | 1.12 | 1.04 | 1.74* | 1.50* | 1.78* | 1.82* |
| Superoxide dismutase [Mn], mitochondrial | SOD2 | 1.20* | 1.69* | 1.12 | 1.74* | 1.96* | 2.13* | 2.03* |
| Arylsulfatase L | ARSL | 1.47* | 1.34* | 1.26 | 1.74* | 2.76* | 3.07* | 2.11* |
| Cartilage oligomeric matrix protein | COMP | 0.57* | 0.84 | 1.22 | 1.73* | 2.91* | 4.29* | 6.18* |
| Osteomodulin | OMD | 0.71 | 1.32 | 1.18 | 1.73* | 2.97* | 4.38* | 5.35* |
| Homologous recombination OB-fold protein | HROB | 0.65* | 1.35* | 1.48* | 1.73* | 1.51* | 2.16* | 1.96* |
| Guided entry of tail-anchored proteins factor 1 | GET1 | 1.03 | 1.19 | 1.10 | 1.73* | 2.26* | 2.28* | 1.64 |
| Macrophage mannose receptor 1 | MRC1 | 0.51* | 0.40* | 0.41* | 1.72* | 6.02* | 7.32* | 7.08* |
| CCR4-NOT transcription complex subunit 7 | CNOT7 | 1.10 | 1.02 | 1.01 | 1.72* | 1.86* | 2.13* | 2.12* |
| Xyloside xylosyltransferase 1 | XXYLT1 | 1.34 | 1.32 | 1.20 | 1.72* | 1.72* | 1.59 | 1.53 |
| Adenosine deaminase | ADA | 0.90 | 1.07 | 1.23 | 1.71* | 1.51 | 1.66* | 1.73* |
| Proteoglycan 4 | PRG4 | 0.74 | 1.15 | 1.30 | 1.71* | 3.11* | 5.08* | 5.60* |
| 2'-5'-oligoadenylate synthase 2 | OAS2 | 0.80 | 3.38* | 3.47* | 1.71* | 2.58* | 9.01* | 8.23* |
| Thrombomodulin | THBD | 0.55* | 0.62* | 0.60* | 1.70* | 7.47* | 6.68* | 7.22* |
| Cell growth regulator with EF hand domain protein 1 | CGREF1 | 1.45* | 1.11 | 1.03 | 1.70* | 3.02* | 3.03* | 2.29* |
| Vitamin D-binding protein | GC | 0.71* | 1.25 | 1.61* | 1.69* | 1.51* | 2.10* | 2.04* |
| Inter-alpha-trypsin inhibitor heavy chain H3 | ITIH3 | 0.66* | 1.15 | 1.11 | 1.68* | 3.30* | 4.88* | 6.20* |
| Sideroflexin-2 | SFXN2 | 1.15 | 1.13 | 1.21 | 1.68* | 2.33* | 2.13* | 1.79* |
| Hemopexin | HPX | 0.65* | 1.28 | 1.58* | 1.68* | 1.54 | 1.99* | 3.50* |
| Complement component C8 alpha chain | C8A | 0.60* | 1.16 | 1.38* | 1.68* | 3.50* | 4.58* | 6.06* |
| STARD3 N-terminal-like protein | STARD3NL | 1.12 | 1.56* | 1.24 | 1.68* | 1.74 | 1.55 | 1.57 |
| Antiviral innate immune response receptor RIG-I | RIGI | 1.05 | 1.14 | 0.93 | 1.68* | 1.84* | 1.91* | 1.56 |

| | | | | | | | | |
|---|---|---|---|---|---|---|---|---|
| E3 ubiquitin-protein ligase RNF213 | RNF213 | 1.05 | 0.94 | 1.06 | 1.68* | 1.91* | 1.85* | 1.84* |
| Sacsin | SACS | 1.37* | 1.56* | 1.45* | 1.67* | 2.82* | 2.91* | 2.32* |
| Adhesion G-protein coupled receptor F1 | ADGRF1 | 1.32* | 1.87* | 1.24 | 1.65* | 1.98* | 1.80* | 1.81* |
| N-acyl-phosphatidylethanolamine-hydrolyzing phospholipase D | NAPEPLD | 1.10 | 1.08 | 1.10 | 1.65* | 1.70* | 1.59 | 1.63 |
| Mitochondrial calcium uniporter regulator 1 | MCUR1 | 1.25* | 1.48* | 1.36* | 1.64* | 2.36* | 2.42* | 2.06* |
| Beta-1,3-galactosyl-O-glycosyl-glycoprotein beta-1,6-N-acetylglucosaminyltransferase | GCNT1 | 1.12 | 1.43 | 1.33 | 1.64* | 2.69* | 2.00* | 1.56 |
| Derlin-2 | DERL2 | 0.83 | 1.02 | 1.18 | 1.64* | 1.74* | 1.70 | 2.16* |
| Apolipoprotein E | APOE | 0.90 | 1.42* | 1.50* | 1.64* | 1.69* | 1.93* | 2.28* |
| Kinesin-like protein KIF6 | KIF6 | 1.14 | 0.64 | 0.99 | 1.63* | 1.91* | 2.10* | 1.79 |
| Mitochondrial import inner membrane translocase subunit Tim17-B | TIMM17B | 1.21 | 1.12 | 1.22 | 1.63* | 2.34* | 2.51* | 2.27* |
| Hepatocyte growth factor-like protein | MST1 | 1.03 | 1.45* | 1.60* | 1.62* | 2.56* | 3.35* | 4.24* |
| Hemoglobin subunit alpha | HBA1 | 0.78* | 1.53* | 1.52* | 1.62* | 1.74* | 2.44* | 3.35* |
| Transgelin | TAGLN | 0.82 | 2.04* | 1.68* | 1.62* | 1.71 | 2.44* | 2.91* |
| Inter-alpha-trypsin inhibitor heavy chain H2 | ITIH2 | 0.86 | 1.71* | 1.75* | 1.61* | 2.05* | 2.42* | 3.06* |
| Laminin subunit alpha-1 | LAMA1 | 0.55* | 2.01* | 1.66* | 1.60* | 2.30* | 2.88* | 3.96* |
| Vesicle transport protein SFT2B | SFT2D2 | 1.27 | 1.27 | 1.21 | 1.60* | 1.75* | 1.59 | 1.81* |
| Parathyroid hormone/parathyroid hormone-related peptide receptor | PTH1R | 0.35* | 0.33* | 0.47* | 1.60* | 8.24* | 7.16* | 7.08* |
| Interferon-induced transmembrane protein 3 | IFITM3 | 0.94 | 2.48* | 1.97* | 1.60* | 2.12* | 4.68* | 3.56* |
| Collagen alpha-1(XVIII) chain | COL18A1 | 0.84 | 1.26 | 1.26 | 1.60* | 1.50* | 1.92* | 1.65* |
| Zinc transporter ZIP6 | SLC39A6 | 1.48* | 1.24 | 1.13 | 1.59* | 1.66* | 1.58* | 1.82* |
| Olfactomedin-like protein 3 | OLFML3 | 0.35* | 1.30 | 0.87 | 1.59* | 3.28* | 5.46* | 7.10* |
| Transmembrane protein 62 | TMEM62 | 0.91 | 1.13 | 1.22 | 1.59* | 1.71* | 1.82* | 1.91* |
| Lysophosphatidylserine lipase ABHD12 | ABHD12 | 1.20 | 1.22 | 1.12 | 1.59* | 1.53* | 1.61* | 1.77* |
| Mitochondrial adenyl nucleotide antiporter SLC25A25 | SLC25A25 | 1.30 | 1.53 | 1.24 | 1.58* | 2.23* | 2.58* | 1.80* |
| Equilibrative nucleoside transporter 2 | SLC29A2 | 1.07 | 0.93 | 0.99 | 1.58* | 1.89* | 1.84* | 1.84* |
| Mitochondrial pyruvate carrier 1 | MPC1 | 0.99 | 1.83* | 1.33 | 1.58 | 2.75* | 2.41* | 2.70* |
| Vitronectin | VTN | 0.53* | 1.33 | 1.51* | 1.58* | 1.55 | 1.93* | 2.76* |
| Beta-galactoside alpha-2,6-sialyltransferase 1 | ST6GAL1 | 1.01 | 1.00 | 1.17 | 1.58* | 3.45* | 2.72* | 2.43* |
| Laminin subunit beta-1 | LAMB1 | 0.54* | 2.04* | 1.65* | 1.58* | 2.39* | 2.83* | 3.78* |
| Gasdermin-D | GSDMD | 1.07 | 1.24 | 0.96 | 1.57* | 1.64* | 1.81* | 1.59* |
| BMP-binding endothelial regulator protein | BMPER | 0.65 | 1.20 | 1.13 | 1.57* | 3.53* | 3.94* | 4.84* |
| Cathepsin L2 | CTSV | 0.87 | 0.99 | 1.14 | 1.57* | 1.53 | 1.52 | 1.54 |
| Apolipoprotein M | APOM | 0.51* | 0.86 | 1.24* | 1.56* | 2.35* | 3.87* | 5.61* |
| Plasminogen activator inhibitor 2 | SERPINB2 | 1.06 | 1.31 | 1.15 | 1.56* | 1.64 | 1.67 | 1.70 |

| | | | | | | | | |
|---|---|---|---|---|---|---|---|---|
| Regulator of microtubule dynamics protein 3 | RMDN3 | 1.19* | 1.27* | 1.16 | 1.56* | 1.76* | 1.81* | 1.56* |
| 3-ketodihydrosphingosine reductase | KDSR | 1.14 | 1.50 | 1.32 | 1.56* | 1.73 | 1.65 | 1.50 |
| Ribonuclease P protein subunit p25 | RPP25 | 1.02 | 1.07 | 1.07 | 1.55 | 1.87* | 1.58 | 1.90* |
| Interferon-stimulated 20 kDa exonuclease-like 2 | ISG20L2 | 1.32 | 1.12 | 1.33 | 1.55 | 2.06* | 2.39* | 2.47* |
| Vacuolar ATPase assembly integral membrane protein VMA21 | VMA21 | 1.29 | 1.16 | 1.02 | 1.54 | 2.33* | 2.16* | 1.98 |
| ER membrane protein complex subunit 7 | EMC7 | 0.97 | 1.51* | 1.43* | 1.54* | 1.50 | 1.59 | 1.99* |
| DNA polymerase subunit gamma-2, mitochondrial | POLG2 | 1.24 | 1.21 | 1.11 | 1.54* | 1.53 | 1.63 | 1.57 |
| Laminin subunit gamma-1 | LAMC1 | 0.61* | 1.97* | 1.62* | 1.54* | 2.28* | 2.68* | 3.65* |
| Bone marrow stromal antigen 2 | BST2 | 0.90 | 0.90 | 0.79 | 1.53 | 1.98* | 2.11* | 1.81 |
| Spectrin beta chain, erythrocytic | SPTB | 0.65 | 0.70 | 1.24 | 1.53 | 3.31* | 3.32* | 4.60* |
| Mediator of RNA polymerase II transcription subunit 18 | MED18 | 0.93 | 0.97 | 1.07 | 1.53* | 2.41* | 1.91* | 2.08* |
| UMP-CMP kinase 2, mitochondrial | CMPK2 | 0.96 | 1.64* | 1.39 | 1.53 | 1.76* | 2.93* | 2.32* |
| Immunoglobulin superfamily member 8 | IGSF8 | 1.11 | 1.61* | 1.44* | 1.53* | 2.10* | 1.93* | 1.61* |
| CMP-N-acetylneuraminate-beta-galactosamide-alpha-2,3-sialyltransferase 1 | ST3GAL1 | 0.97 | 1.05 | 1.11 | 1.53* | 2.23* | 2.22* | 1.84* |
| Adhesion G-protein coupled receptor G7 | ADGRG7 | 1.13 | 1.34 | 1.22 | 1.53 | 1.55 | 1.92* | 1.56 |
| Glutathione peroxidase 3 | GPX3 | 0.72 | 1.00 | 1.35* | 1.52* | 2.96* | 2.09* | 2.86* |
| Sodium/hydrogen exchanger 8 | SLC9A8 | 1.39* | 1.34 | 1.33* | 1.52* | 2.76* | 2.64* | 2.00* |
| Carbamoyl-phosphate synthase [ammonia], mitochondrial | CPS1 | 0.47* | 1.17 | 0.96 | 1.52* | 4.01* | 4.48* | 5.28* |
| Alpha-amylase 1B | AMY1B | 0.60* | 1.01 | 1.13 | 1.52* | 3.33* | 3.94* | 5.43* |
| Surfeit locus protein 1 | SURF1 | 1.27 | 1.28 | 1.12 | 1.51* | 2.16* | 2.07* | 1.84* |
| Adenylyl cyclase-associated protein 2 | CAP2 | 0.49* | 1.23 | 1.16 | 1.51* | 3.15* | 3.71* | 4.96* |
| Prothrombin | F2 | 0.84 | 1.59 | 1.40 | 1.51* | 2.10* | 3.12* | 3.81* |

***B. Up-regulated proteins unique to LPS-Cytokines (LPS-Cyto) [38 proteins]***

| | | Interventions | | | | | | |
|---|---|---|---|---|---|---|---|---|
| | | Control | | | *With LPS & Cytokines* | | | |
| Proteins | Genes | AQ | AQ+MES | MES | ***LPS-Cyto*** | *AQ* | *AQ+MES* | *MES* |
| Small proline-rich protein 2D | SPRR2D | 0.49* | 0.44* | 1.18 | 2.59* | 0.70 | 0.50* | 0.58* |
| Putative nucleoside diphosphate kinase | NME2P1 | 2.99* | 1.90* | 1.48* | 2.39* | 0.55* | 0.75 | 0.60* |
| Nucleoplasmin-3 | NPM3 | 1.99* | 1.31 | 1.49 | 2.28* | 0.37* | 0.65 | 0.30* |
| Allograft inflammatory factor 1-like | AIF1L | 1.33* | 0.98 | 1.34* | 2.00* | 0.61* | 0.64 | 1.17 |
| Zinc-alpha-2-glycoprotein | AZGP1 | 0.88 | 1.21 | 1.26 | 2.00* | 1.12 | 0.56* | 1.08 |
| Hepatoma-derived growth factor-related protein 3 | HDGFL3 | 1.36* | 1.48* | 1.35* | 1.86* | 0.90 | 0.82 | 1.29 |
| Protein max | MAX | 1.28* | 1.31 | 1.42* | 1.85* | 0.24* | 0.39* | 0.75 |
| Putative heat shock protein HSP 90-alpha A4 | HSP90AA4P | 1.25 | 1.35 | 1.36 | 1.84* | 0.78 | 0.94 | 1.30 |

| | | | | | | | | |
|---|---|---|---|---|---|---|---|---|
| Keratin, type I cytoskeletal 14 | KRT14 | 0.51* | 0.49* | 0.57* | 1.84* | 0.71* | 0.52* | 0.61* |
| Keratin, type II cytoskeletal 6B | KRT6B | 0.76* | 0.70* | 0.95 | 1.81* | 1.04 | 0.91 | 1.11 |
| DNA polymerase epsilon subunit 3 | POLE3 | 1.42* | 1.44 | 1.47 | 1.76* | 1.10 | 0.98 | 0.64 |
| Molybdopterin synthase sulfur carrier subunit | MOCS2 | 1.38 | 1.37 | 1.47* | 1.73* | 0.73 | 0.69 | 1.01 |
| Keratin, type I cytoskeletal 17 | KRT17 | 0.71* | 0.64* | 0.61* | 1.72* | 0.68 | 0.55* | 0.77 |
| Protein S100-A7 | S100A7 | 0.33* | 0.70 | 0.94 | 1.71* | 0.60 | 1.01 | 1.30 |
| N-acetyltransferase ESCO1 | ESCO1 | 1.20 | 0.85 | 1.78* | 1.70* | 1.20 | 0.90 | 1.49 |
| Actin-related protein 3B | ACTR3B | 1.21* | 1.28 | 1.27* | 1.69* | 1.17 | 1.10 | 1.14 |
| Protein disulfide-isomerase A2 | PDIA2 | 2.04* | 1.32 | 1.24 | 1.68* | 0.72 | 0.87 | 1.13 |
| Transmembrane protein 115 | TMEM115 | 1.15 | 1.37 | 1.11 | 1.67* | 0.81 | 1.00 | 1.46 |
| Keratin, type II cytoskeletal 5 | KRT5 | 0.65* | 0.63* | 0.87 | 1.64* | 1.09 | 0.72* | 0.86 |
| Keratin, type II cytoskeletal 6A | KRT6A | 0.49* | 1.00 | 1.23 | 1.61* | 0.93 | 1.36 | 1.29 |
| PC4 and SFRS1-interacting protein | PSIP1 | 1.51* | 1.06 | 1.31* | 1.61* | 0.48* | 0.47* | 1.46* |
| Small nuclear ribonucleoprotein F | SNRPF | 2.07* | 1.62* | 1.69* | 1.59* | 0.18* | 0.89 | 0.54 |
| Sodium-coupled monocarboxylate transporter 2 | SLC5A12 | 1.14 | 1.13 | 0.87 | 1.59* | 1.39 | 0.68 | 0.57 |
| Keratin, type I cytoskeletal 16 | KRT16 | 0.57* | 0.74 | 0.81 | 1.57* | 0.80 | 0.85 | 1.03 |
| Arf-GAP with GTPase, ANK repeat and PH domain-containing protein 1 | AGAP1 | 1.36 | 1.20 | 1.47 | 1.56 | 0.69 | 0.89 | 1.04 |
| Nuclear transcription factor Y subunit alpha | NFYA | 1.20 | 1.08 | 1.17 | 1.55* | 0.52* | 0.32* | 0.82 |
| tRNA-dihydrouridine(47) synthase [NAD(P)(+)]-like | DUS3L | 1.03 | 1.35 | 1.33 | 1.55* | 1.22 | 1.21 | 1.46 |
| Protein TEX261 | TEX261 | 1.42 | 1.46 | 1.20 | 1.55* | 1.20 | 0.89 | 1.01 |
| Syntaxin-10 | STX10 | 1.17 | 1.05 | 1.20 | 1.54* | 0.85 | 0.94 | 1.10 |
| Tumor necrosis factor receptor superfamily member 6 | FAS | 1.10 | 1.01 | 1.19 | 1.54* | 1.34 | 1.34 | 1.40* |
| Serum response factor-binding protein 1 | SRFBP1 | 1.01 | 1.10 | 1.12 | 1.54* | 0.80 | 0.63 | 1.07 |
| Sodium- and chloride-dependent neutral and basic amino acid transporter B(0+) | SLC6A14 | 0.91 | 1.09 | 1.08 | 1.53* | 1.41 | 1.38 | 1.44 |
| C-C chemokine receptor-like 2 | CCRL2 | 0.92 | 0.77 | 0.98 | 1.53 | 1.20 | 1.15 | 0.96 |
| Cyclin-dependent kinase inhibitor 2A | CDKN2A | 1.01 | 1.03 | 1.18 | 1.53* | 0.32* | 0.36* | 0.49* |
| Small integral membrane protein 24 | SMIM24 | 1.45 | 1.71* | 0.90 | 1.51 | 1.28 | 1.22 | 0.90 |
| Ubiquitin domain-containing protein 2 | UBTD2 | 1.17 | 1.41 | 1.34 | 1.51 | 1.02 | 1.32 | 1.08 |
| Lysosomal acid phosphatase | ACP2 | 0.92 | 1.08 | 1.06 | 1.51* | 1.45* | 1.41 | 1.35* |
| Solute carrier organic anion transporter family member 4A1 | SLCO4A1 | 0.96 | 1.29 | 1.13 | 1.50* | 1.27 | 1.02 | 1.32 |

***C. Up-regulated proteins unique to Aquamin (AQ) with LPS-Cytokines [64 proteins]***

| | | Interventions | | | | | | |
|---|---|---|---|---|---|---|---|---|
| | | Control | | | *With LPS & Cytokines* | | | |
| Proteins | Genes | AQ | AQ+MES | MES | *LPS-Cyto* | ***AQ*** | *AQ+MES* | *MES* |

| | | | | | | | | |
|---|---|---|---|---|---|---|---|---|
| S-adenosyl-L-methionine-dependent tRNA 4-demethylwyosine synthase TYW1 | TYW1 | 0.98 | 0.77 | 0.69* | 1.30 | 4.94* | 0.92 | 0.83 |
| Haptoglobin | HP | 0.70* | 1.32* | 3.72* | 1.15 | 3.32* | 1.47* | 1.19 |
| Peptidyl-tRNA hydrolase | PTRH1 | 1.02 | 1.23 | 1.02 | 1.25 | 2.58* | 1.25 | 1.26 |
| Arachidonate 12-lipoxygenase, 12R-type | ALOX12B | 2.22* | 1.37 | 7.85* | 0.47* | 2.58* | 0.92 | 1.08 |
| Keratin, type II cytoskeletal 1b | KRT77 | 0.89 | 0.80 | 0.63* | 1.23 | 2.31* | 0.70 | 1.26 |
| Gasdermin-A | GSDMA | 1.05 | 1.25 | 1.66* | 1.00 | 2.19* | 0.72 | 0.83 |
| Membrane-spanning 4-domains subfamily A member 10 | MS4A10 | 1.73* | 2.21* | 1.28 | 1.40 | 2.17* | 1.20 | 1.03 |
| Retinol-binding protein 2 | RBP2 | 1.29* | 2.84* | 1.78* | 1.20 | 2.02* | 1.36* | 1.25* |
| Keratin, type I cytoskeletal 9 | KRT9 | 0.46* | 0.44* | 0.46* | 1.11 | 1.99* | 0.51* | 0.89 |
| Phosphatidylinositol 3-kinase regulatory subunit beta | PIK3R2 | 1.34 | 1.33 | 1.44 | 1.23 | 1.93* | 1.27 | 1.19 |
| Keratin, type II cytoskeletal 78 | KRT78 | 0.71* | 0.67* | 1.26* | 1.50* | 1.92* | 0.51* | 1.49* |
| 5'-nucleotidase domain-containing protein 3 | NT5DC3 | 1.25* | 1.00 | 0.86 | 1.42* | 1.89* | 1.46* | 1.33 |
| Sorbin and SH3 domain-containing protein 1 | SORBS1 | 0.97 | 2.18* | 1.43 | 1.40 | 1.86* | 1.35 | 1.24 |
| Transmembrane protein 125 | TMEM125 | 1.53* | 1.26 | 0.95 | 1.26 | 1.86* | 0.98 | 1.15 |
| Alpha-2-macroglobulin-like protein 1 | A2ML1 | 1.27* | 2.46* | 5.56* | 0.41* | 1.85* | 0.34* | 1.36* |
| Nuclear factor 1 C-type | NFIC | 0.55* | 0.74 | 0.78 | 1.06 | 1.84* | 1.42 | 1.30 |
| Desmocollin-3 | DSC3 | 1.72* | 9.40* | 34.18* | 0.65 | 1.84* | 0.93 | 0.01* |
| OTU domain-containing protein 4 | OTUD4 | 0.76 | 0.99 | 0.99 | 1.05 | 1.83* | 1.10 | 1.24 |
| Aspartate dehydrogenase domain-containing protein | ASPDH | 1.49* | 2.23* | 1.22 | 1.34 | 1.82* | 1.07 | 1.07 |
| Protein GPR108 | GPR108 | 1.15 | 1.09 | 1.08 | 1.41* | 1.81* | 1.48* | 1.38 |
| Keratinocyte proline-rich protein | KPRP | 0.80 | 0.87 | 1.38 | 1.41 | 1.80* | 0.78 | 1.45 |
| Signal peptide peptidase-like 3 | SPPL3 | 1.03 | 0.85 | 1.23 | 0.96 | 1.79 | 1.17 | 1.29 |
| F-box only protein 50 | NCCRP1 | 1.73* | 2.05* | 3.97* | 0.77 | 1.78* | 0.88 | 1.38 |
| Sphingosine-1-phosphate phosphatase 1 | SGPP1 | 1.40 | 1.19 | 1.07 | 1.20 | 1.75* | 1.45 | 1.05 |
| Keratin, type II cytoskeletal 80 | KRT80 | 2.20* | 1.11 | 1.12 | 0.99 | 1.72* | 0.99 | 1.07 |
| NADH-ubiquinone oxidoreductase chain 2 | MT-ND2 | 1.14 | 1.39 | 1.08 | 0.90 | 1.72* | 1.19 | 0.90 |
| Disheveled-associated activator of morphogenesis 1 | DAAM1 | 0.91 | 0.92 | 0.98 | 1.23 | 1.71 | 1.47 | 1.34 |
| Keratin, type I cytoskeletal 25 | KRT25 | 0.44* | 1.22 | 0.25* | 0.41* | 1.70* | 0.23* | 1.17 |
| Cellular retinoic acid-binding protein 2 | CRABP2 | 1.18 | 1.15 | 1.37* | 1.37* | 1.68* | 1.45 | 1.34 |
| Pseudouridylate synthase RPUSD4, mitochondrial | RPUSD4 | 0.89 | 0.95 | 0.90 | 0.83 | 1.66 | 0.63 | 0.78 |
| Immediate early response 3-interacting protein 1 | IER3IP1 | 0.61* | 1.26 | 1.09 | 1.05 | 1.64* | 1.16 | 0.89 |
| Vitamin K-dependent gamma-carboxylase | GGCX | 1.38* | 1.37* | 1.14 | 1.37* | 1.64* | 1.15 | 1.22 |
| Maltase-glucoamylase | MGAM | 1.32* | 3.05* | 1.31* | 1.32* | 1.62* | 1.41* | 0.94 |
| Protein MMP24OS | MMP24OS | 1.36 | 1.67* | 1.29 | 1.11 | 1.61 | 1.43 | 1.33 |
| Phosphatidylserine synthase 2 | PTDSS2 | 1.40* | 1.46* | 1.39* | 1.33 | 1.61 | 1.49 | 1.37 |
| Protein OS-9 | OS9 | 1.38* | 1.38* | 1.13 | 1.15 | 1.61* | 1.43* | 1.26 |
| Nischarin | NISCH | 0.87 | 1.04 | 0.87 | 1.24 | 1.60 | 1.39 | 1.49 |

| | | | | | | | | |
|---|---|---|---|---|---|---|---|---|
| Calcium/calmodulin-dependent protein kinase type 1B | PNCK | 1.12 | 0.39* | 0.14* | 1.00 | 1.60 | 0.82 | 0.13* |
| Mediator of RNA polymerase II transcription subunit 24 | MED24 | 0.92 | 0.82 | 0.82 | 1.25 | 1.60 | 1.43 | 1.45 |
| Keratin, type II cytoskeletal 1 | KRT1 | 0.42* | 0.49* | 0.57* | 1.10 | 1.59* | 0.44* | 1.02 |
| Leucine-rich repeat-containing G-protein coupled receptor 4 | LGR4 | 0.77 | 0.86 | 0.82 | 0.89 | 1.59* | 1.25 | 1.33 |
| Dermokine | DMKN | 0.47* | 0.70 | 0.58* | 0.64* | 1.58 | 0.79 | 1.33 |
| Alpha-1,3-mannosyl-glycoprotein 4-beta-N-acetylglucosaminyltransferase B | MGAT4B | 1.01 | 1.01 | 0.99 | 1.09 | 1.58* | 1.47 | 1.37 |
| Ribonuclease P protein subunit p14 | RPP14 | 1.25 | 1.10 | 1.01 | 1.35 | 1.57 | 1.38 | 1.26 |
| Zinc transporter ZIP5 | SLC39A5 | 1.47* | 0.68 | 0.34* | 1.24 | 1.57 | 0.61 | 0.44* |
| Myelin expression factor 2 | MYEF2 | 0.70* | 1.05 | 1.05 | 0.66* | 1.57 | 1.32 | 1.36 |
| Anaphase-promoting complex subunit 5 | ANAPC5 | 0.39* | 0.38* | 0.39* | 0.53* | 1.56* | 1.01 | 1.02 |
| Golgi SNAP receptor complex member 2 | GOSR2 | 1.03 | 1.20 | 1.10 | 1.29 | 1.55 | 1.38 | 1.34 |
| Dual specificity tyrosine-phosphorylation-regulated kinase 1A | DYRK1A | 0.66* | 0.71* | 0.76* | 0.82 | 1.55* | 1.35 | 1.36 |
| 3 beta-hydroxysteroid dehydrogenase type 7 | HSD3B7 | 1.04 | 1.19 | 0.89 | 1.24 | 1.55 | 1.49 | 1.07 |
| Rhomboid-related protein 4 | RHBDD1 | 0.68 | 1.08 | 1.44 | 1.07 | 1.54 | 1.38 | 1.19 |
| Sucrase-isomaltase, intestinal | SI | 1.39* | 1.36* | 0.98 | 1.15 | 1.53* | 1.07 | 0.72* |
| Transcobalamin-2 | TCN2 | 1.28* | 1.12 | 1.03 | 1.08 | 1.53* | 1.20 | 0.84 |
| Prolyl 4-hydroxylase subunit alpha-1 | P4HA1 | 1.24* | 1.28* | 0.82* | 1.11 | 1.53* | 1.44* | 0.97 |
| Aminopeptidase RNPEPL1 | RNPEPL1 | 1.36 | 0.89 | 0.84 | 1.26 | 1.52 | 0.89 | 0.98 |
| Beta-1,4-N-acetylgalactosaminyltransferase 3 | B4GALNT3 | 1.15 | 1.32 | 1.14 | 1.37 | 1.52 | 1.32 | 1.27 |
| Transmembrane 4 L6 family member 20 | TM4SF20 | 1.61* | 0.93 | 0.65* | 0.63* | 1.51* | 0.97 | 0.50* |
| SPARC-related modular calcium-binding protein 1 | SMOC1 | 0.90 | 1.19 | 1.34* | 1.42* | 1.51 | 1.47 | 1.46 |
| Galectin-3-binding protein | LGALS3BP | 1.19 | 1.21 | 0.91 | 1.29* | 1.51* | 1.16 | 1.08 |
| Dipeptidyl peptidase 4 | DPP4 | 1.02 | 1.06 | 0.91 | 1.22* | 1.51* | 1.32* | 1.17 |
| Proline-rich protein 9 | PRR9 | 1.25 | 15.97* | 11.14* | 0.74 | 1.51 | 0.43* | 0.33* |
| SCAN domain-containing protein 3 | SCAND3 | 1.06 | 0.93 | 1.12 | 1.20 | 1.51 | 1.44 | 1.37 |
| Pumilio homolog 2 | PUM2 | 1.00 | 1.01 | 0.78 | 1.00 | 1.51 | 1.05 | 1.00 |
| CSC1-like protein 2 | TMEM63B | 1.10 | 1.23 | 1.04 | 1.14 | 1.50 | 1.23 | 1.21 |

***D. Up-regulated proteins unique to Aquamin plus Mesalamine (AQ+MES) with LPS-Cytokines [69 proteins]***

| | | Interventions | | | | | | |
|---|---|---|---|---|---|---|---|---|
| | | Control | | | *With LPS & Cytokines* | | | |
| Proteins | Genes | AQ | AQ+MES | MES | *LPS-Cyto* | *AQ* | ***AQ+MES*** | *MES* |
| Dynein axonemal heavy chain 8 | DNAH8 | 0.65* | 0.96 | 0.58* | 0.19* | 0.36* | 2.06* | 0.37* |
| Proteasome subunit beta type-3 | PSMB3 | 1.04 | 1.36* | 0.83 | 1.48* | 1.15 | 2.03* | 1.45* |
| Immunoglobulin heavy variable 3-49 | IGHV3-49 | 0.94 | 2.96* | 25.72* | 0.91 | 1.41 | 2.01* | 0.78 |

| | | | | | | | | |
|---|---|---|---|---|---|---|---|---|
| NEDD4 family-interacting protein 1 | NDFIP1 | 0.94 | 1.39 | 0.98 | 1.05 | 1.44 | 1.98* | 0.90 |
| Proteasome subunit alpha type-7 | PSMA7 | 1.12 | 1.30* | 1.38* | 1.33* | 1.25 | 1.96* | 1.35* |
| Dixin | DIXDC1 | 1.10 | 1.59* | 1.27 | 1.43 | 0.87 | 1.95* | 1.47 |
| Alpha-tocopherol transfer protein | TTPA | 0.89 | 1.02 | 1.13 | 1.20 | 1.39 | 1.93* | 1.21 |
| 55 kDa erythrocyte membrane protein | MPP1 | 1.16 | 2.80* | 1.47* | 1.10 | 1.36 | 1.92* | 1.21 |
| Proteasome subunit alpha type-4 | PSMA4 | 1.07 | 1.25* | 1.27* | 1.30* | 1.14 | 1.92* | 1.32* |
| Proteasome subunit alpha type-2 | PSMA2 | 1.10 | 1.16 | 1.12 | 1.40* | 1.18 | 1.92* | 1.37* |
| Proteasome subunit beta type-2 | PSMB2 | 1.36* | 1.44* | 1.27* | 1.28 | 1.23 | 1.90* | 1.42* |
| V-type proton ATPase 116 kDa subunit a 2 | ATP6V0A2 | 0.97 | 1.32 | 1.43 | 1.11 | 1.37 | 1.88* | 1.45 |
| Trehalase | TREH | 1.60* | 2.35* | 1.52* | 0.90 | 1.42* | 1.88* | 1.13 |
| Phosphatidylethanolamine N-methyltransferase | PEMT | 1.16 | 1.56 | 1.27 | 1.00 | 1.29 | 1.88* | 1.29 |
| Inositol-trisphosphate 3-kinase C | ITPKC | 1.17 | 2.04* | 1.95* | 1.21 | 1.37 | 1.86* | 1.46 |
| Acyl-CoA:lysophosphatidylglycerol acyltransferase 1 | LPGAT1 | 1.10 | 1.50* | 1.18 | 1.37* | 1.49* | 1.82* | 1.33 |
| Proteasome subunit beta type-4 | PSMB4 | 1.07 | 1.17 | 1.33* | 1.33* | 1.24 | 1.80* | 1.33* |
| Enhancer of filamentation 1 | NEDD9 | 1.46* | 1.77* | 1.30 | 0.93 | 1.23 | 1.75* | 0.91 |
| 7-methylguanosine phosphate-specific 5'-nucleotidase | NT5C3B | 1.45* | 1.37 | 1.15 | 1.06 | 0.83 | 1.75 | 0.44* |
| WD repeat and SOCS box-containing protein 2 | WSB2 | 1.15 | 1.43 | 1.24 | 1.40 | 1.21 | 1.73 | 1.48 |
| Proteasome subunit alpha type-3 | PSMA3 | 1.10 | 1.20 | 1.15* | 1.20 | 1.04 | 1.72* | 1.22 |
| E3 ubiquitin-protein ligase TRIM31 | TRIM31 | 0.95 | 1.22 | 1.36 | 1.26 | 0.88 | 1.72 | 1.39 |
| Proteasome subunit alpha type-1 | PSMA1 | 1.10 | 1.22 | 1.30* | 1.35* | 1.16 | 1.72* | 1.30* |
| Proteasome subunit alpha type-6 | PSMA6 | 1.03 | 1.12 | 1.33* | 1.41* | 1.09 | 1.70* | 1.26* |
| Tigger transposable element-derived protein 3 | TIGD3 | 0.93 | 1.74* | 0.94 | 1.15 | 1.30 | 1.69 | 1.19 |
| Mediator of RNA polymerase II transcription subunit 11 | MED11 | 1.14 | 1.04 | 1.11 | 0.99 | 1.25 | 1.69 | 0.78 |
| Multiple coagulation factor deficiency protein 2 | MCFD2 | 1.48* | 1.46* | 1.12 | 0.99 | 1.45 | 1.68* | 1.23 |
| Interferon regulatory factor 1 | IRF1 | 0.85 | 1.36 | 1.21 | 0.90 | 0.93 | 1.68 | 1.24 |
| Reticulophagy regulator 3 | RETREG3 | 0.98 | 0.97 | 1.05 | 1.04 | 1.35 | 1.68* | 1.18 |
| Immunoglobulin heavy constant gamma 4 | IGHG4 | 0.64* | 1.95* | 32.94* | 0.66* | 0.72 | 1.67* | 0.96 |
| Ceramide glucosyltransferase | UGCG | 1.26 | 1.37 | 1.32 | 1.24 | 1.11 | 1.67 | 1.22 |
| Hyaluronan-binding protein 2 | HABP2 | 0.99 | 1.28 | 1.36 | 0.93 | 1.22 | 1.66 | 1.45 |
| Lactadherin | MFGE8 | 1.13 | 1.35* | 1.35* | 1.26 | 1.45* | 1.66* | 1.32 |
| Dual oxidase maturation factor 2 | DUOXA2 | 1.45* | 1.75* | 1.60* | 1.11 | 1.43 | 1.63* | 1.04 |
| Proteasome subunit beta type-1 | PSMB1 | 1.20 | 1.23 | 1.18 | 1.22 | 1.03 | 1.63* | 1.14 |
| Solute carrier family 66 member 3 | SLC66A3 | 1.36 | 2.04* | 1.50* | 0.98 | 1.35 | 1.63 | 1.02 |
| E3 ubiquitin ligase RNF121 | RNF121 | 1.22 | 1.65* | 1.28 | 1.18 | 1.43 | 1.63 | 1.17 |
| Lysophospholipid acyltransferase 1 | MBOAT1 | 1.35 | 1.09 | 1.16 | 1.16 | 1.22 | 1.62 | 1.21 |
| Prenylated Rab acceptor protein 1 | RABAC1 | 0.98 | 1.19 | 1.32 | 1.22 | 1.17 | 1.62 | 1.41 |
| Alkaline phosphatase, placental type | ALPP | 0.98 | 0.67* | 0.93 | 0.51* | 1.19 | 1.61 | 1.36 |

| | | | | | | | | |
|---|---|---|---|---|---|---|---|---|
| Rap guanine nucleotide exchange factor 2 | RAPGEF2 | 1.37 | 1.49 | 0.93 | 1.01 | 1.40 | 1.61 | 0.84 |
| Interferon-stimulated gene 20 kDa protein | ISG20 | 1.14 | 1.25 | 1.35* | 1.12 | 1.39 | 1.60* | 1.46* |
| Toll-like receptor 3 | TLR3 | 1.17 | 1.23 | 0.89 | 1.46* | 1.46* | 1.60* | 1.24 |
| Proteasome subunit alpha type-5 | PSMA5 | 0.97 | 1.09 | 1.21* | 1.23* | 1.09 | 1.60* | 1.16 |
| High affinity copper uptake protein 1 | SLC31A1 | 1.36* | 1.51* | 1.17 | 1.17 | 1.11 | 1.59* | 1.16 |
| Protein RFT1 homolog | RFT1 | 1.24 | 1.24 | 1.18 | 1.31 | 1.43 | 1.58 | 1.48 |
| Peroxisomal membrane protein PEX13 | PEX13 | 1.03 | 1.23 | 1.21 | 1.15 | 1.27 | 1.57 | 1.42 |
| 2'-5'-oligoadenylate synthase 1 | OAS1 | 1.13 | 1.17 | 0.95 | 1.43* | 1.33 | 1.57* | 1.27 |
| Transmembrane channel-like protein 5 | TMC5 | 1.15 | 1.31 | 1.32 | 1.32 | 1.47 | 1.57 | 1.31 |
| Nicotinamide phosphoribosyltransferase | NAMPT | 1.04 | 1.10 | 1.09 | 1.39* | 1.41* | 1.56* | 1.39* |
| Ectonucleotide pyrophosphatase/phosphodiesterase family member 1 | ENPP1 | 0.97 | 1.08 | 1.27 | 1.06 | 1.35 | 1.56 | 1.27 |
| Nuclear pore complex protein Nup160 | NUP160 | 0.94 | 1.01 | 0.89 | 1.13 | 1.30 | 1.56 | 1.27 |
| Alanine aminotransferase 2 | GPT2 | 1.19 | 1.45 | 1.18 | 1.43 | 1.40 | 1.56 | 1.21 |
| Alsin | ALS2 | 1.51* | 1.57 | 1.67* | 1.30 | 1.37 | 1.56 | 1.50 |
| Elongation of very long chain fatty acids protein 5 | ELOVL5 | 1.28 | 1.30 | 1.17 | 1.23 | 1.30 | 1.56 | 1.28 |
| Cytokine receptor common subunit gamma | IL2RG | 1.27 | 1.67* | 1.27 | 1.28 | 1.27 | 1.55 | 0.97 |
| Lipase maturation factor 2 | LMF2 | 1.07 | 1.27 | 1.17 | 1.31 | 1.26 | 1.55 | 1.33 |
| Intestinal-type alkaline phosphatase | ALPI | 1.25 | 1.60* | 1.15 | 1.01 | 1.45 | 1.54 | 1.09 |
| Electrogenic sodium bicarbonate cotransporter 1 | SLC4A4 | 1.22 | 1.21 | 1.08 | 1.27 | 1.30 | 1.54 | 1.28 |
| ORM1-like protein 3 | ORMDL3 | 1.30* | 1.34 | 1.28 | 1.35 | 1.36 | 1.54 | 1.44 |
| H(+)/Cl(-) exchange transporter 7 | CLCN7 | 1.13 | 1.48* | 1.11 | 1.41 | 1.49 | 1.54 | 1.17 |
| Immunoglobulin heavy variable 3-7 | IGHV3-7 | 0.93 | 2.12* | 23.53* | 0.83 | 1.48 | 1.53 | 0.58 |
| Mucin-4 | MUC4 | 1.19 | 0.88 | 0.94 | 1.35 | 1.50 | 1.53 | 1.29 |
| Anion exchange protein 2 | SLC4A2 | 0.96 | 1.19 | 0.95 | 1.45* | 1.49 | 1.52* | 1.10 |
| Transmembrane protein 184B | TMEM184B | 0.97 | 0.97 | 1.02 | 1.10 | 1.29 | 1.52 | 1.19 |
| Torsin-1B | TOR1B | 1.03 | 1.29 | 1.11 | 1.18 | 1.41 | 1.51* | 1.42 |
| Translocating chain-associated membrane protein 1 | TRAM1 | 1.18 | 1.17 | 1.29 | 1.23 | 1.36 | 1.51* | 1.34 |
| Transmembrane 7 superfamily member 3 | TM7SF3 | 0.93 | 0.97 | 0.61* | 1.41* | 1.17 | 1.51 | 1.23 |
| Protein unc-93 homolog B1 | UNC93B1 | 1.21 | 1.29 | 1.14 | 1.29 | 1.42 | 1.50* | 1.22 |

***E. Up-regulated proteins unique to Mesalamine (MES) with LPS-Cytokines [84 proteins]***

| | | Interventions | | | | | | |
|---|---|---|---|---|---|---|---|---|
| | | Control | | | *With LPS & Cytokines* | | | |
| Proteins | Genes | AQ | AQ+MES | MES | *LPS-Cyto* | *AQ* | *AQ+MES* | ***MES*** |
| Keratin, type II cuticular Hb4 | KRT84 | 3.41* | 1.07 | 1.03 | 1.40 | 0.95 | 1.04 | 3.69* |
| Keratin, type I cuticular Ha4 | KRT34 | 0.84 | 63.49* | 2.24* | 0.74 | 1.05 | 0.91 | 3.16* |

| | | | | | | | | |
|---|---|---|---|---|---|---|---|---|
| EF-hand domain-containing protein D1 | EFHD1 | 0.66 | 43.85* | 1.91* | 0.69 | 0.69 | 0.84 | 3.11* |
| Eukaryotic peptide chain release factor GTP-binding subunit ERF3B | GSPT2 | 0.82 | 0.93 | 1.07 | 1.39 | 0.95 | 0.85 | 3.07* |
| Keratin, type I cuticular Ha1 | KRT31 | 0.68* | 49.23* | 2.04* | 0.65* | 0.92 | 0.80 | 2.96* |
| Keratin, type I cuticular Ha3-I | KRT33A | 0.87 | 24.84* | 1.71* | 0.94 | 0.91 | 1.29 | 2.44* |
| Dual specificity protein phosphatase 12 | DUSP12 | 0.92 | 0.94 | 0.94 | 1.02 | 1.45 | 1.43 | 2.31* |
| Thrombospondin-1 | THBS1 | 0.92 | 0.96 | 1.17 | 1.23 | 1.07 | 1.45* | 2.29* |
| Mitotic-spindle organizing protein 1 | MZT1 | 1.14 | 1.09 | 2.73* | 0.92 | 0.63 | 0.71 | 2.13* |
| tRNA methyltransferase 10 homolog A | TRMT10A | 0.87 | 1.03 | 1.66* | 0.99 | 1.01 | 1.30 | 2.12* |
| Spliceosome-associated protein CWC27 homolog | CWC27 | 2.02* | 1.81* | 1.94* | 1.33 | 0.51 | 0.99 | 2.11* |
| Retrotransposon-derived protein PEG10 | PEG10 | 0.68 | 0.94 | 1.32 | 1.06 | 1.31 | 1.39 | 2.08* |
| Retinoic acid receptor responder protein 2 | RARRES2 | 0.54* | 1.61* | 2.51* | 0.66 | 0.68 | 1.46 | 2.08* |
| Cell migration-inducing and hyaluronan-binding protein | CEMIP | 0.69* | 0.80 | 0.90 | 1.04 | 1.29 | 1.30 | 2.08* |
| Nucleoporin Nup43 | NUP43 | 1.06 | 1.34 | 1.38* | 1.45* | 1.26 | 1.43 | 2.05* |
| Fibronectin | FN1 | 0.81 | 1.09 | 1.21 | 1.17 | 0.99 | 1.39 | 1.99* |
| Serine protease HTRA1 | HTRA1 | 0.84 | 1.16 | 1.55* | 0.94 | 1.32 | 1.42 | 1.99* |
| Lysozyme g-like protein 2 | LYG2 | 0.84 | 31.25* | 1.48 | 0.85 | 1.13 | 1.33 | 1.98* |
| Vimentin | VIM | 0.73 | 1.24 | 1.17 | 1.24 | 1.45 | 1.29 | 1.96* |
| Thymosin beta-4 | TMSB4X | 0.55* | 0.94 | 1.71* | 1.20 | 1.01 | 1.17 | 1.93* |
| Collagen alpha-3(VI) chain | COL6A3 | 0.29* | 0.50* | 1.58* | 1.31 | 0.81 | 1.25 | 1.92* |
| Ferroxidase HEPHL1 | HEPHL1 | 0.77 | 61.07* | 3.28* | 0.93 | 1.02 | 0.31* | 1.91 |
| Ribosomal protein eL22-like | RPL22L1 | 0.45* | 0.89 | 1.44 | 0.86 | 0.54 | 1.05 | 1.90* |
| Histone H2B type 3-B | H2BC26 | 0.72 | 1.01 | 1.20 | 1.26 | 1.42 | 1.21 | 1.87* |
| High mobility group protein HMG-I/HMG-Y | HMGA1 | 0.50* | 0.61* | 1.13 | 1.38* | 1.08 | 1.34 | 1.87* |
| Pregnancy zone protein | PZP | 0.57* | 1.21 | 1.00 | 1.42 | 0.69 | 1.38 | 1.86 |
| Exostosin-2 | EXT2 | 1.09 | 1.51* | 1.42* | 1.25 | 1.38 | 1.43 | 1.85* |
| Beta-1,4-glucuronyltransferase 1 | B4GAT1 | 0.84 | 1.35 | 1.30 | 1.08 | 1.38 | 1.22 | 1.79 |
| Syntaxin-binding protein 1 | STXBP1 | 0.71 | 1.08 | 1.38 | 0.90 | 1.16 | 1.49 | 1.77* |
| Tripartite motif-containing protein 14 | TRIM14 | 0.85 | 1.01 | 0.96 | 1.21 | 1.16 | 1.07 | 1.77* |
| Kynureninase | KYNU | 0.82 | 1.00 | 1.61* | 0.52* | 0.92 | 1.21 | 1.74* |
| Testis-expressed protein 10 | TEX10 | 0.89 | 1.21 | 0.82 | 1.30 | 1.49 | 1.41 | 1.74 |
| Nephronectin | NPNT | 0.57* | 0.57* | 0.73 | 0.71 | 1.35 | 1.21 | 1.73 |
| Ribonucleoprotein PTB-binding 2 | RAVER2 | 0.86 | 1.47 | 1.25 | 0.59* | 0.57 | 0.57 | 1.71 |
| ICOS ligand | ICOSLG | 1.00 | 1.25 | 1.37 | 1.26 | 1.24 | 1.50 | 1.70 |
| Iron-sulfur cluster assembly 1 homolog, mitochondrial | ISCA1 | 0.81 | 1.20 | 1.48* | 0.84 | 0.99 | 1.35 | 1.68* |
| Palmitoyltransferase ZDHHC20 | ZDHHC20 | 1.12 | 1.18 | 1.15 | 1.29 | 1.42 | 1.35 | 1.68* |
| 4-hydroxyphenylpyruvate dioxygenase-like protein | HPDL | 1.20 | 1.03 | 0.99 | 1.34 | 1.30 | 1.29 | 1.68 |
| C4b-binding protein alpha chain | C4BPA | 0.48* | 1.00 | 1.41 | 1.50* | 1.06 | 1.25 | 1.68 |

| | | | | | | | | |
|---|---|---|---|---|---|---|---|---|
| Cohesin subunit SA-1 | STAG1 | 0.98 | 1.01 | 0.97 | 1.44 | 1.28 | 1.35 | 1.67 |
| Arylacetamide deacetylase | AADAC | 1.04 | 1.62 | 1.20 | 1.20 | 1.10 | 1.11 | 1.66 |
| Dermcidin | DCD | 0.64* | 0.68* | 0.85 | 0.99 | 1.23 | 0.95 | 1.65* |
| Peroxidasin homolog | PXDN | 1.04 | 1.13 | 1.31 | 1.18 | 1.11 | 1.39 | 1.65* |
| Neurogenic locus notch homolog protein 2 | NOTCH2 | 0.99 | 1.26 | 1.38 | 1.49 | 1.28 | 1.28 | 1.65 |
| Transcription elongation factor SPT4 | SUPT4H1 | 1.14 | 1.63* | 0.98 | 1.44 | 1.15 | 1.33 | 1.64 |
| Proprotein convertase subtilisin/kexin type 5 | PCSK5 | 0.95 | 1.00 | 0.81 | 1.15 | 1.02 | 0.84 | 1.63 |
| Serine/threonine-protein kinase 3 | STK3 | 0.70 | 0.78 | 1.01 | 1.18 | 1.09 | 0.86 | 1.62 |
| Nurim | NRM | 1.27 | 0.90 | 1.16 | 1.15 | 1.00 | 1.40 | 1.62 |
| FACT complex subunit SSRP1 | SSRP1 | 0.93 | 0.95 | 1.07 | 1.26 | 1.01 | 1.03 | 1.59* |
| Caspase-7 | CASP7 | 1.04 | 1.26 | 1.14 | 1.46* | 1.44 | 1.46 | 1.59* |
| Sideroflexin-4 | SFXN4 | 0.93 | 1.26 | 1.05 | 1.30* | 1.46* | 1.48* | 1.59* |
| Urokinase-type plasminogen activator | PLAU | 0.96 | 1.36 | 1.17 | 1.36 | 1.32 | 1.42 | 1.58 |
| N-acetylglucosamine-1-phosphodiester alpha-N-acetylglucosaminidase | NAGPA | 0.82 | 1.28 | 1.80* | 0.75 | 0.93 | 1.45 | 1.58 |
| Myosin-10 | MYH10 | 0.88 | 0.74* | 0.95 | 1.01 | 0.91 | 1.19 | 1.58* |
| AT-rich interactive domain-containing protein 1A | ARID1A | 0.99 | 1.20 | 1.04 | 1.28 | 1.26 | 1.36 | 1.58 |
| Vesicle transport protein SEC20 | BNIP1 | 1.14 | 1.27 | 1.23 | 1.08 | 1.14 | 1.18 | 1.58 |
| Trafficking protein particle complex subunit 2-like protein | TRAPPC2L | 0.96 | 1.06 | 1.01 | 0.93 | 1.05 | 1.32 | 1.57 |
| Sodium-dependent lysophosphatidylcholine symporter 1 | MFSD2A | 1.38 | 1.71* | 1.13 | 1.34 | 0.87 | 1.25 | 1.57 |
| Mitochondrial import inner membrane translocase subunit Tim10 B | TIMM10B | 1.17 | 1.25 | 0.96 | 1.50* | 1.33 | 1.43 | 1.56 |
| Insulin-like growth factor-binding protein 4 | IGFBP4 | 0.88 | 1.45* | 1.57* | 0.83 | 0.89 | 1.38 | 1.56 |
| Glutathione peroxidase 2 | GPX2 | 1.00 | 1.03 | 1.06 | 1.29 | 1.25 | 1.30 | 1.55* |
| ADP-ribosylation factor GTPase-activating protein 1 | ARFGAP1 | 0.99 | 1.39* | 1.38* | 1.49* | 1.20 | 1.21 | 1.55* |
| Golgin-45 | BLZF1 | 0.88 | 1.13 | 1.10 | 1.05 | 1.08 | 0.98 | 1.54 |
| 5'-nucleotidase domain-containing protein 2 | NT5DC2 | 0.93 | 1.08 | 1.18 | 0.94 | 1.22 | 1.20 | 1.54* |
| Ribosome production factor 1 | RPF1 | 0.83 | 0.67* | 0.93 | 1.42* | 1.40 | 1.26 | 1.54 |
| Inter-alpha-trypsin inhibitor heavy chain H1 | ITIH1 | 0.48* | 0.96 | 1.54* | 0.48* | 0.48* | 1.13 | 1.53 |
| Alpha-methylacyl-CoA racemase | AMACR | 1.35 | 1.27 | 1.07 | 1.26 | 1.21 | 1.18 | 1.53 |
| Histone H1.2 | H1-2 | 0.96 | 0.74 | 1.15 | 0.79 | 0.98 | 0.88 | 1.53* |
| Tumor necrosis factor receptor superfamily member 12A | TNFRSF12A | 0.88 | 1.46 | 1.75* | 1.47 | 1.35 | 1.30 | 1.52 |
| Multiple inositol polyphosphate phosphatase 1 | MINPP1 | 1.08 | 1.23 | 1.32* | 1.49* | 1.06 | 1.23 | 1.52 |
| Homeobox protein CDX-2 | CDX2 | 1.29 | 1.40 | 1.36 | 1.43 | 0.92 | 1.24 | 1.52 |
| ATP-binding cassette sub-family B member 10, mitochondrial | ABCB10 | 1.07 | 1.30 | 1.13 | 1.34 | 1.48 | 1.23 | 1.52 |
| Syndecan-4 | SDC4 | 1.04 | 0.89 | 1.12 | 1.48* | 1.49* | 1.32 | 1.52* |
| Non-histone chromosomal protein HMG-17 | HMGN2 | 0.56* | 1.04 | 1.61* | 0.87 | 0.80 | 1.21 | 1.52 |

| | | | | | | | | |
|---|---|---|---|---|---|---|---|---|
| mRNA-decapping enzyme 1B | DCP1B | 1.26 | 2.01* | 1.92* | 0.58* | 1.35 | 0.76 | 1.52 |
| ATP-dependent DNA/RNA helicase DHX36 | DHX36 | 0.77* | 0.80 | 0.81 | 0.93 | 1.17 | 1.34 | 1.52* |
| Tsukushi | TSKU | 0.99 | 1.13 | 1.15 | 1.08 | 1.43 | 1.47 | 1.51 |
| Apolipoprotein B-100 | APOB | 0.77 | 1.20 | 1.37 | 1.26 | 1.18 | 1.26 | 1.51 |
| CDK-activating kinase assembly factor MAT1 | MNAT1 | 0.53* | 0.93 | 1.05 | 0.79 | 0.52 | 0.89 | 1.51 |
| UDP-glucuronic acid decarboxylase 1 | UXS1 | 1.12 | 1.22 | 1.09 | 1.06 | 1.42 | 1.45 | 1.51 |
| GRIP1-associated protein 1 | GRIPAP1 | 1.01 | 0.97 | 1.28 | 1.16 | 1.12 | 1.26 | 1.51 |
| Serpin H1 | SERPINH1 | 1.08 | 1.21 | 1.03 | 1.15 | 1.29 | 1.30* | 1.51* |
| Sodium-dependent multivitamin transporter | SLC5A6 | 1.09 | 1.02 | 1.02 | 1.41* | 1.39 | 1.46 | 1.50* |
| Cell division cycle protein 27 homolog | CDC27 | 1.11 | 0.99 | 1.00 | 0.98 | 0.96 | 1.19 | 1.50 |

***F. Common up-regulated proteins between LPS-Cytokines alone and with Aquamin [17 proteins]***

| | | Interventions | | | | | | |
|---|---|---|---|---|---|---|---|---|
| | | Control | | | *With LPS & Cytokines* | | | |
| Proteins | Genes | AQ | AQ+MES | MES | ***LPS-Cyto*** | ***AQ*** | *AQ+MES* | *MES* |
| Insulin-like growth factor-binding protein 1 | IGFBP1 | 1.38 | 0.82 | 0.97 | 3.49* | 1.56 | 1.39 | 1.48 |
| Interferon-related developmental regulator 1 | IFRD1 | 2.60* | 0.80 | 1.96* | 3.12* | 1.60 | 1.01 | 0.77 |
| Glutathione S-transferase A2 | GSTA2 | 1.36* | 0.40* | 0.10* | 2.33* | 1.56* | 0.19* | 0.50* |
| Proline-rich acidic protein 1 | PRAP1 | 1.21 | 1.06 | 0.84 | 2.23* | 2.24* | 1.21 | 1.08 |
| Bromodomain-containing protein 3 | BRD3 | 1.18 | 1.44 | 1.36 | 1.92* | 1.62 | 1.09 | 1.42 |
| Formin-binding protein 4 | FNBP4 | 0.67 | 1.03 | 0.79 | 1.91* | 1.61 | 1.38 | 1.42 |
| Male-enhanced antigen 1 | MEA1 | 1.28 | 1.17 | 1.37 | 1.81* | 1.59 | 1.28 | 1.50 |
| Loricrin | LORICRIN | 0.48* | 0.44* | 0.64* | 1.79* | 1.63* | 0.30* | 1.10 |
| Sodium-dependent neutral amino acid transporter B(0)AT1 | SLC6A19 | 1.63* | 2.12* | 0.93 | 1.71* | 1.74* | 1.12 | 0.47* |
| Dynein axonemal light chain 1 | DNAL1 | 0.94 | 1.27 | 1.25 | 1.69* | 2.16* | 1.29 | 1.48 |
| Myeloid-associated differentiation marker | MYADM | 1.30 | 1.03 | 1.17 | 1.67* | 1.60 | 1.06 | 1.35 |
| Deleted in malignant brain tumors 1 protein | DMBT1 | 2.24* | 0.89 | 1.16* | 1.67* | 1.57* | 0.60* | 0.73* |
| Matrilysin | MMP7 | 1.52* | 1.02 | 1.05 | 1.66* | 1.74* | 1.24 | 1.42* |
| Cytochrome P450 2C19 | CYP2C19 | 2.12* | 1.15 | 0.61* | 1.64* | 1.98* | 0.88 | 0.62 |
| EKC/KEOPS complex subunit TPRKB | TPRKB | 1.13 | 1.46 | 1.16 | 1.53* | 1.62 | 1.15 | 1.49 |
| F-box only protein 28 | FBXO28 | 0.94 | 1.19 | 1.24 | 1.52 | 1.68 | 1.38 | 1.44 |
| 2-amino-3-carboxymuconate-6-semialdehyde decarboxylase | ACMSD | 1.49* | 1.40 | 0.84 | 1.51* | 1.53 | 1.09 | 1.10 |

***G. Common up-regulated proteins between LPS-Cytokines alone and with Aquamin plus Mesalamine [2 proteins]***

Interventions

| | | Control | | | *With LPS & Cytokines* | | | |
|---|---|---|---|---|---|---|---|---|
| Proteins | Genes | AQ | AQ+MES | MES | ***LPS-Cyto*** | *AQ* | ***AQ+MES*** | *MES* |
| Threonylcarbamoyl-AMP synthase | YRDC | 1.20 | 1.69* | 1.25 | 1.89* | 0.94 | 1.91* | 1.10 |
| Zinc transporter ZIP10 | SLC39A10 | 1.20 | 1.29 | 1.24 | 1.67* | 1.02 | 1.68 | 1.37 |

***H. Common up-regulated proteins between LPS-Cytokines alone and with Mesalamine [18 proteins]***

| | | Interventions | | | | | | |
|---|---|---|---|---|---|---|---|---|
| | | Control | | | *With LPS & Cytokines* | | | |
| Proteins | Genes | AQ | AQ+MES | MES | ***LPS-Cyto*** | *AQ* | *AQ+MES* | ***MES*** |
| Integrin alpha-7 | ITGA7 | 0.48* | 0.78 | 0.74 | 5.21* | 0.84 | 0.49* | 1.76 |
| Non-histone chromosomal protein HMG-14 | HMGN1 | 1.16 | 0.60* | 0.78 | 3.08* | 1.07 | 1.10 | 2.25* |
| Protein-glutamine gamma-glutamyltransferase E | TGM3 | 1.03 | 1.11 | 0.97 | 2.43* | 1.47* | 1.08 | 1.56* |
| Transcription factor 20 | TCF20 | 0.66* | 0.99 | 1.00 | 2.22* | 0.80 | 0.51 | 1.59 |
| Fibrinogen alpha chain | FGA | 0.63* | 0.68 | 2.55* | 2.12* | 0.76 | 1.05 | 2.43* |
| Arrestin-C | ARR3 | 0.73 | 1.04 | 1.01 | 1.99* | 1.12 | 0.86 | 1.82 |
| Bleomycin hydrolase | BLMH | 1.70* | 1.51* | 1.70* | 1.98* | 1.46 | 1.45 | 1.59 |
| Zinc finger protein 284 | ZNF284 | 1.90* | 1.45 | 1.70* | 1.88* | 0.68 | 0.99 | 2.29* |
| Protein FAM193A | FAM193A | 0.75 | 0.87 | 1.40 | 1.83* | 0.96 | 1.32 | 1.98* |
| Transmembrane protein 179B | TMEM179B | 1.32 | 1.24 | 1.18 | 1.82* | 1.44 | 1.48 | 1.67* |
| Ankyrin repeat domain-containing protein 27 | ANKRD27 | 1.10 | 1.61* | 1.51* | 1.81* | 1.46* | 1.06 | 1.92* |
| ABC-type oligopeptide transporter ABCB9 | ABCB9 | 0.40* | 0.67 | 1.05 | 1.77* | 0.59 | 0.93 | 2.42* |
| Ferritin heavy chain | FTH1 | 0.99 | 0.81 | 1.35* | 1.72* | 1.29 | 1.24 | 1.97* |
| tRNA (guanine-N(7)-)-methyltransferase | METTL1 | 1.03 | 1.27 | 1.33 | 1.70* | 1.32 | 1.25 | 1.80* |
| Immortalization up-regulated protein | IMUP | 0.66* | 1.26 | 1.81* | 1.60* | 1.16 | 1.10 | 1.75* |
| Serine/threonine-protein kinase 4 | STK4 | 0.98 | 1.00 | 1.13 | 1.56* | 1.42 | 1.45 | 1.60* |
| Protein S100-A9 | S100A9 | 0.60* | 1.51* | 1.41* | 1.53* | 1.35 | 1.39 | 2.46* |
| Epsin-2 | EPN2 | 1.54* | 1.34* | 1.44* | 1.52* | 1.37 | 1.38 | 1.54* |

***I. Common up-regulated proteins among LPS-Cytokines alone, with Aquamin and with Aquamin plus Mesalamine [9 proteins]***

| | | Interventions | | | | | | |
|---|---|---|---|---|---|---|---|---|
| | | Control | | | *With LPS & Cytokines* | | | |
| Proteins | Genes | AQ | AQ+MES | MES | ***LPS-Cyto*** | ***AQ*** | ***AQ+MES*** | *MES* |
| Peroxisome assembly protein 12 | PEX12 | 1.11 | 1.29 | 1.00 | 2.05* | 2.02* | 1.96* | 1.41 |
| Complement component C8 gamma chain | C8G | 1.96* | 2.13* | 4.51* | 1.81* | 2.79* | 1.96* | 1.26 |
| Apolipoprotein L6 | APOL6 | 1.33 | 1.18 | 0.80 | 1.70* | 2.00* | 2.15* | 1.29 |
| Gamma-interferon-inducible protein 16 | IFI16 | 1.10 | 1.19 | 1.04 | 1.62* | 1.62 | 1.69* | 1.36 |
| Ferredoxin-2, mitochondrial | FDX2 | 1.78* | 1.45 | 1.55* | 1.59* | 2.48* | 1.91* | 1.27 |

| | | | | | | | | |
|---|---|---|---|---|---|---|---|---|
| NK-tumor recognition protein | NKTR | 1.53* | 1.80* | 1.57 | 1.59 | 1.50 | 1.67 | 1.19 |
| Serine protease inhibitor Kazal-type 1 | SPINK1 | 0.67 | 1.40 | 1.08 | 1.56* | 1.71* | 1.57 | 1.39 |
| Armadillo repeat-containing protein 2 | ARMC2 | 1.29 | 1.30 | 1.14 | 1.55* | 1.88* | 1.86* | 1.46 |
| Ileal sodium/bile acid cotransporter | SLC10A2 | 1.33* | 2.98* | 1.64* | 1.53* | 1.67* | 2.16* | 1.42 |

***J. Common up-regulated proteins among LPS-Cytokines alone, with Aquamin and with Mesalamine [6 proteins]***

| | | Interventions | | | | | | |
|---|---|---|---|---|---|---|---|---|
| | | Control | | | *With LPS & Cytokines* | | | |
| Proteins | Genes | AQ | AQ+MES | MES | ***LPS-Cyto*** | ***AQ*** | *AQ+MES* | ***MES*** |
| Apolipoprotein D | APOD | 1.10 | 2.13* | 1.32 | 4.26* | 2.98* | 1.00 | 2.56* |
| Prolactin-inducible protein | PIP | 0.90 | 1.33* | 1.20 | 3.19* | 2.97* | 0.93 | 2.67* |
| Secreted Ly-6/uPAR domain-containing protein 2 | SLURP2 | 0.62* | 0.79 | 0.42* | 2.19* | 4.45* | 1.19 | 3.77* |
| Electron transfer flavoprotein regulatory factor 1 | ETFRF1 | 1.41* | 1.71* | 1.48* | 1.68* | 1.53 | 1.47 | 1.73* |
| U3 small nucleolar RNA-associated protein 15 homolog | UTP15 | 0.85 | 0.85 | 0.86 | 1.57* | 1.57* | 1.37 | 1.90* |
| Prostatic acid phosphatase | ACP3 | 1.83* | 8.61* | 2.97* | 1.52* | 2.09* | 1.08 | 1.62 |

***K. Common up-regulated proteins among LPS-Cytokines alone, with Aquamin plus Mesalamine and with Mesalamine [29 proteins]***

| | | Interventions | | | | | | |
|---|---|---|---|---|---|---|---|---|
| | | Control | | | *With LPS & Cytokines* | | | |
| Proteins | Genes | AQ | AQ+MES | MES | ***LPS-Cyto*** | *AQ* | ***AQ+MES*** | ***MES*** |
| Collagen alpha-6(IV) chain | COL4A6 | 0.76 | 1.01 | 1.42 | 2.54* | 0.98 | 1.86* | 2.68* |
| Nuclear receptor coactivator 6 | NCOA6 | 2.02* | 3.40* | 3.87* | 2.35* | 0.89 | 2.98* | 4.19* |
| CD166 antigen | ALCAM | 1.12 | 1.95* | 1.74* | 2.10* | 1.22 | 2.14* | 1.78* |
| Interferon-induced protein with tetratricopeptide repeats 5 | IFIT5 | 1.04 | 1.34 | 1.60* | 2.00* | 1.09 | 1.96* | 2.05* |
| Midkine | MDK | 0.80 | 1.07 | 1.17 | 1.97* | 1.39 | 1.68* | 1.69* |
| Ferritin light chain | FTL | 0.52* | 2.72* | 3.90* | 1.95* | 0.58 | 4.12* | 9.36* |
| Antithrombin-III | SERPINC1 | 0.69* | 1.19 | 1.55* | 1.84* | 1.44* | 1.87* | 2.27* |
| Alpha-2-HS-glycoprotein | AHSG | 0.70* | 1.45* | 1.73* | 1.82* | 1.47* | 2.35* | 2.40* |
| Bis(5'-adenosyl)-triphosphatase | FHIT | 1.09 | 1.58* | 1.32 | 1.78* | 1.32 | 1.52 | 1.77* |
| Mitoferrin-2 | SLC25A28 | 1.83* | 3.25* | 3.19* | 1.78* | 1.29 | 3.96* | 2.50* |
| Apolipoprotein L5 | APOL5 | 1.57* | 0.98 | 1.90* | 1.78* | 1.11 | 1.62 | 1.76 |
| N-myc-interactor | NMI | 1.08 | 1.13 | 1.26* | 1.73* | 1.34 | 1.52* | 1.63* |
| MARVEL domain-containing protein 3 | MARVELD3 | 1.31 | 1.82* | 1.63* | 1.72* | 0.97 | 1.89* | 1.56 |
| Kininogen-1 | KNG1 | 0.47* | 2.27* | 2.73* | 1.71* | 0.68 | 2.29* | 2.32* |
| Opioid growth factor receptor | OGFR | 1.32* | 2.03* | 1.72* | 1.66* | 1.39 | 1.91* | 2.03* |
| NEDD8 ultimate buster 1 | NUB1 | 0.87 | 1.39* | 1.34* | 1.66* | 0.92 | 1.60* | 1.55* |

| | | | | | | | | |
|---|---|---|---|---|---|---|---|---|
| Cilia- and flagella-associated protein 100 | CFAP100 | 0.89 | 1.31 | 1.78* | 1.65* | 1.48 | 2.37* | 2.52* |
| Gasdermin-B | GSDMB | 0.99 | 1.13 | 1.35* | 1.64* | 0.98 | 1.59 | 1.64* |
| Stomatin | STOM | 1.08 | 1.18 | 1.06 | 1.62* | 1.50 | 1.71* | 1.65* |
| Phospholipid scramblase 1 | PLSCR1 | 0.56* | 1.04 | 1.09 | 1.60* | 0.94 | 1.55 | 1.76* |
| Collagen alpha-1(II) chain | COL2A1 | 1.18 | 4.35* | 5.26* | 1.58* | 1.09 | 3.94* | 5.38* |
| Collagen alpha-1(XV) chain | COL15A1 | 0.83 | 1.81* | 1.43 | 1.57 | 1.31 | 1.62 | 2.31* |
| G patch domain-containing protein 8 | GPATCH8 | 0.78 | 0.72 | 1.11 | 1.55* | 1.25 | 1.53 | 1.63 |
| Putative sodium-coupled neutral amino acid transporter 10 | SLC38A10 | 1.01 | 1.36 | 1.38 | 1.55* | 1.41 | 1.86* | 1.55 |
| Protein C19orf12 | C19orf12 | 1.61* | 2.28* | 1.77* | 1.55 | 1.49 | 1.94* | 1.86 |
| Transcription factor ETV6 | ETV6 | 1.90* | 1.88* | 2.40* | 1.54* | 1.45 | 2.02* | 1.81 |
| Apolipoprotein A-IV | APOA4 | 0.95 | 3.94* | 8.02* | 1.53* | 1.25 | 2.60* | 3.78* |
| ATP-binding cassette sub-family C member 2 | ABCC2 | 1.50* | 2.42* | 1.43* | 1.53* | 1.48* | 1.89* | 1.60* |
| GATOR complex protein MIOS | MIOS | 1.18 | 1.06 | 1.20 | 1.50* | 1.47 | 1.61 | 2.16* |

***L. Common up-regulated proteins between LPS-Cytokines with Aquamin and with Aquamin+Mesalamine [50 proteins]***

| | | Interventions | | | | | | |
|---|---|---|---|---|---|---|---|---|
| | | Control | | | *With LPS & Cytokines* | | | |
| Proteins | Genes | AQ | AQ+MES | MES | *LPS-Cyto* | ***AQ*** | ***AQ+MES*** | *MES* |
| Cadherin-17 | CDH17 | 3.59* | 3.47* | 1.05 | 0.91 | 2.69* | 2.67* | 0.73* |
| Phosphatidylinositol 4-phosphate 5-kinase type-1 beta | PIP5K1B | 1.11 | 1.19 | 0.97 | 0.79 | 2.66* | 2.81* | 1.27 |
| Copine-8 | CPNE8 | 1.21* | 1.04 | 0.87 | 1.35* | 2.62* | 2.32* | 1.37* |
| Fibronectin type III and SPRY domain-containing protein 1 | FSD1 | 3.62* | 3.96* | 1.02 | 0.70 | 2.52* | 1.94* | 0.49* |
| Poly(ADP-ribose) glycohydrolase | PARG | 1.16 | 1.02 | 0.74 | 1.29 | 2.51* | 1.58 | 0.89 |
| Lipase member H | LIPH | 1.16 | 1.37 | 1.18 | 1.37 | 2.38* | 2.40* | 1.48 |
| Calcium/manganese antiporter SLC30A10 | SLC30A10 | 3.21* | 2.20* | 1.22 | 1.36 | 2.24* | 2.25* | 0.82 |
| Sodium/hydrogen exchanger 2 | SLC9A2 | 1.36* | 1.35 | 1.38* | 1.10 | 2.18* | 1.67 | 1.28 |
| Desmoglein-2 | DSG2 | 2.17* | 2.21* | 0.93 | 1.08 | 2.03* | 2.03* | 1.45* |
| Protein FAM3A | FAM3A | 1.18 | 1.25 | 1.22 | 1.39* | 2.02* | 1.88* | 1.48* |
| Protocadherin-1 | PCDH1 | 2.10* | 2.09* | 1.14 | 0.88 | 1.93* | 1.95* | 1.13 |
| Very low-density lipoprotein receptor | VLDLR | 1.58* | 1.36 | 1.44* | 1.24 | 1.93* | 1.56 | 1.43 |
| Sphingolipid delta(4)-desaturase DES1 | DEGS1 | 1.29 | 1.47 | 1.22 | 1.39 | 1.87* | 1.96* | 1.45 |
| Olfactory receptor 1M1 | OR1M1 | 1.48* | 1.95* | 1.32 | 1.06 | 1.87* | 2.10* | 1.19 |
| Copine-2 | CPNE2 | 1.17 | 1.35 | 1.18 | 1.26 | 1.86* | 1.71 | 1.26 |
| Xaa-Pro aminopeptidase 2 | XPNPEP2 | 2.00* | 2.97* | 1.79* | 1.17 | 1.85* | 1.67* | 1.38 |
| Synaptotagmin-7 | SYT7 | 0.99 | 1.23 | 1.00 | 0.99 | 1.84* | 1.91* | 0.91 |
| H(+)/Cl(-) exchange transporter 5 | CLCN5 | 1.32 | 1.34 | 1.07 | 1.43 | 1.83* | 1.83* | 1.19 |

| | | | | | | | | |
|---|---|---|---|---|---|---|---|---|
| Glutamine synthetase | GLUL | 1.31* | 1.26 | 1.08 | 1.40* | 1.83* | 1.78* | 1.38* |
| Pterin-4-alpha-carbinolamine dehydratase 2 | PCBD2 | 1.71* | 1.26 | 1.07 | 1.40 | 1.81* | 1.88* | 1.22 |
| Protein PALS2 | PALS2 | 0.98 | 0.52* | 0.71 | 0.52* | 1.81* | 1.54 | 1.29 |
| Ankyrin-2 | ANK2 | 1.33 | 1.30 | 1.15 | 1.25 | 1.79* | 1.60 | 1.09 |
| Melanotransferrin | MELTF | 1.46* | 1.86* | 1.26 | 1.24 | 1.78 | 1.93* | 1.35 |
| Integrin alpha-5 | ITGA5 | 1.11 | 1.46* | 1.46* | 1.08 | 1.78* | 1.74* | 1.18 |
| Galactosylgalactosylxylosylprotein 3-beta-glucuronosyltransferase 3 | B3GAT3 | 1.46* | 1.65* | 1.29 | 1.47* | 1.77 | 2.12* | 1.07 |
| Cytochrome c oxidase assembly protein COX18, mitochondrial | COX18 | 1.09 | 0.91 | 0.92 | 1.27 | 1.76* | 1.54 | 1.47 |
| Protein SERAC1 | SERAC1 | 1.48* | 1.37 | 1.03 | 1.09 | 1.72 | 1.53 | 1.18 |
| Solute carrier family 52, riboflavin transporter, member 3 | SLC52A3 | 1.21 | 1.54 | 1.45 | 1.29 | 1.72 | 1.52 | 1.49 |
| Neuropilin-2 | NRP2 | 1.63* | 1.43 | 1.45* | 1.26 | 1.69* | 1.56 | 1.33 |
| Heme transporter HRG1 | SLC48A1 | 1.02 | 1.24 | 0.93 | 1.23 | 1.68 | 1.75* | 1.35 |
| Elongation factor 1-alpha 2 | EEF1A2 | 0.84 | 1.52* | 1.19 | 1.02 | 1.66* | 1.71* | 1.43 |
| Protein CLN8 | CLN8 | 1.36 | 1.43 | 1.29 | 1.34 | 1.66 | 1.67 | 1.33 |
| Myosin light chain 6B | MYL6B | 0.56* | 0.71* | 0.75 | 0.80 | 1.66* | 1.68* | 1.15 |
| Signal peptide peptidase-like 2A | SPPL2A | 1.09 | 1.25 | 0.96 | 1.40 | 1.65* | 1.53 | 1.19 |
| CD82 antigen | CD82 | 0.79 | 0.90 | 1.01 | 1.06 | 1.64 | 1.59 | 1.20 |
| Osteopetrosis-associated transmembrane protein 1 | OSTM1 | 1.28 | 1.27 | 1.19 | 1.35 | 1.64 | 1.67 | 1.34 |
| Lathosterol oxidase | SC5D | 1.20 | 1.17 | 1.09 | 1.35 | 1.63 | 1.68 | 1.44 |
| Tetratricopeptide repeat protein 22 | TTC22 | 1.20 | 1.09 | 1.32 | 1.46 | 1.62 | 1.58 | 1.21 |
| Solute carrier family 53 member 1 | XPR1 | 1.58* | 1.51 | 1.38 | 1.17 | 1.61 | 1.70* | 1.32 |
| Sushi domain-containing protein 2 | SUSD2 | 1.30 | 2.95* | 1.70* | 1.09 | 1.57 | 1.60 | 0.92 |
| Glutaminyl-peptide cyclotransferase-like protein | QPCTL | 1.26 | 1.86* | 1.14 | 1.24 | 1.57 | 1.58 | 1.49 |
| UDP-GlcNAc:betaGal beta-1,3-N-acetylglucosaminyltransferase 7 | B3GNT7 | 1.09 | 1.78* | 1.50* | 0.94 | 1.56 | 1.64* | 1.41 |
| Proprotein convertase subtilisin/kexin type 9 | PCSK9 | 1.03 | 1.13 | 1.18 | 1.09 | 1.55 | 1.55 | 1.26 |
| Protein O-linked-mannose beta-1,4-N-acetylglucosaminyltransferase 2 | POMGNT2 | 1.04 | 1.53 | 1.41 | 1.30 | 1.55 | 1.88 | 1.04 |
| Cytochrome P450 4F11 | CYP4F11 | 0.97 | 1.71* | 1.13 | 1.31 | 1.55 | 1.55 | 1.32 |
| Arginase-2, mitochondrial | ARG2 | 1.67* | 1.74* | 1.25 | 1.44 | 1.55 | 1.51 | 0.93 |
| Integral membrane protein 2B | ITM2B | 1.06 | 1.19 | 1.07 | 1.35 | 1.53 | 1.61* | 1.40 |
| tRNA modification GTPase GTPBP3, mitochondrial | GTPBP3 | 0.71 | 1.16 | 1.14 | 1.12 | 1.53 | 1.55 | 0.65 |
| GPI inositol-deacylase | PGAP1 | 1.24 | 1.51* | 1.27 | 1.13 | 1.52 | 1.91* | 1.21 |
| p53 apoptosis effector related to PMP-22 | PERP | 1.05 | 1.22 | 1.25 | 0.82 | 1.51 | 2.65* | 1.46 |

***M. Common up-regulated proteins between LPS-Cytokines with Aquamin and with Mesalamine [50 proteins]***

| | | Interventions | | | | | | |
|---|---|---|---|---|---|---|---|---|
| | | Control | | | *With LPS & Cytokines* | | | |
| Proteins | Genes | AQ | AQ+MES | MES | *LPS-Cyto* | ***AQ*** | *AQ+MES* | ***MES*** |
| Keratin, type II cytoskeletal 4 | KRT4 | 3.27* | 1.56* | 9.03* | 0.70 | 10.27* | 1.13 | 6.70* |
| Cornifin-B | SPRR1B | 0.79 | 1.62* | 1.68* | 1.14 | 4.30* | 0.46* | 3.71* |
| Cystatin-A | CSTA | 1.20* | 1.45* | 1.11 | 1.25 | 3.86* | 0.98 | 3.06* |
| Cornulin | CRNN | 1.73* | 2.38* | 5.70* | 0.44* | 3.63* | 0.61 | 2.57* |
| Protein KPLCE | KPLCE | 1.16 | 0.88 | 0.98 | 0.61* | 3.48* | 0.40* | 1.62* |
| Secretoglobin family 1D member 2 | SCGB1D2 | 0.91 | 1.39 | 1.53* | 1.04 | 3.37* | 0.82 | 2.35* |
| Neutrophil elastase | ELANE | 0.97 | 1.85* | 0.83 | 1.30 | 3.28* | 0.92 | 2.27* |
| Serpin B12 | SERPINB12 | 0.96 | 1.21 | 2.32* | 1.07 | 3.22* | 0.93 | 2.21* |
| Protein-glutamine gamma-glutamyltransferase K | TGM1 | 1.13 | 1.39 | 1.71* | 0.66 | 3.12* | 0.49* | 2.44* |
| Serpin B3 | SERPINB3 | 0.72* | 1.14 | 1.82* | 1.19 | 2.93* | 0.78 | 1.96* |
| Plakophilin-1 | PKP1 | 0.84 | 3.19* | 2.07* | 0.84 | 2.73* | 0.56* | 2.07* |
| Cystatin-S | CST4 | 0.83 | 1.14 | 1.53* | 1.06 | 2.72* | 1.49 | 2.96* |
| Immunoglobulin heavy constant alpha 1 | IGHA1 | 0.68* | 1.29* | 3.28* | 1.11 | 2.69* | 1.30 | 2.32* |
| Keratin, type I cytoskeletal 23 | KRT23 | 2.31* | 1.81* | 6.31* | 0.44* | 2.65* | 0.70 | 2.04* |
| Semenogelin-1 | SEMG1 | 1.74* | 4.10* | 2.91* | 0.71 | 2.56* | 0.55* | 2.67* |
| Histidine ammonia-lyase | HAL | 1.60* | 2.11* | 5.94* | 0.35* | 2.50* | 0.56 | 2.12* |
| BPI fold-containing family A member 1 | BPIFA1 | 1.11 | 2.30* | 1.94* | 1.49* | 2.48* | 1.40 | 3.23* |
| Zymogen granule protein 16 homolog B | EECP | 1.16 | 1.50* | 1.36* | 1.18 | 2.47* | 1.02 | 2.66* |
| Galectin-7 | LGALS7 | 0.90 | 4.40* | 4.72* | 1.18 | 2.20* | 0.97 | 1.65 |
| Repetin | RPTN | 1.60* | 1.00 | 4.84* | 0.42* | 2.18* | 0.67 | 3.40* |
| Carboxypeptidase A4 | CPA4 | 0.89 | 1.16 | 1.45* | 0.83 | 2.09* | 0.70 | 1.86* |
| Hornerin | HRNR | 0.99 | 0.89 | 1.09 | 1.23 | 2.06* | 1.28 | 1.68* |
| Serine/threonine-protein kinase 31 | STK31 | 2.24* | 1.32 | 4.57* | 0.59* | 2.06* | 0.73 | 1.99* |
| Cystatin-M | CST6 | 1.81* | 3.67* | 3.47* | 0.54* | 1.96* | 0.58 | 1.84* |
| Myeloblastin | PRTN3 | 0.84 | 3.59* | 2.95* | 0.55* | 1.93* | 0.45* | 1.61 |
| Inositol polyphosphate-4-phosphatase type I A | INPP4A | 0.52* | 0.62 | 0.73 | 0.89 | 1.91* | 1.37 | 1.56 |
| Immunoglobulin heavy constant alpha 2 | IGHA2 | 1.13 | 3.35* | 14.54* | 0.74 | 1.90* | 1.16 | 4.45* |
| Corneodesmosin | CDSN | 1.08 | 0.72* | 0.79 | 1.44* | 1.86* | 0.87 | 1.75* |
| RING1 and YY1-binding protein | RYBP | 1.77* | 1.81* | 2.11* | 1.28 | 1.80 | 0.92 | 2.13* |
| Mammaglobin-B | SCGB2A1 | 1.32 | 5.33* | 2.76* | 0.64 | 1.80* | 0.64 | 1.75 |
| Mitotic spindle assembly checkpoint protein MAD2A | MAD2L1 | 1.28 | 1.24 | 1.18 | 1.44 | 1.73* | 1.40 | 1.91* |
| tRNA N6-adenosine threonylcarbamoyltransferase, mitochondrial | OSGEPL1 | 1.13 | 0.88 | 0.97 | 1.11 | 1.71 | 1.38 | 1.61 |
| SRSF protein kinase 2 | SRPK2 | 0.77 | 0.95 | 0.96 | 1.06 | 1.64* | 1.38 | 1.84* |

| | | | | | | | | |
|---|---|---|---|---|---|---|---|---|
| DDB1- and CUL4-associated factor 1 | DCAF1 | 0.85 | 0.86 | 0.90 | 1.20 | 1.64* | 1.44 | 1.69* |
| Kallikrein-10 | KLK10 | 1.24* | 0.99 | 1.30* | 1.49* | 1.63* | 1.37 | 1.51* |
| Plasminogen | PLG | 0.62 | 1.31 | 1.39 | 1.44 | 1.62 | 1.48 | 1.84 |
| Cell division cycle protein 16 homolog | CDC16 | 1.12 | 1.46 | 1.40 | 1.32 | 1.61* | 1.11 | 1.58 |
| Lysophosphatidic acid receptor 2 | LPAR2 | 1.31 | 0.98 | 1.18 | 0.93 | 1.60 | 1.36 | 1.54 |
| Transcription initiation factor TFIID subunit 5 | TAF5 | 0.45* | 0.59* | 0.49* | 0.70 | 1.60 | 1.45 | 1.68* |
| YTH domain-containing family protein 1 | YTHDF1 | 0.95 | 1.17 | 1.09 | 1.02 | 1.58* | 1.43 | 1.57* |
| BTB/POZ domain-containing protein KCTD14 | KCTD14 | 0.89 | 1.16 | 1.10 | 1.23 | 1.57 | 1.37 | 1.61 |
| Pyruvate kinase PKLR | PKLR | 0.74* | 1.35 | 1.19 | 1.31 | 1.57 | 1.24 | 1.64* |
| Retroviral-like aspartic protease 1 | ASPRV1 | 0.92 | 3.16* | 10.29* | 0.39* | 1.54* | 0.49* | 1.97* |
| Serine protease 1 | PRSS1 | 0.83 | 1.15 | 1.10 | 1.33 | 1.54* | 1.50 | 1.98* |
| Synaptojanin-1 | SYNJ1 | 0.74 | 0.96 | 0.94 | 1.01 | 1.53 | 1.49 | 1.61 |
| Transport and Golgi organization protein 2 homolog | TANGO2 | 1.01 | 1.34 | 1.26 | 1.11 | 1.52 | 1.26 | 1.79 |
| Amphiregulin | AREG | 0.63 | 1.46 | 1.11 | 1.32 | 1.51 | 1.25 | 3.02* |
| Cyclin-dependent kinase 1 | CDK1 | 0.62* | 0.67* | 0.68* | 0.40* | 1.51 | 1.34 | 1.54* |
| Putative hydroxypyruvate isomerase | HYI | 1.07 | 1.19 | 1.13 | 1.22 | 1.50 | 1.34 | 1.51 |
| Proto-oncogene c-Rel | REL | 0.63* | 0.88 | 0.79 | 1.17 | 1.50 | 0.99 | 1.70* |

***N. Common up-regulated proteins between LPS-Cytokines with Aquamin plus Mesalamine and with Mesalamine [105 proteins]***

| | | Interventions | | | | | | |
|---|---|---|---|---|---|---|---|---|
| | | Control | | | *With LPS & Cytokines* | | | |
| Proteins | Genes | AQ | AQ+MES | MES | *LPS-Cyto* | *AQ* | ***AQ+MES*** | ***MES*** |
| C-X-C motif chemokine 10 | CXCL10 | 0.72 | 34.05* | 43.44* | 0.97 | 0.83 | 30.10* | 22.62* |
| HLA class II histocompatibility antigen, DM beta chain | HLA-DMB | 1.59* | 13.66* | 8.99* | 1.13 | 1.05 | 10.01* | 8.97* |
| HLA class II histocompatibility antigen, DM alpha chain | HLA-DMA | 1.43 | 5.47* | 4.95* | 1.02 | 1.13 | 5.22* | 4.36* |
| HLA class II histocompatibility antigen, DR beta 4 chain | HLA-DRB4 | 0.71 | 3.84* | 4.17* | 0.84 | 1.08 | 4.68* | 3.60* |
| Chromogranin-A | CHGA | 0.64* | 5.56* | 5.04* | 0.86 | 1.34 | 4.03* | 6.57* |
| Fibroleukin | FGL2 | 0.77 | 3.23* | 3.07* | 0.79 | 0.93 | 3.82* | 3.15* |
| Complement C2 | C2 | 0.70 | 2.82* | 4.91* | 0.68 | 0.79 | 3.36* | 3.92* |
| Secretogranin-2 | SCG2 | 0.74 | 3.08* | 4.35* | 0.99 | 1.09 | 3.27* | 5.45* |
| Plasmalemma vesicle-associated protein | PLVAP | 0.75 | 3.13* | 3.90* | 1.02 | 1.13 | 3.00* | 3.92* |
| Centrosomal protein of 85 kDa | CEP85 | 0.93 | 4.39* | 4.17* | 1.04 | 1.23 | 2.87* | 4.16* |
| T-complex protein 10A homolog 1 | TCP10L | 0.85 | 1.26 | 2.12* | 0.87 | 0.83 | 2.79* | 2.40* |
| Synaptic vesicle membrane protein VAT-1 homolog-like | VAT1L | 0.71 | 3.05* | 4.20* | 0.93 | 1.19 | 2.75* | 4.58* |
| Tenascin-X | TNXB | 0.89 | 2.86* | 3.62* | 0.93 | 1.02 | 2.74* | 3.87* |
| Transmembrane protein 236 | TMEM236 | 1.79* | 4.00* | 2.65* | 1.34 | 1.43 | 2.59* | 1.66 |
| Phospholipid transfer protein | PLTP | 0.70 | 2.51* | 3.16* | 0.97 | 0.87 | 2.48* | 3.00* |

| | | | | | | | | |
|---|---|---|---|---|---|---|---|---|
| Cadherin-13 | CDH13 | 0.84 | 2.41* | 3.15* | 0.88 | 1.01 | 2.44* | 3.83* |
| Vasopressin-neurophysin 2-copeptin | AVP | 0.94 | 3.13* | 3.08* | 1.27 | 1.08 | 2.41* | 3.99* |
| EH domain-containing protein 3 | EHD3 | 0.76 | 3.83* | 3.66* | 1.19 | 1.06 | 2.38* | 3.59* |
| Receptor-type tyrosine-protein phosphatase zeta | PTPRZ1 | 0.81 | 2.75* | 4.03* | 0.97 | 1.19 | 2.32* | 3.43* |
| Interleukin-1 receptor accessory protein | IL1RAP | 0.61* | 2.35* | 2.99* | 0.79 | 0.92 | 2.31* | 3.58* |
| Contactin-1 | CNTN1 | 0.65* | 2.25* | 2.79* | 0.80 | 0.88 | 2.29* | 3.42* |
| A disintegrin and metalloproteinase with thrombospondin motifs 13 | ADAMTS13 | 0.68 | 2.03* | 2.82* | 0.65 | 0.85 | 2.28* | 2.94* |
| Hepatocyte growth factor activator | HGFAC | 0.62* | 2.13* | 2.48* | 0.77* | 0.77 | 2.27* | 3.01* |
| Oncoprotein-induced transcript 3 protein | OIT3 | 0.70* | 2.44* | 2.81* | 0.81 | 0.94 | 2.23* | 3.12* |
| Fibromodulin | FMOD | 0.64* | 2.45* | 2.75* | 1.06 | 0.95 | 2.23* | 3.19* |
| Moesin | MSN | 0.69* | 1.72* | 3.19* | 1.45* | 1.46* | 2.21* | 3.03* |
| Collagen alpha-1(XI) chain | COL11A1 | 0.53* | 2.32* | 2.75* | 0.87 | 0.94 | 2.20* | 3.10* |
| Beta-parvin | PARVB | 0.82 | 2.48* | 3.45* | 0.96 | 1.17 | 2.19* | 3.29* |
| Pigment epithelium-derived factor | SERPINF1 | 0.57* | 1.24 | 1.90* | 1.24 | 0.96 | 2.19* | 3.38* |
| NEDD4 family-interacting protein 2 | NDFIP2 | 1.07 | 2.34* | 2.44* | 0.95 | 1.12 | 2.19* | 2.96* |
| Retinol-binding protein 4 | RBP4 | 0.67* | 2.60* | 2.77* | 0.92 | 0.90 | 2.14* | 2.97* |
| Alpha-fetoprotein | AFP | 0.54* | 1.16 | 1.75* | 1.41* | 1.41* | 2.14* | 3.27* |
| Neural cell adhesion molecule 1 | NCAM1 | 0.68* | 2.18* | 2.49* | 0.81 | 0.77 | 2.12* | 3.03* |
| Metalloproteinase inhibitor 3 | TIMP3 | 0.80 | 3.49* | 2.39* | 0.78 | 0.88 | 2.09* | 1.51 |
| Tyrosine-protein kinase receptor Tie-1 | TIE1 | 0.68 | 1.62* | 2.43* | 0.79 | 0.81 | 2.09* | 2.76* |
| C-type mannose receptor 2 | MRC2 | 0.88 | 2.19* | 2.65* | 0.96 | 1.06 | 2.07* | 2.88* |
| Phosphatidylcholine-sterol acyltransferase | LCAT | 0.65* | 2.10* | 2.67* | 0.76 | 0.94 | 2.06* | 2.94* |
| Alpha-1B-glycoprotein | A1BG | 0.55* | 1.99* | 2.73* | 0.77 | 0.76 | 2.05* | 3.14* |
| Dynein axonemal heavy chain 1 | DNAH1 | 0.51* | 1.54* | 2.15* | 1.10 | 0.62* | 2.04* | 2.51* |
| Protein piccolo | PCLO | 0.56* | 1.75* | 2.05* | 0.55* | 0.72 | 2.03* | 2.36* |
| Afamin | AFM | 0.52* | 2.16* | 2.43* | 0.71* | 0.67* | 2.00* | 2.67* |
| Retinal dehydrogenase 2 | ALDH1A2 | 0.81 | 1.80* | 1.94* | 0.84 | 1.03 | 1.97* | 2.31* |
| Sex hormone-binding globulin | SHBG | 0.50* | 2.02* | 2.50* | 0.70 | 0.75 | 1.95* | 2.89* |
| Putative beta-actin-like protein 3 | POTEKP | 3.53* | 4.54* | 4.91* | 1.20 | 1.16 | 1.95* | 5.20* |
| Probable phosphoglycerate mutase 4 | PGAM4 | 0.53* | 1.15 | 2.54* | 0.51* | 0.63 | 1.94* | 2.77* |
| Regucalcin | RGN | 0.59* | 1.19 | 1.30 | 1.08 | 1.30 | 1.94* | 2.66* |
| Glutathione S-transferase A5 | GSTA5 | 0.75 | 1.99* | 2.22* | 0.78 | 0.90 | 1.92* | 2.62* |
| Plasma kallikrein | KLKB1 | 0.59* | 2.06* | 2.86* | 0.72 | 0.82 | 1.89* | 2.72* |
| Insulin | INS | 0.69* | 0.82 | 0.84 | 1.09 | 1.12 | 1.89* | 2.68* |
| Apolipoprotein A-II | APOA2 | 0.90 | 2.24* | 10.88* | 0.84 | 1.17 | 1.89* | 2.53* |
| EGF-containing fibulin-like extracellular matrix protein 1 | EFEMP1 | 0.62* | 2.26* | 2.51* | 0.81 | 0.53* | 1.88* | 2.37* |
| Collagen alpha-2(I) chain | COL1A2 | 0.53* | 2.15* | 2.47* | 0.72 | 0.65 | 1.88* | 2.71* |

| | | | | | | | | |
|---|---|---|---|---|---|---|---|---|
| FERM and PDZ domain-containing protein 1 | FRMPD1 | 0.54* | 2.61* | 2.58* | 0.47* | 0.94 | 1.85* | 2.22* |
| Plastin-2 | LCP1 | 0.68* | 2.28* | 7.32* | 0.81 | 0.92 | 1.85* | 2.39* |
| Interferon-induced protein 44-like | IFI44L | 0.60* | 1.47 | 1.36 | 1.22 | 1.34 | 1.82 | 3.21* |
| Scrapie-responsive protein 1 | SCRG1 | 0.64* | 2.21* | 2.25* | 0.80 | 0.99 | 1.82* | 2.12* |
| Collagen alpha-1(XXI) chain | COL21A1 | 0.75 | 1.78* | 2.36* | 0.97 | 0.96 | 1.81* | 2.34* |
| DENN domain-containing protein 3 | DENND3 | 1.10 | 1.37 | 1.25 | 1.16 | 0.91 | 1.80 | 1.74 |
| Steryl-sulfatase | STS | 1.09 | 1.27 | 1.24 | 1.42* | 1.36 | 1.79* | 1.57 |
| Cell adhesion molecule 1 | CADM1 | 0.65* | 1.89* | 2.04* | 0.63 | 0.89 | 1.76* | 2.25* |
| ERC protein 2 | ERC2 | 0.70 | 2.27* | 2.27* | 0.79 | 0.70 | 1.76 | 2.58* |
| Selenoprotein P | SELENOP | 0.80 | 1.87* | 2.83* | 0.81 | 1.24 | 1.76* | 1.95* |
| DnaJ homolog subfamily B member 9 | DNAJB9 | 1.26 | 1.81* | 1.41 | 1.32 | 1.26 | 1.76 | 1.72 |
| Mucin-3A | MUC3A | 1.00 | 2.47* | 2.01* | 1.05 | 1.21 | 1.75* | 1.67* |
| Acyl-coenzyme A synthetase ACSM3, mitochondrial | ACSM3 | 1.15 | 1.53 | 1.29 | 1.43 | 1.41 | 1.74* | 1.81* |
| Cerebellin-4 | CBLN4 | 0.41* | 1.58 | 2.52* | 0.57* | 0.78 | 1.73 | 1.95* |
| Ubiquitin carboxyl-terminal hydrolase isozyme L1 | UCHL1 | 0.88 | 0.91 | 1.36 | 1.44 | 1.28 | 1.72 | 1.75 |
| Exocyst complex component 3-like protein 4 | EXOC3L4 | 0.82 | 1.70* | 1.67* | 0.79 | 0.71 | 1.71* | 1.62 |
| 2'-5'-oligoadenylate synthase-like protein | OASL | 1.25 | 1.70* | 1.54* | 1.09 | 1.26 | 1.71* | 1.54 |
| TRPM8 channel-associated factor 2 | TCAF2 | 1.14 | 1.76* | 2.34* | 1.03 | 1.43 | 1.71 | 1.71 |
| Carboxypeptidase E | CPE | 0.79 | 2.20* | 2.47* | 0.75 | 0.99 | 1.70 | 2.53* |
| Tissue factor | F3 | 0.97 | 1.60* | 1.48* | 1.34* | 1.23 | 1.69* | 1.58* |
| Transforming growth factor-beta-induced protein ig-h3 | TGFBI | 0.93 | 1.95* | 1.72* | 1.09 | 1.36 | 1.69* | 2.50* |
| Complement factor H | CFH | 0.62* | 2.03* | 27.05* | 0.74 | 0.62 | 1.68* | 1.72* |
| Membrane primary amine oxidase | AOC3 | 0.85 | 2.15* | 3.07* | 0.93 | 1.18 | 1.67 | 2.86* |
| Cytochrome P450 2J2 | CYP2J2 | 0.90 | 1.14 | 1.05 | 1.03 | 1.36 | 1.67 | 1.54 |
| Collagen alpha-1(III) chain | COL3A1 | 0.82 | 1.61* | 2.22* | 0.72 | 0.87 | 1.66* | 2.04* |
| Peptidyl-prolyl cis-trans isomerase E | PPIE | 0.92 | 1.14 | 1.34* | 1.12 | 1.27 | 1.66* | 2.21* |
| Insulin-like growth factor II | IGF2 | 0.62* | 2.07* | 2.04* | 0.74 | 0.48* | 1.65* | 2.22* |
| Protein GUCD1 | GUCD1 | 1.02 | 1.64* | 1.44 | 0.74 | 1.48 | 1.65 | 1.75 |
| Probable serine carboxypeptidase CPVL | CPVL | 0.97 | 1.32 | 1.59* | 1.34 | 1.15 | 1.65* | 2.03* |
| Desumoylating isopeptidase 1 | DESI1 | 0.95 | 1.72* | 1.76* | 1.06 | 1.21 | 1.64 | 2.27* |
| Coagulation factor XIII A chain | F13A1 | 0.79 | 1.13 | 1.36 | 1.44 | 1.37 | 1.62 | 2.10* |
| Latent-transforming growth factor beta-binding protein 4 | LTBP4 | 0.82 | 2.90* | 2.39* | 1.01 | 1.40 | 1.61 | 3.16* |
| Microtubule-associated proteins 1A/1B light chain 3 beta 2 | MAP1LC3B2 | 1.26 | 1.67* | 1.73* | 1.34 | 1.32 | 1.61 | 1.83* |
| DNA ligase 1 | LIG1 | 0.88 | 0.85 | 0.81 | 0.95 | 1.47 | 1.60 | 1.53 |
| Tropomyosin beta chain | TPM2 | 0.68* | 1.91* | 1.88* | 0.86 | 0.83 | 1.59 | 2.29* |
| Coilin | COIL | 0.37* | 0.67 | 0.64* | 0.77 | 1.39 | 1.58 | 1.54 |
| Forkhead box protein G1 | FOXG1 | 0.98 | 1.32 | 1.34 | 1.04 | 1.17 | 1.58 | 1.75* |

| | | | | | | | | |
|---|---|---|---|---|---|---|---|---|
| Cytosolic iron-sulfur assembly component 2B | CIAO2B | 0.45* | 0.60* | 1.29 | 0.47* | 0.38* | 1.57 | 1.87 |
| Desmoglein-3 | DSG3 | 0.68 | 3.24* | 4.23* | 0.59 | 0.94 | 1.57 | 2.39* |
| ER degradation-enhancing alpha-mannosidase-like protein 2 | EDEM2 | 1.23 | 1.67* | 1.68* | 1.01 | 1.30 | 1.56 | 1.89* |
| Tubulointerstitial nephritis antigen-like | TINAGL1 | 0.67* | 0.93 | 1.35* | 1.10 | 1.17 | 1.55* | 2.61* |
| Sorting nexin-18 | SNX18 | 1.13 | 1.23 | 1.20 | 1.12 | 1.42 | 1.55 | 1.51 |
| Complement factor D | CFD | 0.59* | 1.43 | 3.72* | 0.81 | 0.58 | 1.54 | 2.05* |
| Guanidinoacetate N-methyltransferase | GAMT | 0.55* | 1.98* | 1.89* | 0.67 | 0.57 | 1.54 | 1.94* |
| Laminin subunit alpha-2 | LAMA2 | 1.06 | 1.20 | 1.71* | 1.33 | 1.36 | 1.54 | 1.76 |
| E3 ubiquitin-protein ligase DTX3L | DTX3L | 0.93 | 0.99 | 0.95 | 1.42* | 1.45 | 1.53* | 1.62* |
| Glycosylphosphatidylinositol anchor attachment 1 protein | GPAA1 | 1.14 | 1.22 | 1.08 | 1.18 | 1.33 | 1.52* | 1.88* |
| Septin-6 | SEPTIN6 | 0.72 | 0.95 | 0.96 | 1.16 | 1.22 | 1.52 | 1.82* |
| Desmoglein-4 | DSG4 | 1.01 | 65.35* | 2.23* | 0.77 | 1.18 | 1.51 | 2.16* |
| Angiopoietin-related protein 4 | ANGPTL4 | 1.12 | 0.96 | 1.32 | 1.31 | 1.32 | 1.51 | 1.62 |
| Lysyl oxidase homolog 3 | LOXL3 | 0.95 | 1.24 | 1.68* | 0.97 | 1.28 | 1.51 | 1.85* |
| Agrin | AGRN | 0.86 | 1.17 | 1.13 | 1.28* | 1.30 | 1.51* | 2.14* |
| Protein PML | PML | 0.89 | 0.87 | 0.96 | 1.21 | 0.95 | 1.51 | 1.75 |

***O. Common up-regulated proteins among LPS-Cytokines with Aquamin, with Aquamin plus Mesalamine and with Measlamine [137 proteins]***

| | | Interventions | | | | | | |
|---|---|---|---|---|---|---|---|---|
| | | Control | | | *With LPS & Cytokines* | | | |
| Proteins | Genes | AQ | AQ+MES | MES | *LPS-Cyto* | ***AQ*** | ***AQ+MES*** | ***MES*** |
| Ligand of Numb protein X 2 | LNX2 | 1.06 | 1.01 | 0.96 | 1.48 | 6.81* | 4.23* | 4.01* |
| Myosin-2 | MYH2 | 0.56* | 0.80 | 0.45* | 1.12 | 5.57* | 4.93* | 4.82* |
| Endoplasmic reticulum protein SC65 | P3H4 | 0.73 | 0.82 | 0.75 | 1.40 | 5.17* | 4.16* | 5.01* |
| Ephrin type-B receptor 3 | EPHB3 | 0.51* | 0.62* | 0.60* | 1.22 | 4.74* | 3.26* | 4.09* |
| Heparan sulfate glucosamine 3-O-sulfotransferase 1 | HS3ST1 | 0.19* | 0.35* | 0.29* | 1.17 | 4.51* | 3.55* | 5.50* |
| Low-density lipoprotein receptor-related protein 2 | LRP2 | 0.60* | 1.46 | 1.02 | 1.45 | 4.10* | 5.03* | 6.63* |
| Mammaglobin-A | SCGB2A2 | 1.06 | 1.93* | 1.71* | 1.32 | 4.09* | 1.54 | 3.31* |
| Pleckstrin | PLEK | 0.56* | 1.21 | 0.63* | 1.27 | 4.09* | 3.74* | 4.36* |
| Endothelial lipase | LIPG | 0.35* | 0.46* | 0.43* | 1.12 | 4.07* | 3.10* | 3.50* |
| Alpha-1-antichymotrypsin | SERPINA3 | 1.08 | 1.90* | 2.88* | 1.28 | 3.87* | 1.63 | 2.12* |
| Beta-1,3-N-acetylglucosaminyltransferase lunatic fringe | LFNG | 0.67* | 0.50* | 0.41* | 0.90 | 3.55* | 2.99* | 3.10* |
| Bone morphogenetic protein 1 | BMP1 | 0.22* | 0.37* | 0.31* | 0.91 | 3.52* | 2.50* | 3.63* |
| DNA topoisomerase 2-alpha | TOP2A | 0.73* | 0.82 | 0.69* | 0.95 | 3.45* | 3.10* | 3.18* |
| Coagulation factor X | F10 | 0.41* | 0.77 | 0.81 | 1.13 | 3.39* | 5.07* | 5.60* |
| Reticulocalbin-3 | RCN3 | 0.17* | 0.72 | 0.78 | 1.01 | 3.31* | 4.57* | 5.68* |

| | | | | | | | | |
|---|---|---|---|---|---|---|---|---|
| Immunoglobulin lambda constant 2 | IGLC2 | 0.66* | 1.33 | 4.95* | 1.42 | 3.25* | 2.14* | 1.55 |
| Protein Wnt-3a | WNT3A | 0.49* | 0.92 | 0.90 | 1.17 | 3.24* | 4.10* | 5.41* |
| Mimecan | OGN | 0.29* | 0.85 | 0.94 | 1.34 | 3.21* | 4.58* | 5.97* |
| Serum paraoxonase/arylesterase 1 | PON1 | 0.69 | 1.33 | 1.69* | 1.47 | 3.15* | 4.26* | 4.58* |
| Ceruloplasmin | CP | 0.63* | 1.04 | 1.10 | 1.39* | 3.14* | 3.98* | 4.43* |
| Glutathione S-transferase Mu 2 | GSTM2 | 0.74 | 1.19 | 1.67* | 1.22 | 3.12* | 3.94* | 3.58* |
| Carboxypeptidase N catalytic chain | CPN1 | 0.56* | 0.92 | 0.92 | 1.44* | 3.11* | 4.54* | 5.08* |
| Cilia- and flagella-associated protein 45 | CFAP45 | 1.69* | 1.98* | 2.43* | 1.19 | 3.11* | 2.25* | 2.51* |
| Apolipoprotein C-I | APOC1 | 0.88 | 1.73* | 2.48* | 1.47* | 3.07* | 4.84* | 3.83* |
| Matrix metalloproteinase-28 | MMP28 | 0.33* | 0.49* | 0.56* | 0.80 | 3.03* | 2.32* | 2.79* |
| Mast/stem cell growth factor receptor Kit | KIT | 0.70 | 1.13 | 0.66* | 0.79 | 3.01* | 2.43* | 2.74* |
| DNA (cytosine-5)-methyltransferase 1 | DNMT1 | 0.48* | 0.32* | 0.41* | 0.70 | 3.00* | 2.01* | 2.38* |
| Fermitin family homolog 3 | FERMT3 | 0.48* | 1.02 | 0.99 | 1.11 | 2.94* | 4.31* | 4.74* |
| Immunoglobulin kappa constant | IGKC | 0.71* | 1.56* | 4.90* | 1.34* | 2.92* | 1.88* | 2.00* |
| FK506-binding protein-like | FKBPL | 0.89 | 0.94 | 0.99 | 1.11 | 2.90* | 2.69* | 1.57 |
| Tumor necrosis factor ligand superfamily member 9 | TNFSF9 | 0.73 | 0.93 | 0.78 | 1.36 | 2.86* | 2.06* | 2.33* |
| Serine incorporator 5 | SERINC5 | 0.80 | 0.94 | 0.92 | 1.49* | 2.82* | 2.63* | 2.45* |
| Dihydropyrimidinase | DPYS | 0.33* | 0.71 | 0.75 | 1.13 | 2.82* | 3.76* | 4.94* |
| Microtubule-associated protein 1B | MAP1B | 0.42* | 0.77 | 0.85 | 1.20 | 2.79* | 2.86* | 3.79* |
| Protocadherin gamma-C3 | PCDHGC3 | 0.54* | 1.39 | 1.37* | 1.48* | 2.77* | 3.66* | 5.21* |
| Complement component C7 | C7 | 0.72 | 1.09 | 1.33 | 1.47 | 2.77* | 4.26* | 4.88* |
| C-type lectin domain family 11 member A | CLEC11A | 0.51* | 1.23 | 1.22 | 1.20 | 2.68* | 3.66* | 4.12* |
| Transmembrane protein 223 | TMEM223 | 1.04 | 0.97 | 0.93 | 1.24 | 2.66* | 1.67 | 1.61 |
| RAF proto-oncogene serine/threonine-protein kinase | RAF1 | 0.83 | 0.87 | 0.84 | 1.08 | 2.65* | 2.68* | 2.02* |
| Collectin-10 | COLEC10 | 0.62* | 1.16 | 1.14 | 1.23 | 2.63* | 3.78* | 4.85* |
| Dynamin-1 | DNM1 | 0.57* | 1.02 | 0.79 | 1.22 | 2.61* | 1.81* | 2.14* |
| Transmembrane gamma-carboxyglutamic acid protein 1 | PRRG1 | 0.88 | 1.02 | 1.15 | 1.38 | 2.61* | 2.51* | 1.80* |
| Collectin-11 | COLEC11 | 0.28* | 0.79 | 1.14 | 1.30 | 2.59* | 3.59* | 5.27* |
| Solute carrier family 35 member B1 | SLC35B1 | 2.33* | 2.96* | 3.17* | 1.47 | 2.58* | 2.91* | 2.01* |
| Condensin complex subunit 2 | NCAPH | 1.51* | 1.22 | 1.07 | 1.36 | 2.49* | 2.98* | 1.72* |
| Vasorin | VASN | 0.39* | 0.96 | 0.86 | 1.44 | 2.45* | 3.55* | 5.06* |
| Periostin | POSTN | 0.66* | 1.63* | 1.51* | 1.02 | 2.43* | 2.54* | 3.38* |
| CCR4-NOT transcription complex subunit 6 | CNOT6 | 0.63* | 0.77 | 0.73 | 1.15 | 2.38* | 1.85* | 2.10* |
| Anaphase-promoting complex subunit 1 | ANAPC1 | 0.33* | 0.29* | 0.32* | 0.60* | 2.35* | 1.97* | 2.10* |
| Protein spinster homolog 1 | SPNS1 | 1.08 | 1.18 | 1.22 | 1.45 | 2.35* | 2.03* | 2.00* |
| Cartilage-associated protein | CRTAP | 0.97 | 1.09 | 1.04 | 1.21 | 2.34* | 2.51* | 2.31* |
| MANSC domain-containing protein 1 | MANSC1 | 0.94 | 1.34 | 1.50* | 1.10 | 2.34* | 2.35* | 1.72* |

| | | | | | | | | |
|---|---|---|---|---|---|---|---|---|
| Procollagen C-endopeptidase enhancer 1 | PCOLCE | 0.69 | 1.78* | 2.20* | 0.96 | 2.34* | 2.41* | 2.65* |
| Alpha-N-acetylgalactosaminide alpha-2,6-sialyltransferase 6 | ST6GALNAC6 | 1.10 | 1.02 | 1.14 | 1.14 | 2.33* | 2.20* | 1.93* |
| Plasmolipin | PLLP | 1.13 | 1.06 | 1.05 | 1.50* | 2.31* | 1.82* | 1.64* |
| von Willebrand factor A domain-containing protein 1 | VWA1 | 0.61* | 1.07 | 1.14 | 1.04 | 2.29* | 2.51* | 3.15* |
| N-acetyllactosaminide beta-1,3-N-acetylglucosaminyltransferase 2 | B3GNT2 | 1.29 | 1.91* | 1.58* | 1.36 | 2.26* | 2.13* | 2.17* |
| Proton myo-inositol cotransporter | SLC2A13 | 1.34 | 1.49 | 1.29 | 1.28 | 2.22* | 2.37* | 1.80* |
| Complement component C9 | C9 | 0.83 | 1.47* | 1.50* | 1.29 | 2.22* | 3.31* | 3.45* |
| Sushi repeat-containing protein SRPX2 | SRPX2 | 0.71* | 1.06 | 1.21 | 0.82 | 2.21* | 1.83* | 2.35* |
| Noggin | NOG | 0.90 | 1.20 | 1.12 | 1.37 | 2.18* | 3.03* | 2.78* |
| Protein SYS1 homolog | SYS1 | 1.18 | 1.11 | 1.13 | 1.38 | 2.13* | 2.08* | 1.69 |
| Anaphase-promoting complex subunit CDC26 | CDC26 | 0.99 | 1.15 | 1.18 | 1.36 | 2.10* | 2.14* | 1.97 |
| Serpin B7 | SERPINB7 | 1.00 | 1.84* | 1.21 | 1.19 | 2.08* | 2.49* | 1.71* |
| BolA-like protein 1 | BOLA1 | 1.13 | 0.84 | 1.21 | 1.37 | 2.08* | 1.51 | 1.58 |
| Deoxynucleoside triphosphate triphosphohydrolase SAMHD1 | SAMHD1 | 0.65 | 1.18 | 1.07 | 1.16 | 2.07* | 1.70 | 1.86 |
| Glucose-6-phosphate exchanger SLC37A4 | SLC37A4 | 1.22 | 1.37 | 1.24 | 1.36 | 2.07* | 2.39* | 1.67 |
| Bromodomain-containing protein 9 | BRD9 | 0.70* | 1.71* | 1.15 | 1.38* | 2.07* | 3.25* | 5.06* |
| Ectonucleoside triphosphate diphosphohydrolase 6 | ENTPD6 | 1.31 | 1.39 | 1.18 | 1.44 | 2.06* | 2.00* | 1.81* |
| Proteasomal ATPase-associated factor 1 | PAAF1 | 0.79 | 1.33 | 1.03 | 1.34 | 2.05* | 1.58 | 2.35* |
| SPARC-related modular calcium-binding protein 2 | SMOC2 | 0.84 | 1.62* | 1.69* | 1.38 | 2.05* | 1.83* | 1.95* |
| Cytochrome c oxidase assembly protein COX11, mitochondrial | COX11 | 1.28 | 1.96* | 2.23* | 1.39 | 2.04* | 2.52* | 3.10* |
| Trafficking protein particle complex subunit 2 | TRAPPC2 | 1.13 | 1.07 | 1.03 | 1.43* | 2.03* | 1.85* | 1.83* |
| Nidogen-2 | NID2 | 0.73 | 1.89* | 1.61* | 1.45* | 2.02* | 2.55* | 3.43* |
| 4-galactosyl-N-acetylglucosaminide 3-alpha-L-fucosyltransferase FUT6 | FUT6 | 1.30* | 1.36 | 1.24 | 1.39 | 2.02* | 1.83* | 1.84* |
| Galactose-3-O-sulfotransferase 2 | GAL3ST2 | 0.84 | 0.77 | 0.80 | 0.85 | 2.02* | 1.95* | 1.90* |
| Sterile alpha motif domain-containing protein 9-like | SAMD9L | 1.07 | 1.96* | 1.88* | 1.41 | 2.01* | 3.01* | 3.00* |
| Centrin-3 | CETN3 | 1.05 | 1.10 | 0.89 | 1.21 | 2.00* | 2.02* | 1.57 |
| Bcl-2-like protein 1 | BCL2L1 | 1.01 | 1.01 | 0.95 | 1.48* | 2.00* | 1.88* | 1.78* |
| Collagen alpha-1(I) chain | COL1A1 | 0.89 | 1.53* | 1.40* | 1.45* | 1.99* | 2.46* | 3.30* |
| RRP15-like protein | RRP15 | 1.11 | 0.98 | 0.88 | 1.49* | 1.99* | 1.63* | 2.82* |
| Sodium/hydrogen exchanger 6 | SLC9A6 | 1.17 | 1.09 | 1.20 | 1.35 | 1.96* | 2.05* | 1.54 |
| Fumarylacetoacetase | FAH | 0.97 | 0.83 | 0.99 | 1.31 | 1.96* | 1.77* | 1.88* |
| Growth factor receptor-bound protein 7 | GRB7 | 0.74 | 0.68 | 0.71 | 1.04 | 1.95* | 1.91* | 1.61 |
| Acyl-coenzyme A thioesterase MBLAC2 | MBLAC2 | 1.00 | 1.09 | 1.30 | 1.48* | 1.92* | 1.83* | 1.85* |
| Sulfotransferase 2A1 | SULT2A1 | 1.49* | 5.11* | 2.72* | 1.25 | 1.90* | 2.73* | 1.62 |

| | | | | | | | | |
|---|---|---|---|---|---|---|---|---|
| UNC93-like protein MFSD11 | MFSD11 | 1.13 | 1.15 | 1.06 | 1.30 | 1.88* | 1.93* | 1.52 |
| Nidogen-1 | NID1 | 0.75* | 1.56* | 1.44* | 1.41* | 1.86* | 2.21* | 2.89* |
| CD177 antigen | CD177 | 1.04 | 1.45* | 1.41* | 1.26 | 1.85* | 2.23* | 1.55* |
| Laminin subunit beta-2 | LAMB2 | 0.67* | 1.91* | 1.30* | 1.38* | 1.83* | 2.20* | 3.05* |
| HIG1 domain family member 1A, mitochondrial | HIGD1A | 1.46 | 1.58* | 1.10 | 1.43 | 1.83* | 2.31* | 1.94* |
| cAMP-specific 3',5'-cyclic phosphodiesterase 4C | PDE4C | 0.89 | 2.07* | 2.99* | 0.79 | 1.82* | 2.80* | 3.99* |
| Solute carrier organic anion transporter family member 2B1 | SLCO2B1 | 1.05 | 1.32 | 1.49* | 1.04 | 1.81 | 1.86 | 1.72 |
| Stromal cell-derived factor 2 | SDF2 | 0.95 | 1.08 | 1.16 | 1.25 | 1.80* | 1.78* | 1.55 |
| Alpha-1,3-mannosyl-glycoprotein 2-beta-N-acetylglucosaminyltransferase | MGAT1 | 1.12 | 1.44* | 1.50* | 1.39* | 1.79* | 1.89* | 1.90* |
| Fibulin-1 | FBLN1 | 0.88 | 1.47* | 1.43* | 1.41* | 1.79* | 1.99* | 1.99* |
| Cytochrome c oxidase assembly protein COX20, mitochondrial | COX20 | 1.21* | 1.14 | 1.03 | 1.46* | 1.79* | 1.59* | 1.64* |
| UPF0606 protein KIAA1549L | KIAA1549L | 1.36 | 0.96 | 1.23 | 1.26 | 1.77* | 1.54 | 1.62 |
| Retinol-binding protein 1 | RBP1 | 1.18 | 1.15 | 1.11 | 1.21 | 1.75* | 2.10* | 1.53 |
| Carboxypeptidase A2 | CPA2 | 1.24 | 3.77* | 2.77* | 0.76 | 1.74 | 4.32* | 2.79* |
| F-box only protein 2 | FBXO2 | 1.08 | 2.13* | 1.67* | 0.90 | 1.73* | 2.19* | 1.63 |
| Molybdate-anion transporter | MFSD5 | 1.26 | 1.19 | 1.13 | 1.22 | 1.73* | 1.70* | 1.52 |
| Vesicle transport protein GOT1B | GOLT1B | 1.02 | 1.29 | 1.37* | 1.27 | 1.73* | 2.35* | 1.58 |
| Chloride intracellular channel protein 5 | CLIC5 | 1.22* | 1.32 | 1.26* | 1.41* | 1.73* | 1.54* | 1.52* |
| Out at first protein homolog | OAF | 0.98 | 1.36 | 0.99 | 1.38 | 1.71* | 1.51 | 1.61 |
| Testis-expressed protein 9 | TEX9 | 1.68* | 1.53* | 0.95 | 1.42* | 1.70* | 2.57* | 2.77* |
| Acetylcholinesterase | ACHE | 1.68* | 3.16* | 2.26* | 1.09 | 1.69 | 2.30* | 1.62 |
| Lysyl oxidase homolog 4 | LOXL4 | 0.90 | 1.02 | 1.10 | 1.34 | 1.69 | 1.66 | 1.90* |
| Nuclear autoantigen Sp-100 | SP100 | 1.01 | 1.18 | 0.89 | 1.33 | 1.67 | 2.13* | 2.28* |
| Uncharacterized protein MISP3 | MISP3 | 1.18 | 1.36 | 1.56* | 1.30 | 1.66 | 1.70 | 2.06* |
| Collagen alpha-1(V) chain | COL5A1 | 1.02 | 1.90* | 1.77* | 1.10 | 1.65* | 2.12* | 2.62* |
| Mitochondrial thiamine pyrophosphate carrier | SLC25A19 | 1.15 | 1.00 | 0.97 | 1.28 | 1.65 | 1.54 | 1.51 |
| Bisphosphoglycerate mutase | BPGM | 0.83 | 1.30 | 1.11 | 1.07 | 1.64 | 1.94* | 2.03* |
| WD repeat-containing protein 70 | WDR70 | 0.91 | 0.86 | 0.97 | 1.03 | 1.63* | 1.55* | 1.56* |
| Proteinase-activated receptor 2 | F2RL1 | 1.12 | 1.22 | 1.20 | 1.40* | 1.63* | 1.78* | 1.69* |
| Clusterin | CLU | 0.83 | 0.92 | 1.32 | 1.31 | 1.61 | 1.50 | 2.08* |
| Heat shock 70 kDa protein 13 | HSPA13 | 0.96 | 1.49* | 1.51* | 1.27 | 1.61* | 1.99* | 2.46* |
| Mitochondrial fission process protein 1 | MTFP1 | 1.06 | 1.51* | 1.46* | 1.44 | 1.61* | 1.62* | 1.80* |
| Follistatin-related protein 1 | FSTL1 | 0.95 | 4.04* | 4.57* | 1.47* | 1.61 | 3.52* | 5.73* |
| Protein S100-A3 | S100A3 | 1.12 | 26.05* | 2.82* | 0.58* | 1.60 | 1.77 | 3.12* |
| Collagen alpha-2(VI) chain | COL6A2 | 0.64* | 1.24 | 1.33 | 1.16 | 1.60 | 2.85* | 3.58* |

| | | | | | | | | |
|---|---|---|---|---|---|---|---|---|
| eEF1A lysine and N-terminal methyltransferase | METTL13 | 0.70 | 0.78 | 0.82 | 0.87 | 1.59 | 1.63 | 1.60 |
| Mitochondrial glutamate carrier 2 | SLC25A18 | 0.86 | 1.95* | 2.62* | 0.94 | 1.58 | 2.10* | 3.70* |
| Collagen alpha-2(IV) chain | COL4A2 | 0.86 | 1.37 | 1.05 | 1.41* | 1.58* | 2.07* | 3.23* |
| Ras-related protein Rab-8B | RAB8B | 0.94 | 1.25 | 1.10 | 1.33 | 1.58 | 1.67* | 1.51 |
| CD70 antigen | CD70 | 0.94 | 1.26 | 1.25 | 1.30 | 1.58 | 1.63 | 1.60 |
| Type-1 angiotensin II receptor-associated protein | AGTRAP | 1.15 | 1.18 | 1.22 | 1.25 | 1.58 | 1.64 | 1.65 |
| Mediator of RNA polymerase II transcription subunit 1 | MED1 | 0.91 | 1.28 | 1.27 | 1.27 | 1.57 | 1.76* | 1.95* |
| Solute carrier family 2, facilitated glucose transporter member 5 | SLC2A5 | 1.25* | 6.90* | 5.49* | 1.35* | 1.55* | 6.09* | 4.52* |
| Lumican | LUM | 0.67* | 1.73* | 1.81* | 1.41* | 1.54* | 2.50* | 3.78* |
| WD repeat-containing protein 74 | WDR74 | 0.74 | 0.58* | 0.71 | 0.94 | 1.54 | 1.63 | 1.63 |
| Ubiquitin-like modifier-activating enzyme 7 | UBA7 | 1.21 | 1.29 | 1.14 | 1.39* | 1.53* | 1.66* | 1.57* |
| Midasin | MDN1 | 0.66* | 1.29 | 1.56* | 1.48* | 1.53* | 2.03* | 2.59* |
| Dynein axonemal assembly factor 10 | DNAAF10 | 1.08 | 1.30 | 1.22 | 1.36 | 1.52 | 1.55 | 1.65 |
| Collagen alpha-1(VI) chain | COL6A1 | 0.88 | 1.56* | 1.49* | 1.06 | 1.51* | 2.05* | 2.64* |
| Prolyl 3-hydroxylase 1 | P3H1 | 1.03 | 1.04 | 1.15 | 1.16 | 1.51 | 1.55 | 1.79* |
| Basement membrane-specific heparan sulfate proteoglycan core protein | HSPG2 | 0.86* | 1.45* | 1.21* | 1.18 | 1.51* | 1.85* | 2.07* |

Values represent the abundance ratio from organoids (n=4 subjects) compared to the control. These proteins were up-regulated at a 1.5-fold change (<2% FDR). Corresponding abundance ratios from the other treatment groups are provided for comparison. Proteins common among groups and unique to individual groups under proinflammatory conditions (LPS-Cytokines) are presented. *Indicates significance compared to the control (at $p<0.05$).
