## Supplementary material for "Proteomic Profile of Human Colon Organoids: Effects of a multi-mineral intervention alone and in the presence of pro-Inflammatory and anti-inflammatory treatments": S Table 6

**Supplement Table 6. Down-regulated proteins influenced by LPS-Cytokines alone and with Aquamin and Mesalamine (with 1.5-fold)**

***A. Common among four groups: LPS-Cytokines, +Aquamin, +Mesalamine and +Aquamin plus Mesalamine (261 proteins)***

| | | Interventions | | | | | | |
|---|---|---|---|---|---|---|---|---|
| | | Control | | | *With LPS & Cytokines* | | | |
| Proteins | Genes | AQ | AQ+MES | MES | ***LPS-Cyto*** | ***AQ*** | ***AQ+MES*** | ***MES*** |
| WD repeat and FYVE domain-containing protein 3 | WDFY3 | 0.23* | 0.09* | 0.12* | 0.09* | 0.07* | 0.18* | 0.06* |
| TBC1 domain family member 25 | TBC1D25 | 0.21* | 0.09* | 0.08* | 0.10* | 0.11* | 0.15* | 0.07* |
| Rho guanine nucleotide exchange factor 3 | ARHGEF3 | 0.18* | 0.44* | 0.36* | 0.12* | 0.48* | 0.17* | 0.24* |
| Integrin alpha-E | ITGAE | 0.27* | 0.09* | 0.14* | 0.13* | 0.10* | 0.22* | 0.09* |
| PCNA-interacting partner | PARPBP | 0.40* | 0.11* | 0.08* | 0.13* | 0.12* | 0.16* | 0.06* |
| Guanine nucleotide-binding protein-like 3-like protein | GNL3L | 0.23* | 0.38* | 0.37* | 0.14* | 0.22* | 0.31* | 0.20* |
| Polyamine-modulated factor 1-binding protein 1 | PMFBP1 | 0.39* | 0.15* | 0.13* | 0.14* | 0.15* | 0.25* | 0.08* |
| Zinc finger protein 654 | ZNF654 | 0.19* | 0.24* | 0.21* | 0.15* | 0.36* | 0.33* | 0.29* |
| Zinc finger protein with KRAB and SCAN domains 4 | ZKSCAN4 | 0.27* | 0.19* | 0.22* | 0.15* | 0.37* | 0.19* | 0.22* |
| Protein kinase C theta type | PRKCQ | 0.26* | 0.08* | 0.11* | 0.16* | 0.14* | 0.22* | 0.11* |
| Kelch-like ECH-associated protein 1 | KEAP1 | 0.19* | 0.33* | 0.37* | 0.18* | 0.39* | 0.32* | 0.29* |
| E3 SUMO-protein ligase ZNF451 | ZNF451 | 0.32* | 0.37* | 0.35* | 0.19* | 0.43* | 0.44* | 0.36* |
| NEDD4-binding protein 3 | N4BP3 | 0.35* | 0.47* | 0.43* | 0.20* | 0.36* | 0.29* | 0.26* |
| Zinc finger protein 136 | ZNF136 | 0.20* | 0.18* | 0.20* | 0.20* | 0.26* | 0.23* | 0.17* |
| Protein S100-A7A | S100A7A | 0.27* | 0.40* | 4.36* | 0.20* | 0.48* | 0.17* | 0.46* |
| Oxytocin-neurophysin 1 | OXT | 0.28* | 0.52* | 0.47* | 0.20* | 0.20* | 0.50* | 0.37* |
| Centrosomal protein of 131 kDa | CEP131 | 0.35* | 0.36* | 0.35* | 0.21* | 0.33* | 0.09* | 0.09* |
| Protein ECT2 | ECT2 | 0.28* | 0.38* | 0.40* | 0.25* | 0.64 | 0.62 | 0.55* |
| Protein Shroom1 | SHROOM1 | 0.34* | 0.73 | 0.64* | 0.26* | 0.49* | 0.40* | 0.24* |
| Sorting nexin-11 | SNX11 | 0.89 | 1.08 | 1.16 | 0.26* | 0.06* | 0.16* | 0.28* |
| DEP domain-containing mTOR-interacting protein | DEPTOR | 0.53* | 0.74 | 0.75* | 0.26* | 0.46* | 0.39* | 0.27* |
| Transcription factor Sp1 | SP1 | 0.39* | 0.49* | 0.53* | 0.26* | 0.40* | 0.44* | 0.46* |
| Telomere-associated protein RIF1 | RIF1 | 0.11* | 0.18* | 0.18* | 0.26* | 0.35* | 0.29* | 0.28* |
| Mitochondrial ribosome-associated GTPase 2 | MTG2 | 0.63* | 0.83 | 0.94 | 0.26* | 0.29* | 0.28* | 0.25* |
| Calcium/calmodulin-dependent protein kinase type II subunit beta | CAMK2B | 0.12* | 0.10* | 0.17* | 0.26* | 0.64* | 0.53* | 0.47* |
| Probable protein phosphatase 1N | PPM1N | 0.40* | 0.19* | 0.23* | 0.27* | 0.19* | 0.42* | 0.17* |
| Folylpolyglutamate synthase, mitochondrial | FPGS | 0.67* | 0.98 | 0.96 | 0.28* | 0.32* | 0.24* | 0.06* |
| C-X-C motif chemokine 14 | CXCL14 | 0.36* | 0.37* | 0.42* | 0.28* | 0.28* | 0.42* | 0.39* |
| SURP and G-patch domain-containing protein 2 | SUGP2 | 0.29* | 0.33* | 0.39* | 0.28* | 0.24* | 0.23* | 0.19* |
| Helicase-like transcription factor | HLTF | 0.34* | 0.41* | 0.37* | 0.28* | 0.52 | 0.63 | 0.51 |

| | | | | | | | | |
|---|---|---|---|---|---|---|---|---|
| Ribosomal biogenesis protein LAS1L | LAS1L | 0.22* | 0.26* | 0.25* | 0.28* | 0.62* | 0.51* | 0.40* |
| Dihydrofolate reductase | DHFR | 1.10 | 1.27 | 1.41 | 0.29* | 0.02* | 0.21* | 0.06* |
| Molybdenum cofactor sulfurase | MOCOS | 0.36* | 0.52* | 0.58* | 0.29* | 0.24* | 0.26* | 0.25* |
| Ras association domain-containing protein 6 | RASSF6 | 0.72* | 1.04 | 0.98 | 0.29* | 0.22* | 0.19* | 0.08* |
| AT-rich interactive domain-containing protein 2 | ARID2 | 0.24* | 0.38* | 0.30* | 0.30* | 0.48* | 0.35* | 0.48* |
| DNA topoisomerase 2-binding protein 1 | TOPBP1 | 0.34* | 0.50* | 0.60* | 0.31* | 0.44* | 0.44* | 0.51* |
| Inactive ubiquitin carboxyl-terminal hydrolase 53 | USP53 | 0.46* | 0.63* | 0.51* | 0.31* | 0.32* | 0.36* | 0.22* |
| Rab-like protein 2A | RABL2A | 0.67* | 0.64* | 0.77 | 0.32* | 0.45* | 0.43* | 0.28* |
| Serine/threonine-protein kinase 11-interacting protein | STK11IP | 0.63* | 0.66 | 0.79 | 0.32* | 0.11* | 0.24* | 0.36* |
| Zinc finger protein 592 | ZNF592 | 0.25* | 0.28* | 0.27* | 0.33* | 0.59 | 0.55 | 0.49* |
| Keratin, type II cytoskeletal 2 epidermal | KRT2 | 0.21* | 0.26* | 0.47* | 0.33* | 0.50* | 0.23* | 0.35* |
| Rho GTPase-activating protein 29 | ARHGAP29 | 0.19* | 0.35* | 0.41* | 0.33* | 0.58 | 0.50* | 0.35* |
| Circadian locomoter output cycles protein kaput | CLOCK | 0.43* | 0.48* | 0.56* | 0.34* | 0.60 | 0.53* | 0.46* |
| Structural maintenance of chromosomes protein 5 | SMC5 | 0.30* | 0.36* | 0.38* | 0.34* | 0.60 | 0.46* | 0.61* |
| Replication factor C subunit 2 | RFC2 | 0.59* | 0.62* | 0.63* | 0.34* | 0.52* | 0.54* | 0.44* |
| Gametogenetin-binding protein 2 | GGNBP2 | 0.34* | 0.37* | 0.50* | 0.34* | 0.51* | 0.46* | 0.40* |
| Deoxycytidine kinase | DCK | 0.40* | 0.45* | 0.56* | 0.34* | 0.63 | 0.57* | 0.54* |
| ATPase WRNIP1 | WRNIP1 | 0.62* | 0.60* | 0.63* | 0.35* | 0.39* | 0.35* | 0.38* |
| RING finger and CHY zinc finger domain-containing protein 1 | RCHY1 | 0.48* | 0.63* | 0.65* | 0.35* | 0.05* | 0.12* | 0.05* |
| AN1-type zinc finger protein 6 | ZFAND6 | 0.53* | 0.75* | 0.85 | 0.35* | 0.06* | 0.11* | 0.22* |
| Trafficking kinesin-binding protein 1 | TRAK1 | 0.29* | 0.38* | 0.48* | 0.36* | 0.46* | 0.61 | 0.58 |
| Acyl-CoA (8-3)-desaturase | FADS1 | 0.67* | 0.65* | 0.70* | 0.36* | 0.52* | 0.44* | 0.41* |
| NADPH oxidase organizer 1 | NOXO1 | 0.60* | 0.72 | 0.81 | 0.36* | 0.61 | 0.52 | 0.31* |
| Zinc finger FYVE domain-containing protein 26 | ZFYVE26 | 0.41* | 0.71 | 0.58* | 0.36* | 0.55 | 0.65 | 0.44* |
| Actin filament-associated protein 1-like 2 | AFAP1L2 | 0.38* | 0.66 | 0.77 | 0.37* | 0.13* | 0.36* | 0.30* |
| Vam6/Vps39-like protein | VPS39 | 0.50* | 0.46* | 0.46* | 0.37* | 0.63 | 0.54* | 0.58* |
| Dephospho-CoA kinase domain-containing protein | DCAKD | 0.87 | 0.98 | 0.96 | 0.38* | 0.35* | 0.42* | 0.20* |
| Serine/threonine-protein kinase Chk2 | CHEK2 | 0.56* | 0.63* | 0.61* | 0.38* | 0.31* | 0.25* | 0.25* |
| Nuclear receptor corepressor 1 | NCOR1 | 0.36* | 0.52* | 0.51* | 0.38* | 0.32* | 0.33* | 0.51* |
| Tubulin alpha chain-like 3 | TUBAL3 | 0.66* | 0.67* | 0.56* | 0.39* | 0.33* | 0.45* | 0.17* |
| Ankyrin repeat domain-containing protein SOWAHB | SOWAHB | 0.35* | 0.73 | 0.67 | 0.39* | 0.53 | 0.48* | 0.46* |
| Ly6/PLAUR domain-containing protein 8 | LYPD8 | 1.39 | 0.75 | 0.60* | 0.39* | 0.50* | 0.41* | 0.34* |
| Peroxisomal ATPase PEX1 | PEX1 | 0.56* | 0.64* | 0.64* | 0.39* | 0.43* | 0.48* | 0.31* |
| OTU domain-containing protein 3 | OTUD3 | 0.60* | 0.71 | 0.81 | 0.39* | 0.22* | 0.16* | 0.43* |
| Coiled-coil domain-containing protein 85C | CCDC85C | 0.61* | 0.72* | 0.70* | 0.39* | 0.15* | 0.22* | 0.24* |
| NF-X1-type zinc finger protein NFXL1 | NFXL1 | 0.50* | 0.58* | 0.58* | 0.40* | 0.39* | 0.35* | 0.40* |
| Survival motor neuron protein | SMN1 | 0.18* | 0.25* | 0.29* | 0.40* | 0.38* | 0.38* | 0.43* |

| | | | | | | | | |
|---|---|---|---|---|---|---|---|---|
| Lysine-specific demethylase 4B | KDM4B | 0.84 | 0.89 | 0.79 | 0.40* | 0.30* | 0.24* | 0.20* |
| Splicing regulator ARVCF | ARVCF | 0.58* | 0.91 | 0.98 | 0.40* | 0.46* | 0.37* | 0.29* |
| Dual specificity mitogen-activated protein kinase kinase 5 | MAP2K5 | 0.80 | 1.13 | 0.88 | 0.40* | 0.51* | 0.46* | 0.42* |
| E3 ubiquitin-protein ligase Midline-1 | MID1 | 0.32* | 0.37* | 0.40* | 0.41* | 0.38* | 0.40* | 0.35* |
| Beclin-1 | BECN1 | 0.36* | 0.54* | 0.52* | 0.41* | 0.67 | 0.66 | 0.61* |
| Trefoil factor 2 | TFF2 | 1.26 | 0.61* | 0.60* | 0.42* | 0.59 | 0.34* | 0.33* |
| Eukaryotic translation initiation factor 4E transporter | EIF4ENIF1 | 0.69* | 0.95 | 0.98 | 0.42* | 0.18* | 0.18* | 0.11* |
| E3 ubiquitin-protein ligase makorin-2 | MKRN2 | 0.59* | 0.67 | 0.65* | 0.42* | 0.60 | 0.52* | 0.45* |
| High affinity cAMP-specific and IBMX-insensitive 3',5'-cyclic phosphodiesterase 8A | PDE8A | 0.67 | 1.00 | 1.06 | 0.42* | 0.23* | 0.44* | 0.20* |
| Rho GTPase-activating protein 32 | ARHGAP32 | 0.53* | 0.71 | 0.65* | 0.42* | 0.52* | 0.63 | 0.46* |
| Epithelial splicing regulatory protein 2 | ESRP2 | 0.58* | 0.58* | 0.66* | 0.42* | 0.50* | 0.38* | 0.45* |
| Probable ATP-dependent RNA helicase DDX56 | DDX56 | 0.48* | 0.51* | 0.57* | 0.42* | 0.34* | 0.34* | 0.30* |
| S1 RNA-binding domain-containing protein 1 | SRBD1 | 0.60* | 0.53* | 0.64* | 0.43* | 0.58 | 0.40* | 0.23* |
| Ubiquitin-associated and SH3 domain-containing protein B | UBASH3B | 0.46* | 0.57* | 0.63* | 0.43* | 0.52* | 0.51* | 0.44* |
| Protein SSXT | SS18 | 0.94 | 0.98 | 0.94 | 0.43* | 0.17* | 0.37* | 0.57* |
| Bromodomain adjacent to zinc finger domain protein 1A | BAZ1A | 0.43* | 0.62* | 0.56* | 0.43* | 0.58 | 0.43* | 0.43* |
| RalBP1-associated Eps domain-containing protein 2 | REPS2 | 0.62* | 0.72 | 0.76 | 0.43* | 0.43* | 0.24* | 0.25* |
| BAH and coiled-coil domain-containing protein 1 | BAHCC1 | 0.54* | 1.51* | 0.94 | 0.43* | 0.49* | 0.40* | 0.22* |
| GTPase IMAP family member 2 | GIMAP2 | 0.71 | 0.78 | 0.85 | 0.43* | 0.47* | 0.58 | 0.26* |
| Leydig cell tumor 10 kDa protein homolog | C19orf53 | 0.21* | 0.19* | 0.20* | 0.43* | 0.37* | 0.36* | 0.45* |
| Protein Churchill | CHURC1 | 0.97 | 1.48* | 1.21 | 0.43* | 0.23* | 0.14* | 0.19* |
| Death domain-associated protein 6 | DAXX | 0.67* | 0.81 | 0.80 | 0.44* | 0.47* | 0.33* | 0.30* |
| DNA-directed RNA polymerases I and III subunit RPAC2 | POLR1D | 0.68* | 0.68 | 0.74 | 0.44* | 0.44* | 0.49* | 0.46* |
| DmX-like protein 2 | DMXL2 | 0.50* | 0.46* | 0.41* | 0.44* | 0.53* | 0.52* | 0.44* |
| Zinc finger CCCH domain-containing protein 8 | ZC3H8 | 0.48* | 0.52* | 0.58* | 0.44* | 0.51* | 0.43* | 0.45* |
| Uridine-cytidine kinase 2 | UCK2 | 0.65* | 0.69 | 0.67* | 0.44* | 0.39* | 0.36* | 0.34* |
| Putative GTP-binding protein 6 | GTPBP6 | 0.69 | 0.83 | 0.91 | 0.44* | 0.38* | 0.36* | 0.12* |
| HAUS augmin-like complex subunit 3 | HAUS3 | 0.35* | 0.39* | 0.40* | 0.45* | 0.66 | 0.62 | 0.59 |
| Pre-mRNA-splicing factor SLU7 | SLU7 | 0.69* | 0.95 | 0.95 | 0.45* | 0.61 | 0.60 | 0.51* |
| Uncharacterized protein C2orf42 | C2orf42 | 0.42* | 0.67* | 0.53* | 0.45* | 0.41* | 0.45* | 0.28* |
| Inositol hexakisphosphate kinase 1 | IP6K1 | 0.78 | 0.89 | 0.90 | 0.45* | 0.22* | 0.26* | 0.26* |
| Glutathione S-transferase A1 | GSTA1 | 0.72* | 0.38* | 0.47* | 0.45* | 0.49* | 0.64 | 0.41* |
| Zinc finger protein 701 | ZNF701 | 0.43* | 0.34* | 0.36* | 0.45* | 0.46* | 0.42* | 0.48* |
| Eukaryotic translation initiation factor 2-alpha kinase 3 | EIF2AK3 | 0.79 | 0.80 | 0.95 | 0.46* | 0.42* | 0.54 | 0.19* |

| | | | | | | | | |
|---|---|---|---|---|---|---|---|---|
| Neurabin-1 | PPP1R9A | 0.47* | 0.58* | 0.64* | 0.46* | 0.49* | 0.39* | 0.47* |
| A-kinase anchor protein 8-like | AKAP8L | 0.64* | 0.71 | 0.65* | 0.46* | 0.25* | 0.36* | 0.45* |
| SOSS complex subunit C | INIP | 0.86 | 0.86 | 0.75* | 0.46* | 0.17* | 0.19* | 0.05* |
| Pre-mRNA-splicing factor CWC22 homolog | CWC22 | 0.81 | 1.00 | 0.97 | 0.46* | 0.27* | 0.32* | 0.40* |
| Activating transcription factor 7-interacting protein 1 | ATF7IP | 0.27* | 0.43* | 0.38* | 0.47* | 0.46* | 0.50* | 0.46* |
| RRP12-like protein | RRP12 | 0.47* | 0.46* | 0.54* | 0.47* | 0.45* | 0.46* | 0.49* |
| Zinc finger and BTB domain-containing protein 11 | ZBTB11 | 0.25* | 0.37* | 0.31* | 0.47* | 0.42* | 0.29* | 0.49* |
| Caspase-2 | CASP2 | 0.87 | 1.00 | 1.01 | 0.47* | 0.40* | 0.49* | 0.29* |
| Interferon regulatory factor 2-binding protein 1 | IRF2BP1 | 1.00 | 0.92 | 0.82 | 0.47* | 0.50* | 0.43* | 0.40* |
| Echinoderm microtubule-associated protein-like 3 | EML3 | 0.32* | 0.50* | 0.48* | 0.47* | 0.21* | 0.10* | 0.30* |
| Protein FAM110B | FAM110B | 0.57* | 0.56* | 0.69* | 0.47* | 0.56* | 0.52* | 0.45* |
| Serine/threonine-protein phosphatase 6 regulatory subunit 2 | PPP6R2 | 0.61* | 0.71* | 0.58* | 0.47* | 0.49* | 0.54* | 0.32* |
| Ribosomal RNA-processing protein 7 homolog A | RRP7A | 0.57* | 0.49* | 0.52* | 0.47* | 0.39* | 0.41* | 0.46* |
| Neural proliferation differentiation and control protein 1 | NPDC1 | 0.46* | 0.76 | 0.88 | 0.47* | 0.26* | 0.24* | 0.25* |
| Protein polybromo-1 | PBRM1 | 0.57* | 0.81 | 0.74* | 0.47* | 0.32* | 0.33* | 0.27* |
| Dynein regulatory complex protein 10 | IQCD | 0.69 | 2.52* | 1.32 | 0.48* | 0.17* | 0.13* | 0.09* |
| DNA-binding protein SATB2 | SATB2 | 0.51* | 0.92 | 0.75* | 0.48* | 0.45* | 0.42* | 0.63* |
| Ras-related protein Rab-13 | RAB13 | 1.05 | 0.65* | 0.98 | 0.48* | 0.46* | 0.39* | 0.43* |
| Guanine nucleotide-binding protein subunit beta-like protein 1 | GNB1L | 0.54* | 0.61* | 0.64* | 0.48* | 0.50* | 0.58 | 0.57 |
| Protein DENND6B | DENND6B | 0.44* | 1.30 | 0.61* | 0.48* | 0.07* | 0.15* | 0.24* |
| E3 ubiquitin-protein ligase RNF113A | RNF113A | 0.51* | 0.59* | 0.63* | 0.48* | 0.18* | 0.21* | 0.31* |
| Neurogenic locus notch homolog protein 1 | NOTCH1 | 0.46* | 0.54* | 0.85 | 0.49* | 0.63 | 0.65 | 0.50* |
| Biogenesis of lysosome-related organelles complex 1 subunit 3 | BLOC1S3 | 0.30* | 0.42* | 0.36* | 0.49* | 0.50* | 0.39* | 0.62 |
| MAP kinase-activated protein kinase 5 | MAPKAPK5 | 0.66* | 0.99 | 0.88 | 0.49* | 0.63 | 0.44* | 0.45* |
| E3 ubiquitin-protein ligase TRIM36 | TRIM36 | 0.58* | 0.75 | 0.66* | 0.49* | 0.45* | 0.50* | 0.27* |
| Bifunctional polynucleotide phosphatase/kinase | PNKP | 0.87 | 1.01 | 0.99 | 0.49* | 0.66 | 0.45* | 0.56* |
| NFX1-type zinc finger-containing protein 1 | ZNFX1 | 0.46* | 0.56* | 0.59* | 0.49* | 0.55* | 0.63 | 0.56* |
| Synaptojanin-2 | SYNJ2 | 0.61* | 0.68* | 0.76 | 0.49* | 0.41* | 0.32* | 0.33* |
| Coiled-coil domain-containing protein 91 | CCDC91 | 0.46* | 0.97 | 0.95 | 0.49* | 0.54* | 0.49* | 0.49* |
| Breast cancer anti-estrogen resistance protein 3 | BCAR3 | 0.89 | 1.15 | 1.03 | 0.49* | 0.47* | 0.51 | 0.21* |
| NCK-interacting protein with SH3 domain | NCKIPSD | 0.79 | 0.71 | 0.75 | 0.50* | 0.50* | 0.44* | 0.28* |
| Probable ribosome biogenesis protein RLP24 | RSL24D1 | 0.58* | 0.47* | 0.54* | 0.50* | 0.22* | 0.21* | 0.27* |
| Lipid droplet assembly factor 1 | LDAF1 | 0.67* | 0.75 | 0.66 | 0.50* | 0.54 | 0.57 | 0.41* |
| A-kinase anchor protein 8 | AKAP8 | 0.63* | 0.53* | 0.59* | 0.50* | 0.42* | 0.47* | 0.67 |
| Aldo-keto reductase family 1 member C2 | AKR1C2 | 1.21 | 1.04 | 0.87 | 0.50* | 0.61 | 0.49* | 0.66 |

| | | | | | | | | |
|---|---|---|---|---|---|---|---|---|
| Nucleus accumbens-associated protein 1 | NACC1 | 0.48* | 0.68* | 0.69* | 0.50* | 0.31* | 0.32* | 0.34* |
| TATA-binding protein-associated factor 172 | BTAF1 | 0.50* | 0.62* | 0.58* | 0.51* | 0.58 | 0.54 | 0.52* |
| Transmembrane and coiled-coil domain-containing protein 3 | TMCO3 | 0.76 | 1.05 | 1.00 | 0.51* | 0.54 | 0.66 | 0.42* |
| Proteasome subunit beta type-6 | PSMB6 | 0.97 | 1.02 | 0.79 | 0.51* | 0.44* | 0.52* | 0.40* |
| F-box only protein 44 | FBXO44 | 1.23 | 1.11 | 0.97 | 0.51* | 0.38* | 0.61 | 0.44* |
| Activator of basal transcription 1 | ABT1 | 0.75* | 0.79 | 0.72* | 0.51* | 0.42* | 0.47* | 0.48* |
| Dysferlin | DYSF | 0.78* | 0.53* | 0.82 | 0.51* | 0.36* | 0.42* | 0.35* |
| Na(+)/H(+) exchange regulatory cofactor NHE-RF4 | NHERF4 | 0.90 | 0.93 | 0.78 | 0.51* | 0.46* | 0.43* | 0.33* |
| BLOC-1-related complex subunit 6 | BORCS6 | 1.11 | 0.98 | 1.00 | 0.51* | 0.06* | 0.30* | 0.27* |
| Probable ATP-dependent RNA helicase DDX52 | DDX52 | 0.71* | 0.73 | 0.79 | 0.51* | 0.59* | 0.56* | 0.41* |
| Serine/threonine-protein kinase MRCK alpha | CDC42BPA | 0.58* | 0.63* | 0.57* | 0.52* | 0.64 | 0.58* | 0.40* |
| Mitotic deacetylase-associated SANT domain protein | MIDEAS | 0.42* | 0.58* | 0.54* | 0.52* | 0.31* | 0.47* | 0.43* |
| Methionyl-tRNA formyltransferase, mitochondrial | MTFMT | 0.82 | 0.84 | 0.84 | 0.52* | 0.18* | 0.34* | 0.16* |
| Dynamin-binding protein | DNMBP | 0.59* | 0.48* | 0.50* | 0.52* | 0.66 | 0.52 | 0.46* |
| Protein CDV3 homolog | CDV3 | 0.77 | 0.80 | 0.88 | 0.52* | 0.43* | 0.46* | 0.52* |
| ADP-ribosylation factor-like protein 6 | ARL6 | 0.78 | 0.84 | 0.91 | 0.52* | 0.36* | 0.42* | 0.43* |
| Ral GTPase-activating protein subunit alpha-2 | RALGAPA2 | 0.47* | 0.70 | 0.69* | 0.52* | 0.48* | 0.38* | 0.37* |
| Fatty acyl-CoA reductase 2 | FAR2 | 0.82 | 0.49* | 0.55* | 0.52* | 0.11* | 0.25* | 0.27* |
| RNA 3'-terminal phosphate cyclase-like protein | RCL1 | 0.68* | 0.71* | 0.73* | 0.52* | 0.56* | 0.56* | 0.58* |
| Ubiquitin-conjugating enzyme E2 Q1 | UBE2Q1 | 0.72* | 0.69* | 0.69* | 0.53* | 0.52* | 0.54* | 0.38* |
| Alpha-endosulfine | ENSA | 0.47* | 0.44* | 0.55* | 0.53* | 0.36* | 0.35* | 0.55* |
| Interferon regulatory factor 2 | IRF2 | 0.72* | 1.02 | 1.01 | 0.53* | 0.22* | 0.21* | 0.29* |
| Serine/threonine-protein kinase tousled-like 2 | TLK2 | 0.39* | 0.59* | 0.61* | 0.53* | 0.32* | 0.30* | 0.34* |
| ATPase family gene 2 protein homolog B | AFG2B | 0.47* | 0.50* | 0.62* | 0.54* | 0.54 | 0.54 | 0.60 |
| Coiled-coil domain-containing protein 28A | CCDC28A | 0.96 | 0.91 | 1.00 | 0.54* | 0.58 | 0.53 | 0.53* |
| APOBEC1 complementation factor | A1CF | 1.14 | 1.10 | 1.03 | 0.54* | 0.28* | 0.54 | 0.03* |
| Uncharacterized protein C9orf85 | C9orf85 | 0.49* | 0.66 | 0.86 | 0.54* | 0.21* | 0.17* | 0.36* |
| Serine/threonine-protein kinase Sgk2 | SGK2 | 0.83 | 0.88 | 0.66* | 0.55* | 0.63 | 0.60* | 0.50* |
| Aldo-keto reductase family 1 member B10 | AKR1B10 | 1.02 | 0.84 | 0.74* | 0.55* | 0.62* | 0.50* | 0.50* |
| Transmembrane protein 201 | TMEM201 | 0.55* | 0.68* | 0.64* | 0.55* | 0.22* | 0.38* | 0.44* |
| Calcium-binding and coiled-coil domain-containing protein 2 | CALCOCO2 | 0.47* | 0.60* | 0.63* | 0.55* | 0.31* | 0.29* | 0.33* |
| Guanine nucleotide-binding protein-like 3 | GNL3 | 0.66* | 0.68 | 0.79 | 0.55* | 0.56 | 0.46* | 0.47* |
| Periodic tryptophan protein 2 homolog | PWP2 | 0.60* | 0.58* | 0.67* | 0.55* | 0.61* | 0.61 | 0.60* |
| Proteasome subunit beta type-7 | PSMB7 | 0.97 | 0.93 | 0.85 | 0.55* | 0.51* | 0.55* | 0.47* |
| tRNA (uracil-5-)-methyltransferase homolog A | TRMT2A | 0.55* | 0.50* | 0.52* | 0.55* | 0.58 | 0.55* | 0.62 |
| 3-hydroxy-3-methylglutaryl-coenzyme A reductase | HMGCR | 0.62* | 0.67 | 0.59* | 0.55* | 0.62 | 0.57 | 0.53* |

| | | | | | | | | |
|---|---|---|---|---|---|---|---|---|
| SH3 and PX domain-containing protein 2B | SH3PXD2B | 0.86 | 1.06 | 1.09 | 0.55* | 0.37* | 0.41* | 0.59* |
| Kinetochore-associated protein NSL1 homolog | NSL1 | 0.53* | 0.63* | 0.65* | 0.56* | 0.45* | 0.26* | 0.50* |
| Dysbindin | DTNBP1 | 0.83 | 0.78 | 0.88 | 0.56* | 0.64 | 0.53 | 0.55 |
| Inactive ubiquitin thioesterase OTULINL | OTULINL | 0.97 | 0.94 | 0.77 | 0.56* | 0.55* | 0.65 | 0.35* |
| Trypsin-3 | PRSS3 | 1.35 | 0.78 | 1.47 | 0.56* | 0.60 | 0.49* | 0.62 |
| Synergin gamma | SYNRG | 0.65* | 0.69 | 0.84 | 0.56* | 0.42* | 0.37* | 0.55 |
| Zinc finger and BTB domain-containing protein 7A | ZBTB7A | 0.57* | 0.54* | 0.56* | 0.56* | 0.52* | 0.45* | 0.56* |
| tRNA-dihydrouridine(20) synthase [NAD(P)+]-like | DUS2 | 0.80* | 0.85 | 0.93 | 0.56* | 0.41* | 0.35* | 0.37* |
| Phosphatase and actin regulator 2 | PHACTR2 | 0.89 | 0.87 | 1.18 | 0.56* | 0.04* | 0.47* | 0.22* |
| Coiled-coil domain-containing protein 97 | CCDC97 | 0.49* | 0.65 | 1.06 | 0.56* | 0.43* | 0.38* | 0.55 |
| NFATC2-interacting protein | NFATC2IP | 0.85 | 0.85 | 1.09 | 0.56* | 0.21* | 0.20* | 0.18* |
| Tetratricopeptide repeat protein 12 | TTC12 | 0.64* | 0.64* | 0.64* | 0.57* | 0.54* | 0.51* | 0.44* |
| Adipogenesis regulatory factor | ADIRF | 1.03 | 0.94 | 1.13* | 0.57* | 0.51* | 0.53* | 0.62* |
| Transcription factor Sp3 | SP3 | 0.82 | 0.80 | 0.85 | 0.57* | 0.39* | 0.63 | 0.63 |
| Phosphorylase b kinase gamma catalytic chain, liver/testis isoform | PHKG2 | 0.73 | 0.76 | 0.62* | 0.58* | 0.34* | 0.52 | 0.30* |
| Mitochondrial mRNA pseudouridine synthase RPUSD3 | RPUSD3 | 0.99 | 1.04 | 1.05 | 0.58* | 0.24* | 0.31* | 0.18* |
| Zinc finger protein 703 | ZNF703 | 0.68 | 0.86 | 0.91 | 0.58* | 0.54 | 0.33* | 0.54 |
| Ribosome quality control complex subunit TCF25 | TCF25 | 0.76* | 0.96 | 0.83 | 0.58* | 0.49* | 0.59* | 0.41* |
| Constitutive coactivator of peroxisome proliferator-activated receptor gamma | FAM120B | 0.69 | 0.77 | 0.74 | 0.58* | 0.59 | 0.59 | 0.64 |
| Protein Wiz | WIZ | 0.43* | 0.61* | 0.61* | 0.58* | 0.15* | 0.17* | 0.32* |
| Transcriptional repressor protein YY1 | YY1 | 0.68* | 0.92 | 0.97 | 0.59* | 0.40* | 0.51* | 0.57* |
| BRCA1-associated protein | BRAP | 0.58* | 0.73 | 0.74 | 0.59* | 0.33* | 0.42* | 0.44* |
| Mitochondrial amidoxime-reducing component 1 | MTARC1 | 1.04 | 0.96 | 1.13 | 0.59* | 0.21* | 0.41* | 0.49* |
| Ubiquitin-associated protein 2 | UBAP2 | 0.54* | 0.90 | 1.00 | 0.59* | 0.07* | 0.16* | 0.10* |
| Signal-induced proliferation-associated 1-like protein 3 | SIPA1L3 | 0.73* | 0.82 | 0.70* | 0.59* | 0.51* | 0.48* | 0.44* |
| Ubiquitin-associated protein 1 | UBAP1 | 0.75 | 0.91 | 0.87 | 0.59* | 0.25* | 0.55 | 0.49* |
| Exopolyphosphatase PRUNE1 | PRUNE1 | 0.77 | 0.86 | 0.83 | 0.59* | 0.58 | 0.58 | 0.49* |
| tRNA (guanine(6)-N2)-methyltransferase THUMP3 | THUMPD3 | 0.84 | 0.71* | 0.79 | 0.59* | 0.36* | 0.41* | 0.40* |
| Zinc finger CCHC domain-containing protein 8 | ZCCHC8 | 0.56* | 0.55* | 0.56* | 0.60* | 0.26* | 0.25* | 0.39* |
| Cell division cycle protein 123 homolog | CDC123 | 0.72* | 0.81 | 0.88 | 0.60* | 0.49* | 0.57* | 0.44* |
| DNA ligase 3 | LIG3 | 0.85 | 0.86 | 0.93 | 0.60* | 0.48* | 0.62* | 0.56* |
| Nucleolar MIF4G domain-containing protein 1 | NOM1 | 0.38* | 0.40* | 0.40* | 0.60* | 0.37* | 0.27* | 0.32* |
| Pyrin | MEFV | 0.88 | 0.88 | 1.30 | 0.60* | 0.60 | 0.51* | 0.60* |
| Paladin | PALD1 | 0.51* | 0.59* | 0.69* | 0.60* | 0.46* | 0.56* | 0.54* |
| Acyl-CoA-binding domain-containing protein 4 | ACBD4 | 1.11 | 1.04 | 0.85 | 0.61* | 0.56* | 0.54* | 0.39* |
| Actin filament-associated protein 1 | AFAP1 | 0.66 | 1.00 | 0.86 | 0.61* | 0.41* | 0.40* | 0.55 |

| | | | | | | | | |
|---|---|---|---|---|---|---|---|---|
| ATP-dependent RNA helicase DDX3Y | DDX3Y | 0.68 | 0.66 | 0.87 | 0.61* | 0.51* | 0.52* | 0.55* |
| Trefoil factor 1 | TFF1 | 1.54* | 1.08 | 0.91 | 0.61* | 0.63* | 0.48* | 0.37* |
| Butyrophilin-like protein 8 | BTNL8 | 0.98 | 1.50* | 1.43* | 0.61* | 0.47* | 0.65* | 0.47* |
| Arf-GAP domain and FG repeat-containing protein 2 | AGFG2 | 0.81 | 0.62* | 0.76 | 0.61* | 0.61 | 0.45* | 0.54 |
| Kelch repeat and BTB domain-containing protein 11 | KBTBD11 | 0.89 | 0.77 | 0.94 | 0.61* | 0.58 | 0.54 | 0.63 |
| FLYWCH family member 2 | FLYWCH2 | 1.10 | 1.08 | 1.06 | 0.61* | 0.19* | 0.22* | 0.18* |
| Ribosomal protein S6 kinase alpha-4 | RPS6KA4 | 0.69* | 0.47* | 0.63* | 0.61* | 0.49* | 0.63 | 0.55* |
| RNA-binding protein NOB1 | NOB1 | 0.59* | 0.55* | 0.71* | 0.61* | 0.48* | 0.50* | 0.54* |
| Transmembrane channel-like protein 6 | TMC6 | 0.59* | 0.47* | 0.51* | 0.62* | 0.63 | 0.64 | 0.61 |
| Protein FAM83F | FAM83F | 0.88 | 0.91 | 0.86 | 0.62* | 0.54* | 0.54* | 0.48* |
| E3 ubiquitin-protein ligase BRE1B | RNF40 | 0.62* | 0.66* | 0.61* | 0.62* | 0.57* | 0.49* | 0.64 |
| DNA/RNA-binding protein KIN17 | KIN | 0.94 | 1.01 | 1.03 | 0.62* | 0.12* | 0.36* | 0.39* |
| RNA polymerase II subunit A C-terminal domain phosphatase | CTDP1 | 0.63* | 0.67* | 0.70* | 0.62* | 0.63 | 0.55* | 0.62 |
| EKC/KEOPS complex subunit LAGE3 | LAGE3 | 0.82 | 1.02 | 1.07 | 0.62* | 0.23* | 0.19* | 0.31* |
| Peptidyl-prolyl cis-trans isomerase-like 4 | PPIL4 | 1.02 | 0.84 | 0.74 | 0.62* | 0.23* | 0.22* | 0.30* |
| Zinc finger C2HC domain-containing protein 1A | ZC2HC1A | 0.63* | 0.65* | 0.81 | 0.62* | 0.45* | 0.51* | 0.54* |
| Neurabin-2 | PPP1R9B | 0.74 | 0.80 | 0.85 | 0.62* | 0.62 | 0.60 | 0.57 |
| Dematin | DMTN | 0.78 | 1.09 | 1.13 | 0.62* | 0.06* | 0.12* | 0.23* |
| Protein FAM13A | FAM13A | 0.92 | 1.01 | 0.96 | 0.63* | 0.33* | 0.35* | 0.40* |
| Docking protein 1 | DOK1 | 0.79 | 0.84 | 0.98 | 0.63* | 0.38* | 0.46* | 0.44* |
| Diacylglycerol kinase theta | DGKQ | 0.66 | 1.19 | 0.97 | 0.63* | 0.46* | 0.64 | 0.53* |
| Tripartite motif-containing protein 26 | TRIM26 | 0.56* | 0.73* | 0.71* | 0.63* | 0.53* | 0.56* | 0.62* |
| Chromobox protein homolog 8 | CBX8 | 0.49* | 0.66* | 0.71* | 0.63* | 0.26* | 0.20* | 0.51* |
| Ras GTPase-activating-like protein IQGAP2 | IQGAP2 | 0.95 | 0.78* | 0.90 | 0.63* | 0.59* | 0.59* | 0.59* |
| 5'-AMP-activated protein kinase subunit beta-2 | PRKAB2 | 0.76* | 1.08 | 1.05 | 0.63* | 0.41* | 0.60 | 0.58* |
| Thiol S-methyltransferase TMT1B | TMT1B | 0.95 | 0.99 | 0.81 | 0.63* | 0.66 | 0.63 | 0.56* |
| Inositol 1,4,5-trisphosphate receptor type 2 | ITPR2 | 0.94 | 0.85 | 0.65* | 0.64 | 0.58 | 0.66 | 0.55 |
| Zinc finger protein 768 | ZNF768 | 0.73 | 0.67* | 0.83 | 0.64* | 0.19* | 0.28* | 0.47* |
| Breast carcinoma-amplified sequence 1 | BCAS1 | 0.92 | 0.79 | 1.02 | 0.64* | 0.57* | 0.51* | 0.59* |
| Translation initiation factor eIF-2B subunit delta | EIF2B4 | 0.62* | 0.59* | 0.55* | 0.64* | 0.57* | 0.57* | 0.61* |
| Uncharacterized protein C2orf72 | C2orf72 | 1.13 | 0.75 | 0.66* | 0.64* | 0.05* | 0.39* | 0.08* |
| Remodeling and spacing factor 1 | RSF1 | 0.54* | 0.63* | 0.57* | 0.64* | 0.40* | 0.41* | 0.53* |
| La-related protein 1B | LARP1B | 0.95 | 0.73 | 0.88 | 0.64 | 0.59 | 0.59 | 0.59 |
| Motile sperm domain-containing protein 1 | MOSPD1 | 1.07 | 1.64* | 1.30 | 0.65 | 0.27* | 0.58 | 0.13* |
| Large subunit GTPase 1 homolog | LSG1 | 0.76* | 0.64* | 0.66* | 0.65* | 0.54* | 0.52* | 0.52* |
| Proline and serine-rich protein 2 | PROSER2 | 0.90 | 0.94 | 1.01 | 0.65 | 0.32* | 0.42* | 0.44* |
| Transcription termination factor 3, mitochondrial | MTERF3 | 1.00 | 1.19 | 1.30* | 0.65* | 0.24* | 0.33* | 0.40* |

| | | | | | | | | |
|---|---|---|---|---|---|---|---|---|
| Nonsense-mediated mRNA decay factor SMG9 | SMG9 | 0.69* | 0.82 | 0.91 | 0.65* | 0.33* | 0.55* | 0.61 |
| Protein furry homolog | FRY | 0.34* | 0.64* | 0.59* | 0.65 | 0.30* | 0.36* | 0.55 |
| Zinc finger protein 346 | ZNF346 | 0.71 | 0.71 | 0.77 | 0.65 | 0.14* | 0.30* | 0.51* |
| Coiled-coil domain-containing protein 43 | CCDC43 | 1.16 | 1.24 | 1.32 | 0.65 | 0.08* | 0.31* | 0.22* |
| Rab5 GDP/GTP exchange factor | RABGEF1 | 0.67* | 0.92 | 0.93 | 0.65 | 0.50* | 0.62 | 0.53 |
| tRNA N(3)-methylcytidine methyltransferase METTL2B | METTL2B | 0.79* | 0.90 | 0.86 | 0.65* | 0.30* | 0.36* | 0.31* |
| Elongator complex protein 4 | ELP4 | 0.60* | 0.73 | 0.75 | 0.66 | 0.60 | 0.53 | 0.49* |
| Glutathione S-transferase C-terminal domain-containing protein | GSTCD | 0.71* | 0.72* | 0.72* | 0.66* | 0.61* | 0.63 | 0.58* |
| Transcription elongation factor A protein 3 | TCEA3 | 0.93 | 1.00 | 0.97 | 0.66 | 0.33* | 0.51* | 0.60 |
| DNA-directed RNA polymerase I subunit RPA49 | POLR1E | 0.80 | 0.77 | 0.76 | 0.66 | 0.64 | 0.64 | 0.55 |
| SH2 domain-containing protein 3A | SH2D3A | 0.73* | 0.66* | 0.61* | 0.66* | 0.45* | 0.51* | 0.50* |
| Gastrotropin | FABP6 | 1.05 | 0.52* | 0.43* | 0.66* | 0.55* | 0.35* | 0.48* |
| Valine--tRNA ligase, mitochondrial | VARS2 | 1.06 | 0.83 | 0.74* | 0.66* | 0.41* | 0.44* | 0.30* |
| Regenerating islet-derived protein 4 | REG4 | 0.73* | 0.56* | 0.98 | 0.66* | 0.45* | 0.49* | 0.52* |
| RNA polymerase-associated protein LEO1 | LEO1 | 0.98 | 1.02 | 0.90 | 0.67 | 0.22* | 0.41* | 0.30* |
| Dynein axonemal heavy chain 17 | DNAH17 | 1.74* | 2.19* | 1.86* | 0.67 | 0.06* | 0.20* | 0.23* |

***B. Down-regulated proteins unique to LPS-Cytokines (LPS-Cyto) [96 proteins]***

| | | Interventions | | | | | | |
|---|---|---|---|---|---|---|---|---|
| | | Control | | | *With LPS & Cytokines* | | | |
| Proteins | Genes | AQ | AQ+MES | MES | ***LPS-Cyto*** | *AQ* | *AQ+MES* | *MES* |
| Gamma-tubulin complex component 3 | TUBGCP3 | 0.19* | 0.27* | 0.29* | 0.31* | 0.86 | 0.80 | 0.82 |
| Mediator of RNA polymerase II transcription subunit 23 | MED23 | 0.28* | 0.39* | 0.32* | 0.32* | 0.85 | 0.69 | 0.70 |
| Integrator complex subunit 5 | INTS5 | 0.39* | 0.34* | 0.33* | 0.35* | 0.86 | 0.81 | 0.84 |
| Serine/threonine-protein kinase WNK2 | WNK2 | 0.36* | 0.74 | 0.58* | 0.39* | 0.72 | 0.77 | 0.79 |
| Histone-lysine N-methyltransferase EHMT2 | EHMT2 | 0.25* | 0.28* | 0.29* | 0.40* | 1.50 | 1.10 | 1.16 |
| Cyclin-dependent kinase 1 | CDK1 | 0.62* | 0.67* | 0.68* | 0.40* | 1.51 | 1.34 | 1.54* |
| Structural maintenance of chromosomes protein 6 | SMC6 | 0.28* | 0.28* | 0.27* | 0.41* | 1.10 | 0.92 | 1.02 |
| Repetin | RPTN | 1.60* | 1.00 | 4.84* | 0.42* | 2.18* | 0.67 | 3.40* |
| Mediator of RNA polymerase II transcription subunit 12 | MED12 | 0.25* | 0.27* | 0.31* | 0.42* | 0.97 | 0.81 | 0.84 |
| Small subunit processome component 20 homolog | UTP20 | 0.39* | 0.36* | 0.37* | 0.42* | 0.86 | 0.84 | 1.24 |
| Transmembrane protein 209 | TMEM209 | 0.34* | 0.43* | 0.44* | 0.42* | 1.00 | 0.80 | 0.87 |
| WD repeat-containing protein 3 | WDR3 | 0.37* | 0.34* | 0.36* | 0.43* | 0.82 | 0.73 | 0.70* |
| Keratin, type I cytoskeletal 23 | KRT23 | 2.31* | 1.81* | 6.31* | 0.44* | 2.65* | 0.70 | 2.04* |
| Pre-mRNA-processing factor 39 | PRPF39 | 0.97 | 1.06 | 0.98 | 0.46* | 0.82 | 0.98 | 0.80 |
| Endoribonuclease ZC3H12A | ZC3H12A | 0.56* | 0.69* | 0.73* | 0.46* | 0.80 | 0.84 | 0.70 |

| | | | | | | | | |
|---|---|---|---|---|---|---|---|---|
| Progesterone-induced-blocking factor 1 | PIBF1 | 0.51* | 0.67* | 0.65* | 0.46* | 0.97 | 1.00 | 0.67 |
| Structural maintenance of chromosomes protein 4 | SMC4 | 0.46* | 0.45* | 0.55* | 0.46* | 1.22 | 1.09 | 0.96 |
| FERM and PDZ domain-containing protein 1 | FRMPD1 | 0.54* | 2.61* | 2.58* | 0.47* | 0.94 | 1.85* | 2.22* |
| Arachidonate 12-lipoxygenase, 12R-type | ALOX12B | 2.22* | 1.37 | 7.85* | 0.47* | 2.58* | 0.92 | 1.08 |
| CDK5 and ABL1 enzyme substrate 1 | CABLES1 | 0.48* | 0.56* | 0.68* | 0.48* | 1.04 | 0.89 | 0.90 |
| Calpain-8 | CAPN8 | 0.70 | 0.94 | 0.81 | 0.48* | 0.91 | 0.83 | 0.88 |
| E3 ubiquitin-protein ligase TRIM32 | TRIM32 | 0.54* | 0.56* | 0.56* | 0.48* | 0.95 | 0.87 | 0.72 |
| Probable helicase with zinc finger domain | HELZ | 0.61* | 0.70 | 0.67* | 0.49* | 1.11 | 1.01 | 1.01 |
| Integrator complex subunit 7 | INTS7 | 0.46* | 0.58* | 0.57* | 0.49* | 1.12 | 0.88 | 0.83 |
| Alkaline phosphatase, placental type | ALPP | 0.98 | 0.67* | 0.93 | 0.51* | 1.19 | 1.61 | 1.36 |
| Probable ATP-dependent RNA helicase DDX20 | DDX20 | 0.33* | 0.54* | 0.49* | 0.51* | 0.75 | 0.70 | 0.73 |
| Kynureninase | KYNU | 0.82 | 1.00 | 1.61* | 0.52* | 0.92 | 1.21 | 1.74* |
| Protein PALS2 | PALS2 | 0.98 | 0.52* | 0.71 | 0.52* | 1.81* | 1.54 | 1.29 |
| Endoribonuclease Dicer | DICER1 | 0.58* | 0.75 | 0.84 | 0.53* | 0.94 | 0.86 | 0.85 |
| Retrotransposon Gag-like protein 8C | RTL8C | 0.91 | 1.18 | 1.12 | 0.53* | 0.73 | 0.85 | 0.85 |
| Anaphase-promoting complex subunit 5 | ANAPC5 | 0.39* | 0.38* | 0.39* | 0.53* | 1.56* | 1.01 | 1.02 |
| Probable ATP-dependent RNA helicase DDX10 | DDX10 | 0.89 | 0.78 | 0.82 | 0.53* | 0.89 | 0.84 | 0.75 |
| MYND-type zinc finger-containing chromatin reader ZMYND8 | ZMYND8 | 0.68 | 0.70 | 0.69 | 0.54* | 0.69 | 0.82 | 1.45 |
| BTB/POZ domain-containing protein KCTD3 | KCTD3 | 0.53* | 0.58* | 0.51* | 0.54* | 0.97 | 1.07 | 0.69 |
| PWWP domain-containing DNA repair factor 4 | PWWP4 | 1.54* | 1.13 | 1.24 | 0.54* | 1.27 | 1.13 | 0.67 |
| Transformation/transcription domain-associated protein | TRRAP | 0.30* | 0.35* | 0.40* | 0.54* | 1.21 | 1.16 | 1.04 |
| Protein piccolo | PCLO | 0.56* | 1.75* | 2.05* | 0.55* | 0.72 | 2.03* | 2.36* |
| Nitric oxide-associated protein 1 | NOA1 | 0.95 | 1.02 | 1.02 | 0.55* | 0.82 | 0.71 | 0.87 |
| UDP-N-acetylhexosamine pyrophosphorylase-like protein 1 | UAP1L1 | 0.69* | 0.81 | 0.74* | 0.55* | 1.08 | 0.98 | 0.90 |
| Microtubule-associated tumor suppressor 1 | MTUS1 | 0.55* | 0.71 | 0.59* | 0.56* | 0.91 | 0.83 | 0.68 |
| PH-interacting protein | PHIP | 0.81 | 0.72 | 0.81 | 0.56* | 0.82 | 0.81 | 0.68 |
| Baculoviral IAP repeat-containing protein 2 | BIRC2 | 0.41* | 0.58* | 0.49* | 0.57* | 1.37 | 1.08 | 1.31 |
| Target of EGR1 protein 1 | TOE1 | 0.59* | 0.60* | 0.69* | 0.57* | 0.94 | 0.84 | 0.79 |
| Cerebellin-4 | CBLN4 | 0.41* | 1.58 | 2.52* | 0.57* | 0.78 | 1.73 | 1.95* |
| TNF receptor-associated factor 2 | TRAF2 | 0.52* | 0.60* | 0.58* | 0.57* | 0.95 | 0.93 | 0.89 |
| Protein S100-A3 | S100A3 | 1.12 | 26.05* | 2.82* | 0.58* | 1.60 | 1.77 | 3.12* |
| Serine/threonine-protein kinase RIO3 | RIOK3 | 0.81 | 0.76 | 0.69 | 0.58* | 0.85 | 0.71 | 0.80 |
| Single-stranded DNA-binding protein 3 | SSBP3 | 0.56* | 0.77 | 0.66* | 0.58* | 1.24 | 1.30 | 1.34 |
| DNA mismatch repair protein Mlh3 | MLH3 | 1.28 | 0.99 | 1.18 | 0.58* | 1.07 | 0.82 | 0.87 |
| mRNA-decapping enzyme 1B | DCP1B | 1.26 | 2.01* | 1.92* | 0.58* | 1.35 | 0.76 | 1.52 |

| | | | | | | | | |
|---|---|---|---|---|---|---|---|---|
| Single-stranded DNA-binding protein 4 | SSBP4 | 0.65* | 0.96 | 0.80 | 0.58* | 1.08 | 1.15 | 1.07 |
| Desmoglein-3 | DSG3 | 0.68 | 3.24* | 4.23* | 0.59 | 0.94 | 1.57 | 2.39* |
| Serine/threonine-protein kinase 31 | STK31 | 2.24* | 1.32 | 4.57* | 0.59* | 2.06* | 0.73 | 1.99* |
| Ras-related protein Rab-6B | RAB6B | 1.14 | 0.98 | 0.99 | 0.59* | 0.74 | 0.70 | 0.73 |
| Integrator complex subunit 11 | INTS11 | 0.56* | 0.66* | 0.68* | 0.59* | 0.68 | 0.79 | 0.96 |
| Non-structural maintenance of chromosomes element 3 homolog | NSMCE3 | 0.56* | 0.62* | 0.61* | 0.60* | 0.87 | 0.88 | 0.88 |
| Anaphase-promoting complex subunit 1 | ANAPC1 | 0.33* | 0.29* | 0.32* | 0.60* | 2.35* | 1.97* | 2.10* |
| DNA-directed RNA polymerase II subunit RPB2 | POLR2B | 0.86 | 0.81 | 0.74 | 0.60* | 0.80 | 0.76 | 0.85 |
| ATP-dependent RNA helicase DDX19B | DDX19B | 1.02 | 1.19 | 1.08 | 0.60* | 0.92 | 1.22 | 0.87 |
| Sin3 histone deacetylase corepressor complex component SDS3 | SUDS3 | 0.45* | 0.57* | 0.52* | 0.61* | 0.92 | 0.93 | 0.83 |
| Histidine-rich glycoprotein | HRG | 0.76 | 1.27 | 4.14* | 0.61 | 0.90 | 1.03 | 1.39 |
| Unconventional myosin-IXb | MYO9B | 0.70* | 0.80 | 0.75* | 0.61* | 0.88 | 0.89 | 0.78 |
| Integrator complex subunit 4 | INTS4 | 0.39* | 0.43* | 0.46* | 0.61* | 1.10 | 1.22 | 1.14 |
| Pericentriolar material 1 protein | PCM1 | 0.68* | 0.68* | 0.65* | 0.61* | 0.75 | 0.76 | 0.69 |
| ER lumen protein-retaining receptor 1 | KDELR1 | 0.93 | 0.91 | 1.01 | 0.61* | 0.99 | 1.19 | 0.93 |
| Mediator of RNA polymerase II transcription subunit 22 | MED22 | 0.54* | 0.67 | 0.61* | 0.62* | 1.13 | 1.42 | 1.22 |
| Glycerol-3-phosphate acyltransferase 4 | GPAT4 | 1.16 | 1.21 | 1.18 | 0.63* | 0.96 | 0.87 | 0.70 |
| Serine/threonine-protein kinase PAK 5 | PAK5 | 1.18 | 0.85 | 0.91 | 0.63 | 0.94 | 1.28 | 0.83 |
| Ribonuclease P protein subunit p29 | POP4 | 0.51* | 0.61* | 0.62* | 0.63* | 1.31 | 1.21 | 1.20 |
| Cell adhesion molecule 1 | CADM1 | 0.65* | 1.89* | 2.04* | 0.63 | 0.89 | 1.76* | 2.25* |
| Replication factor C subunit 4 | RFC4 | 0.59* | 0.80 | 0.92 | 0.63* | 0.81 | 0.77 | 0.95 |
| Guanine nucleotide-binding protein subunit alpha-15 | GNA15 | 0.90 | 2.12* | 3.41* | 0.64 | 1.28 | 1.26 | 1.03 |
| DNA topoisomerase 3-beta-1 | TOP3B | 0.68 | 0.69 | 0.76 | 0.64* | 0.82 | 0.74 | 0.68 |
| Tetratricopeptide repeat protein 13 | TTC13 | 0.85 | 1.07 | 1.31 | 0.64 | 1.18 | 1.21 | 1.20 |
| Heat shock-related 70 kDa protein 2 | HSPA2 | 0.97 | 1.11 | 1.15 | 0.64* | 0.76 | 0.82 | 0.72 |
| Dermokine | DMKN | 0.47* | 0.70 | 0.58* | 0.64* | 1.58 | 0.79 | 1.33 |
| GEM-interacting protein | GMIP | 0.91 | 0.79 | 0.86 | 0.64 | 0.71 | 0.75 | 0.88 |
| Protein phosphatase 1 regulatory subunit 14C | PPP1R14C | 1.14 | 0.99 | 1.29 | 0.65 | 0.96 | 0.84 | 0.85 |
| RAB6A-GEF complex partner protein 2 | RGP1 | 0.41* | 0.67 | 0.93 | 0.65 | 0.92 | 1.04 | 0.97 |
| eIF-2-alpha kinase GCN2 | EIF2AK4 | 0.51* | 0.63 | 0.70 | 0.65 | 0.82 | 0.77 | 0.79 |
| A disintegrin and metalloproteinase with thrombospondin motifs 13 | ADAMTS13 | 0.68 | 2.03* | 2.82* | 0.65 | 0.85 | 2.28* | 2.94* |
| Mitochondrial antiviral-signaling protein | MAVS | 0.91 | 1.11 | 1.16 | 0.65* | 0.70 | 0.89 | 0.89 |
| Death-associated protein kinase 2 | DAPK2 | 0.99 | 0.95 | 0.94 | 0.65 | 1.31 | 0.93 | 0.96 |
| TBC1 domain family member 17 | TBC1D17 | 0.73 | 0.69 | 0.65* | 0.65* | 0.98 | 0.78 | 0.76 |
| B-cell lymphoma/leukemia 11B | BCL11B | 0.76 | 0.76 | 0.86 | 0.65 | 0.86 | 0.79 | 0.99 |

| | | | | | | | | |
|---|---|---|---|---|---|---|---|---|
| Keratin, type I cuticular Ha1 | KRT31 | 0.68* | 49.23* | 2.04* | 0.65* | 0.92 | 0.80 | 2.96* |
| Renin | REN | 1.34 | 1.56* | 1.57* | 0.66 | 0.96 | 0.87 | 0.83 |
| Myelin expression factor 2 | MYEF2 | 0.70* | 1.05 | 1.05 | 0.66* | 1.57 | 1.32 | 1.36 |
| DDB1- and CUL4-associated factor 6 | DCAF6 | 0.75 | 0.86 | 1.02 | 0.66 | 0.91 | 1.16 | 0.82 |
| Immunoglobulin heavy constant gamma 4 | IGHG4 | 0.64* | 1.95* | 32.94* | 0.66* | 0.72 | 1.67* | 0.96 |
| Leucine-rich repeat-containing protein 75A | LRRC75A | 0.79 | 1.07 | 1.06 | 0.66 | 0.68 | 0.75 | 0.72 |
| Retinoic acid receptor responder protein 2 | RARRES2 | 0.54* | 1.61* | 2.51* | 0.66 | 0.68 | 1.46 | 2.08* |
| 15-hydroxyprostaglandin dehydrogenase [NAD(+)] | HPGD | 1.05 | 1.10 | 1.06 | 0.67* | 0.71 | 0.85 | 0.80 |
| Translation factor GUF1, mitochondrial | GUF1 | 0.81 | 0.67* | 0.68* | 0.67* | 0.68 | 0.81 | 0.86 |
| Protein Daple | CCDC88C | 0.91 | 0.72* | 0.75* | 0.67* | 0.86 | 0.69 | 0.69 |
| U3 small nucleolar RNA-associated protein 6 homolog | UTP6 | 0.56* | 0.49* | 0.59* | 0.67 | 0.68 | 0.68 | 0.75 |

***C. Down-regulated proteins unique to Aquamin (AQ) with LPS-Cytokines [137 proteins]***

| | | Interventions | | | | | | |
|---|---|---|---|---|---|---|---|---|
| | | Control | | | *With LPS & Cytokines* | | | |
| Proteins | Genes | AQ | AQ+MES | MES | *LPS-Cyto* | ***AQ*** | *AQ+MES* | *MES* |
| Nuclear envelope pore membrane protein POM 121 | POM121 | 0.75 | 1.26 | 1.56* | 0.87 | 0.22* | 1.01 | 1.38 |
| Polyglutamine-binding protein 1 | PQBP1 | 1.06 | 1.73* | 1.85* | 1.25 | 0.26* | 0.69 | 1.16 |
| Protein Aster-B | GRAMD1B | 0.77 | 0.85 | 0.73 | 0.83 | 0.35* | 0.83 | 0.90 |
| Chromatin modification-related protein MEAF6 | MEAF6 | 0.73 | 0.77 | 0.68* | 1.03 | 0.46* | 0.77 | 0.86 |
| Protein C10 | C12orf57 | 0.85 | 0.95 | 1.28 | 1.09 | 0.46* | 0.72 | 0.88 |
| TNFAIP3-interacting protein 1 | TNIP1 | 0.92 | 1.07 | 1.14 | 0.73 | 0.47* | 0.83 | 0.97 |
| Insulin-like growth factor II | IGF2 | 0.62* | 2.07* | 2.04* | 0.74 | 0.48* | 1.65* | 2.22* |
| Pre-mRNA 3'-end-processing factor FIP1 | FIP1L1 | 0.92 | 0.96 | 0.98 | 0.75 | 0.48* | 0.67 | 1.03 |
| Protein O-glucosyltransferase 3 | POGLUT3 | 1.62* | 1.67* | 1.49* | 0.96 | 0.48* | 0.78 | 1.14 |
| Upstream stimulatory factor 1 | USF1 | 0.92 | 0.94 | 1.04 | 0.77 | 0.49* | 0.71 | 0.86 |
| Peptidyl-prolyl cis-trans isomerase G | PPIG | 0.85 | 0.81 | 0.98 | 0.96 | 0.49* | 0.82 | 1.10 |
| Neurotensin/neuromedin N | NTS | 0.74 | 1.54* | 1.30 | 1.01 | 0.50* | 1.03 | 0.72 |
| Large ribosomal subunit protein bL32m | MRPL32 | 1.35* | 1.33 | 1.23 | 1.38 | 0.50* | 0.88 | 1.01 |
| Transcriptional repressor p66-alpha | GATAD2A | 0.64* | 0.72 | 0.99 | 0.78 | 0.50* | 0.69 | 0.85 |
| Regulator of G-protein signaling 10 | RGS10 | 1.14 | 1.18 | 1.22 | 1.43 | 0.50* | 0.92 | 0.86 |
| Spliceosome-associated protein CWC27 homolog | CWC27 | 2.02* | 1.81* | 1.94* | 1.33 | 0.51 | 0.99 | 2.11* |
| CD2 antigen cytoplasmic tail-binding protein 2 | CD2BP2 | 1.03 | 1.11 | 1.35 | 1.15 | 0.52* | 0.93 | 1.03 |
| CDK-activating kinase assembly factor MAT1 | MNAT1 | 0.53* | 0.93 | 1.05 | 0.79 | 0.52 | 0.89 | 1.51 |
| Biogenesis of lysosome-related organelles complex 1 subunit 5 | BLOC1S5 | 0.92 | 0.82 | 1.14 | 0.96 | 0.52 | 0.73 | 0.94 |
| SAP domain-containing ribonucleoprotein | SARNP | 1.10 | 1.28 | 1.51* | 0.98 | 0.53* | 0.92 | 0.93 |

| | | | | | | | | |
|---|---|---|---|---|---|---|---|---|
| Protein LSM14 homolog A | LSM14A | 0.79 | 0.85 | 1.16 | 0.92 | 0.53 | 0.74 | 1.02 |
| EGF-containing fibulin-like extracellular matrix protein 1 | EFEMP1 | 0.62* | 2.26* | 2.51* | 0.81 | 0.53* | 1.88* | 2.37* |
| DNA polymerase beta | POLB | 0.99 | 1.03 | 1.08 | 1.13 | 0.53* | 0.70 | 0.85 |
| Serine/Arginine-related protein 53 | RSRC1 | 0.97 | 1.19 | 1.35 | 1.16 | 0.53* | 0.75 | 1.14 |
| Ribosomal protein eL22-like | RPL22L1 | 0.45* | 0.89 | 1.44 | 0.86 | 0.54 | 1.05 | 1.90* |
| Ubiquitin carboxyl-terminal hydrolase 17-like protein 15 | USP17L15 | 1.01 | 0.98 | 0.76* | 0.89 | 0.54* | 0.79 | 0.99 |
| Uncharacterized protein C7orf50 | C7orf50 | 0.69 | 1.04 | 1.15 | 1.07 | 0.54 | 0.84 | 1.23 |
| Protein TASOR | TASOR | 0.74 | 0.65 | 0.64* | 0.68 | 0.54 | 0.68 | 0.76 |
| CCHC-type zinc finger nucleic acid binding protein | CNBP | 0.62* | 0.99 | 0.95 | 1.03 | 0.55* | 0.93 | 1.04 |
| E3 ubiquitin-protein ligase TRIM47 | TRIM47 | 1.07 | 0.74 | 0.89 | 0.70 | 0.55 | 0.67 | 0.72 |
| MAPK regulated corepressor interacting protein 2 | MCRIP2 | 0.94 | 1.03 | 1.06 | 0.69 | 0.55 | 0.78 | 1.01 |
| Calpain-15 | CAPN15 | 0.79 | 0.74 | 0.87 | 0.77 | 0.55 | 0.69 | 0.82 |
| Methylated-DNA--protein-cysteine methyltransferase | MGMT | 0.91 | 1.07 | 1.06 | 0.80 | 0.55* | 0.67 | 0.68 |
| Interferon regulatory factor 2-binding protein 2 | IRF2BP2 | 0.90 | 1.10 | 0.99 | 0.83 | 0.56 | 0.74 | 0.86 |
| LYR motif-containing protein 1 | LYRM1 | 0.95 | 1.23 | 1.16 | 0.90 | 0.56 | 0.72 | 0.85 |
| Elongin-A | ELOA | 0.98 | 1.04 | 1.25* | 1.07 | 0.56* | 0.68 | 1.05 |
| Caveolae-associated protein 3 | CAVIN3 | 1.26 | 1.02 | 1.14 | 0.88 | 0.56 | 0.67 | 0.82 |
| PHD and RING finger domain-containing protein 1 | PHRF1 | 0.89 | 0.83 | 0.97 | 0.76 | 0.56 | 0.70 | 0.83 |
| Pre-mRNA-splicing factor ISY1 homolog | ISY1 | 0.93 | 1.03 | 0.99 | 1.04 | 0.56* | 0.70 | 0.90 |
| Nik-related protein kinase | NRK | 0.87 | 0.59* | 0.80 | 1.03 | 0.57 | 0.92 | 1.26 |
| NudC domain-containing protein 3 | NUDCD3 | 0.96 | 0.69 | 1.21 | 1.09 | 0.57 | 0.80 | 1.01 |
| Thymosin beta-10 | TMSB10 | 0.55* | 0.69 | 1.12 | 1.08 | 0.57 | 0.67 | 0.93 |
| Nucleolar protein 4-like | NOL4L | 0.74 | 0.81 | 0.72 | 0.74 | 0.58 | 0.73 | 0.71 |
| Transforming acidic coiled-coil-containing protein 2 | TACC2 | 1.05 | 0.98 | 1.01 | 0.83 | 0.58 | 0.69 | 0.78 |
| Ferritin light chain | FTL | 0.52* | 2.72* | 3.90* | 1.95* | 0.58 | 4.12* | 9.36* |
| Complement factor D | CFD | 0.59* | 1.43 | 3.72* | 0.81 | 0.58 | 1.54 | 2.05* |
| Rab GTPase-activating protein 1-like, isoform 10 | RABGAP1L | 0.99 | 0.79 | 1.10 | 0.99 | 0.58 | 0.68 | 0.68 |
| [Pyruvate dehydrogenase [acetyl-transferring]]-phosphatase 2, mitochondrial | PDP2 | 1.70* | 1.56* | 1.29 | 1.09 | 0.59 | 0.86 | 0.90 |
| MRG/MORF4L-binding protein | MRGBP | 1.05 | 0.96 | 1.20 | 1.19 | 0.59 | 0.95 | 0.98 |
| Protein IWS1 homolog | IWS1 | 1.07 | 0.99 | 1.21 | 0.95 | 0.59* | 0.80 | 0.92 |
| Probable dimethyladenosine transferase | DIMT1 | 0.65* | 0.59* | 0.64* | 0.70 | 0.59 | 0.75 | 0.90 |
| ABC-type oligopeptide transporter ABCB9 | ABCB9 | 0.40* | 0.67 | 1.05 | 1.77* | 0.59 | 0.93 | 2.42* |
| Transcription initiation factor IIB | GTF2B | 0.93 | 0.87 | 0.79 | 0.79 | 0.59 | 0.80 | 0.83 |
| Signal transducing adapter molecule 2 | STAM2 | 1.02 | 0.88 | 1.07 | 0.83 | 0.59 | 0.70 | 0.90 |
| Axin interactor, dorsalization-associated protein | AIDA | 0.80 | 0.86 | 0.90 | 1.01 | 0.60 | 0.75 | 0.84 |
| Exosome component 10 | EXOSC10 | 0.98 | 0.95 | 0.99 | 1.16 | 0.60 | 0.67 | 0.93 |

| | | | | | | | | |
|---|---|---|---|---|---|---|---|---|
| Probable glutamate--tRNA ligase, mitochondrial | EARS2 | 0.88 | 0.94 | 0.94 | 0.76 | 0.60 | 0.80 | 0.73 |
| Cyclin-dependent kinase 12 | CDK12 | 0.76 | 0.75 | 0.79 | 0.78 | 0.60 | 0.71 | 0.72 |
| Keratin, type II cytoskeletal 7 | KRT7 | 1.02 | 1.08 | 1.12* | 0.93 | 0.60* | 0.84 | 0.89 |
| DNA replication licensing factor MCM5 | MCM5 | 0.54* | 0.55* | 0.65* | 0.72 | 0.60 | 0.67 | 1.03 |
| Protein phosphatase 1 regulatory subunit 14B | PPP1R14B | 0.87 | 1.14 | 0.95 | 1.07 | 0.60 | 1.03 | 0.93 |
| Protein S100-A7 | S100A7 | 0.33* | 0.70 | 0.94 | 1.71* | 0.60 | 1.01 | 1.30 |
| Microtubule-associated serine/threonine-protein kinase 4 | MAST4 | 1.07 | 0.86 | 1.26 | 0.73 | 0.60 | 0.72 | 0.94 |
| Telomerase RNA component interacting RNase | TRIR | 0.74 | 0.98 | 1.04 | 0.84 | 0.61 | 0.85 | 0.85 |
| E3 ubiquitin-protein ligase UHRF2 | UHRF2 | 0.79 | 0.63 | 1.05 | 1.05 | 0.61 | 0.74 | 0.85 |
| N-alpha-acetyltransferase 20 | NAA20 | 0.97 | 1.21 | 1.16 | 1.11 | 0.61* | 0.84 | 1.01 |
| Segment polarity protein dishevelled homolog DVL-1 | DVL1 | 0.77 | 0.80 | 0.68* | 0.75 | 0.61 | 0.79 | 0.84 |
| Negative elongation factor A | NELFA | 1.04 | 1.11 | 1.26 | 0.88 | 0.61 | 0.69 | 0.85 |
| Ubiquitin-associated protein 2-like | UBAP2L | 0.83 | 0.94 | 1.39* | 0.90 | 0.61* | 0.76 | 1.09 |
| Splicing factor 45 | RBM17 | 0.93 | 0.98 | 0.93 | 1.00 | 0.62 | 0.83 | 0.97 |
| Complement factor H | CFH | 0.62* | 2.03* | 27.05* | 0.74 | 0.62 | 1.68* | 1.72* |
| START domain-containing protein 10 | STARD10 | 0.87 | 0.93 | 0.72* | 0.79 | 0.62 | 0.78 | 0.78 |
| ADP-ribose glycohydrolase OARD1 | OARD1 | 0.92 | 0.80 | 0.87 | 0.88 | 0.62 | 0.72 | 0.75 |
| WD repeat domain phosphoinositide-interacting protein 4 | WDR45 | 0.99 | 0.70 | 0.58* | 0.84 | 0.62 | 0.94 | 0.79 |
| RCC1-like G exchanging factor-like protein | RCC1L | 1.23 | 1.34 | 1.22 | 1.17 | 0.62 | 0.70 | 0.69 |
| Serine/threonine-protein kinase VRK1 | VRK1 | 0.78* | 0.82 | 0.76* | 0.77* | 0.62* | 0.67* | 0.79 |
| Zinc finger protein 185 | ZNF185 | 0.88 | 0.90 | 1.10 | 0.81 | 0.62 | 0.80 | 0.77 |
| Rab proteins geranylgeranyltransferase component A 2 | CHML | 1.17 | 1.18 | 1.06 | 1.06 | 0.62 | 0.68 | 0.70 |
| Dynein axonemal heavy chain 1 | DNAH1 | 0.51* | 1.54* | 2.15* | 1.10 | 0.62* | 2.04* | 2.51* |
| Importin subunit alpha-7 | KPNA6 | 0.83 | 0.83 | 0.94 | 0.80 | 0.62 | 0.67 | 0.92 |
| Mitotic-spindle organizing protein 1 | MZT1 | 1.14 | 1.09 | 2.73* | 0.92 | 0.63 | 0.71 | 2.13* |
| Prolyl endopeptidase-like | PREPL | 0.82 | 0.84 | 0.74 | 0.74 | 0.63 | 0.80 | 0.67 |
| Large ribosomal subunit protein eL24 | RPL24 | 0.78* | 0.79 | 0.91 | 0.75* | 0.63* | 0.74 | 0.97 |
| AP2-associated protein kinase 1 | AAK1 | 0.97 | 0.99 | 0.91 | 0.85 | 0.63* | 0.80 | 0.90 |
| Magnesium-dependent phosphatase 1 | MDP1 | 1.09 | 1.23 | 1.15 | 0.96 | 0.63 | 0.75 | 0.84 |
| Nucleolar GTP-binding protein 2 | GNL2 | 0.79 | 1.10 | 0.81 | 1.06 | 0.63 | 0.75 | 1.19 |
| Peptidyl-prolyl cis-trans isomerase NIMA-interacting 4 | PIN4 | 0.96 | 1.03 | 1.04 | 0.95 | 0.63 | 0.92 | 0.96 |
| Pre-mRNA-splicing factor 38A | PRPF38A | 1.16 | 1.06 | 1.00 | 1.01 | 0.63 | 0.77 | 0.71 |
| Inhibitor of Bruton tyrosine kinase | IBTK | 0.71 | 0.73 | 0.70 | 0.74 | 0.63 | 0.70 | 0.77 |
| Protein SCAF11 | SCAF11 | 0.77 | 0.71 | 0.80 | 0.74 | 0.63 | 0.73 | 0.75 |
| Nucleoprotein TPR | TPR | 0.93 | 0.93 | 1.03 | 1.02 | 0.63* | 0.77 | 0.91 |

| | | | | | | | | |
|---|---|---|---|---|---|---|---|---|
| Segment polarity protein dishevelled homolog DVL-2 | DVL2 | 1.07 | 1.18 | 1.11 | 1.08 | 0.63 | 0.70 | 0.96 |
| Uncharacterized protein C1orf21 | C1orf21 | 1.00 | 1.07 | 1.02 | 1.18 | 0.63 | 0.80 | 0.89 |
| Zinc finger protein castor homolog 1 | CASZ1 | 0.92 | 0.95 | 0.79 | 0.78 | 0.63 | 0.78 | 0.70 |
| Zinc finger CCCH domain-containing protein 18 | ZC3H18 | 0.84 | 0.69 | 0.99 | 0.94 | 0.63 | 0.76 | 0.87 |
| Ras GTPase-activating protein 2 | RASA2 | 0.84 | 0.82 | 0.94 | 0.75 | 0.64 | 0.82 | 0.77 |
| Negative elongation factor B | NELFB | 0.86 | 0.85 | 0.85 | 0.86 | 0.64 | 0.78 | 0.69 |
| PDZ and LIM domain protein 2 | PDLIM2 | 0.86 | 1.11 | 1.11 | 0.84 | 0.64 | 0.78 | 0.90 |
| Histone deacetylase 8 | HDAC8 | 0.95 | 0.98 | 0.89 | 0.91 | 0.64 | 0.79 | 0.74 |
| Lysophosphatidylcholine acyltransferase 2 | LPCAT2 | 1.02 | 1.01 | 1.02 | 0.81 | 0.64 | 0.68 | 0.73 |
| Serine/threonine-protein phosphatase 1 regulatory subunit 10 | PPP1R10 | 1.08 | 0.91 | 1.17 | 1.31 | 0.64 | 0.75 | 0.99 |
| SH3 domain-binding protein 5-like | SH3BP5L | 1.15 | 1.08 | 1.06 | 1.07 | 0.64 | 0.88 | 0.89 |
| DNA-directed RNA polymerase I subunit RPA2 | POLR1B | 0.75 | 0.69 | 0.76 | 0.76 | 0.64 | 0.81 | 0.84 |
| FYVE and coiled-coil domain-containing protein 1 | FYCO1 | 0.95 | 0.85 | 0.89 | 0.73* | 0.64* | 0.76 | 0.67* |
| AFG1-like ATPase | AFG1L | 1.06 | 0.82 | 0.71 | 0.82 | 0.64 | 0.77 | 0.86 |
| Nitric oxide synthase-interacting protein | NOSIP | 0.68* | 0.84 | 0.92 | 0.78 | 0.65 | 0.69 | 0.92 |
| High mobility group protein 20A | HMG20A | 0.84 | 0.90 | 0.92 | 0.94 | 0.65 | 0.69 | 0.99 |
| Collagen alpha-2(I) chain | COL1A2 | 0.53* | 2.15* | 2.47* | 0.72 | 0.65 | 1.88* | 2.71* |
| mRNA cap guanine-N7 methyltransferase | RNMT | 1.08 | 0.83 | 0.97 | 0.83 | 0.65 | 0.78 | 0.85 |
| Sesquipedalian-1 | PHETA1 | 0.89 | 1.00 | 0.85 | 0.89 | 0.65 | 0.76 | 0.70 |
| Alcohol dehydrogenase 1C | ADH1C | 0.94 | 0.94 | 0.70* | 0.95 | 0.65* | 0.72* | 1.22* |
| Jupiter microtubule associated homolog 2 | JPT2 | 0.73 | 0.98 | 0.90 | 0.77 | 0.65 | 0.70 | 0.83 |
| Endothelial differentiation-related factor 1 | EDF1 | 0.86 | 0.81 | 1.04 | 0.89 | 0.65 | 0.72 | 0.90 |
| Islet cell autoantigen 1 | ICA1 | 0.87 | 0.81 | 0.93 | 0.70* | 0.65 | 0.76 | 0.74 |
| Cell death regulator Aven | AVEN | 0.76* | 0.71* | 0.76* | 0.80 | 0.65 | 0.70 | 0.73 |
| Zinc finger CCCH domain-containing protein 11A | ZC3H11A | 0.80 | 1.03 | 1.07 | 0.74 | 0.65 | 0.67 | 0.71 |
| Microtubule-associated protein 4 | MAP4 | 1.03 | 0.96 | 1.19* | 0.75* | 0.65* | 0.67* | 0.84 |
| Nuclear cap-binding protein subunit 2 | NCBP2 | 0.94 | 1.01 | 1.11 | 1.06 | 0.65 | 0.75 | 1.18 |
| UBAP1-MVB12-associated (UMA)-domain containing protein 1 | UMAD1 | 0.77 | 0.75 | 0.90 | 0.85 | 0.65 | 0.80 | 0.72 |
| SCY1-like protein 2 | SCYL2 | 1.02 | 0.98 | 1.17 | 0.98 | 0.65 | 0.78 | 0.89 |
| Thyrotroph embryonic factor | TEF | 1.06 | 0.95 | 0.91 | 0.90 | 0.65 | 0.85 | 0.96 |
| dCTP pyrophosphatase 1 | DCTPP1 | 0.86 | 0.75 | 0.70* | 0.86 | 0.66 | 0.77 | 0.84 |
| Trafficking protein particle complex subunit 14 | TRAPPC14 | 1.04 | 0.81 | 0.89 | 0.91 | 0.66 | 0.96 | 0.79 |
| Small ribosomal subunit protein uS12m | MRPS12 | 0.94 | 0.74 | 1.12 | 0.90 | 0.66 | 0.71 | 1.01 |
| Tripartite motif-containing protein 2 | TRIM2 | 1.00 | 0.85 | 0.86 | 0.81 | 0.66 | 0.74 | 0.74 |
| Sulfotransferase 2B1 | SULT2B1 | 0.98 | 0.88 | 1.25 | 0.75 | 0.66 | 0.77 | 0.83 |
| B-cell CLL/lymphoma 7 protein family member A | BCL7A | 0.98 | 1.13 | 1.15 | 1.43 | 0.66 | 0.83 | 1.23 |

| | | | | | | | | |
|---|---|---|---|---|---|---|---|---|
| Nuclear factor NF-kappa-B p105 subunit | NFKB1 | 0.95 | 0.97 | 1.04 | 0.90 | 0.66 | 0.81 | 0.97 |
| Exportin-6 | XPO6 | 0.67 | 0.58* | 0.69 | 0.91 | 0.66 | 0.82 | 0.82 |
| Filamin-binding LIM protein 1 | FBLIM1 | 0.77 | 0.92 | 0.79 | 0.93 | 0.66 | 0.87 | 0.74 |
| Protein FAM83B | FAM83B | 0.84 | 0.75 | 1.00 | 0.98 | 0.66 | 0.90 | 1.07 |
| Protein PRRC2C | PRRC2C | 0.87 | 0.87 | 1.02 | 0.88 | 0.67 | 0.80 | 0.98 |
| Plectin | PLEC | 1.17 | 0.85 | 0.67* | 0.71* | 0.67* | 0.75 | 0.94 |
| Mitotic interactor and substrate of PLK1 | MISP | 0.92 | 1.07 | 1.11 | 0.92 | 0.67* | 0.89 | 1.06 |
| Suppressor of SWI4 1 homolog | PPAN | 1.18 | 0.98 | 1.15 | 1.26 | 0.67 | 0.74 | 1.07 |
| Zinc finger CCCH domain-containing protein 15 | ZC3H15 | 0.65* | 0.74* | 0.80* | 0.74* | 0.67* | 0.67 | 0.80 |
| Mitochondrial import inner membrane translocase subunit Tim8 A | TIMM8A | 1.13 | 1.09 | 0.74* | 1.13 | 0.67 | 0.86 | 0.84 |

***D. Down-regulated proteins unique to Aquamin plus Mesalamine (AQ+MES) with LPS-Cytokines [90 proteins]***

| | | Interventions | | | | | | |
|---|---|---|---|---|---|---|---|---|
| | | Control | | | *With LPS & Cytokines* | | | |
| Proteins | Genes | AQ | AQ+MES | MES | *LPS-Cyto* | *AQ* | ***AQ+MES*** | *MES* |
| Calmodulin-like protein 3 | CALML3 | 1.02 | 52.36* | 12.16* | 0.75 | 1.15 | 0.24* | 0.82 |
| Loricrin | LORICRIN | 0.48* | 0.44* | 0.64* | 1.79* | 1.63* | 0.30* | 1.10 |
| Ferroxidase HEPHL1 | HEPHL1 | 0.77 | 61.07* | 3.28* | 0.93 | 1.02 | 0.31* | 1.91 |
| Filaggrin-2 | FLG2 | 0.24* | 0.30* | 0.44* | 0.94 | 0.69* | 0.34* | 0.69* |
| Keratin, type II cuticular Hb2 | KRT82 | 0.91 | 23.09* | 1.49* | 0.74 | 1.00 | 0.40* | 1.04 |
| Keratin, type II cytoskeletal 1 | KRT1 | 0.42* | 0.49* | 0.57* | 1.10 | 1.59* | 0.44* | 1.02 |
| UPF0235 protein C15orf40 | C15orf40 | 0.86 | 0.40* | 0.33* | 1.30 | 0.91 | 0.45* | 0.67 |
| Solute carrier family 13 member 2 | SLC13A2 | 0.79 | 0.59* | 0.49* | 0.99 | 0.97 | 0.46* | 0.81 |
| Cornifin-B | SPRR1B | 0.79 | 1.62* | 1.68* | 1.14 | 4.30* | 0.46* | 3.71* |
| Complement C1r subcomponent-like protein | C1RL | 1.53* | 1.26 | 1.35 | 1.05 | 1.08 | 0.47* | 0.76 |
| Keratin, type I cytoskeletal 27 | KRT27 | 0.48* | 0.64 | 0.52* | 1.06 | 0.68 | 0.48* | 1.03 |
| Integrin alpha-7 | ITGA7 | 0.48* | 0.78 | 0.74 | 5.21* | 0.84 | 0.49* | 1.76 |
| Keratin, type I cytoskeletal 9 | KRT9 | 0.46* | 0.44* | 0.46* | 1.11 | 1.99* | 0.51* | 0.89 |
| Keratin, type II cytoskeletal 78 | KRT78 | 0.71* | 0.67* | 1.26* | 1.50* | 1.92* | 0.51* | 1.49* |
| Kinesin-like protein KIF14 | KIF14 | 1.08 | 1.04 | 1.02 | 1.08 | 1.33 | 0.51 | 0.80 |
| Ubiquitin-conjugating enzyme E2 E1 | UBE2E1 | 0.95 | 0.75 | 1.03 | 0.83 | 0.72 | 0.51 | 0.78 |
| Transcription factor 20 | TCF20 | 0.66* | 0.99 | 1.00 | 2.22* | 0.80 | 0.51 | 1.59 |
| Early estrogen-induced gene 1 protein | EEIG1 | 0.91 | 0.93 | 1.05 | 0.79 | 0.78 | 0.52 | 0.90 |
| RELT-like protein 1 | RELL1 | 0.81 | 1.11 | 1.18 | 0.95 | 0.84 | 0.52 | 0.77 |
| Trypsin-2 | PRSS2 | 1.21 | 0.69 | 0.67* | 1.28 | 0.72 | 0.53 | 0.68 |
| Semenogelin-1 | SEMG1 | 1.74* | 4.10* | 2.91* | 0.71 | 2.56* | 0.55* | 2.67* |

| | | | | | | | | |
|---|---|---|---|---|---|---|---|---|
| Keratin, type I cytoskeletal 17 | KRT17 | 0.71* | 0.64* | 0.61* | 1.72* | 0.68 | 0.55* | 0.77 |
| Ubiquitin thioesterase OTU1 | YOD1 | 1.22 | 1.23 | 2.20* | 0.87 | 0.79 | 0.55 | 0.91 |
| Zinc-alpha-2-glycoprotein | AZGP1 | 0.88 | 1.21 | 1.26 | 2.00* | 1.12 | 0.56* | 1.08 |
| E3 ubiquitin-protein ligase MIB1 | MIB1 | 0.62* | 0.64* | 0.55* | 0.69 | 0.71 | 0.56 | 0.72 |
| Lysosomal thioesterase PPT2 | PPT2 | 0.78 | 1.58* | 0.68 | 1.00 | 0.92 | 0.56 | 0.82 |
| Plakophilin-1 | PKP1 | 0.84 | 3.19* | 2.07* | 0.84 | 2.73* | 0.56* | 2.07* |
| Alpha-2-macroglobulin receptor-associated protein | LRPAP1 | 0.90 | 0.82 | 0.91 | 0.82 | 0.73 | 0.56* | 0.67* |
| Peroxisomal ATPase PEX6 | PEX6 | 0.87 | 0.82 | 0.92 | 0.98 | 0.72 | 0.57 | 0.76 |
| UPF0538 protein C2orf76 | C2orf76 | 0.62* | 0.76 | 0.83 | 0.77 | 0.71 | 0.58 | 0.73 |
| TBC1 domain family member 4 | TBC1D4 | 0.72 | 0.82 | 0.99 | 0.92 | 0.82 | 0.58 | 0.85 |
| Cytotoxic granule associated RNA binding protein TIA1 | TIA1 | 0.87 | 0.80 | 0.88 | 0.99 | 0.75 | 0.60 | 0.74 |
| 3'-5' RNA helicase YTHDC2 | YTHDC2 | 0.61* | 0.59* | 0.64* | 0.82 | 0.72 | 0.60* | 0.78 |
| UPF0488 protein C8orf33 | C8orf33 | 0.95 | 1.00 | 0.97 | 0.80 | 0.84 | 0.60* | 0.68* |
| Deleted in malignant brain tumors 1 protein | DMBT1 | 2.24* | 0.89 | 1.16* | 1.67* | 1.57* | 0.60* | 0.73* |
| Engulfment and cell motility protein 2 | ELMO2 | 1.06 | 1.12 | 1.49 | 1.10 | 1.17 | 0.60 | 1.01 |
| Arginase-1 | ARG1 | 0.45* | 0.50* | 0.84 | 0.93 | 0.87 | 0.60 | 0.98 |
| Large ribosomal subunit protein uL23m | MRPL23 | 0.93 | 0.93 | 1.09 | 0.89 | 0.76 | 0.61 | 0.73 |
| SH3 domain-containing protein 21 | SH3D21 | 1.28 | 1.49* | 1.04 | 1.06 | 0.84 | 0.61 | 0.68 |
| Elafin | PI3 | 1.00 | 0.70 | 1.30 | 0.83 | 0.80 | 0.61 | 0.77 |
| Uridine-cytidine kinase-like 1 | UCKL1 | 0.71 | 0.77 | 0.75 | 0.90 | 0.75 | 0.61 | 0.74 |
| Probable rRNA-processing protein EBP2 | EBNA1BP2 | 0.94 | 0.83 | 1.12 | 1.17 | 0.71 | 0.61* | 0.96 |
| Calmodulin-regulated spectrin-associated protein 1 | CAMSAP1 | 0.59* | 0.92 | 1.06 | 0.77 | 0.86 | 0.61 | 0.70 |
| Synaptotagmin-like protein 2 | SYTL2 | 0.92 | 0.83 | 0.86 | 0.89 | 0.71 | 0.62 | 0.79 |
| Doublecortin domain-containing protein 1 | DCDC1 | 0.92 | 0.96 | 0.81 | 0.79 | 0.75 | 0.62 | 1.03 |
| WD repeat-containing protein 36 | WDR36 | 0.60* | 0.52* | 0.56* | 0.76 | 0.69 | 0.62 | 0.71 |
| PRKC apoptosis WT1 regulator protein | PAWR | 0.92 | 1.01 | 1.18 | 0.97 | 0.71 | 0.62 | 0.85 |
| Ankyrin repeat and MYND domain-containing protein 2 | ANKMY2 | 0.84 | 0.78 | 0.81 | 0.79 | 0.75 | 0.62 | 0.76 |
| Methylthioribulose-1-phosphate dehydratase | APIP | 1.24 | 0.94 | 1.17 | 0.99 | 0.71 | 0.62 | 0.97 |
| Zinc finger CCCH domain-containing protein 13 | ZC3H13 | 0.93 | 0.91 | 0.95 | 1.07 | 0.68 | 0.62 | 0.74 |
| Pseudouridylate synthase RPUSD4, mitochondrial | RPUSD4 | 0.89 | 0.95 | 0.90 | 0.83 | 1.66 | 0.63 | 0.78 |
| CD99 antigen | CD99 | 0.94 | 1.06 | 1.08 | 0.75 | 0.82 | 0.63 | 1.13 |
| Tyrosine-protein kinase BAZ1B | BAZ1B | 0.67* | 0.66* | 0.69* | 0.85 | 0.68 | 0.63 | 0.72 |
| Interleukin-18 | IL18 | 0.96 | 0.82 | 0.87 | 0.80 | 0.75 | 0.63* | 0.72* |
| Glia-derived nexin | SERPINE2 | 0.74* | 0.75* | 1.04 | 0.74* | 0.67* | 0.63* | 0.68* |
| Monocyte differentiation antigen CD14 | CD14 | 1.13 | 0.54* | 1.02 | 0.95 | 1.09 | 0.63 | 0.72 |
| Liprin-beta-2 | PPFIBP2 | 0.82 | 0.79 | 0.86 | 0.80 | 0.74 | 0.63 | 0.73 |
| Myosin light chain kinase, smooth muscle | MYLK | 0.98 | 0.96 | 1.07 | 0.75 | 0.68 | 0.63 | 0.76 |

| | | | | | | | | |
|---|---|---|---|---|---|---|---|---|
| Serum response factor-binding protein 1 | SRFBP1 | 1.01 | 1.10 | 1.12 | 1.54* | 0.80 | 0.63 | 1.07 |
| Xaa-Pro dipeptidase | PEPD | 1.04 | 0.94 | 0.90 | 0.96 | 0.82 | 0.64 | 0.90 |
| Phosphoribosylformylglycinamidine synthase | PFAS | 0.86 | 0.97 | 0.85 | 1.16 | 0.82 | 0.64 | 1.06 |
| Keratin, type I cytoskeletal 13 | KRT13 | 0.75* | 0.53* | 0.92 | 0.69* | 0.87 | 0.64 | 1.46* |
| Neuroblastoma suppressor of tumorigenicity 1 | NBL1 | 1.17 | 0.99 | 0.92 | 0.90 | 0.90 | 0.64 | 0.87 |
| Rho guanine nucleotide exchange factor 10-like protein | ARHGEF10L | 0.96 | 0.79 | 0.85 | 0.76 | 0.73 | 0.64 | 0.79 |
| Large ribosomal subunit protein eL39 | RPL39 | 0.80 | 0.87 | 0.91 | 0.94 | 0.70 | 0.64 | 1.00 |
| Nucleolar protein 16 | NOP16 | 0.95 | 0.81 | 0.95 | 1.18 | 0.70 | 0.64 | 1.03 |
| Nucleolar protein 6 | NOL6 | 0.67* | 0.57* | 0.61* | 0.81 | 0.75 | 0.64 | 0.74 |
| cAMP-dependent protein kinase catalytic subunit beta | PRKACB | 0.90 | 0.84 | 0.75 | 0.73 | 0.70 | 0.64 | 0.72 |
| Syntaxin-binding protein 6 | STXBP6 | 0.91 | 0.70 | 1.02 | 1.04 | 0.74 | 0.64 | 0.97 |
| A-kinase anchor protein 7 isoform gamma | AKAP7 | 1.47 | 1.45 | 1.24 | 0.82 | 0.98 | 0.64 | 0.75 |
| Tumor protein D53 | TPD52L1 | 1.13 | 0.91 | 1.09 | 0.93 | 0.75 | 0.64 | 0.70 |
| TSC22 domain family protein 4 | TSC22D4 | 0.99 | 0.86 | 1.00 | 0.97 | 0.98 | 0.65 | 1.02 |
| NAD-dependent protein deacetylase sirtuin-2 | SIRT2 | 0.98 | 0.70 | 1.14 | 0.95 | 0.77 | 0.65 | 0.89 |
| Claudin-1 | CLDN1 | 0.52* | 0.82 | 1.07 | 0.92 | 0.85 | 0.65 | 1.06 |
| Ubiquitin-ribosomal protein eL40 fusion protein | UBA52 | 0.91 | 0.84 | 0.99 | 0.77 | 0.67 | 0.65 | 0.80 |
| Ceramide-1-phosphate transfer protein | CPTP | 0.90 | 0.78 | 0.78 | 0.83 | 0.81 | 0.65 | 0.83 |
| Mesothelin | MSLN | 0.98 | 0.86 | 1.13 | 0.86 | 0.77 | 0.65* | 0.79 |
| MutS protein homolog 5 | MSH5 | 1.03 | 0.80 | 1.02 | 0.91 | 0.80 | 0.65 | 0.69 |
| Nucleolar protein 14 | NOP14 | 0.79* | 0.81 | 1.18* | 1.45* | 0.75 | 0.65* | 1.42* |
| Alcohol dehydrogenase 6 | ADH6 | 1.07 | 0.66* | 0.66* | 0.79 | 0.97 | 0.65* | 0.74 |
| KxDL motif-containing protein 1 | KXD1 | 0.83 | 0.78 | 0.80 | 0.89 | 0.92 | 0.66 | 0.85 |
| Kalirin | KALRN | 0.90 | 0.85 | 0.95 | 0.95 | 0.73 | 0.66 | 0.89 |
| E3 ubiquitin-protein ligase RBBP6 | RBBP6 | 1.01 | 0.94 | 1.17 | 0.97 | 0.70 | 0.66 | 1.02 |
| Lysozyme C | LYZ | 0.97 | 0.91 | 0.94 | 0.94 | 0.79 | 0.66* | 0.92 |
| Mitogen-activated protein kinase kinase kinase 4 | MAP3K4 | 0.79 | 0.70 | 0.71 | 0.86 | 0.71 | 0.66 | 0.68 |
| Coiled-coil domain-containing protein 9 | CCDC9 | 1.01 | 1.07 | 0.94 | 1.18 | 0.77 | 0.66 | 0.69 |
| DNA-directed RNA polymerase II subunit RPB9 | POLR2I | 0.70* | 0.79 | 0.78 | 0.75 | 0.73 | 0.66 | 0.85 |
| Cordon-bleu protein-like 1 | COBLL1 | 0.84 | 0.83 | 0.95 | 0.76 | 0.73 | 0.66 | 0.86 |
| Protein MANBAL | MANBAL | 1.08 | 1.01 | 0.99 | 0.91 | 0.84 | 0.66 | 0.79 |
| Xylulose kinase | XYLB | 1.01 | 0.80 | 0.73* | 0.98 | 0.76 | 0.66* | 0.70* |

***E. Down-regulated proteins unique to Mesalamine (MES) with LPS-Cytokines [113 proteins]***

| | | Interventions | | | |
|---|---|---|---|---|---|
| | | Control | | | *With LPS & Cytokines* |

| Proteins | Genes | AQ | AQ+MES | MES | *LPS-Cyto* | *AQ* | *AQ+MES* | ***MES*** |
|---|---|---|---|---|---|---|---|---|
| Calcium/calmodulin-dependent protein kinase type 1B | PNCK | 1.12 | 0.39* | 0.14* | 1.00 | 1.60 | 0.82 | 0.13* |
| CD5 antigen-like | CD5L | 0.66 | 1.53 | 23.17* | 0.77 | 0.69 | 0.82 | 0.29* |
| StAR-related lipid transfer protein 7, mitochondrial | STARD7 | 0.92 | 1.44 | 1.26 | 0.95 | 0.85 | 1.26 | 0.30* |
| Profilin-3 | PFN3 | 2.06* | 0.44* | 0.48* | 0.82 | 0.81 | 0.84 | 0.37* |
| Ubiquinone biosynthesis protein COQ4 homolog, mitochondrial | COQ4 | 1.99* | 1.66* | 0.97 | 1.02 | 0.88 | 0.76 | 0.40* |
| Cytochrome P450 4F2 | CYP4F2 | 1.28 | 1.00 | 0.89 | 0.76 | 0.96 | 0.81 | 0.43* |
| 7-methylguanosine phosphate-specific 5'-nucleotidase | NT5C3B | 1.45* | 1.37 | 1.15 | 1.06 | 0.83 | 1.75 | 0.44* |
| Rho GTPase-activating protein 42 | ARHGAP42 | 0.55* | 0.74 | 0.71 | 0.78 | 0.67 | 0.97 | 0.45* |
| Cdc42 effector protein 5 | CDC42EP5 | 0.44* | 0.80 | 0.94 | 1.05 | 0.88 | 0.85 | 0.47* |
| Angiotensin-converting enzyme | ACE | 1.37* | 1.37* | 0.72* | 1.10 | 1.34* | 0.70* | 0.47* |
| Sodium-dependent neutral amino acid transporter B(0)AT1 | SLC6A19 | 1.63* | 2.12* | 0.93 | 1.71* | 1.74* | 1.12 | 0.47* |
| Protein O-mannose kinase | POMK | 1.35 | 1.28 | 1.31 | 0.78 | 0.83 | 0.81 | 0.48* |
| Dimethylaniline monooxygenase [N-oxide-forming] 4 | FMO4 | 1.07 | 0.86 | 0.56* | 1.06 | 0.72 | 0.70 | 0.48* |
| Serine/threonine-protein kinase 38-like | STK38L | 1.41 | 0.92 | 0.85 | 1.13 | 0.74 | 1.02 | 0.49* |
| Intermembrane lipid transfer protein VPS13D | VPS13D | 0.63* | 0.69 | 0.51* | 0.69 | 0.82 | 0.95 | 0.49* |
| Fibronectin type III and SPRY domain-containing protein 1 | FSD1 | 3.62* | 3.96* | 1.02 | 0.70 | 2.52* | 1.94* | 0.49* |
| UDP-glucuronosyltransferase 2B7 | UGT2B7 | 1.26 | 1.15 | 0.68* | 0.75 | 1.37 | 0.97 | 0.50* |
| UDP-glucuronosyltransferase 2A3 | UGT2A3 | 1.71* | 1.27 | 0.93 | 0.94 | 1.03 | 0.73 | 0.51* |
| Tyrosine-protein kinase STYK1 | STYK1 | 1.26 | 1.26 | 1.03 | 0.83 | 0.95 | 0.75 | 0.51* |
| DNA repair protein complementing XP-C cells | XPC | 1.02 | 0.73 | 0.86 | 0.91 | 0.69 | 0.76 | 0.51* |
| Carcinoembryonic antigen-related cell adhesion molecule 7 | CEACAM7 | 1.29* | 0.98 | 0.79* | 0.69* | 0.82 | 0.91 | 0.52* |
| Syntabulin | SYBU | 1.08 | 0.71 | 0.67 | 0.89 | 1.10 | 0.71 | 0.52 |
| Bile salt export pump | ABCB11 | 1.44* | 1.28 | 0.96 | 0.92 | 1.08 | 1.03 | 0.52 |
| Serine/threonine-protein kinase ULK3 | ULK3 | 0.96 | 1.00 | 1.00 | 0.78 | 0.74 | 0.67 | 0.52* |
| Protein AMN1 homolog | AMN1 | 0.83 | 0.93 | 0.95 | 0.67 | 0.92 | 0.82 | 0.53* |
| Phosphatidylserine decarboxylase proenzyme, mitochondrial | PISD | 1.17 | 0.99 | 0.92 | 0.78 | 0.77 | 0.72 | 0.53* |
| GPI ethanolamine phosphate transferase 1 | PIGN | 1.17 | 1.12 | 1.08 | 0.80 | 0.72 | 0.74 | 0.53* |
| Sulfotransferase 1C2 | SULT1C2 | 1.49* | 0.71* | 0.76 | 0.94 | 1.13 | 0.68 | 0.54* |
| NEDD4-like E3 ubiquitin-protein ligase WWP1 | WWP1 | 0.82 | 0.76 | 0.69 | 0.90 | 0.98 | 0.69 | 0.54 |
| Interleukin-1 receptor-associated kinase 4 | IRAK4 | 0.94 | 0.92 | 0.91 | 0.80 | 0.74 | 0.75 | 0.54* |
| RAB6-interacting golgin | GORAB | 0.98 | 0.96 | 1.26 | 0.80 | 0.99 | 1.05 | 0.55 |
| Immunoglobulin lambda variable 1-51 | IGLV1-51 | 0.85 | 1.31 | 8.76* | 0.95 | 0.92 | 0.94 | 0.55* |

| | | | | | | | | |
|---|---|---|---|---|---|---|---|---|
| Ephrin-A2 | EFNA2 | 0.88 | 0.60* | 0.64* | 0.88 | 0.70 | 0.70 | 0.55* |
| Transmembrane 6 superfamily member 2 | TM6SF2 | 1.03 | 0.80 | 0.84 | 0.74 | 0.89 | 1.02 | 0.56 |
| Sorting nexin-30 | SNX30 | 1.10 | 0.73 | 0.66* | 0.80 | 0.73 | 0.69 | 0.56* |
| Kinase D-interacting substrate of 220 kDa | KIDINS220 | 0.66 | 0.81 | 0.67 | 0.81 | 0.76 | 0.85 | 0.57 |
| Ribonuclease H2 subunit A | RNASEH2A | 1.15 | 1.09 | 1.21 | 1.09 | 0.71 | 0.82 | 0.57 |
| Sodium-coupled monocarboxylate transporter 2 | SLC5A12 | 1.14 | 1.13 | 0.87 | 1.59* | 1.39 | 0.68 | 0.57 |
| Indian hedgehog protein | IHH | 1.38* | 1.29 | 0.84 | 0.97 | 1.24 | 0.92 | 0.57 |
| Heat shock factor protein 1 | HSF1 | 0.93 | 0.99 | 0.95 | 0.75 | 0.70 | 0.76 | 0.57* |
| Valacyclovir hydrolase | BPHL | 1.04 | 1.03 | 0.86 | 0.69* | 0.84 | 0.77 | 0.58* |
| Protein FAM98C | FAM98C | 0.89 | 0.85 | 0.91 | 0.67 | 0.78 | 0.67 | 0.58 |
| Immunoglobulin heavy variable 3-7 | IGHV3-7 | 0.93 | 2.12* | 23.53* | 0.83 | 1.48 | 1.53 | 0.58 |
| Oxysterol-binding protein-related protein 5 | OSBPL5 | 0.87 | 0.71 | 0.76 | 0.68 | 0.98 | 1.08 | 0.58* |
| Calcium channel flower homolog | CACFD1 | 0.89 | 0.81 | 0.77 | 0.82 | 1.01 | 0.91 | 0.58 |
| AKT-interacting protein | AKTIP | 0.85 | 0.90 | 0.81 | 0.67* | 0.75 | 0.73 | 0.58* |
| Creatine kinase B-type | CKB | 1.86* | 1.48* | 1.47* | 0.69* | 0.99 | 1.05 | 0.58* |
| Bridge-like lipid transfer protein family member 1 | BLTP1 | 0.98 | 0.65 | 0.59* | 0.88 | 1.01 | 0.80 | 0.58 |
| Peroxisomal bifunctional enzyme | EHHADH | 1.10 | 1.00 | 0.90 | 0.70* | 0.71 | 0.74 | 0.59* |
| Inhibitor of growth protein 1 | ING1 | 1.02 | 0.47* | 0.62* | 0.86 | 1.16 | 0.73 | 0.59 |
| Patatin-like phospholipase domain-containing protein 2 | PNPLA2 | 1.07 | 1.23 | 0.89 | 0.71 | 0.90 | 0.92 | 0.59* |
| Transcription cofactor vestigial-like protein 4 | VGLL4 | 0.92 | 1.59* | 1.92* | 0.98 | 0.94 | 0.98 | 0.60 |
| Mitochondrial peptide methionine sulfoxide reductase | MSRA | 1.01 | 0.90 | 0.78 | 0.73 | 0.88 | 0.69 | 0.60 |
| Retinol dehydrogenase 10 | RDH10 | 1.07 | 0.96 | 0.96 | 0.67 | 0.90 | 0.68 | 0.61 |
| Guanylyl cyclase C | GUCY2C | 1.25 | 1.13 | 1.00 | 0.75 | 0.80 | 0.74 | 0.61 |
| Serine hydrolase RBBP9 | RBBP9 | 1.10 | 0.93 | 0.99 | 1.02 | 0.85 | 0.76 | 0.61 |
| Myelin regulatory factor-like protein | MYRFL | 0.96 | 0.80 | 0.57* | 1.19 | 0.89 | 0.77 | 0.61 |
| Cadherin-related family member 2 | CDHR2 | 1.07 | 1.07 | 0.89 | 0.79 | 0.96 | 0.68 | 0.61* |
| Keratinocyte differentiation factor 1 | KDF1 | 0.96 | 1.14 | 1.14 | 0.92 | 0.82 | 0.74 | 0.61 |
| Epidermal retinol dehydrogenase 2 | SDR16C5 | 1.03 | 1.10 | 0.91 | 0.76 | 0.82 | 0.71 | 0.61* |
| Mitochondrial Rho GTPase 1 | RHOT1 | 0.78 | 1.00 | 1.00 | 0.87 | 0.83 | 0.92 | 0.61 |
| Alanine aminotransferase 1 | GPT | 1.14 | 1.16 | 0.80 | 0.68* | 0.84 | 0.80 | 0.62* |
| Polyprenol reductase | SRD5A3 | 1.02 | 0.92 | 0.90 | 0.68 | 0.89 | 0.78 | 0.62 |
| A disintegrin and metalloproteinase with thrombospondin motifs 2 | ADAMTS2 | 1.04 | 0.96 | 0.98 | 0.90 | 0.79 | 0.82 | 0.62 |
| Cytochrome P450 2C19 | CYP2C19 | 2.12* | 1.15 | 0.61* | 1.64* | 1.98* | 0.88 | 0.62 |
| Transmembrane protein 54 | TMEM54 | 1.23 | 1.25 | 1.27 | 0.97 | 0.70 | 0.74 | 0.62 |
| Protein asteroid homolog 1 | ASTE1 | 0.94 | 1.10 | 1.01 | 0.75 | 1.22 | 1.40 | 0.62 |
| Sarcoplasmic/endoplasmic reticulum calcium ATPase 3 | ATP2A3 | 1.17 | 0.63* | 0.89 | 0.69 | 0.72 | 0.68 | 0.62 |

| | | | | | | | | |
|---|---|---|---|---|---|---|---|---|
| Interleukin-1 receptor-associated kinase 1 | IRAK1 | 0.88 | 0.82 | 0.84 | 0.75 | 0.74 | 0.78 | 0.62 |
| SEC14 domain and spectrin repeat-containing protein 1 | SESTD1 | 0.79 | 0.84 | 0.82 | 0.77 | 0.84 | 0.68 | 0.63 |
| All-trans-retinol 13,14-reductase | RETSAT | 1.09 | 0.92 | 0.82* | 0.79* | 0.98 | 0.76 | 0.63* |
| Tuftelin | TUFT1 | 0.84 | 0.99 | 1.01 | 0.78 | 0.97 | 0.67 | 0.63 |
| Gap junction beta-3 protein | GJB3 | 0.93 | 0.70 | 0.74 | 0.68 | 1.01 | 0.70 | 0.63 |
| Sorting nexin-13 | SNX13 | 1.11 | 0.85 | 0.80 | 0.69 | 0.75 | 0.99 | 0.63 |
| Protein angel homolog 2 | ANGEL2 | 0.98 | 0.89 | 0.54* | 0.99 | 0.91 | 0.95 | 0.63 |
| UDP-N-acetylglucosamine transferase subunit ALG14 homolog | ALG14 | 0.63 | 0.81 | 1.10 | 1.15 | 1.16 | 0.88 | 0.64 |
| DNA polymerase epsilon subunit 3 | POLE3 | 1.42* | 1.44 | 1.47 | 1.76* | 1.10 | 0.98 | 0.64 |
| Transmembrane protein 82 | TMEM82 | 1.16 | 1.11 | 0.78 | 0.86 | 0.98 | 0.95 | 0.64 |
| Rab GTPase-activating protein 1-like | HHL | 0.91 | 0.86 | 1.11 | 0.77 | 0.78 | 0.89 | 0.64* |
| Chromobox protein homolog 1 | CBX1 | 0.86 | 1.05 | 0.93 | 0.94 | 0.76 | 0.73 | 0.64* |
| Fatty acid-binding protein, liver | FABP1 | 1.10 | 0.92 | 0.87 | 0.78* | 0.83 | 0.71* | 0.65* |
| Acidic fibroblast growth factor intracellular-binding protein | FIBP | 0.73* | 0.79 | 0.73* | 0.74 | 0.80 | 0.76 | 0.65* |
| Receptor tyrosine-protein kinase erbB-2 | ERBB2 | 0.79 | 0.87 | 0.96 | 1.03 | 0.85 | 0.87 | 0.65 |
| EGF domain-specific O-linked N-acetylglucosamine transferase | EOGT | 0.96 | 0.86 | 0.72 | 0.85 | 0.81 | 1.00 | 0.65 |
| Protein N-terminal asparagine amidohydrolase | NTAN1 | 1.23 | 0.94 | 1.05 | 0.94 | 0.70 | 0.82 | 0.65 |
| Peroxiredoxin-6 | PRDX6 | 1.15 | 1.16 | 1.01 | 0.75* | 0.82 | 0.86 | 0.65* |
| Coenzyme Q-binding protein COQ10 homolog B, mitochondrial | COQ10B | 0.88 | 1.05 | 0.84 | 0.80 | 0.74 | 0.78 | 0.65 |
| Amiloride-sensitive sodium channel subunit alpha | SCNN1A | 1.60* | 1.38 | 0.85 | 0.99 | 0.86 | 1.44 | 0.65 |
| tRNA modification GTPase GTPBP3, mitochondrial | GTPBP3 | 0.71 | 1.16 | 1.14 | 1.12 | 1.53 | 1.55 | 0.65 |
| Fatty acid desaturase 6 | FADS6 | 1.32* | 1.26 | 0.88 | 0.94 | 1.21 | 1.03 | 0.65 |
| SH2 domain-containing protein 4A | SH2D4A | 0.91 | 0.83 | 0.99 | 0.97 | 0.87 | 0.86 | 0.65 |
| Protein unc-13 homolog B | UNC13B | 0.77 | 0.75 | 0.94 | 0.80 | 0.69 | 0.74 | 0.65 |
| Phospholipid-transporting ATPase IA | ATP8A1 | 1.05 | 1.10 | 0.94 | 0.73* | 0.82 | 0.84 | 0.65* |
| Long-chain fatty acid transport protein 1 | SLC27A1 | 1.02 | 1.10 | 0.96 | 0.81 | 1.10 | 1.02 | 0.65 |
| Transmembrane protein 263 | TMEM263 | 0.67* | 0.91 | 0.93 | 0.67* | 0.71 | 0.72 | 0.65* |
| Acid sphingomyelinase-like phosphodiesterase 3a | SMPDL3A | 0.99 | 0.75 | 0.81 | 0.81 | 0.76 | 0.76 | 0.66* |
| Group XIIB secretory phospholipase A2-like protein | PLA2G12B | 1.01 | 1.25 | 0.97 | 0.86 | 0.74 | 1.13 | 0.66 |
| Mediator of RNA polymerase II transcription subunit 25 | MED25 | 0.80 | 0.95 | 0.70 | 0.87 | 0.75 | 0.84 | 0.66 |
| ATP-dependent RNA helicase DQX1 | DQX1 | 1.09 | 0.84 | 0.71 | 1.10 | 0.78 | 0.90 | 0.66 |
| ATP-binding cassette sub-family D member 3 | ABCD3 | 1.04 | 1.08 | 0.96 | 0.80* | 0.84 | 0.93 | 0.66* |
| 1-acylglycerol-3-phosphate O-acyltransferase ABHD5 | ABHD5 | 0.95 | 0.84 | 0.89 | 0.94 | 1.04 | 0.76 | 0.66 |
| Ribosomal protein S6 kinase beta-1 | RPS6KB1 | 0.98 | 1.31 | 1.08 | 0.88 | 0.77 | 0.76 | 0.66 |

| | | | | | | | | |
|---|---|---|---|---|---|---|---|---|
| Cytoplasmic phosphatidylinositol transfer protein 1 | PITPNC1 | 0.99 | 0.98 | 0.88 | 1.09 | 0.69 | 0.71 | 0.66 |
| Aldehyde dehydrogenase 1A1 | ALDH1A1 | 1.18 | 0.95 | 0.87 | 0.73* | 0.91 | 0.68* | 0.66* |
| Breakpoint cluster region protein | BCR | 0.83 | 0.60* | 0.73 | 0.72 | 0.82 | 0.81 | 0.66 |
| Transmembrane protein 256 | TMEM256 | 0.85 | 0.72 | 0.87 | 0.82 | 0.85 | 0.91 | 0.66 |
| Zinc transporter ZIP4 | SLC39A4 | 1.32* | 1.15 | 0.98 | 0.85 | 0.94 | 1.15 | 0.66 |
| Zinc finger FYVE domain-containing protein 1 | ZFYVE1 | 1.06 | 0.74 | 0.59* | 0.76 | 0.82 | 0.87 | 0.67 |
| Eukaryotic translation initiation factor 4E type 3 | EIF4E3 | 1.03 | 0.83 | 0.75 | 1.11 | 1.08 | 0.96 | 0.67 |
| Dual specificity mitogen-activated protein kinase kinase 6 | MAP2K6 | 1.09 | 1.18 | 0.99 | 0.76 | 0.72 | 0.69 | 0.67* |
| Long-chain-fatty-acid--CoA ligase 1 | ACSL1 | 1.16 | 0.88 | 0.84 | 0.79 | 0.91 | 0.73 | 0.67* |
| NF-kappa-B inhibitor alpha | NFKBIA | 1.02 | 1.07 | 1.14 | 0.89 | 0.91 | 1.27 | 0.67 |
| Transmembrane protein 35B | TMEM35B | 0.99 | 1.13 | 0.70 | 1.06 | 0.86 | 0.89 | 0.67 |

***F. Common down-regulated proteins between LPS-Cytokines alone and with Aquamin [22 proteins]***

| | | Interventions | | | | | | |
|---|---|---|---|---|---|---|---|---|
| | | Control | | | *With LPS & Cytokines* | | | |
| Proteins | Genes | AQ | AQ+MES | MES | ***LPS-Cyto*** | ***AQ*** | *AQ+MES* | *MES* |
| Rho guanine nucleotide exchange factor 40 | ARHGEF40 | 0.35* | 0.74 | 1.05 | 0.31* | 0.49* | 0.88 | 0.97 |
| Tetratricopeptide repeat protein 7A | TTC7A | 0.60* | 0.57* | 0.61* | 0.46* | 0.66 | 0.77 | 0.73 |
| Cytosolic iron-sulfur assembly component 2B | CIAO2B | 0.45* | 0.60* | 1.29 | 0.47* | 0.38* | 1.57 | 1.87 |
| Rapamycin-insensitive companion of mTOR | RICTOR | 0.52* | 0.69 | 0.57* | 0.47* | 0.57 | 0.72 | 0.69 |
| Inter-alpha-trypsin inhibitor heavy chain H1 | ITIH1 | 0.48* | 0.96 | 1.54* | 0.48* | 0.48* | 1.13 | 1.53 |
| Prolyl 3-hydroxylase 3 | P3H3 | 1.07 | 1.08 | 1.14 | 0.50* | 0.61 | 0.82 | 0.93 |
| Probable phosphoglycerate mutase 4 | PGAM4 | 0.53* | 1.15 | 2.54* | 0.51* | 0.63 | 1.94* | 2.77* |
| Ubiquitin carboxyl-terminal hydrolase 48 | USP48 | 0.66* | 0.45* | 0.72 | 0.53* | 0.62 | 0.70 | 0.74 |
| Coiled-coil domain-containing protein 9B | CCDC9B | 0.53* | 0.96 | 1.03 | 0.55* | 0.41* | 0.73 | 0.79 |
| Uncharacterized protein FLJ45252 | | 0.82 | 0.79 | 1.02 | 0.58* | 0.64 | 0.69 | 0.76 |
| Serine/threonine-protein kinase A-Raf | ARAF | 0.87 | 0.65* | 0.81 | 0.60* | 0.66 | 0.73 | 0.87 |
| Chromodomain-helicase-DNA-binding protein 1-like | CHD1L | 0.79 | 0.73 | 0.72 | 0.60* | 0.67 | 0.84 | 0.76 |
| AP-5 complex subunit beta-1 | AP5B1 | 0.82 | 0.77 | 0.66* | 0.61* | 0.56 | 0.72 | 0.69 |
| Low-density lipoprotein receptor-related protein 5 | LRP5 | 0.62* | 1.06 | 0.81 | 0.61* | 0.55 | 0.72 | 0.76 |
| Poly(A)-specific ribonuclease PARN | PARN | 0.59* | 0.68* | 0.67* | 0.62* | 0.66* | 0.71 | 0.71 |
| Pachytene checkpoint protein 2 homolog | TRIP13 | 0.77* | 0.84 | 0.92 | 0.64* | 0.65 | 0.81 | 0.73 |
| Protein FAM107B | FAM107B | 0.81 | 0.92 | 0.67* | 0.64 | 0.61 | 0.69 | 0.69 |
| Transducin beta-like protein 3 | TBL3 | 0.43* | 0.52* | 0.55* | 0.64* | 0.60 | 0.68 | 0.74 |
| AMP deaminase 3 | AMPD3 | 0.58* | 0.83 | 0.81 | 0.65 | 0.64 | 0.73 | 0.77 |
| ADP-ribosylation factor-like protein 2 | ARL2 | 0.75 | 0.69 | 0.75 | 0.66 | 0.60 | 0.68 | 0.69 |

| | | | | | | | | |
|---|---|---|---|---|---|---|---|---|
| Guanidinoacetate N-methyltransferase | GAMT | 0.55* | 1.98* | 1.89* | 0.67 | 0.57 | 1.54 | 1.94* |
| GRB10-interacting GYF protein 2 | GIGYF2 | 0.85 | 0.78 | 0.97 | 0.67* | 0.55* | 0.67 | 0.83 |

***G. Common down-regulated proteins between LPS-Cytokines alone and with Aquamin plus Mesalamine [21 proteins]***

| | | Interventions | | | | | | |
|---|---|---|---|---|---|---|---|---|
| | | Control | | | *With LPS & Cytokines* | | | |
| Proteins | Genes | AQ | AQ+MES | MES | ***LPS-Cyto*** | *AQ* | ***AQ+MES*** | *MES* |
| Cap-specific mRNA (nucleoside-2'-O-)-methyltransferase 2 | CMTR2 | 0.25* | 0.45* | 0.57* | 0.26* | 0.82 | 0.62 | 0.87 |
| Vacuolar protein sorting-associated protein 72 homolog | VPS72 | 0.62* | 0.73 | 0.82 | 0.28* | 0.92 | 0.17* | 0.70 |
| Histidine ammonia-lyase | HAL | 1.60* | 2.11* | 5.94* | 0.35* | 2.50* | 0.56 | 2.12* |
| Retroviral-like aspartic protease 1 | ASPRV1 | 0.92 | 3.16* | 10.29* | 0.39* | 1.54* | 0.49* | 1.97* |
| Keratin, type I cytoskeletal 25 | KRT25 | 0.44* | 1.22 | 0.25* | 0.41* | 1.70* | 0.23* | 1.17 |
| Alpha-2-macroglobulin-like protein 1 | A2ML1 | 1.27* | 2.46* | 5.56* | 0.41* | 1.85* | 0.34* | 1.36* |
| Cornulin | CRNN | 1.73* | 2.38* | 5.70* | 0.44* | 3.63* | 0.61 | 2.57* |
| Cystatin-M | CST6 | 1.81* | 3.67* | 3.47* | 0.54* | 1.96* | 0.58 | 1.84* |
| Mismatch repair endonuclease PMS2 | PMS2 | 0.47* | 0.47* | 0.58* | 0.54* | 1.06 | 0.65 | 0.77 |
| Myeloblastin | PRTN3 | 0.84 | 3.59* | 2.95* | 0.55* | 1.93* | 0.45* | 1.61 |
| Histone deacetylase complex subunit SAP30 | SAP30 | 0.72 | 0.63* | 0.71 | 0.57* | 0.95 | 0.62 | 0.72 |
| Protein LRATD1 | LRATD1 | 0.99 | 0.73 | 0.82 | 0.58* | 0.76 | 0.67 | 0.70 |
| DNA-directed RNA polymerase I subunit RPA1 | POLR1A | 0.74* | 0.60* | 0.57* | 0.61* | 0.73 | 0.64 | 0.71 |
| Proline-rich protein 15 | PRR15 | 0.69 | 0.84 | 1.11 | 0.61* | 0.69 | 0.62 | 0.77 |
| Protein KPLCE | KPLCE | 1.16 | 0.88 | 0.98 | 0.61* | 3.48* | 0.40* | 1.62* |
| Protein arginine N-methyltransferase 3 | PRMT3 | 0.53* | 0.56* | 0.71* | 0.61* | 0.69 | 0.66 | 0.73 |
| Helicase SRCAP | SRCAP | 0.47* | 0.38* | 0.76 | 0.62 | 0.78 | 0.59 | 0.85 |
| Ribonuclease 7 | RNASE7 | 0.79 | 0.80 | 0.67* | 0.64* | 1.10 | 0.43* | 1.35 |
| Mammaglobin-B | SCGB2A1 | 1.32 | 5.33* | 2.76* | 0.64 | 1.80* | 0.64 | 1.75 |
| Uncharacterized protein KIAA1143 | KIAA1143 | 1.23 | 0.88 | 1.10 | 0.64* | 0.71 | 0.43* | 0.72 |
| Protein-glutamine gamma-glutamyltransferase K | TGM1 | 1.13 | 1.39 | 1.71* | 0.66 | 3.12* | 0.49* | 2.44* |

***H. Common down-regulated proteins between LPS-Cytokines alone and with Mesalamine [32 proteins]***

| | | Interventions | | | | | | |
|---|---|---|---|---|---|---|---|---|
| | | Control | | | *With LPS & Cytokines* | | | |
| Proteins | Genes | AQ | AQ+MES | MES | ***LPS-Cyto*** | *AQ* | *AQ+MES* | ***MES*** |
| TBC1 domain family member 2B | TBC1D2B | 0.37* | 0.52* | 0.55* | 0.35* | 0.89 | 0.73 | 0.60 |
| Ubiquitin carboxyl-terminal hydrolase 27 | USP27X | 0.39* | 0.49* | 0.71* | 0.43* | 0.82 | 0.81 | 0.63 |

| | | | | | | | | |
|---|---|---|---|---|---|---|---|---|
| E3 ubiquitin-protein transferase MAEA | MAEA | 0.36* | 0.45* | 0.47* | 0.45* | 0.72 | 0.67 | 0.61* |
| Condensin complex subunit 3 | NCAPG | 0.58* | 0.53* | 0.54* | 0.45* | 0.97 | 0.80 | 0.67 |
| WD repeat-containing protein 37 | WDR37 | 0.44* | 0.58* | 0.50* | 0.48* | 0.72 | 0.69 | 0.64 |
| General transcription factor 3C polypeptide 1 | GTF3C1 | 0.42* | 0.44* | 0.37* | 0.48* | 0.68 | 0.68 | 0.43* |
| Vacuolar protein sorting-associated protein 18 homolog | VPS18 | 0.56* | 0.56* | 0.55* | 0.49* | 0.67* | 0.70 | 0.59* |
| AP-4 complex subunit beta-1 | AP4B1 | 0.37* | 0.55* | 0.63* | 0.50* | 0.67 | 0.68 | 0.55 |
| Pleckstrin homology domain-containing family A member 2 | PLEKHA2 | 0.73 | 0.77 | 0.79 | 0.50* | 0.71 | 0.87 | 0.39* |
| Serine incorporator 2 | SERINC2 | 0.90 | 0.86 | 0.58* | 0.50* | 0.89 | 0.86 | 0.55* |
| Prostate stem cell antigen | PSCA | 1.17 | 0.85 | 1.13 | 0.51* | 0.78 | 0.87 | 0.67 |
| Transmembrane protein 225B | TMEM225B | 0.44* | 0.39* | 0.80 | 0.51* | 0.70 | 1.23 | 0.59 |
| Pleckstrin homology domain-containing family S member 1 | PLEKHS1 | 0.77 | 1.46 | 1.10 | 0.52* | 0.69 | 0.70 | 0.17* |
| Glucocorticoid modulatory element-binding protein 2 | GMEB2 | 0.60* | 0.73 | 0.76 | 0.54* | 0.68 | 0.74 | 0.57 |
| Trafficking protein particle complex subunit 10 | TRAPPC10 | 0.69* | 0.66* | 0.63* | 0.55* | 0.71 | 0.72 | 0.54* |
| Tripartite motif-containing protein 5 | TRIM5 | 0.65* | 0.68 | 0.64* | 0.55* | 0.75 | 0.87 | 0.57 |
| CNK3/IPCEF1 fusion protein | CNK3/IPCEF1 | 1.00 | 1.11 | 0.82 | 0.56* | 0.99 | 1.03 | 0.64 |
| Nuclear factor 1 A-type | NFIA | 0.51* | 0.65* | 0.67* | 0.57* | 0.73 | 0.68 | 0.60* |
| Flavin-containing monooxygenase 5 | FMO5 | 1.01 | 0.96 | 0.80 | 0.57* | 0.78 | 0.91 | 0.58* |
| Protein pelota homolog | PELO | 0.60* | 0.72* | 0.69* | 0.59* | 0.70 | 0.70 | 0.65* |
| F-box only protein 38 | FBXO38 | 0.77 | 0.74 | 0.67* | 0.60* | 0.72 | 0.72 | 0.56* |
| DNA mismatch repair protein Msh6 | MSH6 | 0.85 | 0.62* | 0.61* | 0.60* | 0.81 | 0.76 | 0.63 |
| Serine/threonine-protein kinase 17B | STK17B | 1.11 | 1.30 | 0.93 | 0.61* | 0.76 | 0.73 | 0.48* |
| Peptide chain release factor 1, mitochondrial | MTRF1 | 1.00 | 1.14 | 1.03 | 0.62* | 0.95 | 1.03 | 0.40* |
| Unconventional myosin-Vc | MYO5C | 0.66 | 0.63* | 0.70 | 0.62* | 0.67 | 0.78 | 0.66 |
| 17-beta-hydroxysteroid dehydrogenase type 2 | HSD17B2 | 1.40* | 1.31* | 0.98 | 0.63* | 0.70* | 0.67* | 0.42* |
| Transmembrane 4 L6 family member 20 | TM4SF20 | 1.61* | 0.93 | 0.65* | 0.63* | 1.51* | 0.97 | 0.50* |
| Tripartite motif-containing protein 3 | TRIM3 | 0.85 | 0.53* | 0.45* | 0.64* | 0.79 | 0.92 | 0.52 |
| Ras-related protein Rab-3B | RAB3B | 0.90 | 0.94 | 1.15 | 0.64* | 0.81 | 0.78 | 0.61 |
| Desmocollin-3 | DSC3 | 1.72* | 9.40* | 34.18* | 0.65 | 1.84* | 0.93 | 0.01* |
| EEF1A lysine methyltransferase 1 | EEF1AKMT1 | 0.97 | 1.13 | 1.04 | 0.65* | 0.75 | 0.70 | 0.63 |
| Calpain-7 | CAPN7 | 0.75 | 0.70 | 0.69* | 0.66* | 0.76 | 0.71 | 0.61 |

***I. Common down-regulated proteins among LPS-Cytokines alone, with Aquamin and with Aquamin plus Mesalamine [23 proteins]***

| | | Interventions | | | | | | |
|---|---|---|---|---|---|---|---|---|
| | | Control | | | *With LPS & Cytokines* | | | |
| Proteins | Genes | AQ | AQ+MES | MES | ***LPS-Cyto*** | ***AQ*** | ***AQ+MES*** | *MES* |

| | | | | | | | | |
|---|---|---|---|---|---|---|---|---|
| PHD finger protein 6 | PHF6 | 0.32* | 0.43* | 0.46* | 0.26* | 0.65* | 0.64* | 0.69* |
| AN1-type zinc finger protein 1 | ZFAND1 | 0.53* | 0.50* | 0.58* | 0.40* | 0.41* | 0.34* | 0.71 |
| Splicing factor, suppressor of white-apricot homolog | SFSWAP | 0.77 | 0.87 | 0.71* | 0.42* | 0.48* | 0.56 | 0.78 |
| ATP-dependent RNA helicase DDX39A | DDX39A | 0.72 | 0.78 | 0.83 | 0.48* | 0.52* | 0.64 | 0.68 |
| tRNA (guanine(26)-N(2))-dimethyltransferase | TRMT1 | 0.64* | 0.82 | 0.82 | 0.50* | 0.50* | 0.39* | 0.72 |
| Synaptopodin | SYNPO | 0.94 | 1.12 | 1.25 | 0.56* | 0.48* | 0.64 | 0.88 |
| Ubiquitin carboxyl-terminal hydrolase 16 | USP16 | 0.76* | 0.88 | 0.87 | 0.56* | 0.59* | 0.54* | 0.80 |
| FAST kinase domain-containing protein 1, mitochondrial | FASTKD1 | 0.94 | 0.96 | 0.93 | 0.58* | 0.31* | 0.45* | 0.82 |
| Refilin-A | RFLNA | 1.15 | 1.15 | 1.16 | 0.58* | 0.61 | 0.65 | 0.94 |
| Prothymosin alpha | PTMA | 0.77 | 0.65* | 1.07 | 0.59* | 0.45* | 0.66 | 1.38* |
| Ribonucleoprotein PTB-binding 2 | RAVER2 | 0.86 | 1.47 | 1.25 | 0.59* | 0.57 | 0.57 | 1.71 |
| POTE ankyrin domain family member E | POTEE | 1.02 | 0.49* | 0.95 | 0.59* | 0.50* | 0.51* | 0.90 |
| Atypical kinase COQ8B, mitochondrial | COQ8B | 0.71 | 0.60* | 0.66* | 0.60* | 0.50* | 0.61 | 0.72 |
| Protein FAM117B | FAM117B | 0.47* | 0.87 | 0.70 | 0.60* | 0.12* | 0.28* | 0.71 |
| DNA mismatch repair protein Msh3 | MSH3 | 0.66* | 0.67* | 0.70* | 0.60* | 0.66 | 0.65 | 0.69 |
| Hyccin 2 | HYCC2 | 0.90 | 0.89 | 0.69 | 0.60* | 0.58 | 0.62 | 0.71 |
| FH1/FH2 domain-containing protein 1 | FHOD1 | 0.77 | 0.69 | 0.76 | 0.61* | 0.59 | 0.63 | 0.67 |
| Secretion-regulating guanine nucleotide exchange factor | SERGEF | 0.98 | 1.22 | 1.06 | 0.62* | 0.61 | 0.61 | 0.82 |
| Overexpressed in colon carcinoma 1 protein | OCC1 | 0.65* | 0.70 | 0.87 | 0.63* | 0.55* | 0.62 | 0.87 |
| Cytosolic carboxypeptidase 1 | AGTPBP1 | 0.58* | 0.73 | 0.86 | 0.64* | 0.62 | 0.35* | 0.67 |
| Serine/threonine-protein kinase LMTK2 | LMTK2 | 0.80 | 0.95 | 0.98 | 0.65* | 0.60 | 0.63 | 0.68 |
| Protein Tob2 | TOB2 | 0.73 | 0.84 | 0.54* | 0.66 | 0.35* | 0.37* | 0.95 |
| ATP-dependent RNA helicase DDX51 | DDX51 | 0.64* | 0.66* | 0.72* | 0.66* | 0.55* | 0.60 | 0.70 |

***J. Common down-regulated proteins among LPS-Cytokines alone, with Aquamin and with Mesalamine [25 proteins]***

| | | Interventions | | | | | | |
|---|---|---|---|---|---|---|---|---|
| | | Control | | | *With LPS & Cytokines* | | | |
| Proteins | Genes | AQ | AQ+MES | MES | ***LPS-Cyto*** | ***AQ*** | *AQ+MES* | ***MES*** |
| Dynein axonemal heavy chain 8 | DNAH8 | 0.65* | 0.96 | 0.58* | 0.19* | 0.36* | 2.06* | 0.37* |
| Replication factor C subunit 5 | RFC5 | 0.51* | 0.49* | 0.62* | 0.29* | 0.55* | 0.70 | 0.43* |
| Tumor necrosis factor receptor superfamily member 10D | TNFRSF10D | 0.96 | 1.40* | 1.27 | 0.41* | 0.56* | 1.02 | 0.45* |
| Structural maintenance of chromosomes protein 2 | SMC2 | 0.55* | 0.44* | 0.54* | 0.42* | 0.65 | 0.74 | 0.48* |
| Heat shock protein beta-1 | HSPB1 | 1.21* | 1.07 | 0.98 | 0.50* | 0.60* | 0.67* | 0.56* |
| Deoxynucleotidyltransferase terminal-interacting protein 2 | DNTTIP2 | 0.57* | 0.77 | 0.76 | 0.51* | 0.67 | 0.67 | 0.56* |

| | | | | | | | | |
|---|---|---|---|---|---|---|---|---|
| Helicase with zinc finger domain 2 | HELZ2 | 0.61* | 0.88 | 0.77 | 0.53* | 0.36* | 0.97 | 0.57 |
| Conserved oligomeric Golgi complex subunit 6 | COG6 | 0.87 | 1.08 | 0.80 | 0.55* | 0.59 | 0.70 | 0.45* |
| Membrane-bound transcription factor site-1 protease | MBTPS1 | 0.93 | 0.92 | 1.00 | 0.56* | 0.56 | 0.69 | 0.60 |
| SUN domain-containing protein 1 | SUN1 | 0.53* | 0.64* | 0.58* | 0.57* | 0.65 | 0.69 | 0.49* |
| NTF2-related export protein 2 | NXT2 | 1.07 | 1.05 | 0.93 | 0.57* | 0.44* | 0.78 | 0.66 |
| Trinucleotide repeat-containing gene 6B protein | TNRC6B | 1.51* | 1.07 | 0.90 | 0.59* | 0.65 | 1.27 | 0.62 |
| Zinc finger FYVE domain-containing protein 16 | ZFYVE16 | 0.52* | 0.76 | 0.62* | 0.59* | 0.40* | 0.71 | 0.60 |
| Phosphomannomutase 1 | PMM1 | 0.91 | 1.20 | 1.12 | 0.61* | 0.65 | 0.73 | 0.49* |
| TBC1 domain family member 8B | TBC1D8B | 0.65* | 0.82 | 0.74* | 0.61* | 0.63 | 0.67 | 0.63 |
| Protein kish-A | TMEM167A | 0.87 | 0.92 | 0.94 | 0.62* | 0.60* | 1.03 | 0.64 |
| RNA-binding protein 12B | RBM12B | 0.80 | 0.82 | 0.78 | 0.63 | 0.58 | 0.68 | 0.58 |
| Complement C1r subcomponent | C1R | 0.94 | 1.31 | 14.51* | 0.63 | 0.54 | 1.27 | 0.15* |
| Threonine synthase-like 1 | THNSL1 | 0.88 | 1.16 | 1.27 | 0.63* | 0.62 | 0.71 | 0.66 |
| Sulfate transporter | SLC26A2 | 2.55* | 1.80* | 0.90 | 0.65* | 0.48* | 0.77 | 0.44* |
| Proteasome subunit beta type-5 | PSMB5 | 0.90 | 0.81 | 0.84 | 0.65* | 0.60 | 0.71 | 0.57* |
| Probable proline--tRNA ligase, mitochondrial | PARS2 | 0.84 | 0.87 | 0.88 | 0.65* | 0.55* | 0.68 | 0.51* |
| Telomerase-binding protein EST1A | SMG6 | 0.65* | 0.61* | 0.87 | 0.65 | 0.60 | 0.67 | 0.66 |
| Zinc finger protein 718 | ZNF718 | 0.75 | 0.71 | 0.69* | 0.65* | 0.60 | 0.70 | 0.63 |
| Ras and Rab interactor 2 | RIN2 | 0.79 | 0.80 | 0.71 | 0.66 | 0.56 | 0.72 | 0.55 |

***K. Common down-regulated proteins among LPS-Cytokines alone, with Aquamin plus Mesalamine and with Mesalamine [40 proteins]***

| | | Interventions | | | | | | |
|---|---|---|---|---|---|---|---|---|
| | | Control | | | *With LPS & Cytokines* | | | |
| Proteins | Genes | AQ | AQ+MES | MES | ***LPS-Cyto*** | *AQ* | ***AQ+MES*** | ***MES*** |
| Primary cilium assembly protein FAM149B1 | FAM149B1 | 0.29* | 0.24* | 0.24* | 0.30* | 0.85 | 0.39* | 0.60 |
| Putative ATP-dependent RNA helicase DHX57 | DHX57 | 0.45* | 0.47* | 0.46* | 0.30* | 0.71 | 0.63 | 0.46* |
| SWI/SNF-related matrix-associated actin-dependent regulator of chromatin subfamily A-like protein 1 | SMARCAL1 | 0.57* | 0.55* | 0.67* | 0.31* | 0.77 | 0.53* | 0.44* |
| Myotubularin-related protein 13 | SBF2 | 0.39* | 0.59* | 0.60* | 0.32* | 0.67 | 0.42* | 0.40* |
| E3 ubiquitin-protein ligase NRDP1 | RNF41 | 0.43* | 0.55* | 0.45* | 0.33* | 0.81 | 0.61 | 0.52 |
| ADP-ribosylation factor-binding protein GGA2 | GGA2 | 0.61* | 0.80 | 0.81 | 0.40* | 0.69 | 0.44* | 0.48* |
| Mitogen-activated protein kinase kinase kinase 20 | MAP3K20 | 0.44* | 0.46* | 0.61* | 0.40* | 0.79 | 0.65 | 0.61* |
| Monocarboxylate transporter 6 | SLC16A5 | 0.77 | 0.92 | 0.78 | 0.43* | 0.68 | 0.55 | 0.46* |
| Tensin-3 | TNS3 | 0.53* | 0.80 | 0.74* | 0.43* | 0.72 | 0.61 | 0.47* |
| Tensin-4 | TNS4 | 0.30* | 0.47* | 0.46* | 0.43* | 0.67 | 0.52* | 0.45* |
| Serine/threonine-protein phosphatase 6 regulatory ankyrin repeat subunit A | ANKRD28 | 0.46* | 0.44* | 0.46* | 0.44* | 0.67 | 0.55 | 0.51* |
| Mitogen-activated protein kinase 7 | MAPK7 | 0.47* | 0.62* | 0.67* | 0.45* | 0.68 | 0.63 | 0.63 |

| | | | | | | | | |
|---|---|---|---|---|---|---|---|---|
| Arylamine N-acetyltransferase 1 | NAT1 | 0.88 | 0.42* | 0.46* | 0.46* | 0.77 | 0.60 | 0.62 |
| Serine/threonine-protein phosphatase 6 regulatory ankyrin repeat subunit C | ANKRD52 | 0.54* | 0.63 | 0.71 | 0.49* | 0.75 | 0.59 | 0.49* |
| Probable JmjC domain-containing histone demethylation protein 2C | JMJD1C | 0.47* | 0.42* | 0.33* | 0.50* | 0.72 | 0.42* | 0.29* |
| Meprin A subunit alpha | MEP1A | 1.35* | 1.47* | 0.85 | 0.50* | 0.69* | 0.57* | 0.47* |
| Atypical kinase COQ8A, mitochondrial | COQ8A | 0.82 | 1.19 | 1.04 | 0.52* | 0.72 | 0.62 | 0.45* |
| Leucine-rich alpha-2-glycoprotein | LRG1 | 1.03 | 0.46* | 0.83 | 0.52* | 0.97 | 0.66 | 0.54* |
| GTP-binding protein 10 | GTPBP10 | 1.01 | 0.90 | 0.62* | 0.52* | 0.78 | 0.66* | 0.52* |
| Ubiquitin carboxyl-terminal hydrolase 3 | USP3 | 0.63* | 0.71 | 0.70 | 0.52* | 0.67 | 0.41* | 0.28* |
| Smad nuclear-interacting protein 1 | SNIP1 | 0.49* | 0.69 | 0.71 | 0.53* | 0.69 | 0.66 | 0.48* |
| Cleavage and polyadenylation specificity factor subunit 4 | CPSF4 | 0.61* | 0.69* | 0.66* | 0.55* | 0.69 | 0.65 | 0.61* |
| Intelectin-2 | ITLN2 | 1.04 | 1.01 | 0.45* | 0.56* | 1.13 | 0.38* | 0.42* |
| Protein-arginine deiminase type-2 | PADI2 | 1.11 | 1.05 | 0.91 | 0.57* | 0.73 | 0.64* | 0.53* |
| DDB1- and CUL4-associated factor 13 | DCAF13 | 0.82* | 0.72* | 0.75* | 0.58* | 0.70 | 0.63* | 0.65* |
| RNA-binding protein 6 | RBM6 | 0.84 | 0.77 | 0.79 | 0.58* | 0.73 | 0.64 | 0.51* |
| Pleckstrin homology domain-containing family A member 4 | PLEKHA4 | 1.12 | 0.99 | 1.45 | 0.58* | 0.71 | 0.54 | 0.62 |
| Cytosolic iron-sulfur assembly component 3 | CIAO3 | 0.48* | 0.47* | 0.55* | 0.58* | 0.69 | 0.48* | 0.58* |
| Cytochrome c oxidase assembly factor 4 homolog, mitochondrial | COA4 | 1.02 | 1.08 | 0.99 | 0.59* | 0.81 | 0.61 | 0.63 |
| Annexin A13 | ANXA13 | 1.19* | 0.98 | 0.76* | 0.60* | 0.83 | 0.55* | 0.39* |
| Hydroxymethylglutaryl-CoA synthase, mitochondrial | HMGCS2 | 1.33* | 1.13 | 0.88 | 0.61* | 0.73* | 0.66* | 0.55* |
| Sulfotransferase 1B1 | SULT1B1 | 1.02 | 0.90 | 0.84 | 0.61* | 0.67 | 0.54* | 0.47* |
| 5'-AMP-activated protein kinase subunit gamma-2 | PRKAG2 | 0.84 | 1.01 | 0.80 | 0.62* | 0.81 | 0.57 | 0.48* |
| Mucosa-associated lymphoid tissue lymphoma translocation protein 1 | MALT1 | 0.55* | 0.63 | 0.79 | 0.62 | 0.67 | 0.61 | 0.58 |
| Poly(rC)-binding protein 3 | PCBP3 | 1.00 | 0.97 | 0.95 | 0.63 | 0.72 | 0.66 | 0.58 |
| Transferrin receptor protein 1 | TFRC | 1.04 | 0.83 | 0.67* | 0.63* | 0.93 | 0.58* | 0.52* |
| Phosphoinositide 3-kinase regulatory subunit 4 | PIK3R4 | 0.61* | 0.65* | 0.59* | 0.64* | 0.72 | 0.62 | 0.58 |
| Carbonic anhydrase 1 | CA1 | 0.97 | 0.86 | 0.67* | 0.66* | 0.70 | 0.66 | 0.66* |
| Lactase/phlorizin hydrolase | LCT | 0.79 | 0.74 | 1.16 | 0.66 | 0.69 | 0.48* | 0.39* |
| Steroid hormone receptor ERR1 | ESRRA | 0.67* | 0.77 | 0.76 | 0.66 | 0.67 | 0.61 | 0.61 |

***L. Common down-regulated proteins between LPS-Cytokines with Aquamin and with Aquamin plus Mesalamine [123 proteins]***

| | | Interventions | | | | | | |
|---|---|---|---|---|---|---|---|---|
| | | Control | | | *With LPS & Cytokines* | | | |
| Proteins | Genes | AQ | AQ+MES | MES | *LPS-Cyto* | ***AQ*** | ***AQ+MES*** | *MES* |

| | | | | | | | | |
|---|---|---|---|---|---|---|---|---|
| Nuclear speckle splicing regulatory protein 1 | NSRP1 | 1.69* | 2.06* | 2.33* | 1.09 | 0.02* | 0.40* | 1.14 |
| Nucleolar protein 7 | NOL7 | 1.42* | 1.20 | 1.38* | 1.46* | 0.21* | 0.42* | 0.68 |
| Fos-related antigen 1 | FOSL1 | 0.58* | 0.55* | 0.85 | 0.79 | 0.21* | 0.42* | 0.78 |
| Protein max | MAX | 1.28* | 1.31 | 1.42* | 1.85* | 0.24* | 0.39* | 0.75 |
| Vesicle transport protein SFT2C | SFT2D3 | 0.91 | 1.08 | 0.75* | 1.19 | 0.25* | 0.28* | 0.85 |
| Transmembrane protein 51 | TMEM51 | 1.19 | 1.38 | 1.70* | 1.29 | 0.27* | 0.66 | 0.80 |
| Phosphorylated adapter RNA export protein | PHAX | 1.12 | 0.93 | 1.21 | 0.85 | 0.29* | 0.50 | 0.80 |
| Protein Dr1 | DR1 | 1.21 | 1.20 | 1.30 | 1.10 | 0.29* | 0.62 | 0.80 |
| RNA exonuclease 4 | REXO4 | 1.08 | 0.91 | 1.02 | 1.10 | 0.29* | 0.36* | 0.80 |
| Zinc finger CCCH domain-containing protein 4 | ZC3H4 | 1.14 | 1.02 | 1.15 | 1.18 | 0.29* | 0.57 | 0.81 |
| U6 snRNA-associated Sm-like protein LSm5 | LSM5 | 1.17 | 1.00 | 1.01 | 1.07 | 0.30* | 0.48* | 0.68 |
| Microfibrillar-associated protein 1 | MFAP1 | 1.11 | 0.96 | 1.09 | 1.09 | 0.31* | 0.38* | 0.68* |
| Translation machinery-associated protein 16 | TMA16 | 0.81 | 0.94 | 1.04 | 1.01 | 0.34* | 0.54* | 0.83 |
| Protein ELYS | AHCTF1 | 0.73* | 0.88 | 0.87 | 0.95 | 0.35* | 0.29* | 0.76 |
| Extracellular sulfatase Sulf-2 | SULF2 | 0.59* | 0.93 | 1.25* | 0.86 | 0.35* | 0.37* | 0.73* |
| Mediator of DNA damage checkpoint protein 1 | MDC1 | 0.91 | 0.65 | 0.62* | 0.70 | 0.35* | 0.50* | 1.13 |
| Zinc finger protein ubi-d4 | DPF2 | 0.75 | 0.98 | 0.71 | 0.73 | 0.35* | 0.50* | 0.81 |
| KRR1 small subunit processome component homolog | KRR1 | 1.24* | 1.05 | 1.20* | 0.95 | 0.35* | 0.43* | 0.82 |
| U3 small nucleolar RNA-associated protein 14 homolog A | UTP14A | 0.74 | 0.96 | 1.11 | 1.00 | 0.35* | 0.52* | 0.74 |
| Nuclear receptor subfamily 2 group F member 6 | NR2F6 | 0.53* | 0.55* | 0.85 | 1.03 | 0.36* | 0.53 | 0.69 |
| KICSTOR subunit 2 | KICS2 | 0.48* | 0.38* | 1.09 | 1.29 | 0.39* | 0.38* | 0.78 |
| U4/U6.U5 small nuclear ribonucleoprotein 27 kDa protein | SNRNP27 | 1.30 | 0.82 | 1.06 | 1.32 | 0.39* | 0.59 | 0.89 |
| Nucleolar complex protein 3 homolog | NOC3L | 0.55* | 0.80 | 0.98 | 1.23 | 0.41* | 0.42* | 0.82 |
| Mth938 domain-containing protein | AAMDC | 0.97 | 1.03 | 0.99 | 0.89 | 0.42* | 0.55* | 0.67* |
| YLP motif-containing protein 1 | YLPM1 | 0.84 | 0.74 | 0.93 | 0.85 | 0.42* | 0.46* | 0.75 |
| Thioredoxin-like protein 4A | TXNL4A | 1.23* | 1.22 | 1.01 | 1.41* | 0.42* | 0.56* | 0.75 |
| Protein FRG1 | FRG1 | 0.78 | 0.72 | 0.85 | 0.69 | 0.43* | 0.44* | 0.68 |
| Lysine-specific demethylase 2A | KDM2A | 0.49* | 0.82 | 0.77 | 0.73 | 0.44* | 0.45* | 0.70 |
| RWD domain-containing protein 1 | RWDD1 | 1.11 | 1.03 | 1.05 | 1.13 | 0.44* | 0.58* | 0.71 |
| Translation machinery-associated protein 7 | TMA7 | 0.44* | 0.46* | 0.65* | 0.80 | 0.44* | 0.40* | 0.71 |
| Receptor-interacting serine/threonine-protein kinase 2 | RIPK2 | 0.69 | 0.83 | 0.85 | 1.23 | 0.45* | 0.63 | 0.68 |
| ATP-dependent RNA helicase DHX8 | DHX8 | 0.85 | 1.18 | 1.11 | 1.00 | 0.45* | 0.65 | 0.87 |
| Mitotic-spindle organizing protein 2B | MZT2B | 0.98 | 0.92 | 1.03 | 0.95 | 0.45* | 0.63 | 0.74 |
| Ribosome biogenesis regulatory protein homolog | RRS1 | 0.57* | 0.64* | 0.98 | 1.07 | 0.46* | 0.53* | 0.78 |
| Deoxyhypusine hydroxylase | DOHH | 0.99 | 1.19 | 1.04 | 1.07 | 0.46* | 0.59 | 0.97 |
| Receptor-interacting serine/threonine-protein kinase 3 | RIPK3 | 0.92 | 0.79 | 0.97 | 0.85 | 0.46* | 0.58* | 0.69* |

| | | | | | | | | |
|---|---|---|---|---|---|---|---|---|
| Histone chaperone ASF1A | ASF1A | 0.86 | 0.97 | 0.99 | 1.00 | 0.46* | 0.54* | 0.79 |
| SERPINE1 mRNA-binding protein 1 | SERBP1 | 0.81* | 0.82 | 1.07 | 0.78 | 0.47* | 0.51* | 0.70* |
| B-cell linker protein | BLNK | 0.97 | 1.17 | 1.10 | 1.18 | 0.47* | 0.53* | 0.86 |
| Junctophilin-1 | JPH1 | 1.14 | 0.54* | 1.41 | 1.04 | 0.48* | 0.49* | 1.10 |
| PC4 and SFRS1-interacting protein | PSIP1 | 1.51* | 1.06 | 1.31* | 1.61* | 0.48* | 0.47* | 1.46* |
| Large ribosomal subunit protein mL55 | MRPL55 | 1.09 | 1.07 | 1.04 | 0.96 | 0.48* | 0.57* | 0.69 |
| Transcription initiation factor IIE subunit beta | GTF2E2 | 0.97 | 1.01 | 1.04 | 1.06 | 0.48* | 0.58 | 0.75 |
| Proliferation marker protein Ki-67 | MKI67 | 0.53* | 0.44* | 0.69 | 0.89 | 0.49* | 0.62 | 1.02 |
| Oxidoreductase-like domain-containing protein 1 | OXLD1 | 1.07 | 1.12 | 1.14 | 1.01 | 0.49* | 0.52* | 0.75 |
| Ubiquitin-conjugating enzyme E2 R2 | UBE2R2 | 1.01 | 1.25 | 1.22 | 1.37 | 0.50* | 0.62 | 1.04 |
| WD repeat-containing protein 55 | WDR55 | 0.88 | 0.60* | 0.71 | 0.76 | 0.50* | 0.65 | 0.79 |
| Death-inducer obliterator 1 | DIDO1 | 0.93 | 1.03 | 1.03 | 0.98 | 0.50* | 0.51 | 0.98 |
| Ribosome biogenesis protein BMS1 homolog | BMS1 | 0.71 | 0.65 | 0.98 | 0.73 | 0.51* | 0.57 | 0.79 |
| ATP-dependent RNA helicase DDX24 | DDX24 | 0.56* | 0.55* | 0.62* | 0.81 | 0.51* | 0.51* | 0.67* |
| Envoplakin | EVPL | 0.87 | 0.87 | 1.06 | 0.80 | 0.52* | 0.56* | 0.74 |
| HEAT repeat-containing protein 3 | HEATR3 | 0.74 | 0.44* | 0.85 | 0.72 | 0.52 | 0.37* | 0.80 |
| Nuclear transcription factor Y subunit alpha | NFYA | 1.20 | 1.08 | 1.17 | 1.55* | 0.52* | 0.32* | 0.82 |
| PEST proteolytic signal-containing nuclear protein | PCNP | 0.88 | 0.65* | 0.93 | 0.78 | 0.53* | 0.64 | 0.76 |
| Survival of motor neuron-related-splicing factor 30 | SMNDC1 | 1.02 | 1.21 | 1.27 | 1.27 | 0.53* | 0.65 | 0.91 |
| Large ribosomal subunit protein eL29 | RPL29 | 0.89 | 0.85 | 1.02 | 1.01 | 0.53* | 0.65* | 0.78 |
| Phosphopantothenoylcysteine decarboxylase | PPCDC | 0.91 | 0.83 | 0.80 | 1.01 | 0.53* | 0.49* | 0.69 |
| Chromobox protein homolog 5 | CBX5 | 1.01 | 0.95 | 1.04 | 0.82 | 0.53* | 0.60* | 0.72 |
| Zinc finger protein 638 | ZNF638 | 0.68* | 0.84 | 0.84 | 0.71 | 0.53* | 0.62 | 0.78 |
| RNA polymerase II-associated factor 1 homolog | PAF1 | 0.83 | 0.80 | 0.85 | 0.92 | 0.53* | 0.58 | 0.75 |
| Protein LSM14 homolog B | LSM14B | 0.75 | 0.77 | 1.15 | 0.88 | 0.54 | 0.42* | 0.89 |
| Insulin-like growth factor 2 mRNA-binding protein 3 | IGF2BP3 | 1.18 | 0.90 | 0.98 | 0.88 | 0.54* | 0.57* | 0.72 |
| High mobility group protein HMGI-C | HMGA2 | 0.68 | 0.54* | 0.81 | 0.74 | 0.54 | 0.64 | 0.71 |
| Nucleolar and spindle-associated protein 1 | NUSAP1 | 1.44* | 1.64* | 1.78* | 1.33 | 0.54 | 0.27* | 0.86 |
| 7SK snRNA methylphosphate capping enzyme | MEPCE | 0.76 | 0.81 | 0.96 | 0.78 | 0.55 | 0.62 | 0.74 |
| Pumilio homolog 1 | PUM1 | 0.85 | 0.82 | 0.93 | 0.70 | 0.55* | 0.66 | 0.71 |
| Probable RNA-binding protein 19 | RBM19 | 0.69* | 0.75 | 0.83 | 0.70 | 0.55* | 0.54* | 0.74 |
| Signal transducing adapter molecule 1 | STAM | 0.89 | 0.89 | 1.02 | 0.83 | 0.55* | 0.59* | 0.92 |
| Calcium homeostasis endoplasmic reticulum protein | CHERP | 0.82 | 0.84 | 0.95 | 0.90 | 0.55* | 0.65* | 0.91 |
| Ladinin-1 | LAD1 | 0.97 | 0.78* | 0.94 | 0.69* | 0.55* | 0.50* | 0.71* |
| Cancer-related nucleoside-triphosphatase | NTPCR | 0.70* | 0.79 | 0.64* | 0.86 | 0.55* | 0.65 | 0.85 |
| Keratin, type II cuticular Hb5 | KRT85 | 0.66* | 8.93* | 1.06 | 0.69* | 0.55* | 0.40* | 1.22 |
| Spondin-1 | SPON1 | 0.67* | 0.66* | 1.05 | 0.73* | 0.56* | 0.49* | 0.69* |

| | | | | | | | | |
|---|---|---|---|---|---|---|---|---|
| Protein AHNAK2 | AHNAK2 | 0.92 | 0.86 | 0.88 | 0.74* | 0.57* | 0.49* | 0.68* |
| Mortality factor 4-like protein 1 | MORF4L1 | 0.86 | 1.13 | 1.08 | 0.98 | 0.57 | 0.59 | 0.88 |
| Regulator of nonsense transcripts 3B | UPF3B | 0.93 | 0.84 | 0.83 | 1.03 | 0.57 | 0.50* | 0.75 |
| Ribosome production factor 2 homolog | RPF2 | 0.73* | 0.71* | 0.77 | 0.81 | 0.57* | 0.62 | 0.76 |
| Neuroblast differentiation-associated protein AHNAK | AHNAK | 0.93 | 0.93 | 1.07 | 0.69* | 0.58* | 0.60* | 0.75 |
| FAS-associated factor 1 | FAF1 | 0.96 | 0.81 | 0.77 | 1.04 | 0.58 | 0.59 | 0.69 |
| Protein PAXX | PAXX | 1.12 | 0.97 | 1.07 | 1.00 | 0.58 | 0.64 | 0.77 |
| Histone H2B type 1-B | H2BC3 | 0.66* | 0.83 | 0.98 | 1.05 | 0.58 | 0.63 | 0.97 |
| RNA-binding motif, single-stranded-interacting protein 2 | RBMS2 | 1.06 | 0.91 | 1.23 | 0.90 | 0.58 | 0.65 | 0.83 |
| Glia maturation factor gamma | GMFG | 1.27 | 1.38 | 1.29 | 1.34 | 0.59 | 0.62 | 0.74 |
| Negative elongation factor E | NELFE | 0.80 | 0.86 | 1.02 | 0.81 | 0.59 | 0.61 | 0.71 |
| Mediator of RNA polymerase II transcription subunit 15 | MED15 | 0.90 | 0.70 | 0.92 | 0.76 | 0.59 | 0.56 | 0.82 |
| KICSTOR complex protein ITFG2 | ITFG2 | 0.90 | 0.77 | 0.67* | 0.92 | 0.59 | 0.61 | 0.81 |
| TOX high mobility group box family member 4 | TOX4 | 0.76* | 0.97 | 0.96 | 0.95 | 0.59* | 0.60 | 0.67* |
| PWWP domain-containing protein 2A | PWWP2A | 1.12 | 1.65* | 1.22 | 1.07 | 0.59 | 0.61 | 0.74 |
| TOM1-like protein 1 | TOM1L1 | 0.99 | 0.93 | 0.92 | 1.00 | 0.59* | 0.61 | 0.76 |
| F-box/LRR-repeat protein 18 | FBXL18 | 0.74* | 0.59* | 0.64* | 0.70 | 0.60 | 0.61 | 0.67 |
| Arginyl-tRNA--protein transferase 1 | ATE1 | 0.90 | 1.01 | 1.07 | 0.68 | 0.60 | 0.60 | 0.67 |
| Histone-lysine N-methyltransferase SETD1A | SETD1A | 0.61* | 0.70 | 0.79 | 0.76 | 0.60 | 0.55 | 0.68 |
| Alpha-taxilin | TXLNA | 0.77* | 0.83 | 0.89 | 0.78* | 0.60* | 0.64* | 0.84 |
| [F-actin]-monooxygenase MICAL3 | MICAL3 | 1.05 | 1.30 | 1.09 | 0.84 | 0.60 | 0.62 | 0.74 |
| DNA polymerase delta catalytic subunit | POLD1 | 0.62* | 0.55* | 0.53* | 0.73 | 0.61 | 0.62 | 0.85 |
| Allograft inflammatory factor 1-like | AIF1L | 1.33* | 0.98 | 1.34* | 2.00* | 0.61* | 0.64 | 1.17 |
| Ubiquitin-conjugating enzyme E2 R1 | CDC34 | 0.89 | 0.78 | 1.01 | 0.88 | 0.61 | 0.63 | 0.95 |
| Krueppel-like factor 5 | KLF5 | 0.83 | 0.76 | 0.82 | 0.67* | 0.61* | 0.63* | 0.69* |
| Insulin-like growth factor-binding protein 2 | IGFBP2 | 1.02 | 1.19 | 1.29 | 0.69 | 0.62 | 0.57 | 0.83 |
| Coiled-coil domain-containing protein 50 | CCDC50 | 0.65* | 0.81 | 0.99 | 1.02 | 0.62 | 0.67 | 1.29 |
| Eukaryotic translation initiation factor 4B | EIF4B | 0.92 | 0.78 | 1.00 | 0.78* | 0.62* | 0.60* | 0.77 |
| Methyl-CpG-binding domain protein 2 | MBD2 | 0.74 | 0.56* | 0.76 | 0.69 | 0.63 | 0.60 | 0.68 |
| Protein strawberry notch homolog 1 | SBNO1 | 0.73 | 0.68 | 0.89 | 0.70 | 0.63 | 0.56 | 0.90 |
| Nucleoredoxin | NXN | 0.92 | 0.89 | 1.01 | 0.93 | 0.63 | 0.67 | 0.72 |
| Centrosomal protein of 170 kDa | CEP170 | 0.89 | 1.03 | 1.10 | 1.06 | 0.63 | 0.57 | 0.98 |
| Microtubule-associated protein RP/EB family member 2 | MAPRE2 | 0.71 | 0.65* | 0.80 | 0.83 | 0.63 | 0.66 | 0.75 |
| Cytoplasmic polyadenylation element-binding protein 2 | CPEB2 | 0.79 | 1.18 | 1.09 | 0.69 | 0.63 | 0.60 | 0.67 |
| BAG family molecular chaperone regulator 3 | BAG3 | 0.91 | 0.84 | 1.06 | 0.89 | 0.63 | 0.58* | 0.89 |
| NudC domain-containing protein 2 | NUDCD2 | 0.81 | 0.88 | 0.94 | 0.71 | 0.64 | 0.63 | 0.68 |

| | | | | | | | | |
|---|---|---|---|---|---|---|---|---|
| Tuftelin-interacting protein 11 | TFIP11 | 0.90 | 0.97 | 1.04 | 0.90 | 0.64 | 0.62 | 0.85 |
| Histone-lysine N-methyltransferase SETD7 | SETD7 | 0.89 | 0.70 | 1.06 | 0.91 | 0.64 | 0.63 | 0.70 |
| Sciellin | SCEL | 0.81* | 0.77* | 1.09 | 0.81 | 0.64* | 0.64* | 0.86 |
| BMP and activin membrane-bound inhibitor homolog | BAMBI | 1.00 | 1.31* | 0.93 | 1.39* | 0.64* | 0.61* | 1.07 |
| Probable ATP-dependent RNA helicase DDX41 | DDX41 | 0.76* | 0.85 | 0.83 | 0.79 | 0.65 | 0.65 | 0.78 |
| Sorbin and SH3 domain-containing protein 2 | SORBS2 | 1.28 | 1.02 | 0.92 | 0.96 | 0.65 | 0.55 | 0.71 |
| MKI67 FHA domain-interacting nucleolar phosphoprotein | NIFK | 0.99 | 0.98 | 1.05 | 1.01 | 0.65 | 0.65 | 0.83 |
| Macoilin | MACO1 | 1.00 | 0.99 | 1.06 | 0.97 | 0.65 | 0.66 | 0.69 |
| Neugrin | NGRN | 1.04 | 1.57* | 1.71* | 1.11 | 0.65 | 0.56* | 0.90 |
| Asparagine synthetase [glutamine-hydrolyzing] | ASNS | 0.76 | 0.68* | 0.97 | 0.95 | 0.65 | 0.66 | 0.99 |
| Phosphatidate phosphatase LPIN3 | LPIN3 | 0.77 | 0.49* | 0.79 | 0.70 | 0.65 | 0.66 | 1.02 |
| Pre-mRNA-splicing factor ATP-dependent RNA helicase PRP16 | DHX38 | 0.84 | 0.66* | 0.64* | 0.68* | 0.65 | 0.66 | 0.68* |
| DNA mismatch repair protein Msh2 | MSH2 | 0.60* | 0.57* | 0.67* | 0.70 | 0.66 | 0.64 | 0.75 |
| Syndecan-1 | SDC1 | 0.64* | 0.52* | 0.65* | 0.81 | 0.67 | 0.59 | 0.73 |

***M. Common down-regulated proteins between LPS-Cytokines with Aquamin and with Mesalamine [37 proteins]***

| | | Interventions | | | | | | |
|---|---|---|---|---|---|---|---|---|
| | | Control | | | *With LPS & Cytokines* | | | |
| Proteins | Genes | AQ | AQ+MES | MES | *LPS-Cyto* | ***AQ*** | *AQ+MES* | ***MES*** |
| Group 10 secretory phospholipase A2 | PLA2G10 | 1.13 | 1.83* | 1.30 | 0.87 | 0.14* | 0.82 | 0.58 |
| Small nuclear ribonucleoprotein F | SNRPF | 2.07* | 1.62* | 1.69* | 1.59* | 0.18* | 0.89 | 0.54 |
| Cyclic AMP-dependent transcription factor ATF-2 | ATF2 | 0.70* | 0.75 | 0.87 | 0.71* | 0.29* | 0.70 | 0.51* |
| Protein cordon-bleu | COBL | 0.75 | 0.96 | 0.75 | 0.77 | 0.33* | 0.72 | 0.60 |
| Eukaryotic translation initiation factor 2D | EIF2D | 0.75 | 0.61* | 0.65* | 0.73 | 0.40* | 0.77 | 0.65 |
| DNA dC->dU-editing enzyme APOBEC-3C | APOBEC3C | 0.70 | 0.87 | 0.84 | 0.69 | 0.40* | 0.73 | 0.37* |
| Pre-mRNA-splicing factor SYF2 | SYF2 | 0.91 | 1.19 | 1.22 | 1.07 | 0.43* | 0.74 | 0.51* |
| Mitochondrial assembly of ribosomal large subunit protein 1 | MALSU1 | 0.92 | 0.82 | 0.88 | 1.04 | 0.44* | 0.73 | 0.53 |
| Receptor-type tyrosine-protein phosphatase gamma | PTPRG | 1.17 | 1.42 | 1.35 | 1.14 | 0.45* | 0.71 | 0.35* |
| Cyclin-D1-binding protein 1 | CCNDBP1 | 0.91 | 0.91 | 0.82 | 0.69 | 0.46* | 0.76 | 0.52* |
| Transmembrane 4 L6 family member 5 | TM4SF5 | 1.24 | 1.38 | 0.43* | 0.94 | 0.47* | 0.88 | 0.52 |
| Spermatogenesis-associated protein 20 | SPATA20 | 0.96 | 0.88 | 0.77 | 0.75 | 0.50* | 0.67 | 0.61* |
| Tetratricopeptide repeat protein 17 | TTC17 | 0.85 | 1.15 | 0.96 | 0.76 | 0.52 | 0.70 | 0.55 |
| Diacylglycerol kinase zeta | DGKZ | 0.78 | 0.87 | 0.94 | 1.11 | 0.54 | 0.73 | 0.57 |
| Sodium-dependent phosphate transporter 1 | SLC20A1 | 1.47* | 1.85* | 1.42* | 0.90 | 0.54 | 0.70 | 0.57 |
| Protein spire homolog 1 | SPIRE1 | 0.86 | 0.87 | 0.90 | 0.70 | 0.55 | 0.75 | 0.50* |

| | | | | | | | | |
|---|---|---|---|---|---|---|---|---|
| Putative nucleoside diphosphate kinase | NME2P1 | 2.99* | 1.90* | 1.48* | 2.39* | 0.55* | 0.75 | 0.60* |
| Elongation of very long chain fatty acids protein 7 | ELOVL7 | 0.95 | 0.94 | 1.07 | 0.92 | 0.55* | 0.74 | 0.58* |
| Mitogen-activated protein kinase kinase kinase 2 | MAP3K2 | 0.81 | 0.83 | 0.67 | 0.76 | 0.57 | 0.70 | 0.44* |
| Negative elongation factor C/D | NELFCD | 1.15 | 1.00 | 1.02 | 0.80 | 0.58* | 0.72 | 0.62* |
| Mitochondrial fission regulator 1 | MTFR1 | 0.73 | 1.14 | 1.06 | 0.72 | 0.59 | 0.69 | 0.63 |
| Testis development-related protein | TDRP | 1.05 | 1.04 | 1.14 | 0.78 | 0.60 | 0.70 | 0.44* |
| Natural cytotoxicity triggering receptor 3 ligand 1 | NCR3LG1 | 1.33 | 0.74 | 1.17 | 0.69 | 0.61 | 0.77 | 0.55 |
| Rab9 effector protein with kelch motifs | RABEPK | 0.97 | 0.97 | 0.96 | 0.98 | 0.61 | 0.68 | 0.65 |
| Ribosomal protein eL42-like | RPL36AL | 0.59* | 0.84 | 0.84 | 0.70 | 0.61 | 0.72 | 0.65 |
| Metalloreductase STEAP3 | STEAP3 | 1.43* | 1.45* | 1.12 | 0.90 | 0.61 | 0.82 | 0.59 |
| Interferon regulatory factor 6 | IRF6 | 1.13 | 1.02 | 1.10 | 0.84 | 0.63 | 0.71 | 0.63* |
| Caspase activity and apoptosis inhibitor 1 | CAAP1 | 0.97 | 0.96 | 0.83 | 0.74 | 0.63 | 0.80 | 0.66 |
| Kelch-like protein 3 | KLHL3 | 1.27 | 0.99 | 0.89 | 0.86 | 0.63 | 1.17 | 0.61 |
| DNA-directed RNA polymerase, mitochondrial | POLRMT | 1.04 | 0.95 | 0.97 | 0.83 | 0.64 | 0.74 | 0.57 |
| Mitofusin-1 | MFN1 | 0.84 | 1.05 | 1.04 | 0.77 | 0.64 | 0.78 | 0.51* |
| E3 ubiquitin-protein ligase RNF14 | RNF14 | 0.84 | 1.01 | 0.80 | 0.73 | 0.64 | 0.73 | 0.60 |
| Keratin, type I cytoskeletal 20 | KRT20 | 1.04 | 1.03 | 0.88 | 0.94 | 0.65* | 0.84 | 0.64* |
| Proline-rich protein 15-like protein | PRR15L | 0.92 | 0.94 | 0.94 | 0.69 | 0.65 | 0.80 | 0.57 |
| Threonylcarbamoyladenosine tRNA methylthiotransferase | CDKAL1 | 0.82 | 0.96 | 1.01 | 0.68 | 0.65 | 0.69 | 0.57 |
| Thioredoxin-like protein 4B | TXNL4B | 1.26 | 1.11 | 1.35 | 1.29 | 0.66 | 1.11 | 0.52 |
| Mucin-19 | MUC19 | 1.02 | 0.89 | 0.72 | 0.69 | 0.67 | 0.73 | 0.41* |

***N. Common down-regulated proteins between LPS-Cytokines with Aquamin plus Mesalamine and with Mesalamine [44 proteins]***

| | | Interventions | | | | | | |
|---|---|---|---|---|---|---|---|---|
| | | Control | | | *With LPS & Cytokines* | | | |
| Proteins | Genes | AQ | AQ+MES | MES | *LPS-Cyto* | *AQ* | ***AQ+MES*** | ***MES*** |
| Glutathione S-transferase A2 | GSTA2 | 1.36* | 0.40* | 0.10* | 2.33* | 1.56* | 0.19* | 0.50* |
| 3 beta-hydroxysteroid dehydrogenase/Delta 5-->4-isomerase type 2 | HSD3B2 | 1.43* | 0.44* | 0.24* | 0.88 | 1.24 | 0.30* | 0.21* |
| Meprin A subunit beta | MEP1B | 1.18 | 0.37* | 0.25* | 1.11 | 1.11 | 0.36* | 0.29* |
| Keratin, type I cytoskeletal 10 | KRT10 | 0.29* | 0.34* | 0.79* | 0.85 | 0.85 | 0.37* | 0.57* |
| Large ribosomal subunit protein mL54 | MRPL54 | 1.08 | 0.82 | 0.81 | 0.94 | 0.84 | 0.38* | 0.33* |
| Keratin, type II cytoskeletal 71 | KRT71 | 0.36* | 0.52* | 0.41* | 0.94 | 1.15 | 0.39* | 0.51* |
| Carboxypeptidase O | CPO | 1.36* | 0.54* | 0.34* | 1.41* | 1.27 | 0.41* | 0.39* |
| Ornithine transcarbamylase, mitochondrial | OTC | 1.13 | 0.62* | 0.42* | 0.86 | 1.03 | 0.41* | 0.48* |
| Proline-rich protein 9 | PRR9 | 1.25 | 15.97* | 11.14* | 0.74 | 1.51 | 0.43* | 0.33* |
| Growth arrest-specific protein 6 | GAS6 | 1.31* | 0.74 | 0.52* | 1.10 | 1.23 | 0.47* | 0.54* |

| | | | | | | | | |
|---|---|---|---|---|---|---|---|---|
| DCC-interacting protein 13-alpha | APPL1 | 0.87 | 0.65* | 0.89 | 0.72* | 0.67 | 0.48* | 0.61* |
| Small proline-rich protein 2D | SPRR2D | 0.49* | 0.44* | 1.18 | 2.59* | 0.70 | 0.50* | 0.58* |
| Integrin alpha-1 | ITGA1 | 0.90 | 0.54* | 0.51* | 0.80 | 0.98 | 0.51* | 0.52* |
| Keratin, type I cytoskeletal 14 | KRT14 | 0.51* | 0.49* | 0.57* | 1.84* | 0.71* | 0.52* | 0.61* |
| Carboxymethylenebutenolidase homolog | CMBL | 1.06 | 0.80 | 0.84* | 0.74* | 0.83 | 0.53* | 0.63* |
| Methyltransferase-like protein 17, mitochondrial | METTL17 | 1.00 | 0.69 | 0.99 | 0.86 | 0.78 | 0.53 | 0.65 |
| Rhomboid-related protein 2 | RHBDL2 | 0.90 | 0.80 | 1.42 | 0.77 | 0.73 | 0.54 | 0.60 |
| Desmocollin-1 | DSC1 | 0.54* | 0.35* | 0.53* | 0.79 | 1.12 | 0.55* | 0.56* |
| E3 ubiquitin-protein ligase SH3RF1 | SH3RF1 | 0.66 | 0.85 | 0.79 | 0.70 | 0.69 | 0.55 | 0.45* |
| Beta-chimaerin | CHN2 | 0.86 | 0.57* | 0.62* | 0.94 | 0.85 | 0.56* | 0.58* |
| Polyhomeotic-like protein 2 | PHC2 | 0.54* | 0.75 | 0.61* | 0.74 | 0.79 | 0.56 | 0.55 |
| Iodotyrosine deiodinase 1 | IYD | 0.94 | 0.57* | 0.46* | 0.68* | 1.05 | 0.57* | 0.51* |
| Large ribosomal subunit protein bL34m | MRPL34 | 1.01 | 0.83 | 0.83 | 0.75 | 0.79 | 0.57 | 0.59 |
| Mitochondrial inner membrane protease ATP23 homolog | ATP23 | 1.03 | 0.58* | 0.47* | 0.82 | 0.73 | 0.58 | 0.46* |
| Thiosulfate:glutathione sulfurtransferase | TSTD1 | 0.92 | 0.78 | 0.80 | 0.84 | 0.71 | 0.59 | 0.57 |
| Protein TANC1 | TANC1 | 0.62* | 0.71 | 0.76 | 0.69 | 0.87 | 0.59 | 0.63 |
| Kinesin-like protein KIF23 | KIF23 | 0.95 | 0.64* | 0.70* | 0.85 | 0.70* | 0.59* | 0.66* |
| Coiled-coil domain-containing protein 124 | CCDC124 | 0.57* | 0.56* | 0.69 | 0.84 | 0.69 | 0.60 | 0.56 |
| AF4/FMR2 family member 4 | AFF4 | 1.09 | 1.30 | 1.21 | 1.27 | 1.00 | 0.60 | 0.59 |
| ADP-ribosylation factor-like protein 14 | ARL14 | 0.93 | 0.60* | 0.53* | 0.92 | 0.82 | 0.60 | 0.63 |
| Uncharacterized protein C1orf198 | C1orf198 | 0.84 | 1.29 | 0.88 | 0.77 | 0.71 | 0.60 | 0.26* |
| Zinc transporter ZIP5 | SLC39A5 | 1.47* | 0.68 | 0.34* | 1.24 | 1.57 | 0.61 | 0.44* |
| Rho guanine nucleotide exchange factor 12 | ARHGEF12 | 0.78 | 0.82 | 0.82 | 0.68 | 0.76 | 0.61 | 0.66 |
| UDP-glucuronosyltransferase 2B15 | UGT2B15 | 0.87 | 0.69* | 0.53* | 0.73 | 0.92 | 0.62 | 0.61* |
| Putative bifunctional UDP-N-acetylglucosamine transferase and deubiquitinase ALG13 | ALG13 | 1.06 | 0.87 | 0.79 | 0.89 | 0.68 | 0.63 | 0.59* |
| Dipeptidase 1 | DPEP1 | 1.38* | 0.85 | 0.70* | 0.98 | 1.13 | 0.63* | 0.58* |
| All trans-polyprenyl-diphosphate synthase PDSS2 | PDSS2 | 1.18 | 0.97 | 0.91 | 0.90 | 0.77 | 0.64 | 0.52* |
| TGF-beta receptor type-2 | TGFBR2 | 0.83 | 0.58* | 0.65* | 0.69 | 0.94 | 0.64 | 0.60 |
| tRNA (cytosine(34)-C(5))-methyltransferase, mitochondrial | NSUN3 | 0.94 | 1.02 | 1.03 | 0.85 | 0.78 | 0.64 | 0.53 |
| Alpha-galactosidase A | GLA | 0.72* | 1.44* | 1.19* | 0.98 | 0.69* | 0.65* | 0.65* |
| Membrane-associated guanylate kinase, WW and PDZ domain-containing protein 1 | MAGI1 | 1.18 | 1.17 | 0.88 | 1.02 | 0.70 | 0.65 | 0.65 |
| UDP-glucuronosyltransferase 2B17 | UGT2B17 | 1.16 | 1.09 | 0.79* | 0.69* | 0.72* | 0.66* | 0.49* |
| Acireductone dioxygenase | ADI1 | 1.00 | 0.84 | 0.96 | 0.89 | 0.71 | 0.66 | 0.66 |
| GTP cyclohydrolase 1 feedback regulatory protein | GCHFR | 1.02 | 0.93 | 0.70* | 0.96 | 1.12 | 0.67 | 0.67 |

***O. Common down-regulated proteins among LPS-Cytokines with Aquamin, with Aquamin plus Mesalamine and with Mesalamine [184 proteins]***

| | | Interventions | | | | | | |
|---|---|---|---|---|---|---|---|---|
| | | Control | | | *With LPS & Cytokines* | | | |
| Proteins | Genes | AQ | AQ+MES | MES | *LPS-Cyto* | ***AQ*** | ***AQ+MES*** | ***MES*** |
| Multiple PDZ domain protein | MPDZ | 1.42* | 1.28 | 1.27 | 1.35 | 0.05* | 0.20* | 0.42* |
| M-phase phosphoprotein 6 | MPHOSPH6 | 1.02 | 0.90 | 1.03 | 0.68 | 0.05* | 0.42* | 0.48* |
| Pre-rRNA-processing protein TSR2 homolog | TSR2 | 1.13 | 1.05 | 1.08 | 1.05 | 0.06* | 0.23* | 0.35* |
| Zinc finger protein 595 | ZNF595 | 1.36 | 1.35 | 1.34 | 1.19 | 0.07* | 0.59 | 0.32* |
| Lysine-specific demethylase 9 | RSBN1 | 0.47* | 0.57* | 0.46* | 0.83 | 0.08* | 0.42* | 0.30* |
| Protein TMED8 | TMED8 | 1.19 | 1.33 | 1.27 | 1.01 | 0.08* | 0.53 | 0.22* |
| Surfeit locus protein 6 | SURF6 | 0.69* | 0.68* | 0.69* | 0.87 | 0.08* | 0.19* | 0.58* |
| PRKR-interacting protein 1 | PRKRIP1 | 1.25 | 0.93 | 1.09 | 1.01 | 0.09* | 0.20* | 0.34* |
| Cleavage stimulation factor subunit 2 tau variant | CSTF2T | 0.90 | 1.00 | 1.09 | 0.73 | 0.10* | 0.38* | 0.55 |
| Transcription initiation factor IIA subunit 1 | GTF2A1 | 1.15 | 1.08 | 1.24 | 1.08 | 0.12* | 0.35* | 0.48* |
| Cyclic AMP-responsive element-binding protein 1 | CREB1 | 0.74* | 0.72 | 0.81 | 0.71 | 0.13* | 0.32* | 0.43* |
| Glutamyl-tRNA(Gln) amidotransferase subunit A, mitochondrial | QRSL1 | 1.26* | 1.24* | 1.30* | 0.75* | 0.13* | 0.24* | 0.08* |
| Tudor domain-containing protein 3 | TDRD3 | 1.39 | 1.39 | 2.19* | 0.80 | 0.14* | 0.30* | 0.42* |
| Ribosome biogenesis protein NOP53 | NOP53 | 0.97 | 0.95 | 1.15 | 1.14 | 0.14* | 0.25* | 0.62* |
| Signal-induced proliferation-associated 1-like protein 1 | SIPA1L1 | 0.86 | 1.21 | 1.14 | 1.06 | 0.15* | 0.17* | 0.49* |
| E3 ubiquitin-protein ligase RING2 | RNF2 | 0.64* | 0.81 | 0.86 | 0.85 | 0.15* | 0.24* | 0.38* |
| PSME3-interacting protein | PSME3IP1 | 0.84 | 1.03 | 1.07 | 0.86 | 0.16* | 0.12* | 0.33* |
| Chromosome alignment-maintaining phosphoprotein 1 | CHAMP1 | 0.53* | 0.76 | 0.62* | 0.74 | 0.17* | 0.25* | 0.60* |
| Liprin-alpha-4 | PPFIA4 | 0.39* | 0.45* | 0.66* | 1.37 | 0.17* | 0.20* | 0.32* |
| Proteasome assembly chaperone 4 | PSMG4 | 1.35* | 1.22 | 1.32* | 1.11 | 0.18* | 0.24* | 0.43* |
| Ribosome biogenesis protein SLX9 homolog | SLX9 | 1.03 | 1.15 | 1.25 | 0.93 | 0.18* | 0.26* | 0.30* |
| Nibrin | NBN | 0.82 | 1.02 | 1.01 | 1.25 | 0.18* | 0.31* | 0.55* |
| Caspase-5 | CASP5 | 1.06 | 0.65* | 0.95 | 1.37 | 0.19* | 0.37* | 0.45* |
| Small integral membrane protein 20 | SMIM20 | 1.53* | 1.65* | 1.62* | 1.27* | 0.19* | 0.38* | 0.30* |
| Uncharacterized protein C1orf122 | C1orf122 | 0.98 | 1.23 | 1.01 | 1.22 | 0.20* | 0.46* | 0.54 |
| Acyl-CoA-binding domain-containing protein 6 | ACBD6 | 0.78 | 0.77 | 0.90 | 0.72 | 0.21* | 0.40* | 0.35* |
| Endoribonuclease YbeY | YBEY | 1.15 | 0.98 | 0.88 | 0.84 | 0.21* | 0.49* | 0.50* |
| Uncharacterized protein C11orf98 | C11orf98 | 1.07 | 0.66* | 0.84 | 0.91 | 0.21* | 0.21* | 0.25* |
| Leukocyte receptor cluster member 8 | LENG8 | 1.48* | 1.79* | 1.57* | 1.20 | 0.21* | 0.51* | 0.13* |
| Pseudouridylate synthase RPUSD2 | RPUSD2 | 1.20 | 1.09 | 1.13 | 0.98 | 0.22* | 0.29* | 0.31* |
| GPALPP motifs-containing protein 1 | GPALPP1 | 1.39* | 1.10 | 1.30 | 1.27 | 0.24* | 0.24* | 0.58 |

| | | | | | | | | |
|---|---|---|---|---|---|---|---|---|
| ATP-dependent RNA helicase DDX54 | DDX54 | 0.64* | 0.60* | 0.72 | 0.75 | 0.24* | 0.34* | 0.56 |
| ASNSD1 upstream open reading frame protein | ASDURF | 0.89 | 1.15 | 1.16* | 0.89 | 0.24* | 0.27* | 0.37* |
| Selenoprotein H | SELENOH | 1.24* | 1.05 | 1.06 | 1.15 | 0.25* | 0.35* | 0.53* |
| Coiled-coil-helix-coiled-coil-helix domain-containing protein 5 | CHCHD5 | 1.36* | 1.07 | 0.87 | 0.88 | 0.26* | 0.40* | 0.29* |
| SH2 domain-containing adapter protein B | SHB | 0.86 | 0.96 | 0.92 | 0.74 | 0.26* | 0.37* | 0.55* |
| DNA-directed RNA polymerase I subunit RPA43 | POLR1F | 0.99 | 1.01 | 1.03 | 0.77 | 0.26* | 0.46* | 0.40* |
| Coiled-coil domain-containing protein 86 | CCDC86 | 0.97 | 0.88 | 0.95 | 1.04 | 0.27* | 0.30* | 0.61 |
| DNA excision repair protein ERCC-1 | ERCC1 | 0.84 | 0.80 | 0.90 | 0.89 | 0.27* | 0.36* | 0.41* |
| Transcription factor E2F4 | E2F4 | 0.76 | 0.86 | 0.77 | 0.73 | 0.27* | 0.51* | 0.31* |
| Peptide deformylase, mitochondrial | PDF | 1.14 | 1.27 | 1.13 | 1.04 | 0.28* | 0.48* | 0.58 |
| Ribosomal RNA processing protein 36 homolog | RRP36 | 1.13 | 0.92 | 1.27 | 1.22 | 0.28* | 0.30* | 0.43* |
| Myomegalin | PDE4DIP | 0.52* | 0.90 | 0.94 | 0.87 | 0.30* | 0.31* | 0.60* |
| Formin-binding protein 1 | FNBP1 | 0.90 | 1.21 | 1.14 | 0.80 | 0.30* | 0.34* | 0.43* |
| Kynurenine--oxoglutarate transaminase 1 | KYAT1 | 0.87 | 0.84 | 0.98 | 0.85 | 0.30* | 0.48* | 0.51* |
| Y-box-binding protein 3 | YBX3 | 1.16 | 1.01 | 1.30 | 0.77 | 0.31* | 0.47* | 0.65 |
| WASH complex subunit 2C | WASHC2C | 0.85 | 0.84 | 0.96 | 1.02 | 0.31* | 0.42* | 0.58* |
| Protein FAN | NSMAF | 0.85 | 0.90 | 1.07 | 0.78 | 0.31* | 0.36* | 0.36* |
| Ribosomal RNA processing protein 1 homolog B | RRP1B | 0.84 | 0.85 | 0.81* | 0.71* | 0.31* | 0.39* | 0.45* |
| Cdc42 effector protein 4 | CDC42EP4 | 1.18 | 0.89 | 1.25 | 0.79 | 0.32* | 0.51* | 0.40* |
| Cyclin-dependent kinase inhibitor 2A | CDKN2A | 1.01 | 1.03 | 1.18 | 1.53* | 0.32* | 0.36* | 0.49* |
| KAT8 regulatory NSL complex subunit 3 | KANSL3 | 1.13 | 0.85 | 0.80 | 0.99 | 0.33* | 0.35* | 0.33* |
| Regulation of nuclear pre-mRNA domain-containing protein 1A | RPRD1A | 0.79* | 0.77 | 0.81* | 0.78 | 0.33* | 0.53* | 0.50* |
| Myocyte-specific enhancer factor 2D | MEF2D | 0.78 | 1.08 | 1.11 | 0.83 | 0.33* | 0.41* | 0.65 |
| Max-like protein X | MLX | 0.96 | 1.07 | 0.98 | 0.86 | 0.33* | 0.38* | 0.47* |
| Tubulin-specific chaperone C | TBCC | 0.92 | 0.86 | 0.97 | 1.05 | 0.34* | 0.47* | 0.60* |
| Splicing factor Cactin | CACTIN | 1.09 | 1.00 | 1.12 | 1.04 | 0.35* | 0.40* | 0.56* |
| Protein SDA1 homolog | SDAD1 | 0.65* | 0.70 | 0.69* | 0.69 | 0.35* | 0.42* | 0.47* |
| Ubiquitin-like domain-containing CTD phosphatase 1 | UBLCP1 | 0.93 | 1.03 | 1.08 | 0.98 | 0.35* | 0.51* | 0.63* |
| N6-adenosine-methyltransferase catalytic subunit | METTL3 | 1.04 | 1.24 | 1.19 | 0.71* | 0.36* | 0.32* | 0.49* |
| Transmembrane 4 L6 family member 4 | TM4SF4 | 0.69 | 0.68 | 0.62* | 0.67 | 0.36* | 0.58 | 0.59 |
| A-kinase anchor protein 2 | PALM2AKAP2 | 1.01 | 0.97 | 1.24 | 0.92 | 0.36* | 0.58 | 0.67 |
| Nucleoplasmin-3 | NPM3 | 1.99* | 1.31 | 1.49 | 2.28* | 0.37* | 0.65 | 0.30* |
| DNA repair protein XRCC4 | XRCC4 | 1.08 | 1.04 | 1.20 | 0.99 | 0.37* | 0.48* | 0.49* |
| Ubiquinol-cytochrome-c reductase complex assembly factor 6 | UQCC6 | 1.50* | 1.63* | 1.62* | 1.17 | 0.37* | 0.43* | 0.39* |
| 5-methylcytosine rRNA methyltransferase NSUN4 | NSUN4 | 0.91 | 0.99 | 0.97 | 0.81 | 0.38* | 0.56 | 0.35* |

| | | | | | | | | |
|---|---|---|---|---|---|---|---|---|
| GrpE protein homolog 2, mitochondrial | GRPEL2 | 1.26* | 1.12 | 1.07 | 0.79 | 0.38* | 0.43* | 0.38* |
| Bromodomain-containing protein 4 | BRD4 | 1.20* | 1.20 | 1.19* | 0.96 | 0.38* | 0.50* | 0.58* |
| DnaJ homolog subfamily C member 21 | DNAJC21 | 0.94 | 0.94 | 0.87 | 0.82 | 0.38* | 0.41* | 0.55* |
| Actin-related protein 5 | ACTR5 | 0.96 | 0.83 | 1.04 | 0.80 | 0.39* | 0.64 | 0.54 |
| Large ribosomal subunit protein mL65 | MRPS30 | 0.95 | 0.89 | 0.84 | 0.72* | 0.39* | 0.45* | 0.34* |
| Endoplasmic reticulum resident protein 27 | ERP27 | 1.40* | 1.22 | 1.11 | 0.83 | 0.39* | 0.57 | 0.37* |
| Sorting nexin-15 | SNX15 | 1.13 | 1.03 | 0.91 | 0.76 | 0.40* | 0.45* | 0.36* |
| Fos-related antigen 2 | FOSL2 | 0.85 | 0.88 | 1.00 | 0.74 | 0.40* | 0.37* | 0.41* |
| E3 ubiquitin-protein ligase ZNRF2 | ZNRF2 | 0.99 | 1.08 | 1.00 | 0.81 | 0.40* | 0.52* | 0.51* |
| Pleckstrin homology-like domain family B member 1 | PHLDB1 | 0.73* | 0.61* | 0.53* | 0.70* | 0.41* | 0.32* | 0.43* |
| AN1-type zinc finger protein 2B | ZFAND2B | 0.91 | 0.99 | 0.88 | 0.76 | 0.42* | 0.59 | 0.51* |
| SID1 transmembrane family member 2 | SIDT2 | 1.03 | 1.02 | 0.97 | 0.88 | 0.42* | 0.50 | 0.40* |
| DNA polymerase subunit gamma-1 | POLG | 0.77 | 0.98 | 0.89 | 0.71 | 0.42* | 0.45* | 0.42* |
| Zinc finger protein 330 | ZNF330 | 0.83 | 0.85 | 0.81 | 0.70 | 0.42* | 0.55 | 0.52* |
| Metallothionein-1H | MT1H | 0.61* | 0.90 | 0.77 | 0.97 | 0.42* | 0.55 | 0.62 |
| Shootin-1 | SHTN1 | 0.79* | 0.84 | 0.87 | 0.76* | 0.43* | 0.45* | 0.54* |
| p53 and DNA damage-regulated protein 1 | PDRG1 | 0.91 | 1.18 | 1.19 | 1.01 | 0.43* | 0.49* | 0.65* |
| Ribosome biogenesis protein NSA2 homolog | NSA2 | 0.71* | 0.71* | 0.76 | 0.88 | 0.44* | 0.48* | 0.62 |
| Fatty acyl-CoA reductase 1 | FAR1 | 0.73 | 0.75 | 0.99 | 0.67 | 0.44* | 0.48* | 0.36* |
| E3 ubiquitin-protein ligase RNF25 | RNF25 | 0.77 | 0.99 | 0.84 | 0.79 | 0.44* | 0.47* | 0.62 |
| Nucleolar protein 10 | NOL10 | 0.72* | 0.67* | 0.82 | 0.68* | 0.44* | 0.49* | 0.51* |
| TP53-regulated inhibitor of apoptosis 1 | TRIAP1 | 1.35 | 1.33 | 1.20 | 1.16 | 0.45* | 0.59 | 0.67 |
| Death-associated protein 1 | DAP | 0.68 | 0.53* | 0.55* | 0.77 | 0.45* | 0.43* | 0.48* |
| SUN domain-containing ossification factor | SUCO | 0.95 | 0.99 | 0.77 | 0.68 | 0.45* | 0.39* | 0.35* |
| Alpha-ketoglutarate-dependent dioxygenase alkB homolog 7, mitochondrial | ALKBH7 | 0.95 | 0.89 | 0.84 | 0.79 | 0.45* | 0.45* | 0.29* |
| 2-(3-amino-3-carboxypropyl)histidine synthase subunit 2 | DPH2 | 1.00 | 1.13 | 1.16 | 0.89 | 0.46* | 0.30* | 0.45* |
| Replication termination factor 2 | RTF2 | 0.68* | 0.83 | 0.82 | 0.68 | 0.46* | 0.48* | 0.49* |
| Kanadaptin | SLC4A1AP | 0.79 | 0.88 | 0.82 | 0.89 | 0.46* | 0.61 | 0.55 |
| Protein FAM83G | FAM83G | 1.12 | 1.58* | 1.13 | 0.85 | 0.46* | 0.56 | 0.54 |
| 28S rRNA (cytosine-C(5))-methyltransferase | NSUN5 | 0.76* | 0.72* | 0.80* | 0.75* | 0.47* | 0.51* | 0.53* |
| ADP-ribosylation factor-like protein 15 | ARL15 | 0.91 | 0.97 | 0.78 | 0.84 | 0.47* | 0.53* | 0.53* |
| Serine/threonine-protein kinase N1 | PKN1 | 0.71 | 0.71 | 0.84 | 0.82 | 0.47* | 0.63 | 0.64 |
| Receptor-type tyrosine-protein phosphatase F | PTPRF | 0.52* | 0.48* | 0.95 | 0.70* | 0.47* | 0.47* | 0.64* |
| DENN domain-containing protein 1A | DENND1A | 0.80 | 0.86 | 1.18 | 0.78 | 0.48* | 0.59 | 0.61 |
| MAP kinase-interacting serine/threonine-protein kinase 1 | MKNK1 | 0.76 | 0.68 | 0.83 | 0.71 | 0.48* | 0.55* | 0.61 |

| | | | | | | | | |
|---|---|---|---|---|---|---|---|---|
| Zinc finger FYVE domain-containing protein 21 | ZFYVE21 | 0.95 | 0.83 | 0.83 | 0.77 | 0.48* | 0.56* | 0.57* |
| Katanin p60 ATPase-containing subunit A1 | KATNA1 | 0.84 | 0.95 | 1.29 | 0.80 | 0.48* | 0.51 | 0.43* |
| Putative monooxygenase p33MONOX | KIAA1191 | 1.29 | 1.25 | 1.40* | 0.92 | 0.48* | 0.44* | 0.43* |
| CCAAT/enhancer-binding protein zeta | CEBPZ | 0.71* | 0.70* | 0.69* | 0.74 | 0.48* | 0.58* | 0.67 |
| tRNA (guanine(37)-N1)-methyltransferase | TRMT5 | 1.09 | 0.88 | 0.98 | 0.94 | 0.48* | 0.53* | 0.40* |
| tRNA dimethylallyltransferase | TRIT1 | 0.93 | 0.94 | 0.77 | 0.89 | 0.49* | 0.52* | 0.31* |
| Zinc finger protein 22 | ZNF22 | 0.46* | 0.37* | 0.55* | 0.77 | 0.49* | 0.35* | 0.36* |
| UPF0462 protein C4orf33 | C4orf33 | 1.07 | 0.98 | 0.89 | 0.77 | 0.49* | 0.46* | 0.43* |
| Glycylpeptide N-tetradecanoyltransferase 2 | NMT2 | 0.89 | 1.06 | 1.00 | 0.70* | 0.49* | 0.54* | 0.55* |
| Lariat debranching enzyme | DBR1 | 0.90 | 0.70 | 0.90 | 0.80 | 0.49* | 0.60 | 0.58 |
| Multivesicular body subunit 12A | MVB12A | 0.94 | 0.64* | 0.98 | 0.89 | 0.49* | 0.65 | 0.61 |
| TRAF family member-associated NF-kappa-B activator | TANK | 0.61* | 0.64 | 0.66* | 0.91 | 0.50* | 0.50* | 0.55 |
| Cysteine protease ATG4C | ATG4C | 1.22 | 1.51 | 1.51* | 1.04 | 0.50* | 0.27* | 0.39* |
| Rootletin | CROCC | 0.49* | 1.51* | 0.96 | 0.74 | 0.50* | 0.53* | 0.43* |
| MOB kinase activator 3B | MOB3B | 0.71 | 0.93 | 0.82 | 0.73 | 0.51* | 0.65 | 0.56 |
| Selenoprotein W | SELENOW | 1.07 | 0.68 | 0.66* | 0.86 | 0.51* | 0.45* | 0.40* |
| Prostaglandin reductase 3 | PTGR3 | 1.10 | 1.09 | 1.08 | 0.75 | 0.51* | 0.60* | 0.37* |
| N-terminal Xaa-Pro-Lys N-methyltransferase 1 | NTMT1 | 0.83 | 0.88 | 0.84 | 0.84 | 0.51 | 0.55 | 0.54* |
| Translational activator of cytochrome c oxidase 1 | TACO1 | 0.98 | 1.04 | 0.92 | 0.89 | 0.51* | 0.64* | 0.60* |
| Small ribosomal subunit protein bS21m | MRPS21 | 0.94 | 0.91 | 0.82 | 0.79 | 0.52* | 0.58* | 0.56* |
| Pseudouridylate synthase TRUB1 | TRUB1 | 1.04 | 1.11 | 1.03 | 0.73* | 0.52* | 0.57* | 0.56* |
| Ribosome biogenesis protein BOP1 | BOP1 | 0.66* | 0.58* | 0.70* | 0.77 | 0.53* | 0.54* | 0.56 |
| SNARE-associated protein Snapin | SNAPIN | 0.84 | 0.89 | 1.04 | 0.78 | 0.53* | 0.64 | 0.60 |
| tRNA (guanine(10)-N2)-methyltransferase homolog | TRMT11 | 0.96 | 0.98 | 0.90 | 0.96 | 0.53 | 0.67 | 0.61 |
| Protein N-terminal glutamine amidohydrolase | NTAQ1 | 0.79 | 1.04 | 0.94 | 0.70 | 0.53 | 0.64 | 0.47* |
| Peroxisomal acyl-coenzyme A oxidase 2 | ACOX2 | 1.19 | 1.07 | 0.89 | 0.75* | 0.53* | 0.51* | 0.41* |
| Transcription initiation factor TFIID subunit 7 | TAF7 | 0.91 | 0.90 | 0.80 | 1.04 | 0.53* | 0.52* | 0.61 |
| Protein KTI12 homolog | KTI12 | 1.00 | 1.10 | 1.20 | 0.98 | 0.54 | 0.47* | 0.47* |
| Actin-binding LIM protein 2 | ABLIM2 | 0.78 | 0.89 | 0.89 | 0.68* | 0.54* | 0.59 | 0.58* |
| IgGFc-binding protein | FCGBP | 1.09 | 1.13 | 1.23 | 0.82 | 0.54 | 0.51* | 0.57 |
| Protein AATF | AATF | 0.61* | 0.52* | 0.65* | 0.82 | 0.54* | 0.54* | 0.65 |
| DNA repair endonuclease XPF | ERCC4 | 0.69 | 0.78 | 0.78 | 0.75 | 0.54 | 0.57 | 0.50* |
| Phenylalanine--tRNA ligase, mitochondrial | FARS2 | 0.88 | 0.83 | 0.80 | 0.71* | 0.54* | 0.52* | 0.50* |
| SURP and G-patch domain-containing protein 1 | SUGP1 | 1.18 | 1.25 | 1.06 | 1.09 | 0.55 | 0.66 | 0.65 |
| BAG family molecular chaperone regulator 4 | BAG4 | 0.91 | 0.89 | 0.93 | 0.81 | 0.55 | 0.58 | 0.67 |
| Probable N-acetyltransferase 14 | NAT14 | 0.90 | 0.93 | 1.08 | 0.95 | 0.55* | 0.50* | 0.43* |
| Transforming acidic coiled-coil-containing protein 1 | TACC1 | 0.94 | 0.62* | 0.65* | 0.84 | 0.56* | 0.47* | 0.61 |

| | | | | | | | | |
|---|---|---|---|---|---|---|---|---|
| Calcium/calmodulin-dependent protein kinase type 1D | CAMK1D | 0.92 | 1.15 | 0.95 | 0.81 | 0.56* | 0.61 | 0.39* |
| Ephrin type-A receptor 1 | EPHA1 | 0.71* | 0.85 | 0.88 | 0.73 | 0.56* | 0.65 | 0.55* |
| Lon protease homolog 2, peroxisomal | LONP2 | 0.89 | 0.94 | 0.85 | 0.71 | 0.56 | 0.54* | 0.48* |
| Aminomethyltransferase, mitochondrial | AMT | 0.98 | 0.99 | 0.96 | 0.77 | 0.57* | 0.63 | 0.52* |
| Importin subunit alpha-1 | KPNA2 | 0.83 | 1.20 | 1.27* | 0.89 | 0.58* | 0.59* | 0.63* |
| Jupiter microtubule associated homolog 1 | JPT1 | 0.63* | 0.58* | 0.86 | 0.78 | 0.58 | 0.49* | 0.65 |
| Phosphoprotein associated with glycosphingolipid-enriched microdomains 1 | PAG1 | 1.00 | 1.02 | 1.11 | 0.85 | 0.58 | 0.56* | 0.61 |
| Ligand-dependent corepressor | LCOR | 0.88 | 0.85 | 0.89 | 0.72 | 0.58 | 0.62 | 0.58 |
| StAR-related lipid transfer protein 5 | STARD5 | 1.16 | 1.12 | 1.05 | 0.89 | 0.59 | 0.63 | 0.55* |
| Tetratricopeptide repeat protein 39A | TTC39A | 1.06 | 0.89 | 0.75* | 0.77 | 0.59* | 0.63* | 0.47* |
| Nucleolar protein 9 | NOP9 | 0.70 | 0.71 | 0.85 | 0.84 | 0.59 | 0.55 | 0.65 |
| Cytosolic iron-sulfur assembly component 2A | CIAO2A | 1.03 | 0.91 | 0.80* | 0.78 | 0.59* | 0.62 | 0.58* |
| Transcription initiation factor IIA subunit 2 | GTF2A2 | 1.36 | 1.35 | 1.54* | 1.37 | 0.59 | 0.36* | 0.48* |
| Kynurenine formamidase | AFMID | 0.96 | 0.74 | 0.63* | 0.86 | 0.60* | 0.58* | 0.56* |
| Kinesin-like protein KIF1B | KIF1B | 0.75 | 0.64* | 0.78 | 0.67 | 0.60 | 0.64 | 0.56 |
| Mitochondrial tRNA methylthiotransferase CDK5RAP1 | CDK5RAP1 | 0.68 | 0.62* | 0.56* | 0.69 | 0.60 | 0.56 | 0.58 |
| Telomeric repeat-binding factor 2 | TERF2 | 1.54* | 1.23 | 1.29 | 1.43 | 0.60 | 0.56 | 0.66 |
| Ethanolamine kinase 1 | ETNK1 | 1.21 | 1.20 | 0.99 | 0.88 | 0.61 | 0.66 | 0.64 |
| Frizzled-6 | FZD6 | 0.99 | 1.22 | 1.31 | 0.81 | 0.61 | 0.51 | 0.65 |
| Protein FAM118B | FAM118B | 0.81* | 0.72* | 0.75* | 0.89 | 0.61* | 0.60* | 0.62* |
| Mothers against decapentaplegic homolog 5 | SMAD5 | 0.84 | 0.87 | 0.86 | 0.67 | 0.61 | 0.51* | 0.53* |
| Biotinidase | BTD | 1.46* | 1.07 | 0.92 | 0.97 | 0.61* | 0.62* | 0.58* |
| Ral GTPase-activating protein subunit alpha-1 | RALGAPA1 | 0.64* | 0.71 | 0.75 | 0.76 | 0.62 | 0.55 | 0.61 |
| Histone-lysine N-trimethyltransferase SMYD5 | SMYD5 | 0.97 | 0.88 | 0.75 | 1.14 | 0.62 | 0.63 | 0.67 |
| ATP-dependent RNA helicase DDX50 | DDX50 | 0.81 | 0.80 | 0.84 | 0.81 | 0.62 | 0.52* | 0.62* |
| Unconventional prefoldin RPB5 interactor 1 | URI1 | 0.73 | 0.74 | 0.72 | 0.89 | 0.62 | 0.53 | 0.64 |
| NAD kinase | NADK | 0.84 | 0.85 | 0.77 | 0.89 | 0.63 | 0.66 | 0.65 |
| 8-oxo-dGDP phosphatase NUDT18 | NUDT18 | 1.02 | 0.93 | 0.92 | 0.83 | 0.63 | 0.53 | 0.62 |
| [F-actin]-monooxygenase MICAL2 | MICAL2 | 0.79 | 0.67* | 0.86 | 0.69 | 0.63 | 0.51* | 0.60* |
| Inositol polyphosphate multikinase | IPMK | 0.88 | 0.88 | 0.70* | 0.70 | 0.63 | 0.64 | 0.55* |
| NEDD8-conjugating enzyme UBE2F | UBE2F | 0.80 | 0.74 | 0.74 | 0.71 | 0.64 | 0.65 | 0.65 |
| Rac GTPase-activating protein 1 | RACGAP1 | 0.98 | 0.66* | 0.69* | 0.80 | 0.64* | 0.52* | 0.57* |
| COX assembly mitochondrial protein 2 homolog | CMC2 | 1.24* | 1.12 | 1.05 | 0.96 | 0.64 | 0.56* | 0.51* |
| Fatty acid-binding protein, intestinal | FABP2 | 1.03 | 0.91 | 0.93 | 0.72* | 0.64* | 0.59* | 0.63* |
| Transcription factor Sp6 | SP6 | 0.94 | 1.06 | 0.92 | 0.83 | 0.65 | 0.53 | 0.32* |
| 5-phosphohydroxy-L-lysine phospho-lyase | PHYKPL | 0.94 | 0.88 | 0.73* | 0.80 | 0.65 | 0.64 | 0.58* |
| Solute carrier family 12 member 9 | SLC12A9 | 1.02 | 0.83 | 0.88 | 0.80 | 0.65 | 0.57* | 0.53* |

| | | | | | | | | |
|---|---|---|---|---|---|---|---|---|
| Thioredoxin-related transmembrane protein 4 | TMX4 | 0.79 | 0.58* | 0.73 | 0.70 | 0.65 | 0.46* | 0.43* |
| RalA-binding protein 1 | RALBP1 | 0.81 | 0.83 | 0.73* | 0.79 | 0.65 | 0.59 | 0.64 |
| Origin recognition complex subunit 2 | ORC2 | 0.72* | 0.82 | 0.81 | 0.76 | 0.65 | 0.66 | 0.61* |
| Ras-related protein Rab-20 | RAB20 | 0.88 | 0.81 | 0.76* | 0.72* | 0.66 | 0.60* | 0.55* |
| Gap junction beta-1 protein | GJB1 | 0.62* | 0.56* | 0.56* | 0.75 | 0.66 | 0.53* | 0.58* |
| Protein phosphatase 1 regulatory subunit 1B | PPP1R1B | 1.00 | 1.05 | 1.01 | 0.79 | 0.66 | 0.66 | 0.53* |
| Kinesin-like protein KIF13A | KIF13A | 0.62* | 0.77 | 0.74* | 0.69 | 0.66 | 0.63 | 0.66 |
| Nuclear prelamin A recognition factor | NARF | 0.85 | 0.61* | 0.79 | 0.81 | 0.66 | 0.61 | 0.60 |
| Ribonucleoside-diphosphate reductase subunit M2 | RRM2 | 1.74* | 1.38 | 1.58* | 0.79 | 0.67 | 0.60 | 0.55* |

Values represent the abundance ratio from organoids (n=4 subjects) compared to the control. These proteins were down-regulated at a 1.5-fold change (<2% FDR). Corresponding abundance ratios from the other treatment groups are provided for comparison. Proteins common among groups and unique to individual groups under proinflammatory conditions (LPS-Cytokines) are presented. *Indicates significance compared to the control (at $p<0.05$).
